# Identification of potential key genes and pathway linked with sporadic Creutzfeldt-Jakob disease based on integrated bioinformatics analyses

**DOI:** 10.1101/2020.12.21.20248688

**Authors:** Basavaraj Vastrad, Chanabasayya Vastrad, Iranna Kotturshetti

## Abstract

Sporadic Creutzfeldt-Jakob disease (sCJD) is neurodegenerative disease also called prion disease linked with poor prognosis. The aim of the current study was to illuminate the underlying molecular mechanisms of sCJD. The mRNA microarray dataset GSE124571 was downloaded from the Gene Expression Omnibus database. Differentially expressed genes (DEGs) were screened. Pathway and GO enrichment analyses of DEGs were performed. Furthermore, the protein-protein interaction (PPI) network was predicted using the IntAct Molecular Interaction Database and visualized with Cytoscape software. In addition, hub genes and important modules were selected based on the network. Finally, we constructed target genes - miRNA regulatory network and target genes - TF regulatory network. Hub genes were validated. A total of 891 DEGs 448 of these DEGs presented significant up regulated, and the remaining 443 down regulated were obtained. Pathway enrichment analysis indicated that up regulated genes were mainly linked with glutamine degradation/glutamate biosynthesis, while the down regulated genes were involved in melatonin degradation. GO enrichment analyses indicated that up regulated genes were mainly linked with chemical synaptic transmission, while the down regulated genes were involved in regulation of immune system process. hub and target genes were selected from the PPI network, modules, and target genes - miRNA regulatory network and target genes - TF regulatory network namely YWHAZ, GABARAPL1, EZR, CEBPA, HSPB8, TUBB2A and CDK14. The current study sheds light on the molecular mechanisms of sCJD and may provide molecular targets and diagnostic biomarkers for sCJD.

## Introduction

Prion diseases of human beings are linked with the aggregation in the brain of an abnormal, slightly protease-resistant isoform, of cellular prion protein (PrPC) [1]. Prion diseases are neurodegenerative diseases that affect human beings [2]. However, dissimilar with the most common diseases of this group, such as Alzheimer’s disease, Lewy body disorders and frontotemporal dementias [3]. Sporadic Creutzfeldt-Jakob disease (sCJD) is the most prevalent human prion disease occurs through unknown origin [4]. sCJD is considered as a gene-related disease [5]. Expanding confirmation also has demonstrated that multiple genes and cellular pathways participate in the occurrence and advancement of sCJD [6-7]. Alteration and polymorhisum in prion protein was associated with progression of sCJD [8-9]. To date, a deficiency of understanding the accurate molecular mechanisms underlying sCJD development limits the ability to treat advanced disease. In recent years, microarray technology is now undergoing a revolution, which could be used on tissue or blood samples to diagnose key biomarkers and novel pathways in individual person [10-11]. Therefore, further knowledge of molecular mechanism associated in genetic expression disorder of sCJD, which is extremely key for the future advancement of diagnosis and treatment could be learned through this latest technology. Gene expression analysis [12] now is a popular approach to analyze the expression changes of gene in the advancement and progression of sCJD, comprehensively. In this current study, we downloaded the original gene expression dataset (GSE124571) from the Gene Expression Omnibus (GEO), which is repository leads to the archiving as a hub for microarray data deposit and retrieval. Gene expression profiles of brain tissue in patients with sCJD were compared with those in normal controls to diagnose the differentially expressed genes (DEGs). Selected DEGs were screened by pathway enrichment and gene ontology (GO) enrichment analysis. Subsequently, PPI network construction, module analysis, target genes - miRNA regulatory network construction and target genes - TF regulatory network construction were performed. Finally validation of hub genes was carried out. By using various bioinformatics tools, we may take a further insight of sCJD at molecular level and explore the potential diagnostic and prognostics biomarkers for the therapeutic strategies of sCJD.

## Materials and methods

### Microarray data

The gene expression profile dataset GSE124571 [13] was downloaded from the GEO database (www.ncbi.nlm.nih.gov/geo/) [14]. The platform for GSE124571 is GPL14951, [Human] Illumina HumanHT-12 WG-DASL V4.0 R2 expression beadchip. The platform files and raw data were downloaded as TXT files. Only data from sCJD and normal control samples were extracted and further analyzed.

### Data Preprocessing and DEG Screening

The R software package beadarray was used to preprocess the downloaded original TXT data. This process involved background adjustment, normalization and expression calculation. Probes not matching any known genes were deleted, and the mean was determined, when numerous probes were balanced to the same gene. The probe ID was converted into gene symbol and saved in a TXT file. The DEGs of sCJD and normal control samples were identified using the R bioconductor package limma [15]. Genes with an adjusted p value < 0.05, |logFC| > 0.856 for up regulated genes and |logFC| > −0.84 for down regulated genes were considered DEGs.

### Pathway enrichment analysis of DEGs

To analyze the identified DEGs at the functional level, BIOCYC (https://biocyc.org/) [16], Kyoto Encyclopedia of Genes and Genomes (KEGG) (http://www.genome.jp/kegg/pathway.html) [17], Pathway Interaction Database (PID) (https://wiki.nci.nih.gov/pages/viewpage.action?pageId=315491760) [18], REACTOME (https://reactome.org/) [19], GenMAPP (http://www.genmapp.org/) [20], MSigDB C2 BIOCARTA (v6.0) (http://software.broadinstitute.org/gsea/msigdb/collections.jsp) [21], PantherDB (http://www.pantherdb.org/) [22], Pathway Ontology (http://www.obofoundry.org/ontology/pw.html) [23] and Small Molecule Pathway Database (SMPDB) (http://smpdb.ca/) [24] pathway analysis were performed using ToppGene (https://toppgene.cchmc.org/enrichment.jsp) [25]. ToppGene is an updated web server which provides a comprehensive set of functional annotation tools for researchers to understand biological meanings behind a large list of genes. In this current study, we analyzed the candidate DEGs that were importantly up and down regulated, and P < 0.05 was set as the threshold value.

### Gene ontology (GO) enrichment analysis of DEGs

GO enrichment analysis (http://www.geneontology.org/) [26] is progressively tested for functional studies of large-scale genomic or transcriptomic data, which constitute three separate ontologies such as biological process (BP), molecular function (MF), and cellular component (CC). The usual up regulated and down regulated genes were analyzed using ToppGene (https://toppgene.cchmc.org/enrichment.jsp) [25], an online program that provides a complete set of function annotation tools for investigators to understand the biological meaning lists of genes. GO enrichment an analysis was performed using ToppGene. P < 0.05 was considered to indicate statistically significant difference.

### Protein–protein interaction network (PPI) and modular analysis

To study the interactive relationships among the DEGs, a PPI network of up and down regulated genes were constructed using the IntAct Molecular Interaction Database (https://www.ebi.ac.uk/intact/) [27], which integrates different PPI databases such as Molecular INTeraction Database (MINT, https://mint.bio.uniroma2.it/) [28], UniProt (https://www.uniprot.org/) [29], Interologous interaction database (I2D, http://ophid.utoronto.ca/ophidv2.204/) [30], InnateDB (https://www.innatedb.com/) [31], MatrixDB (http://matrixdb.univ-lyon1.fr/) [32] and The International Molecular Exchange Consortium (IMEx, http://www.imexconsortium.org/) [33]. Then, the PPI network was visualized using the Cytoscape software (http://www.cytoscape.org) [34]. In this network, each node is a DEG (up or down regulated genes), and the connections between nodes represent the interactions between these genes. PPI network topological properties nodes such as node degree [35], betweenness [36], stress [37], closeness [38] and clustering coefficient [39] were calculated. Modular analysis was conducted with the Molecular Complex Detection PEWCC1 [40] (version 1.3) app of Cytoscape software, and an PEWCC1 score > 2 was set as the cut-off criterion.

### Construction of target genes - miRNA regulatory network

miRNet (https://www.mirnet.ca/) [41] is a comprehensive database of miRNAs, which hosts predicted as well as validated miRNA binding sites. miRNet compares the identified miRNA binding sites with the results obtained from ten established target gene – miRNA prediction databases such as TarBase (http://diana.imis.athena-innovation.gr/DianaTools/index.php?r=tarbase/index) [42], miRTarBase (http://mirtarbase.mbc.nctu.edu.tw/php/download.php) [43], miRecords (http://miRecords.umn.edu/miRecords) [44], miR2Disease (http://www.mir2disease.org/) [45], HMDD (http://www.cuilab.cn/hmdd) [46], PhenomiR (http://mips.helmholtz-muenchen.de/phenomir/) [47], SM2miR (http://bioinfo.hrbmu.edu.cn/SM2miR/) [48], PharmacomiR (http://www.pharmaco-mir.org/) [49], EpimiR (http://bioinfo.hrbmu.edu.cn/EpimiR/) [50] and starBase (http://starbase.sysu.edu.cn/) [51]. The target gene – miRNA pairs were visualized by Cytoscape software (http://www.cytoscape.org) [34].

### Construction of target genes - TF regulatory network

NetworkAnalyst (https://www.networkanalyst.ca/) [52] is a web based tool which can search transcription factors for the input up and down regulated genes as well as assess the effect of a transcription factor on the expression of the target genes. In this study, the transcription factors of the target genes were predicted from NetworkAnalyst database which ingrates TF database ChEA (http://amp.pharm.mssm.edu/lib/chea.jsp) [53] and target genes - TF regulatory network was constructed and visualized by Cytoscape software(http://www.cytoscape.org) [34].

### Validation of hub genes

Receiver operating characteristic (ROC) curves were used to analyze the potential clinical significance of these hub genes as molecular prognostic markers in sCJD. Based on the obtained optimal prognostic gene biomarkers for sCJD, we established the GLM (generalized linear models) was a managed learning model by using the pROC package [54] in R software. The prognostic ability of this models was accessed by obtaining the area under a ROC curve (AUC), accuracy, sensitivity and specificity. P<0.05 was considered to indicate a statistically significant difference.

## Results

### Identification of DEGs in sCJD

After gene expression profile data processing and standardization, we screened DEGs in GSE124571 dataset using integrated bioinformatics analysis and the results are shown in Fig. 1A and Fig. 1B. The limma package of the R software identified 891 DEGs (Table 1), with the cutoff standard of P value < 0.05, |logFC| > 0.856 for up regulated genes and |logFC| > −0.84 for down regulated genes; 448 of these DEGs presented significant up regulated, and the remaining 443 down regulated. Fig. 2 presents the volcano plot of DEGs for data set. A cluster heatmap developed with R software showed the distribution of all up and down regulated genes (Fig. 3 and Fig. 4).

**Fig. 1.**
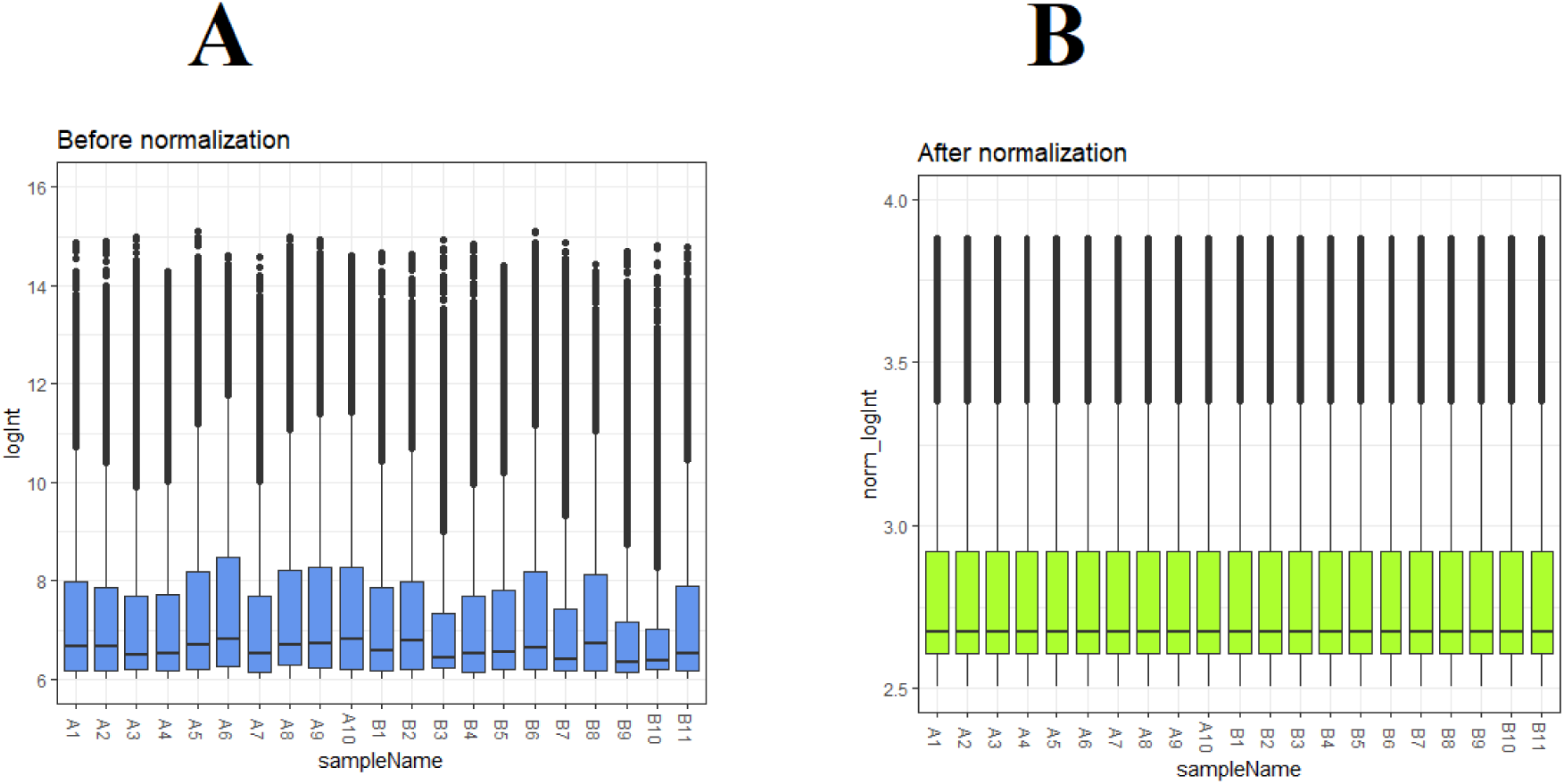
Box plots of the gene expression data before normalization (A) and after normalization (B). Horizontal axis represents the sample symbol and the vertical axis represents the gene expression values. The black line in the box plot represents the median value of gene expression. (A1, A2, A3, A4, A5, A6, A7, A8, A9, A10 = normal controls samples; B1,B2, B3, B4,B5, B6, B7,B8, B9, B10 = sCJD samples)

**Fig. 2.**
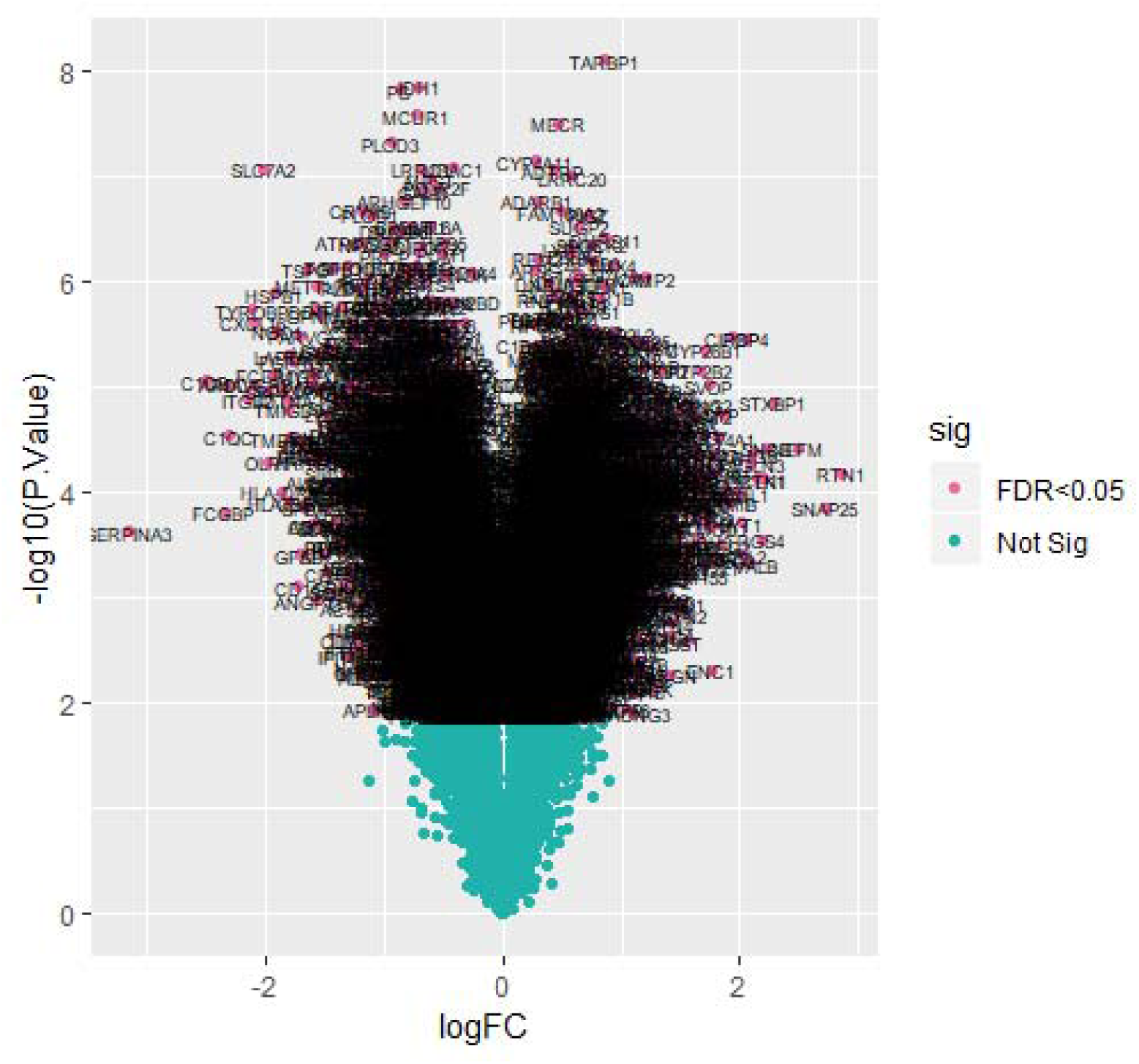
Volcano plot of differentially expressed genes. Genes with a significant change of more than two-fold were selected.

**Fig. 3.**
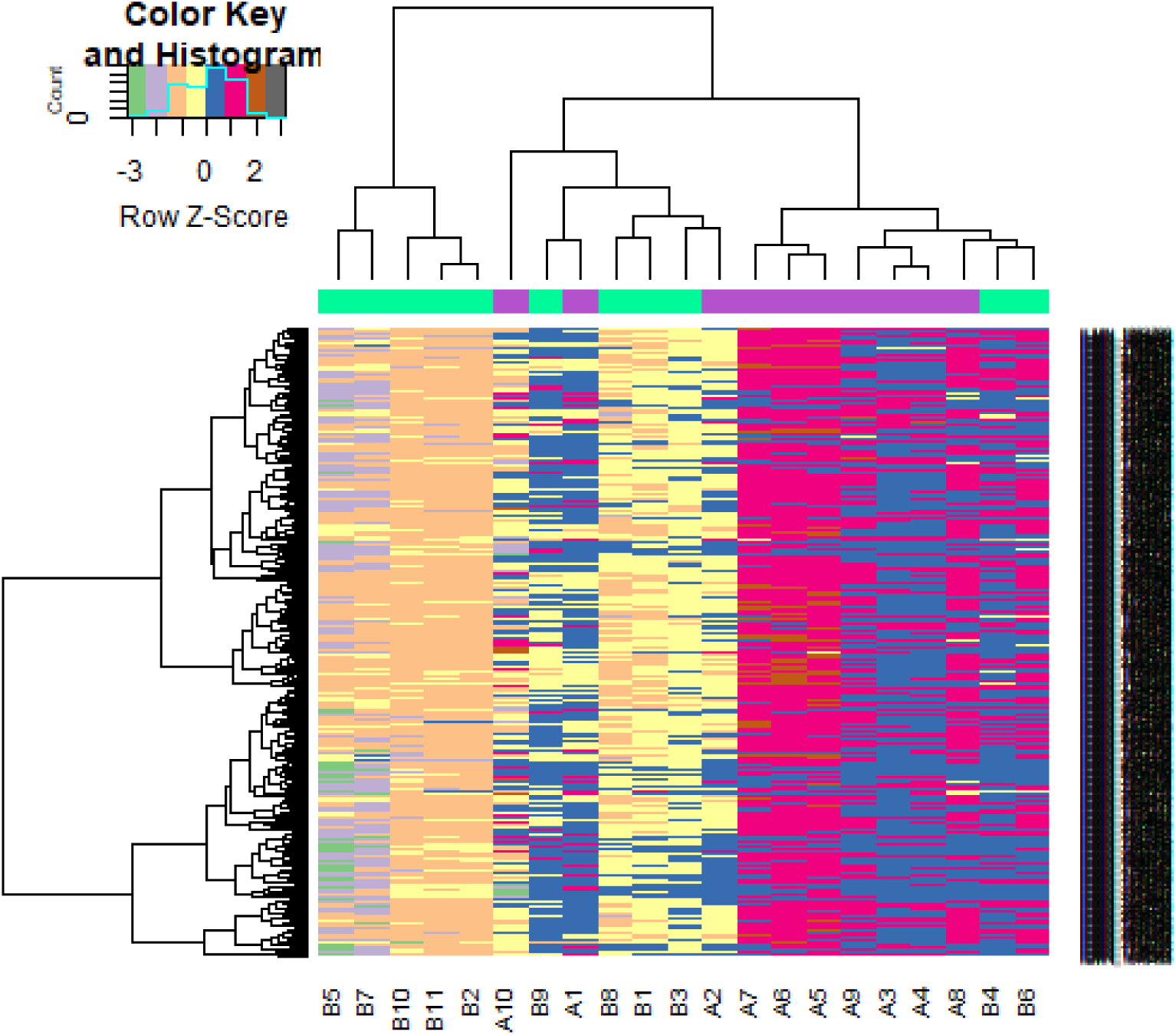
Heat map of up regulated differentially expressed genes. Legend on the top left indicate log fold change of genes. (A1, A2, A3, A4, A5, A6, A7, A8, A9, A10 = normal controls samples; B1,B2, B3, B4,B5, B6, B7,B8, B9, B10 = sCJD samples)

**Fig. 4.**
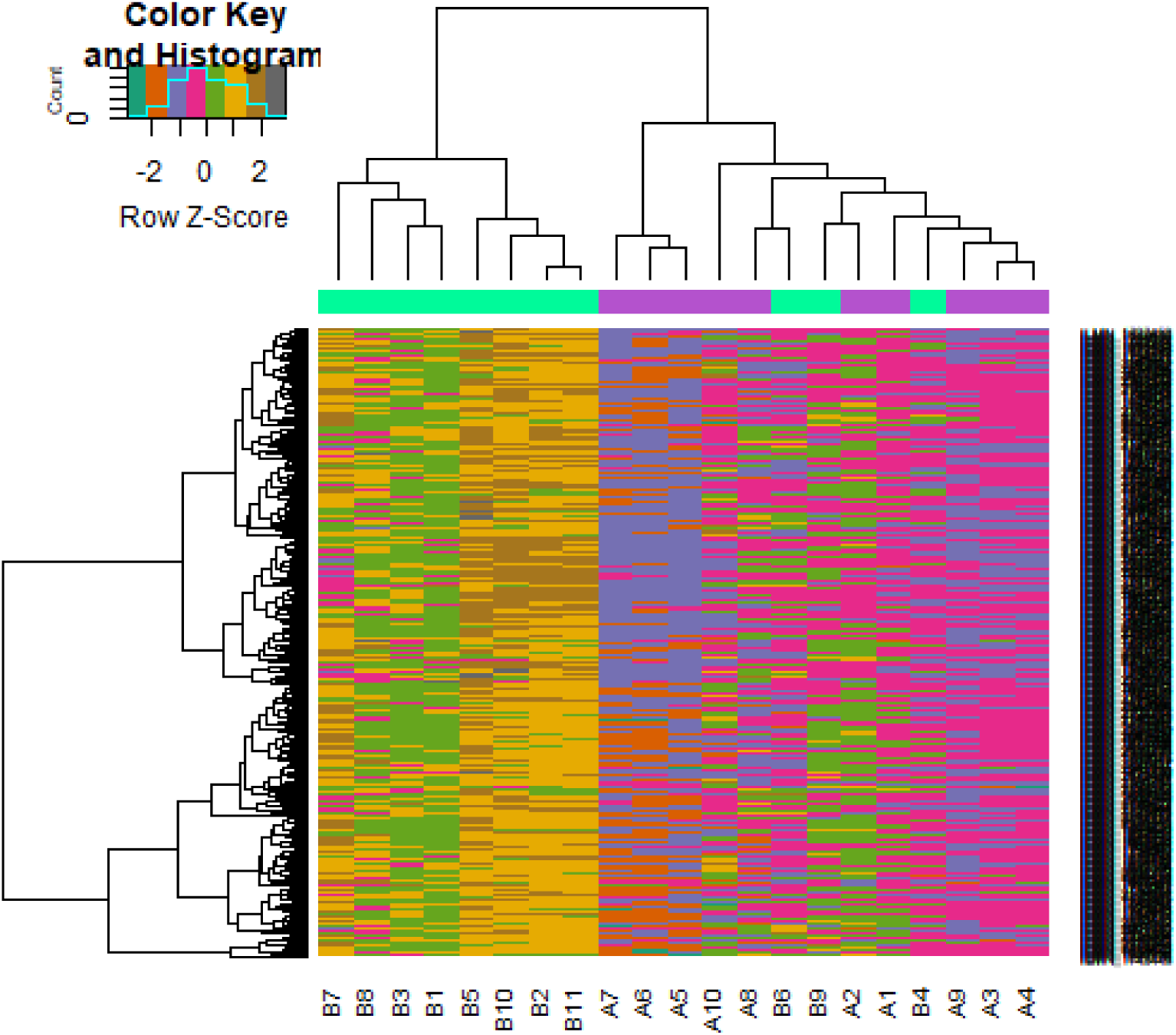
Heat map of down regulated differentially expressed genes. Legend on the top left indicate log fold change of genes. (A1, A2, A3, A4, A5, A6, A7, A8, A9, A10 = normal controls samples; B1,B2, B3, B4,B5, B6, B7,B8, B9, B10 = sCJD samples)

**Table 1.**
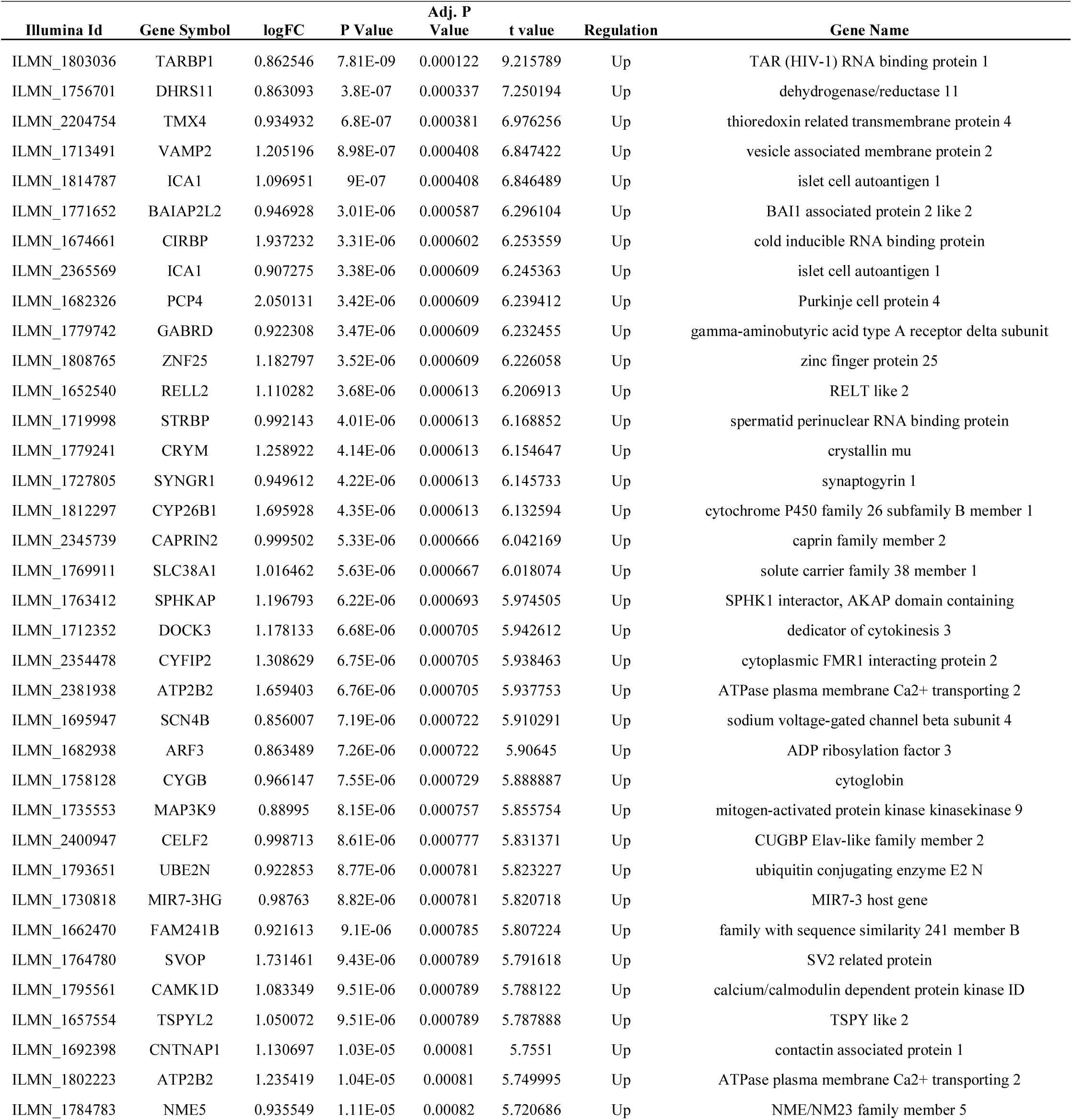
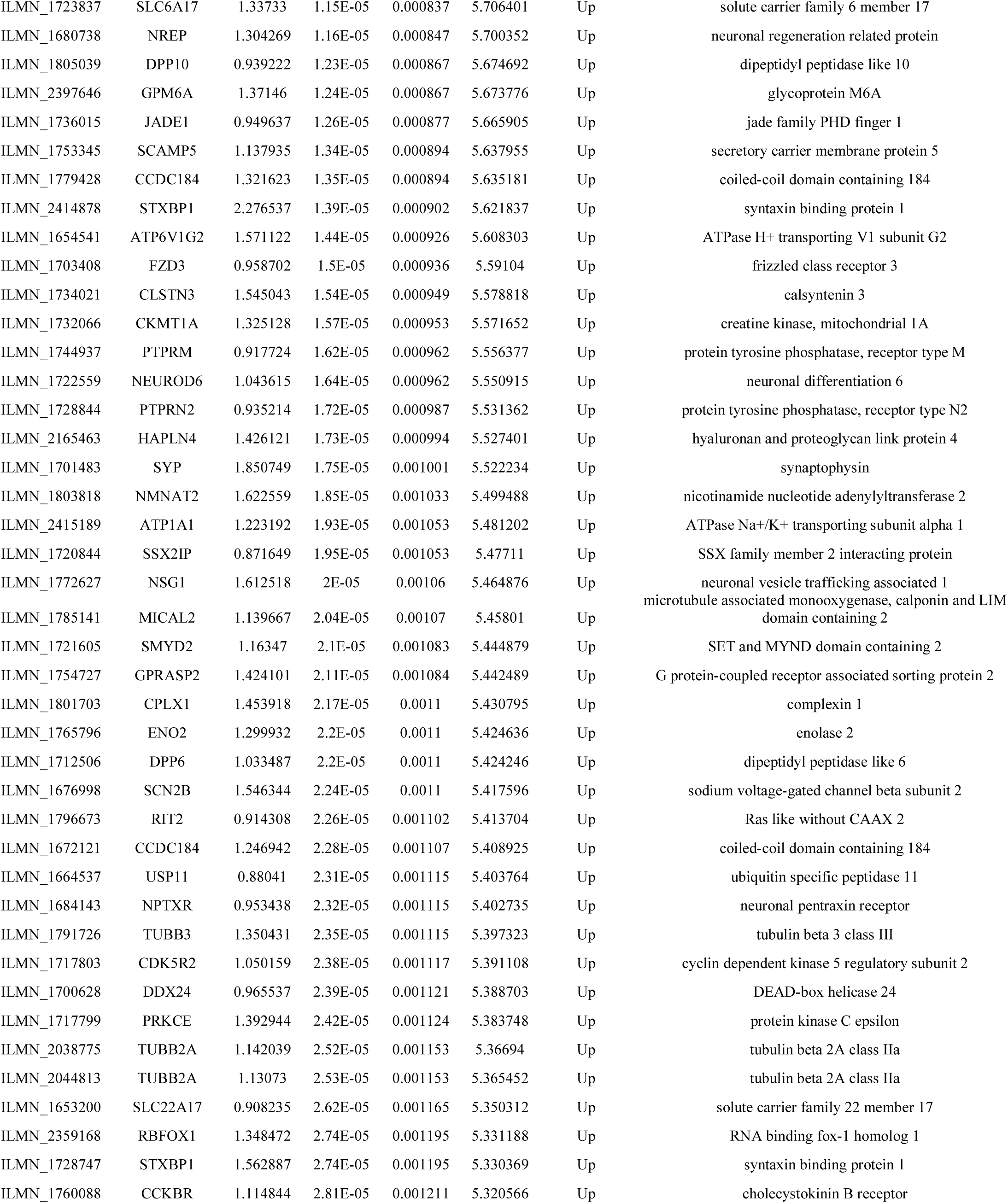
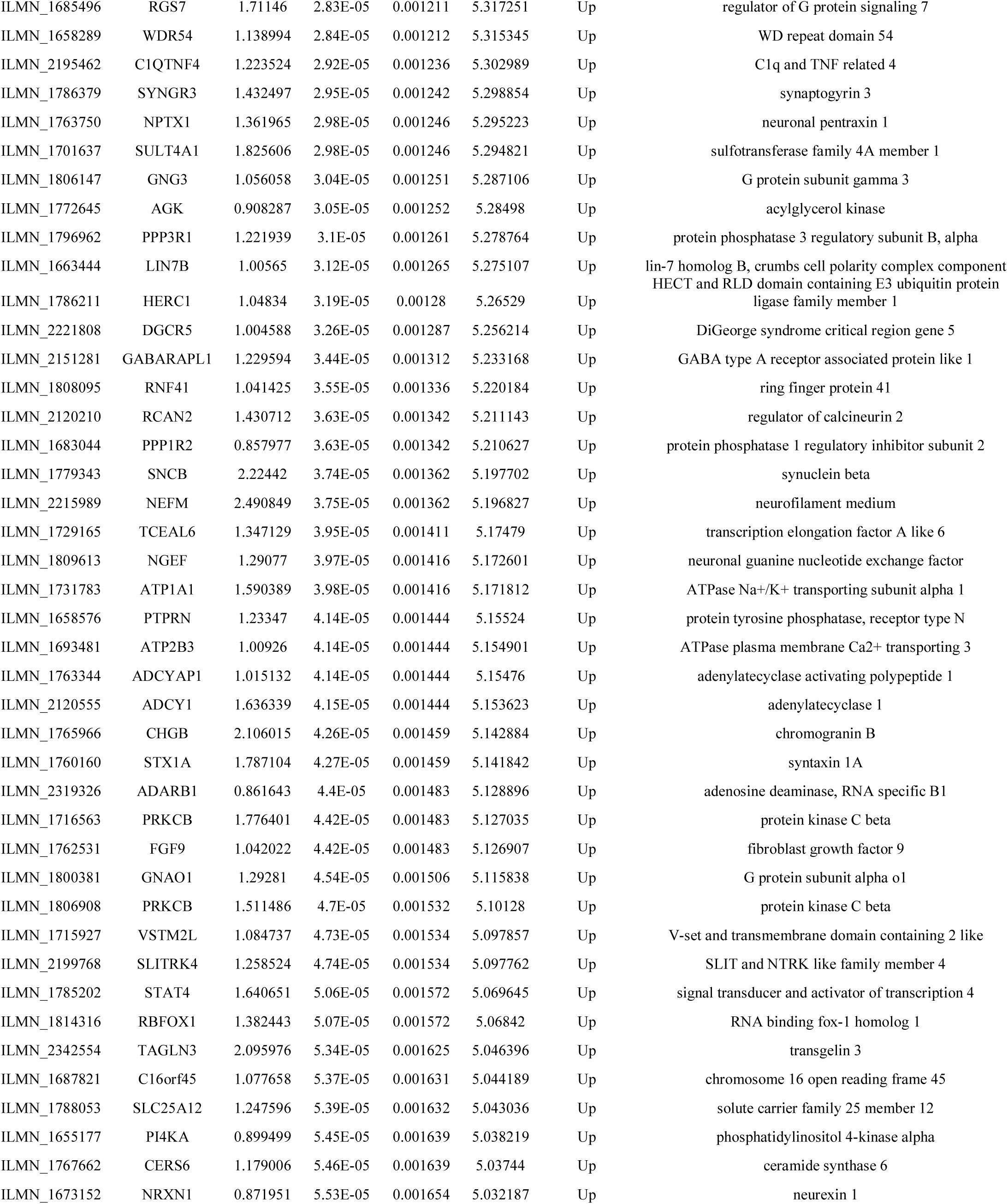
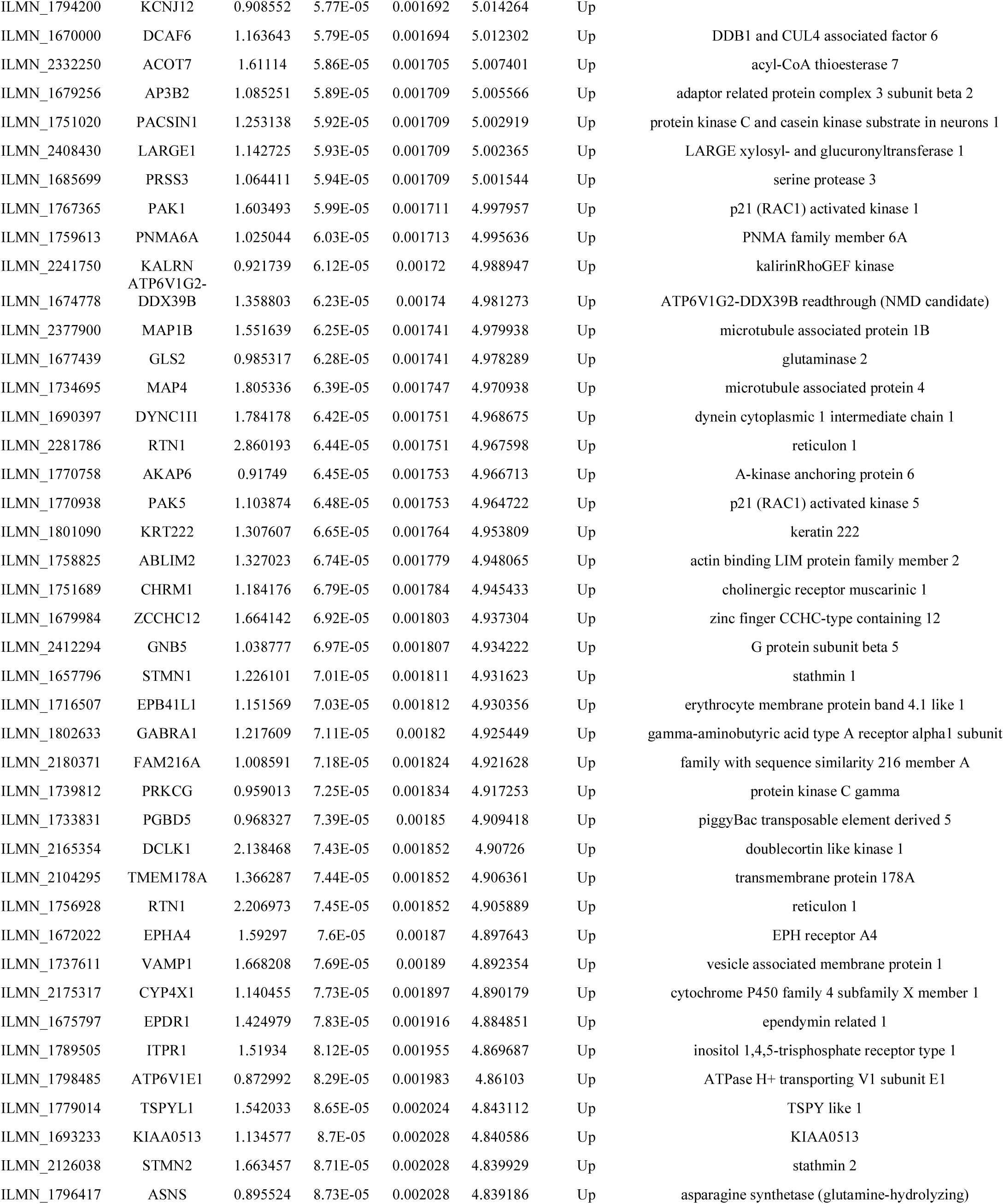
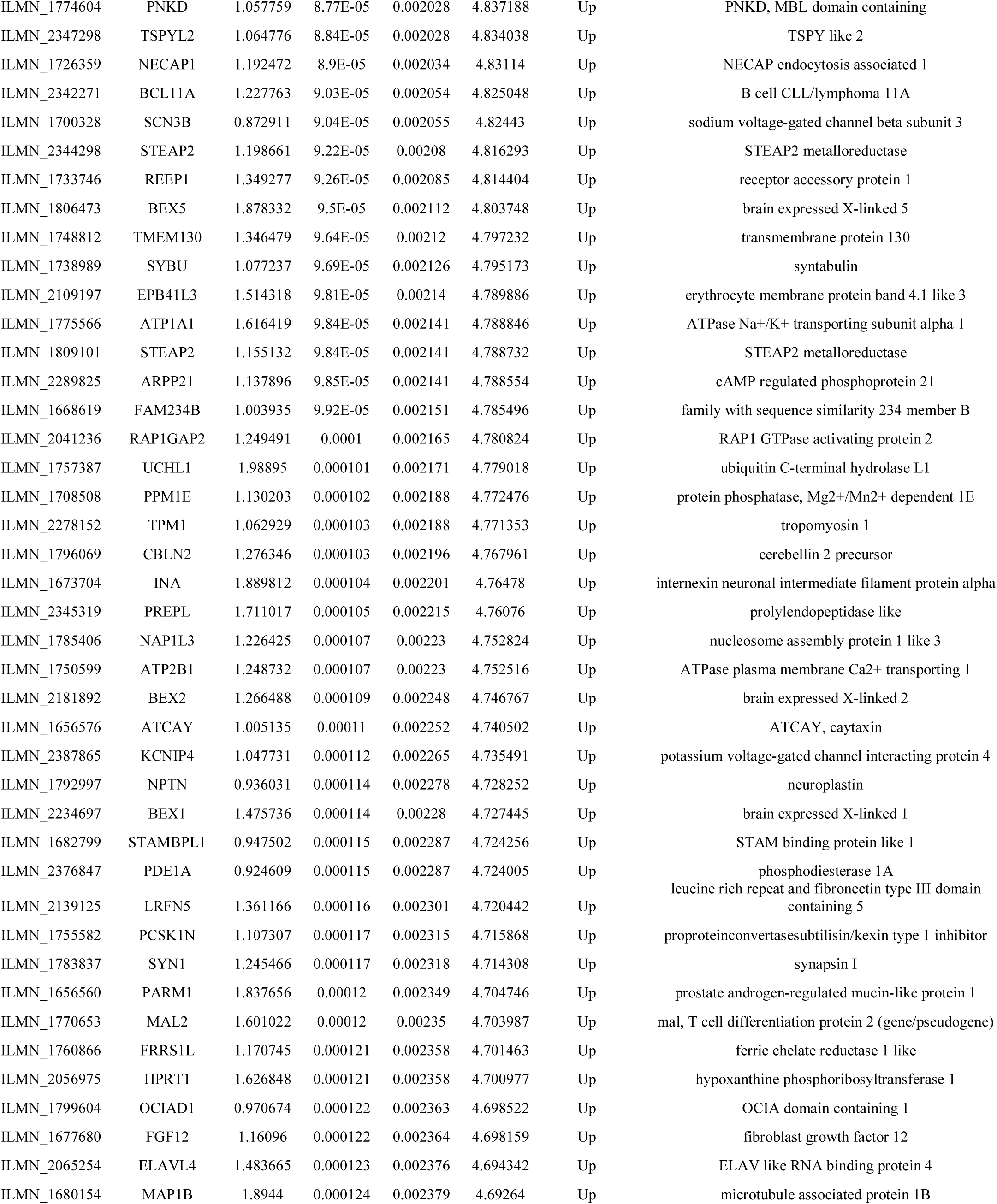
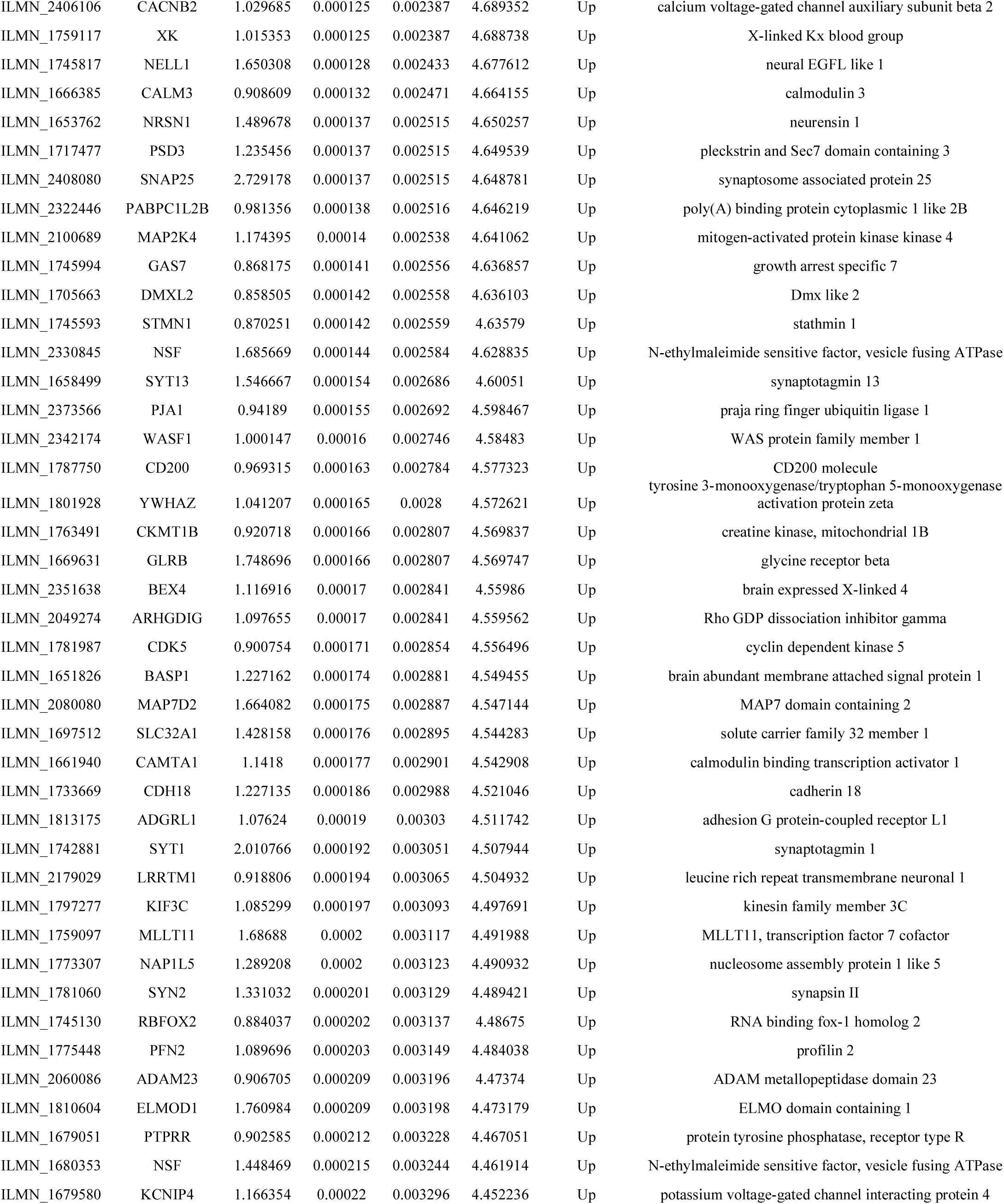
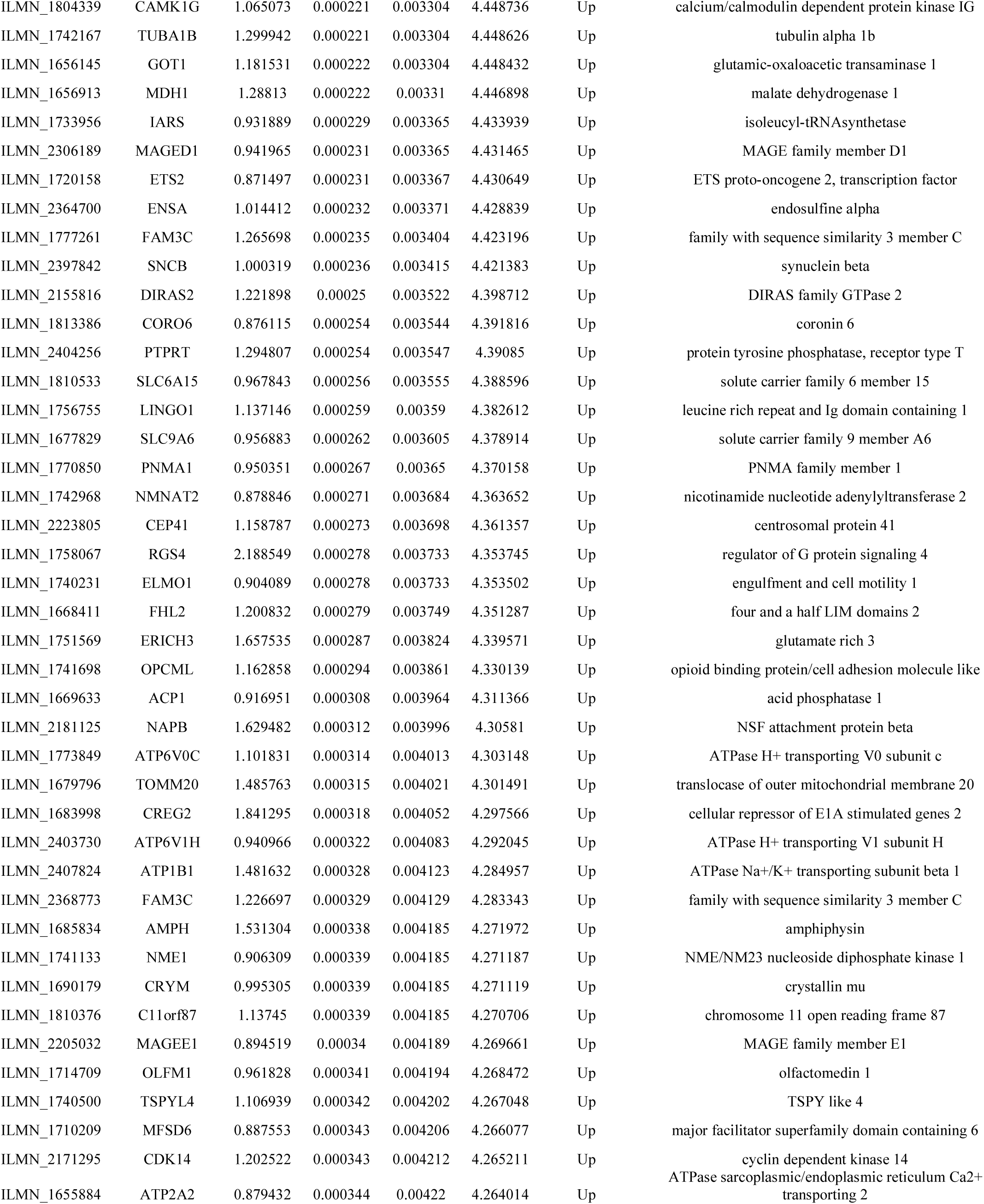
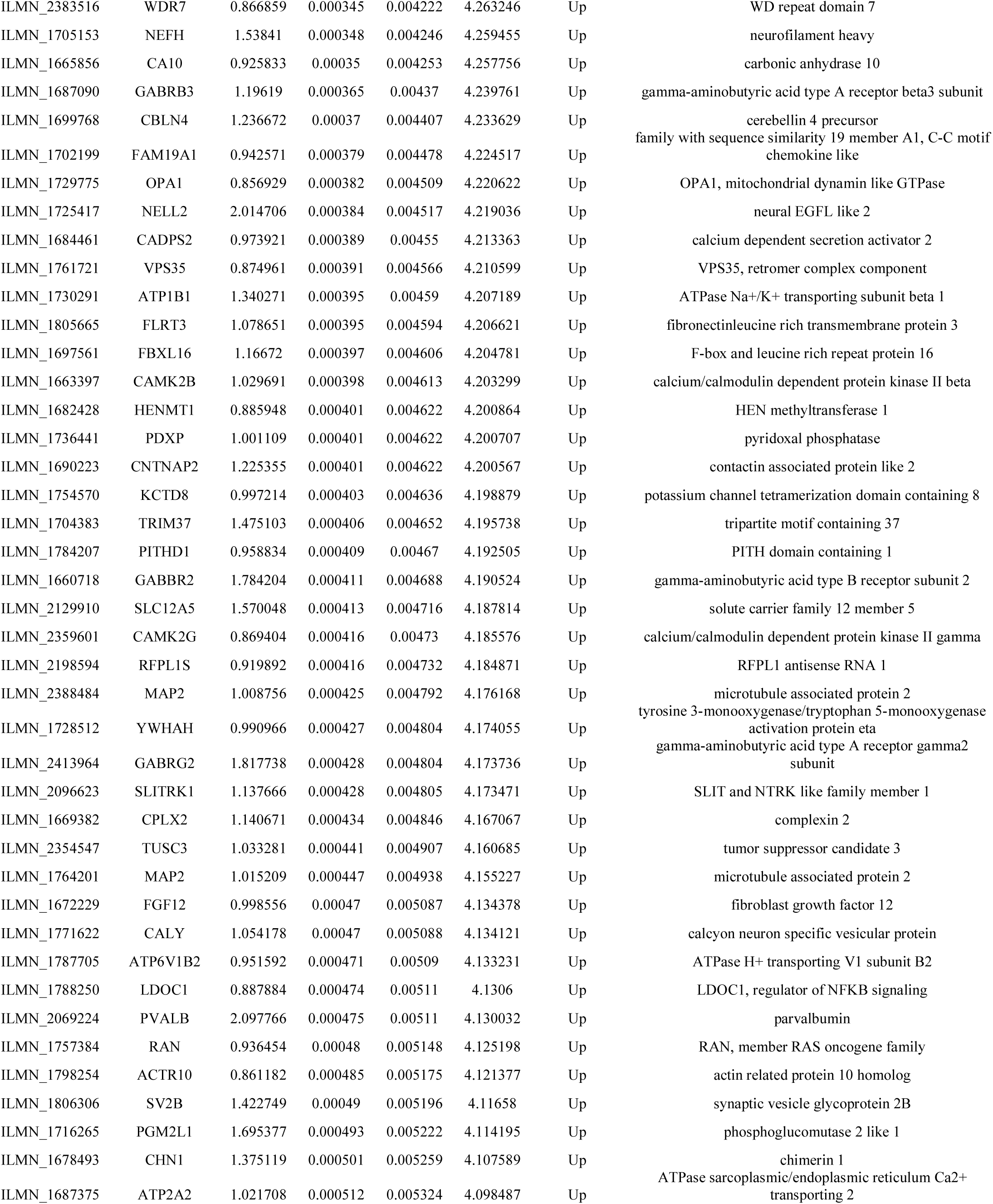
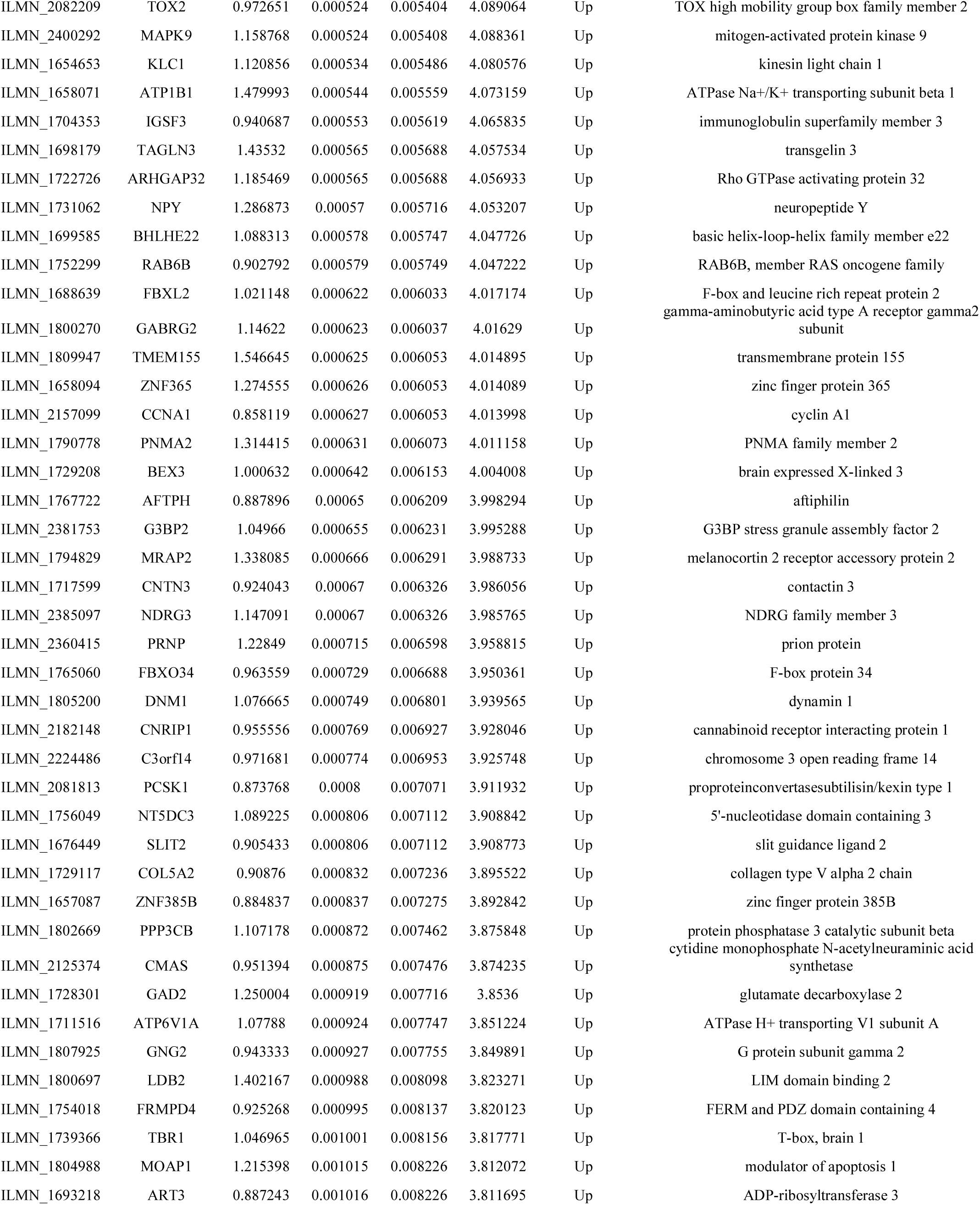
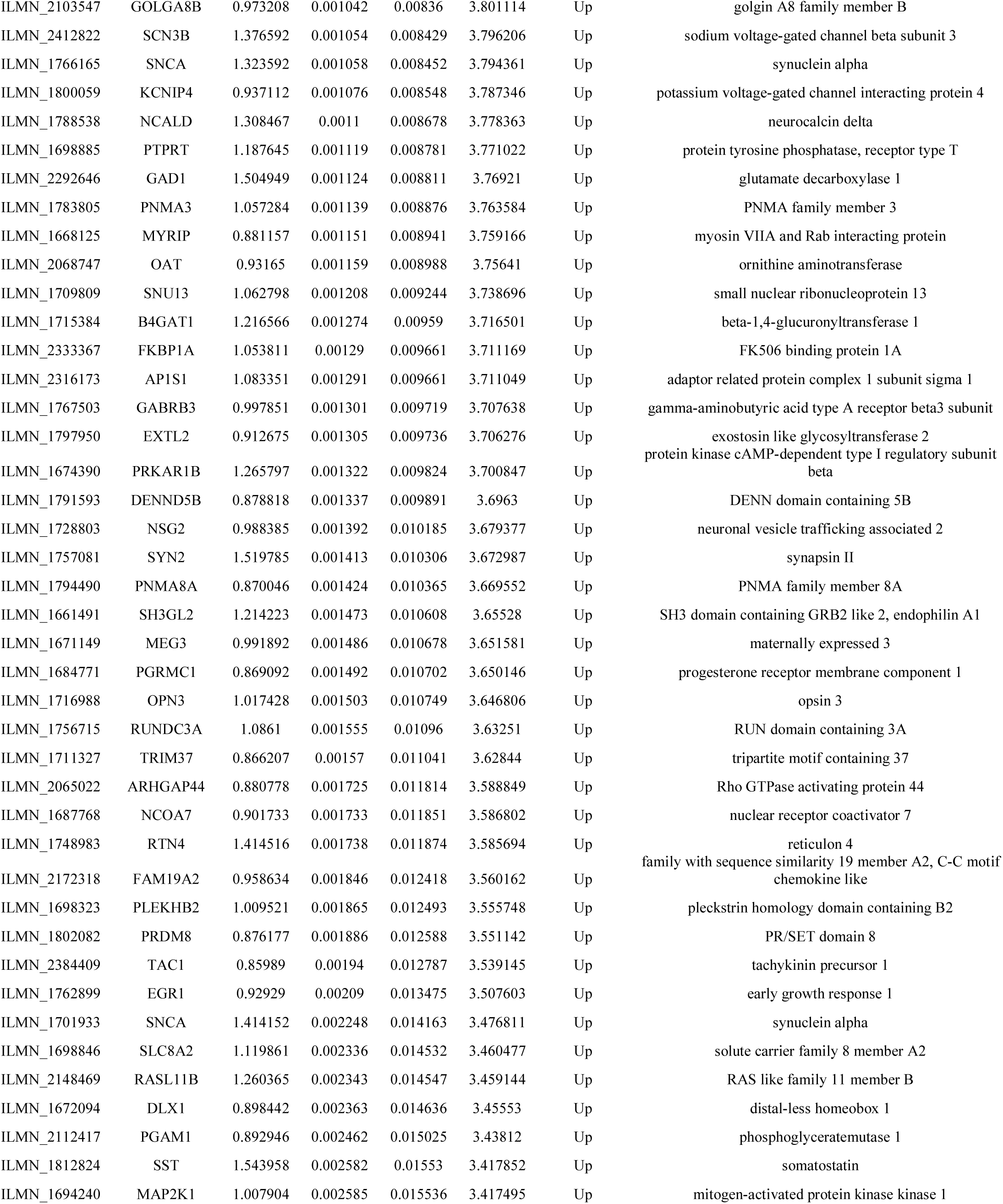
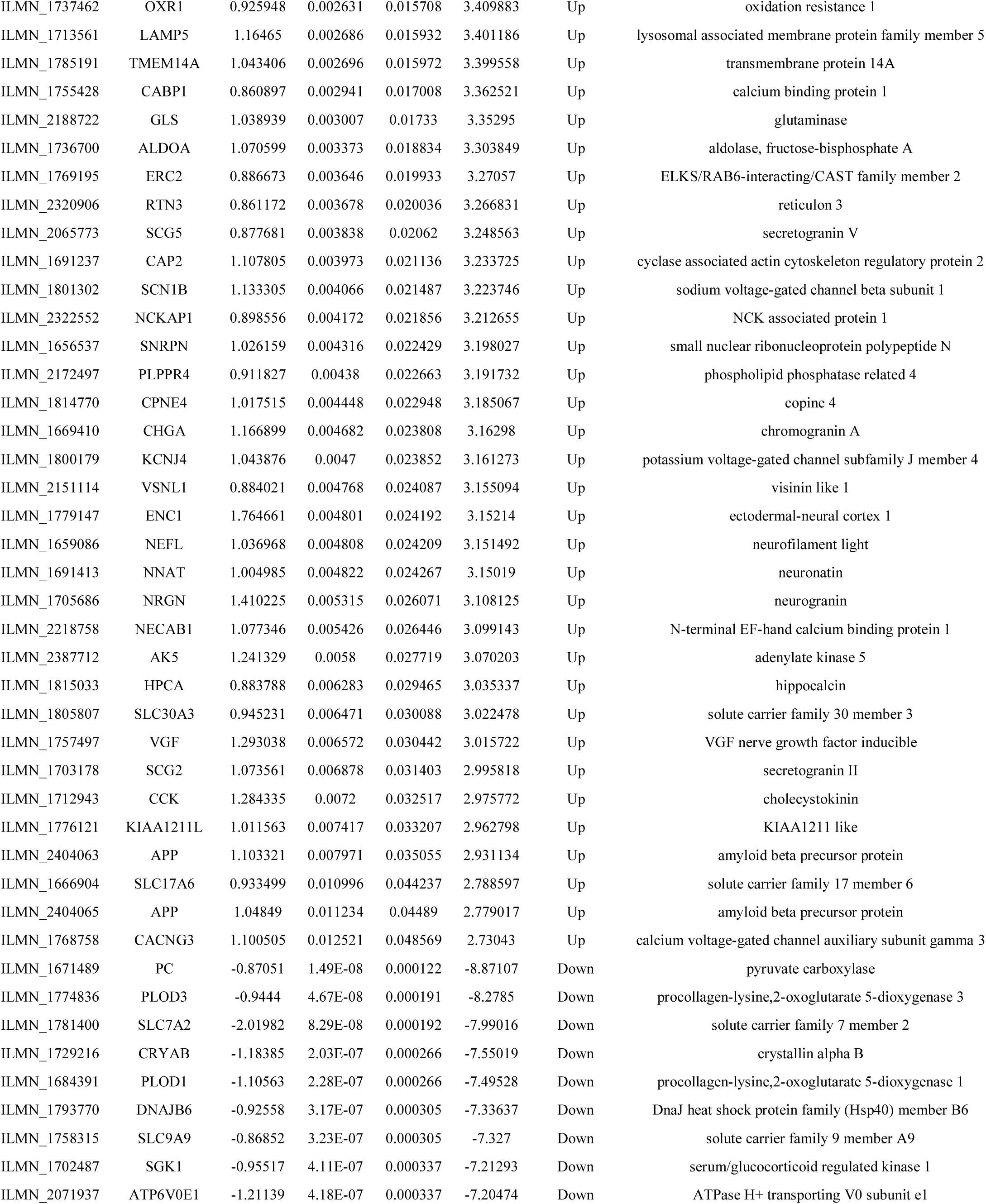
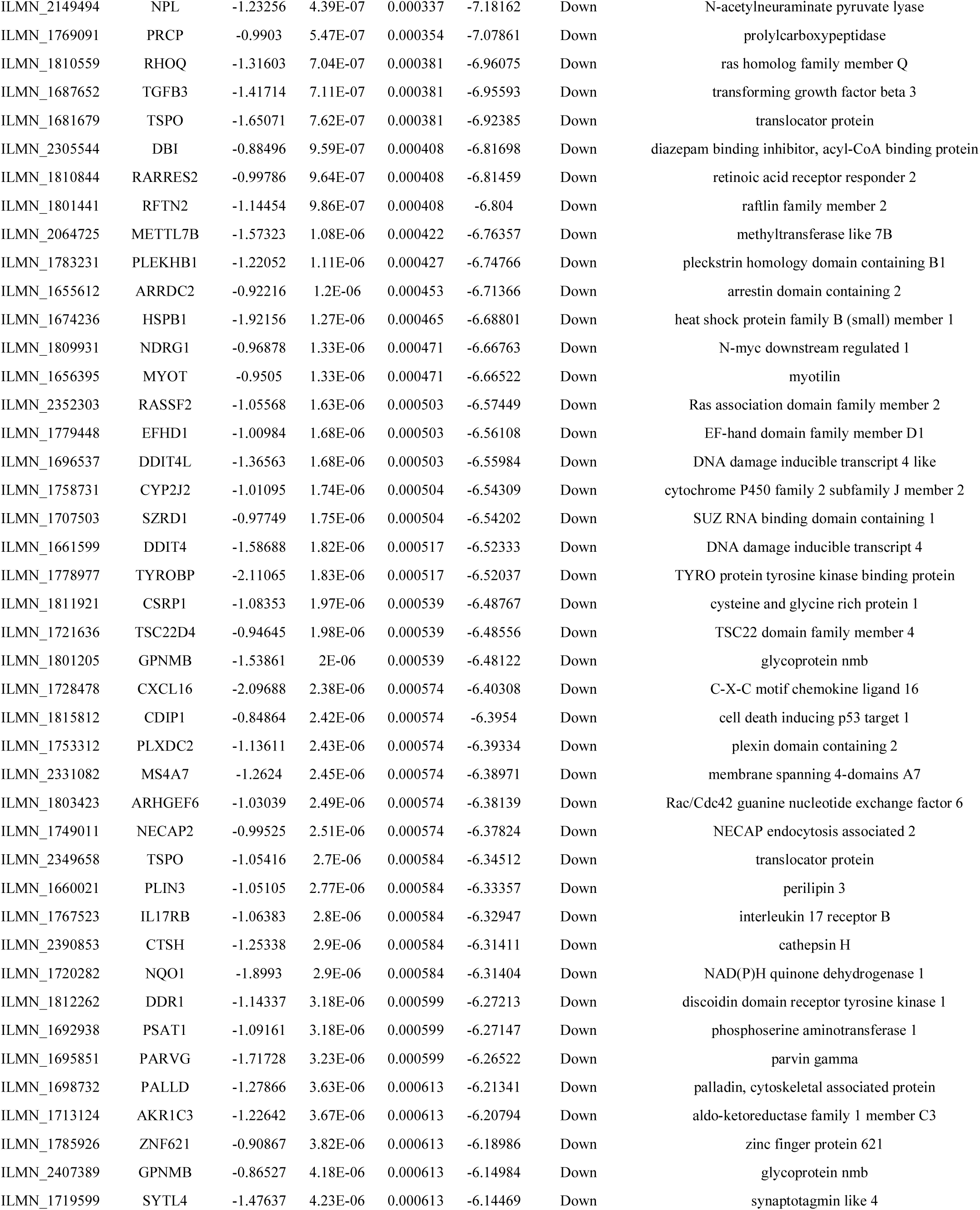
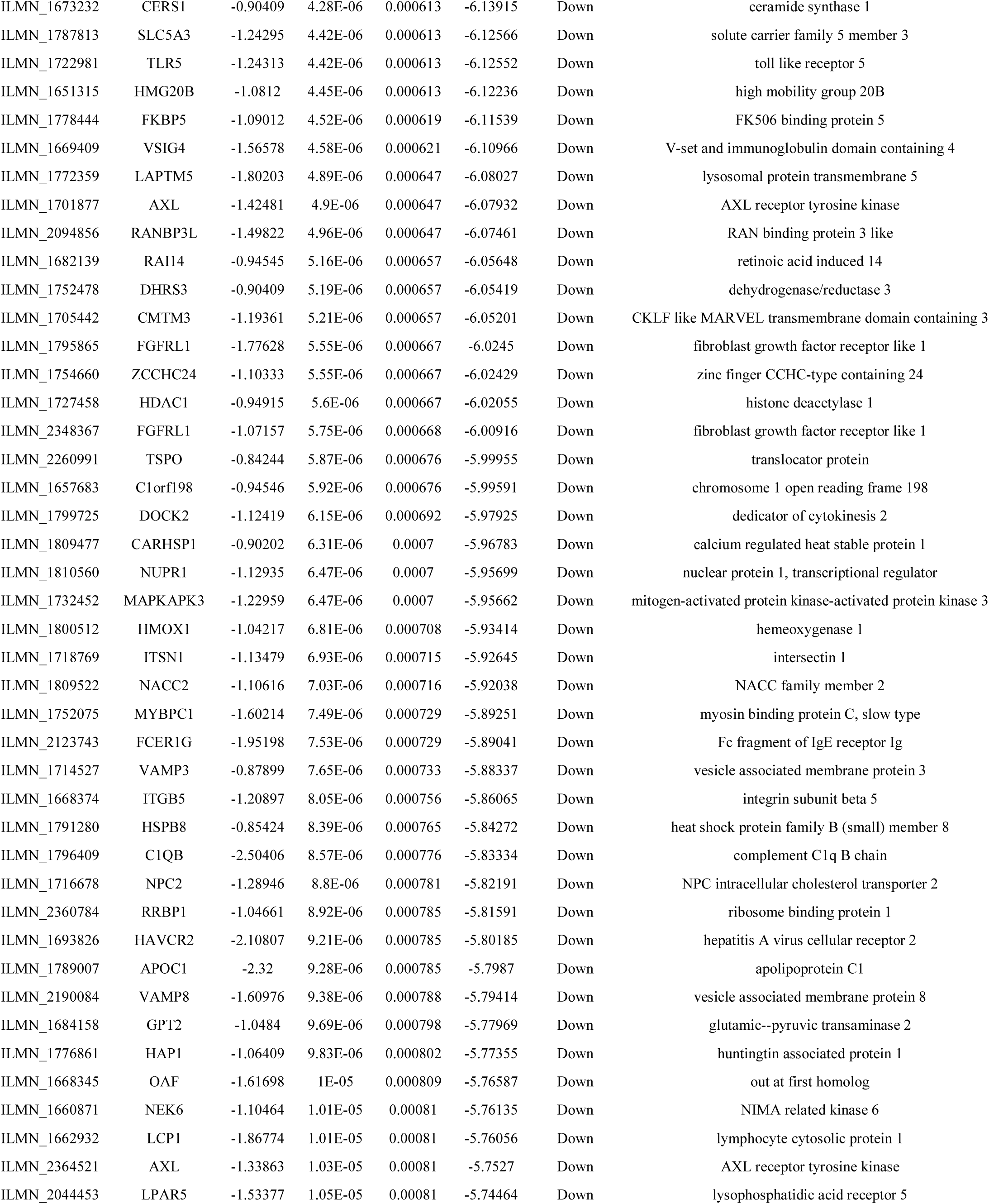
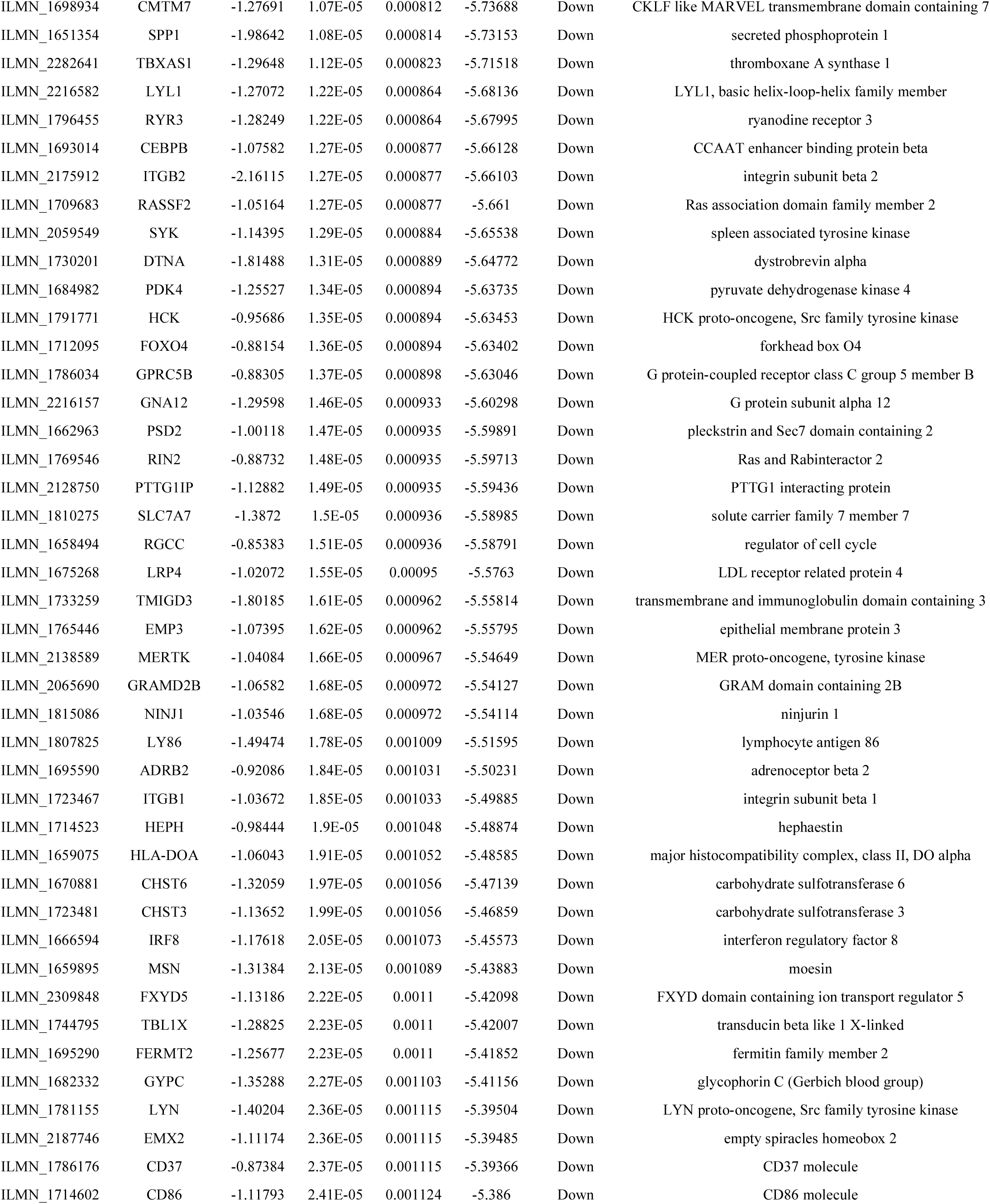
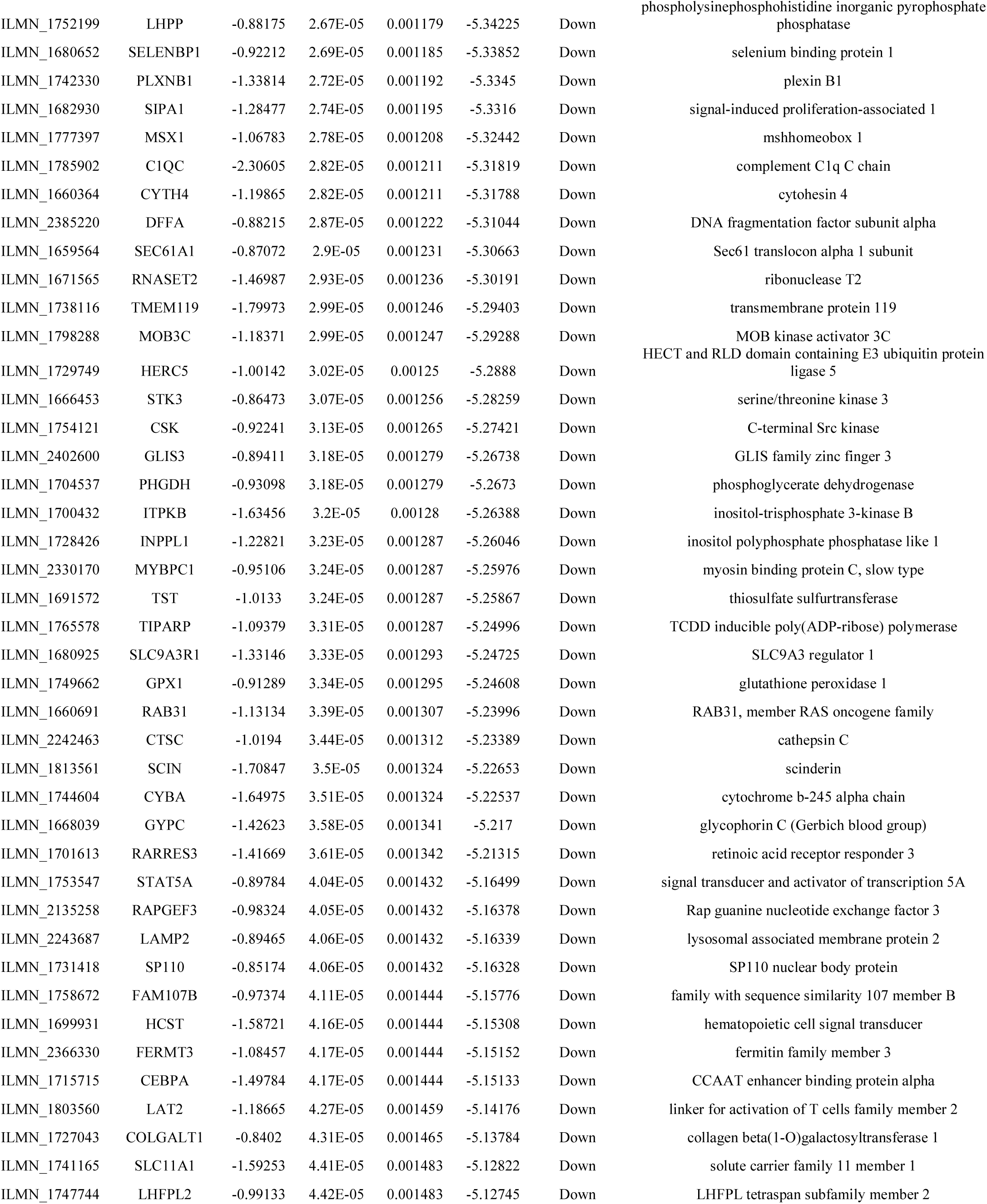
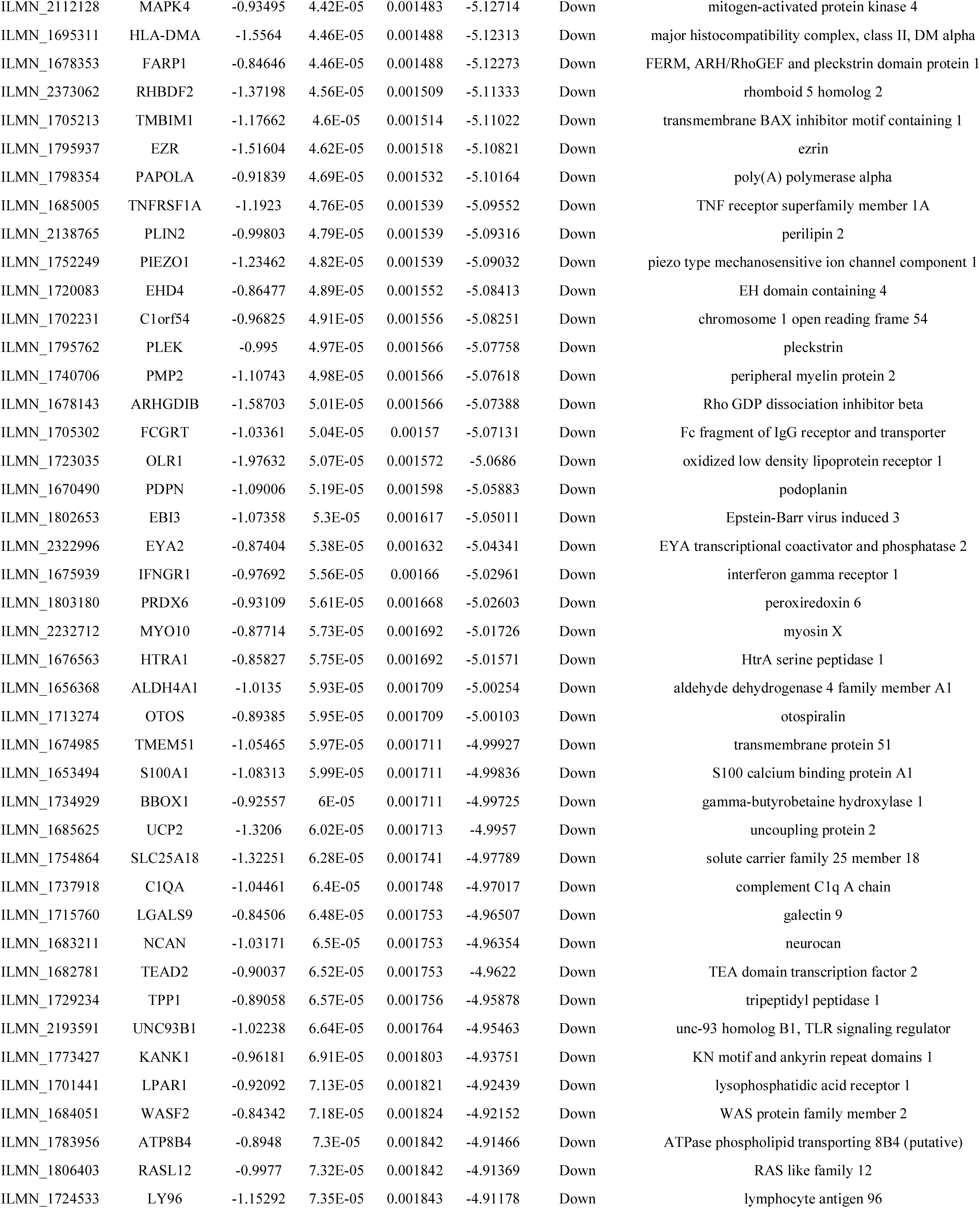
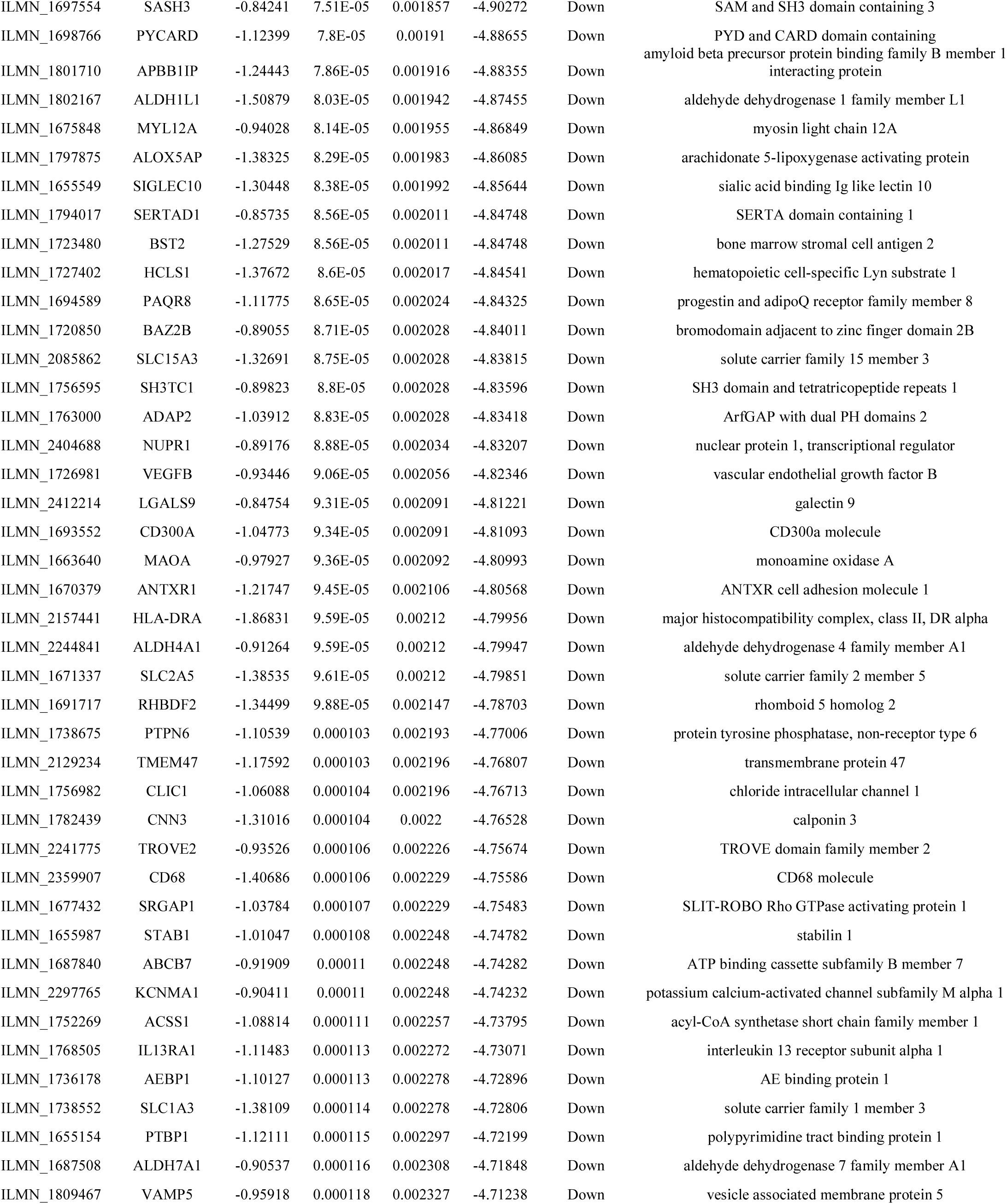
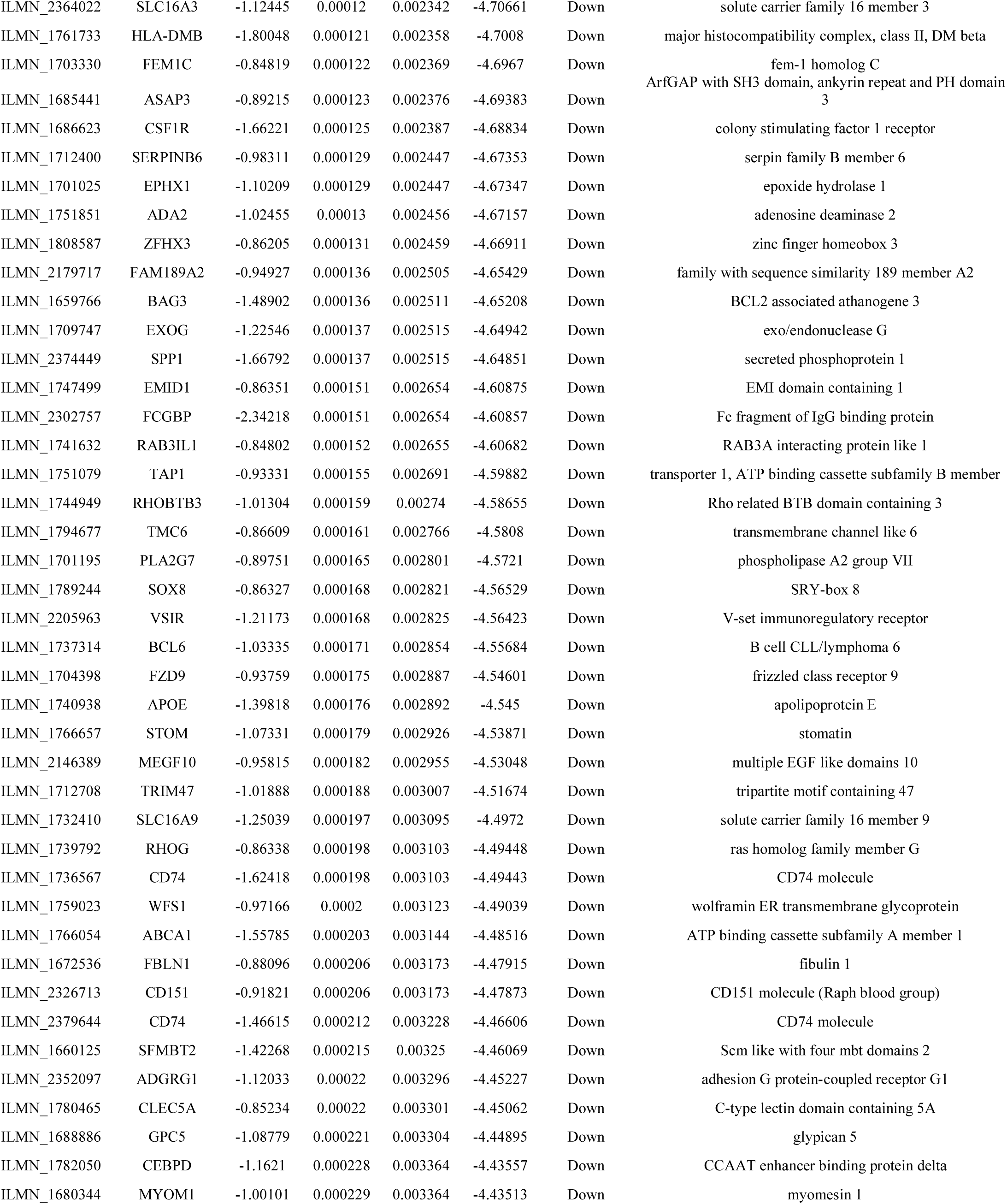
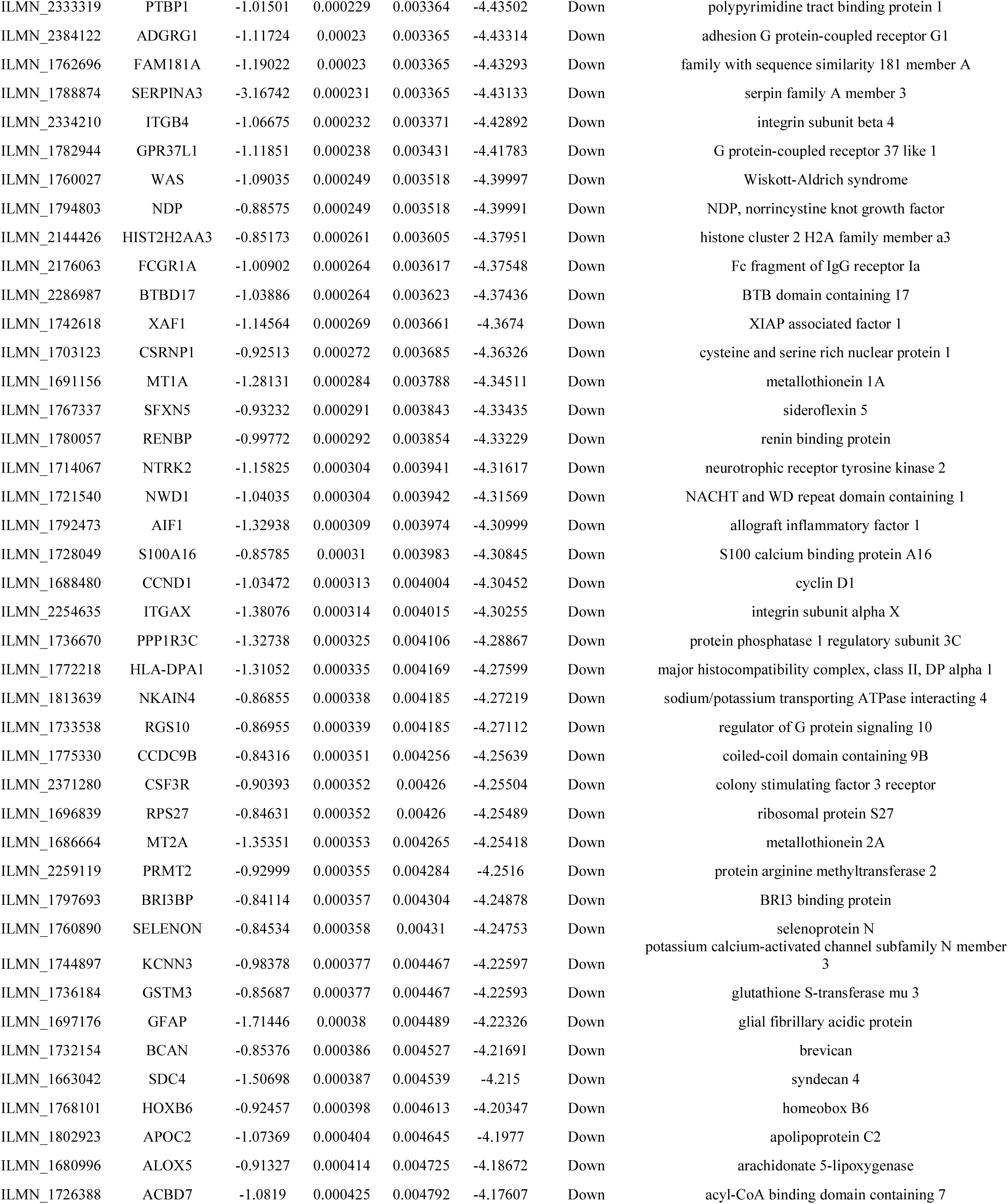
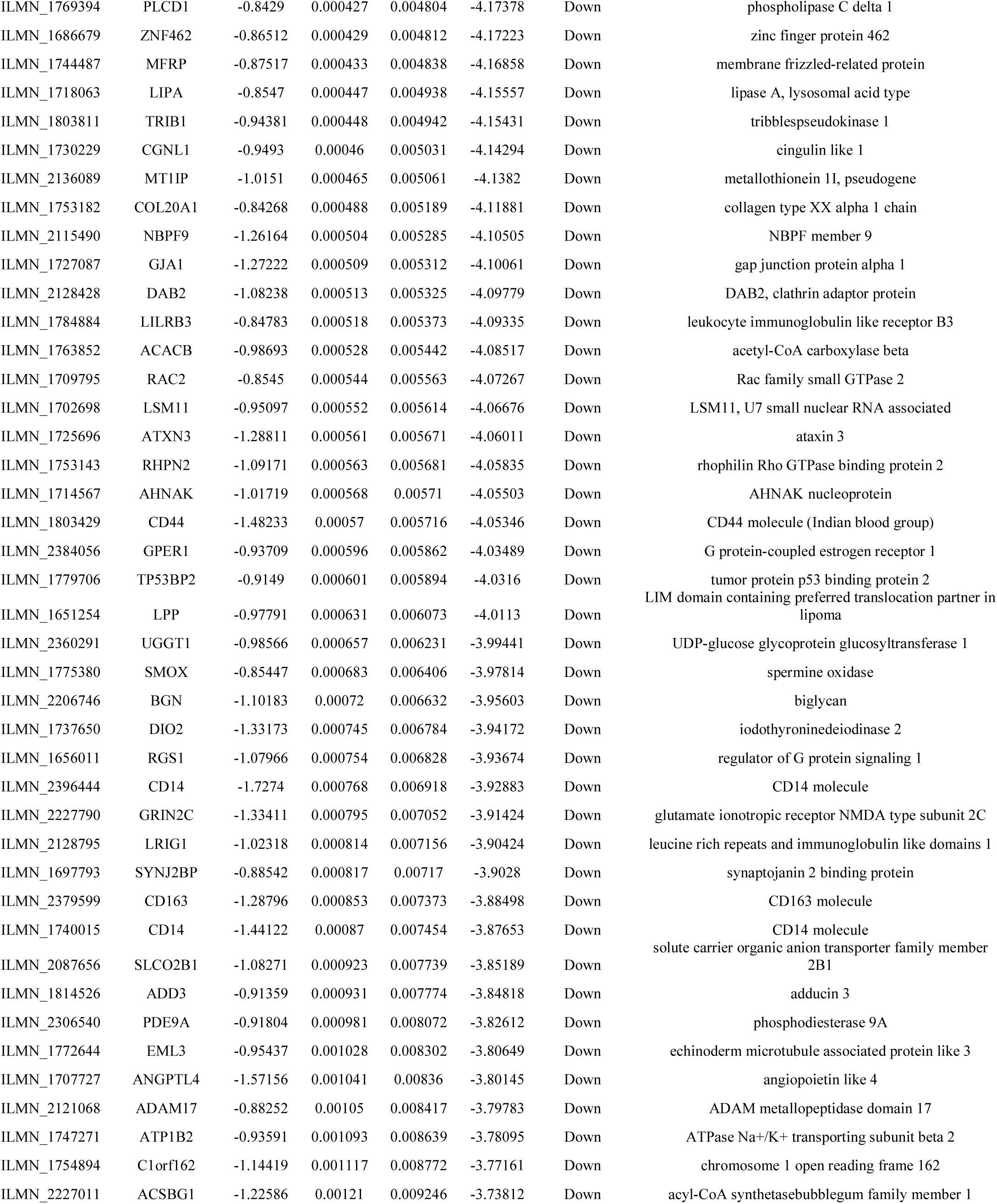
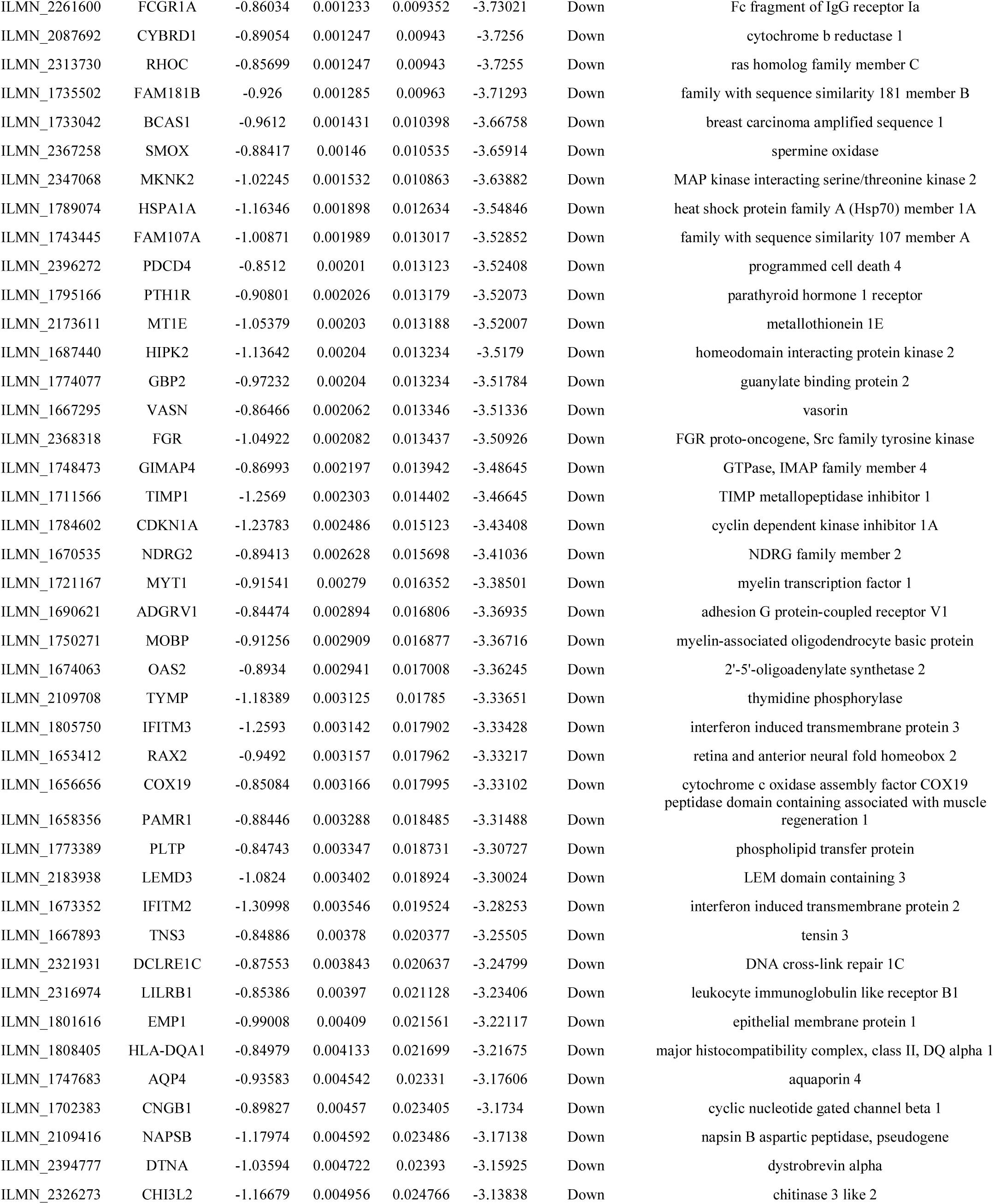
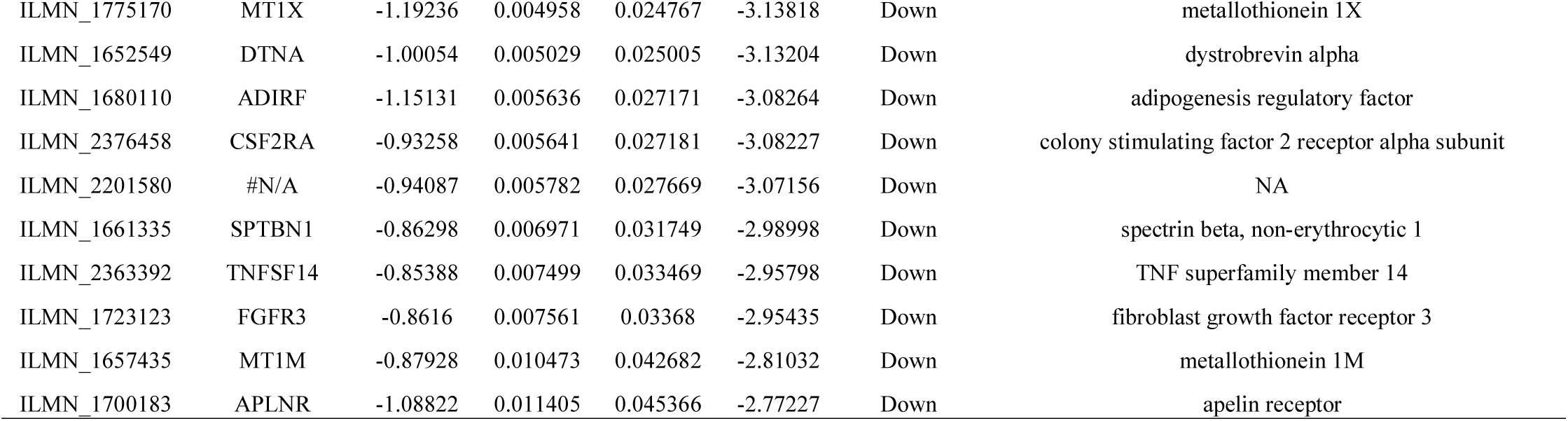
The statistical metrics for key differentially expressed genes (DEGs)

### Pathway enrichment analysis of DEGs

Pathway enrichment analysis of DEGs was conducted with ToppGene, and the results are given in Table 2 and Table 3. The signaling pathways of up regulated were mainly enriched in glutamine degradation/glutamate biosynthesis, asparagine biosynthesis, GABAergic synapse, synaptic vesicle cycle, effects of Botulinum toxin, role of calcineurin-dependent NFAT signaling in lymphocytes, transmission across chemical synapses, neuronal system, type III secretion system, ATP synthesis, regulation of PGC-1a, bioactive peptide induced signaling pathway, muscarinic acetylcholine receptor 1 and 3 signaling pathway, oxytocin receptor mediated signaling pathway, glutamate metabolic, insulin secretion pathway, cimetidine pathway and malate-aspartate shuttle, whereas down regulated were mainly enriched in melatonin degradation II, proline degradation, leishmaniasis, phagosome, validated targets of C-MYC transcriptional repression, CXCR4-mediated signaling events, neutrophil degranulation, cytokine signaling in immune system, inositol phosphate metabolism, monocyte and its surface molecules, cystic fibrosis transmembrane conductance regulator and beta 2 adrenergic receptor pathway, T cell activation, 5-hydroxytryptamine degredation, integrin signaling, altered lipoprotein metabolic, lactic acidemia and lysosomal acid lipase deficiency (Wolman Disease).

**Table 2.**
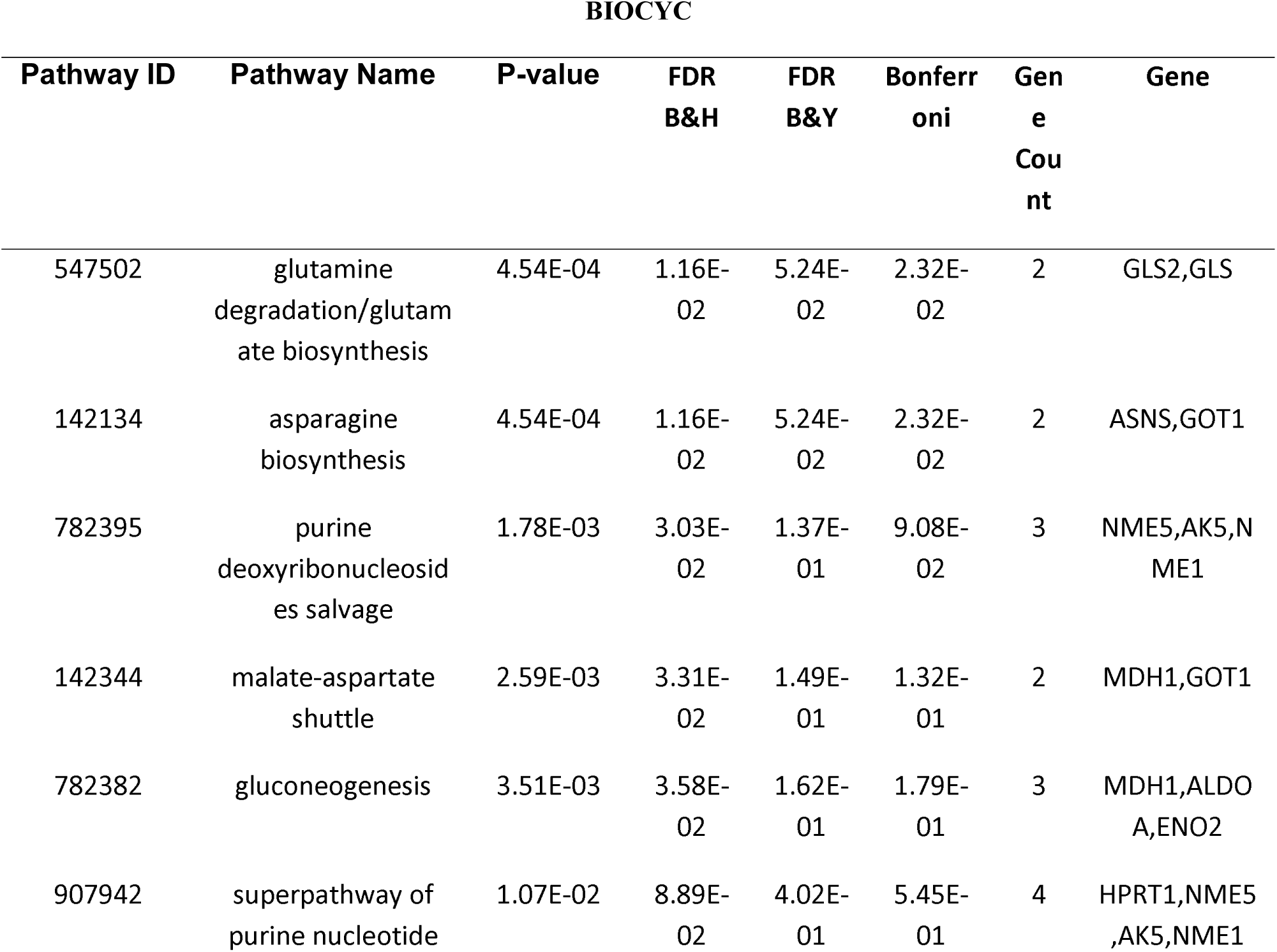
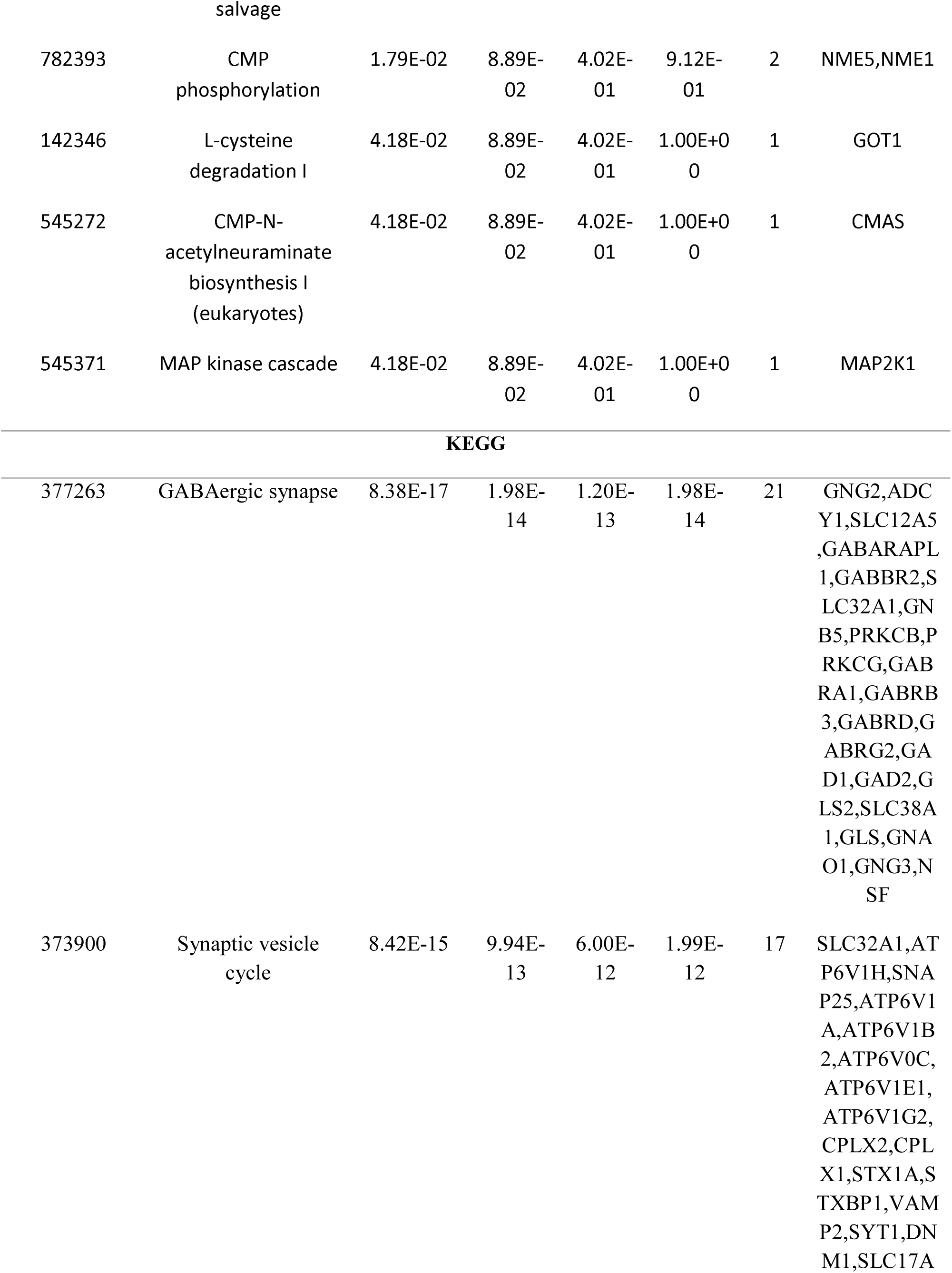
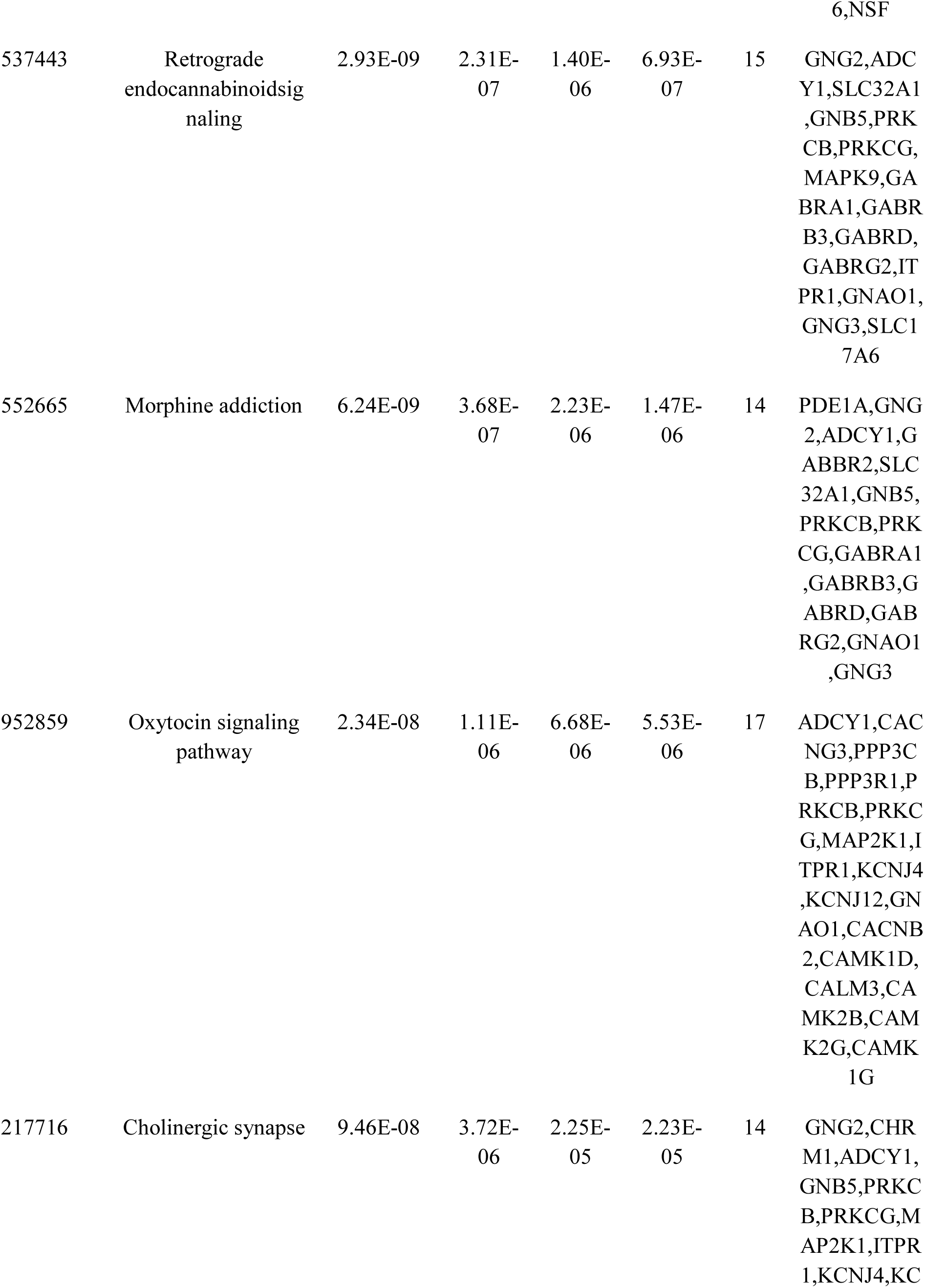
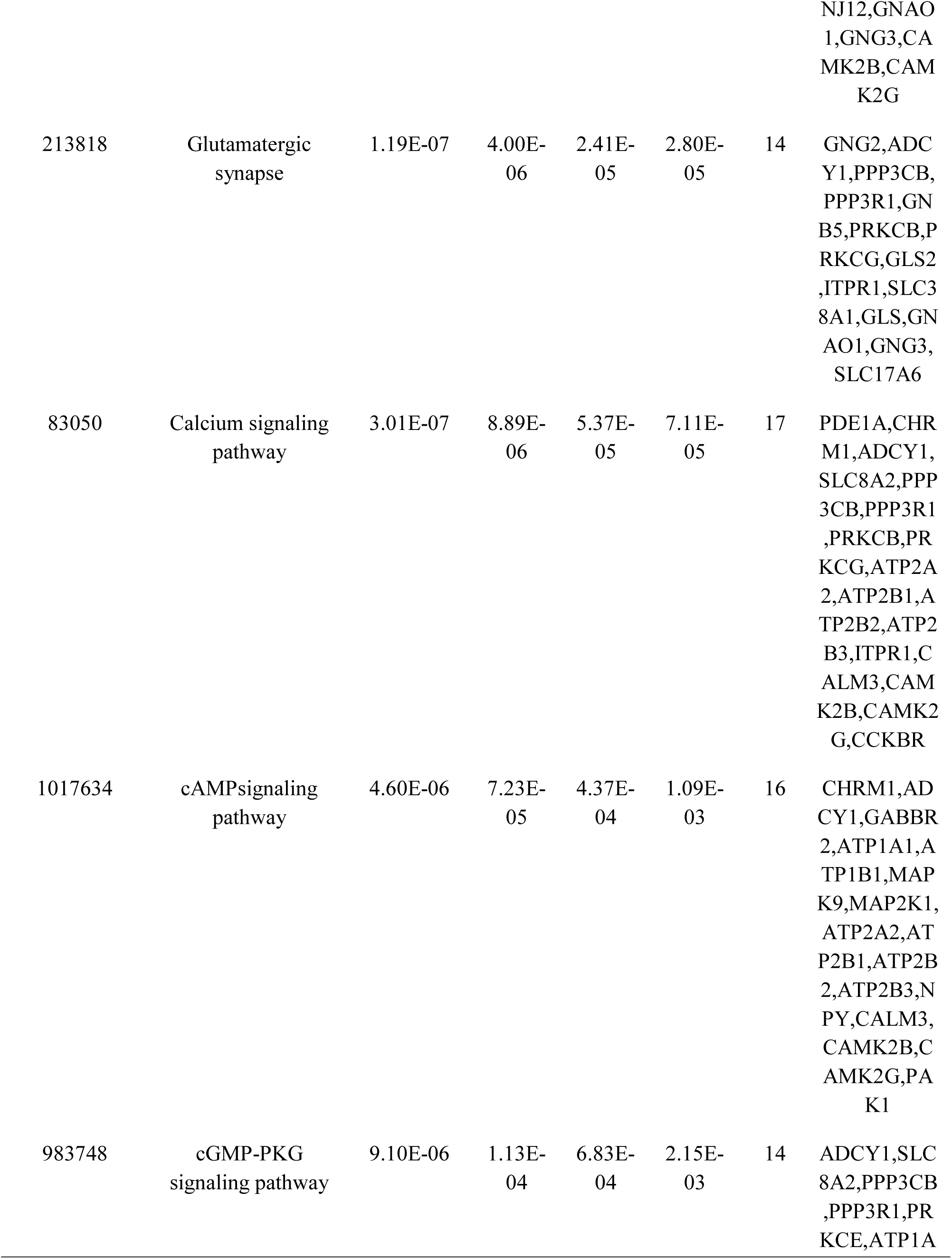
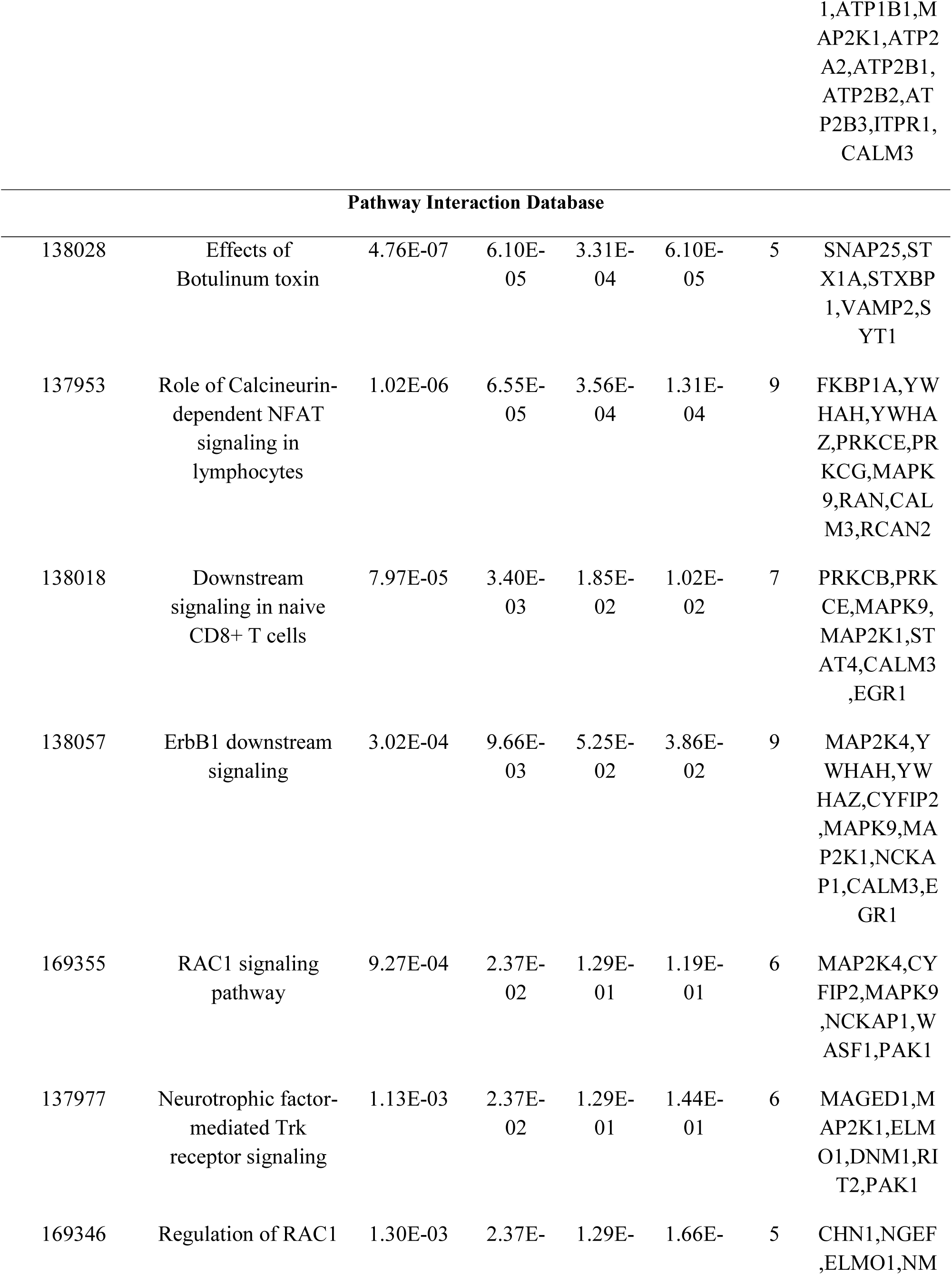
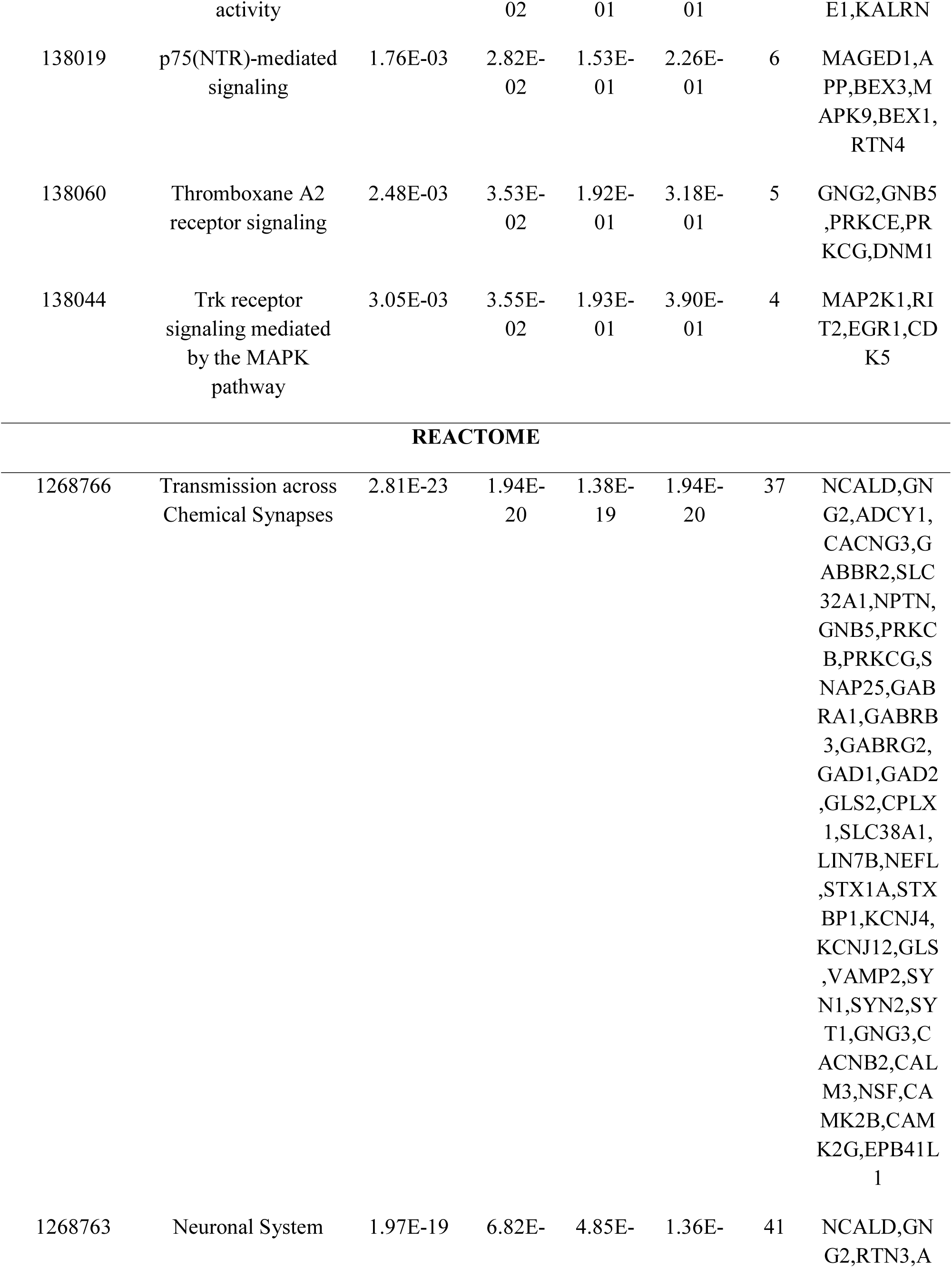
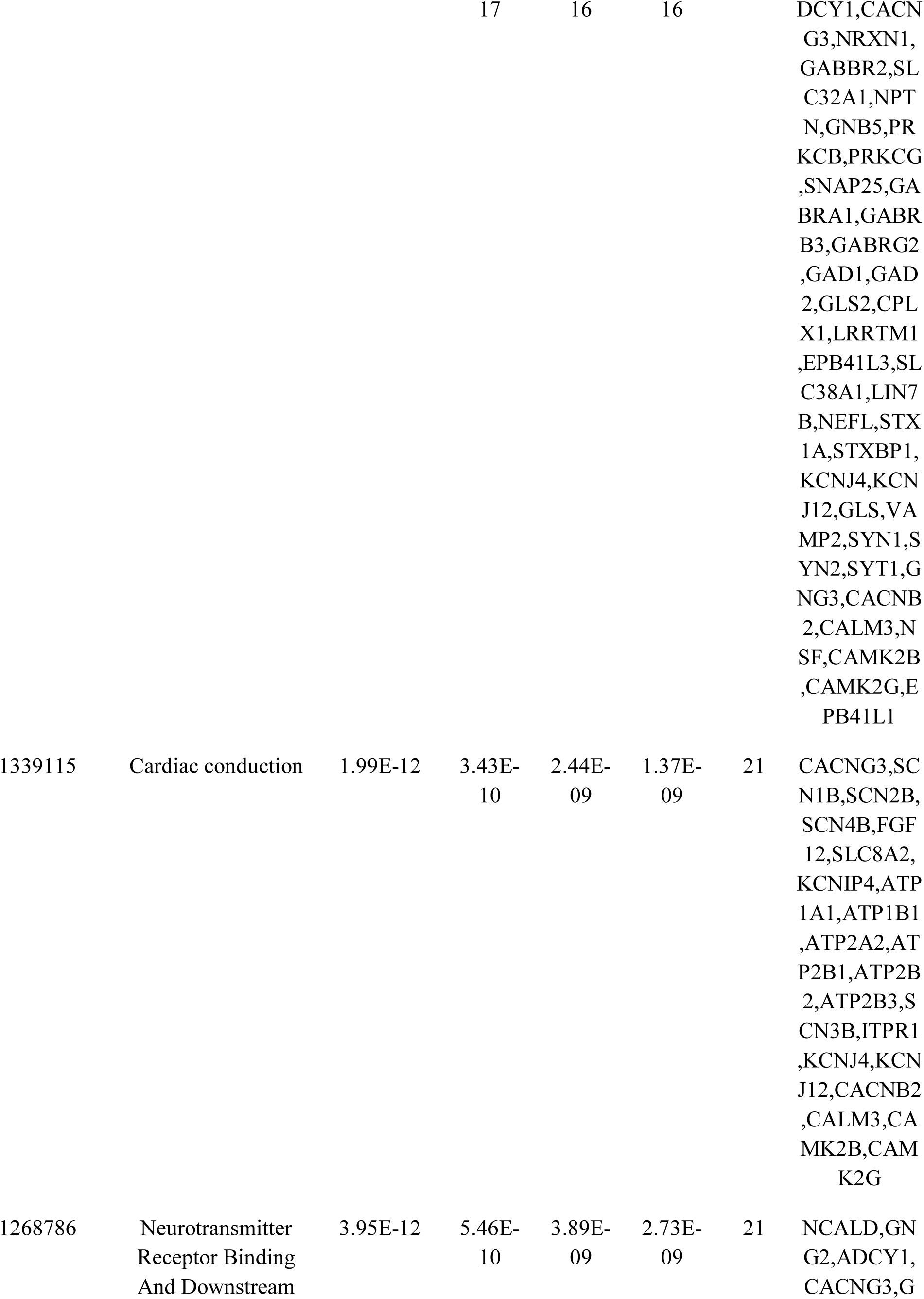
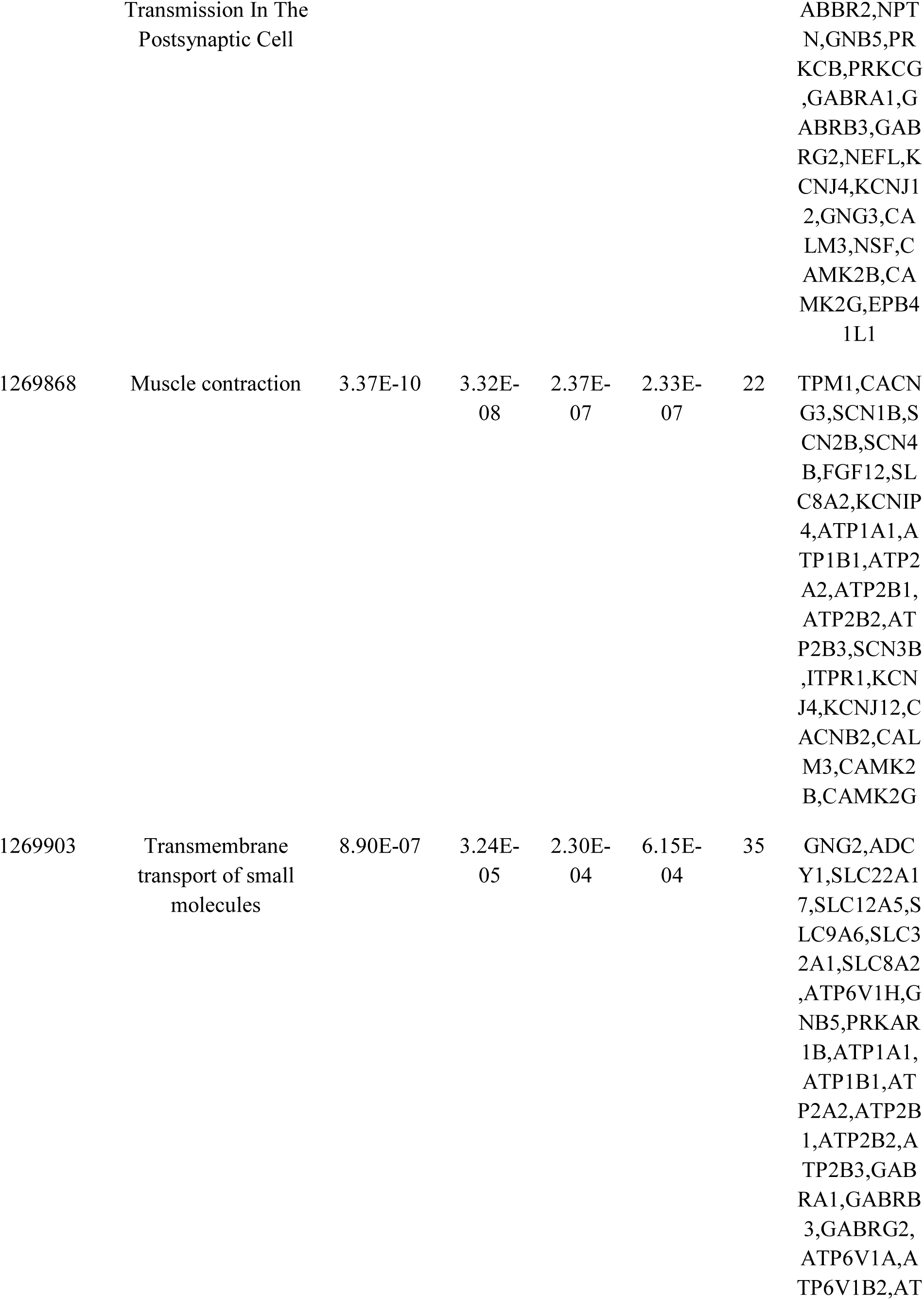
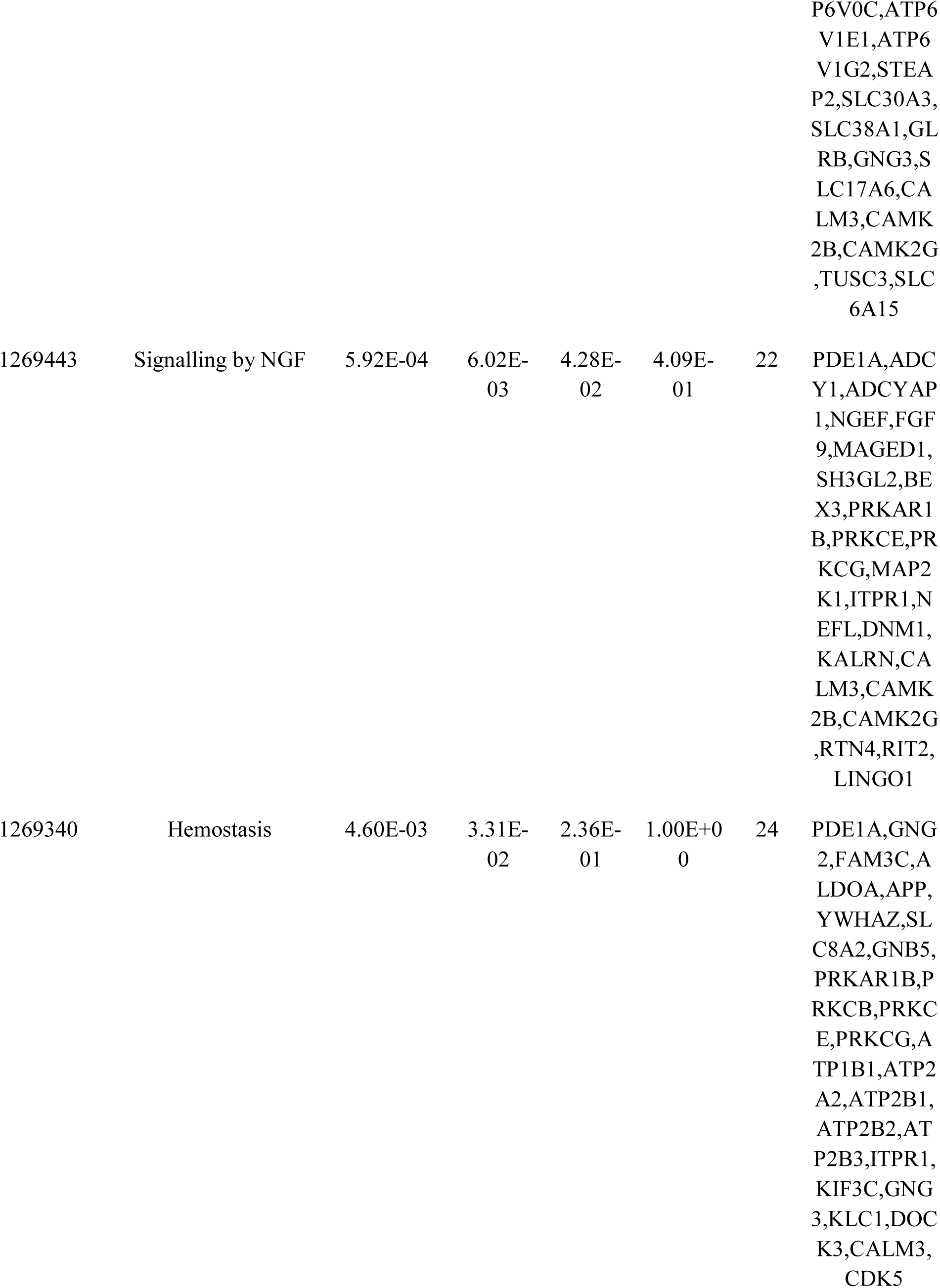
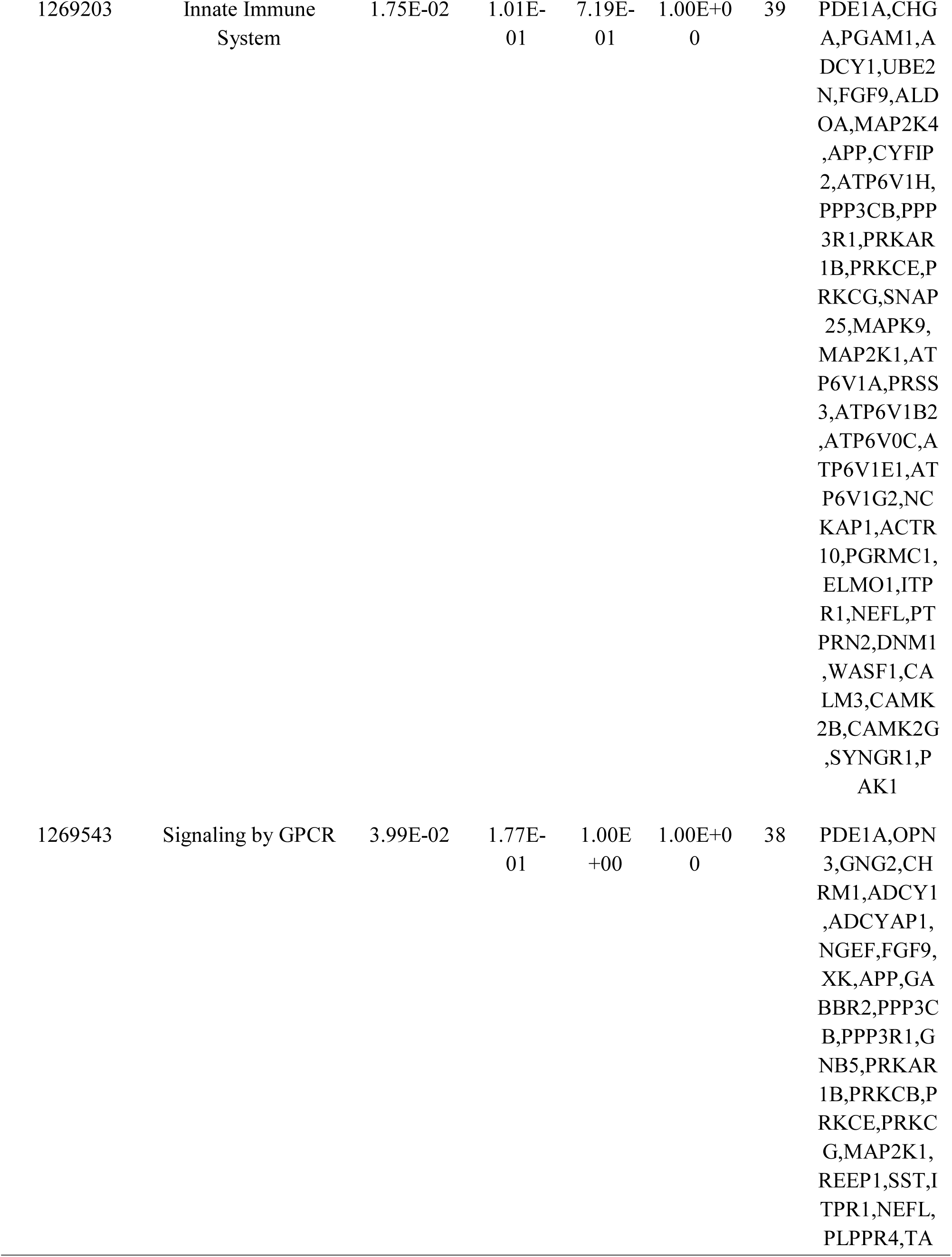
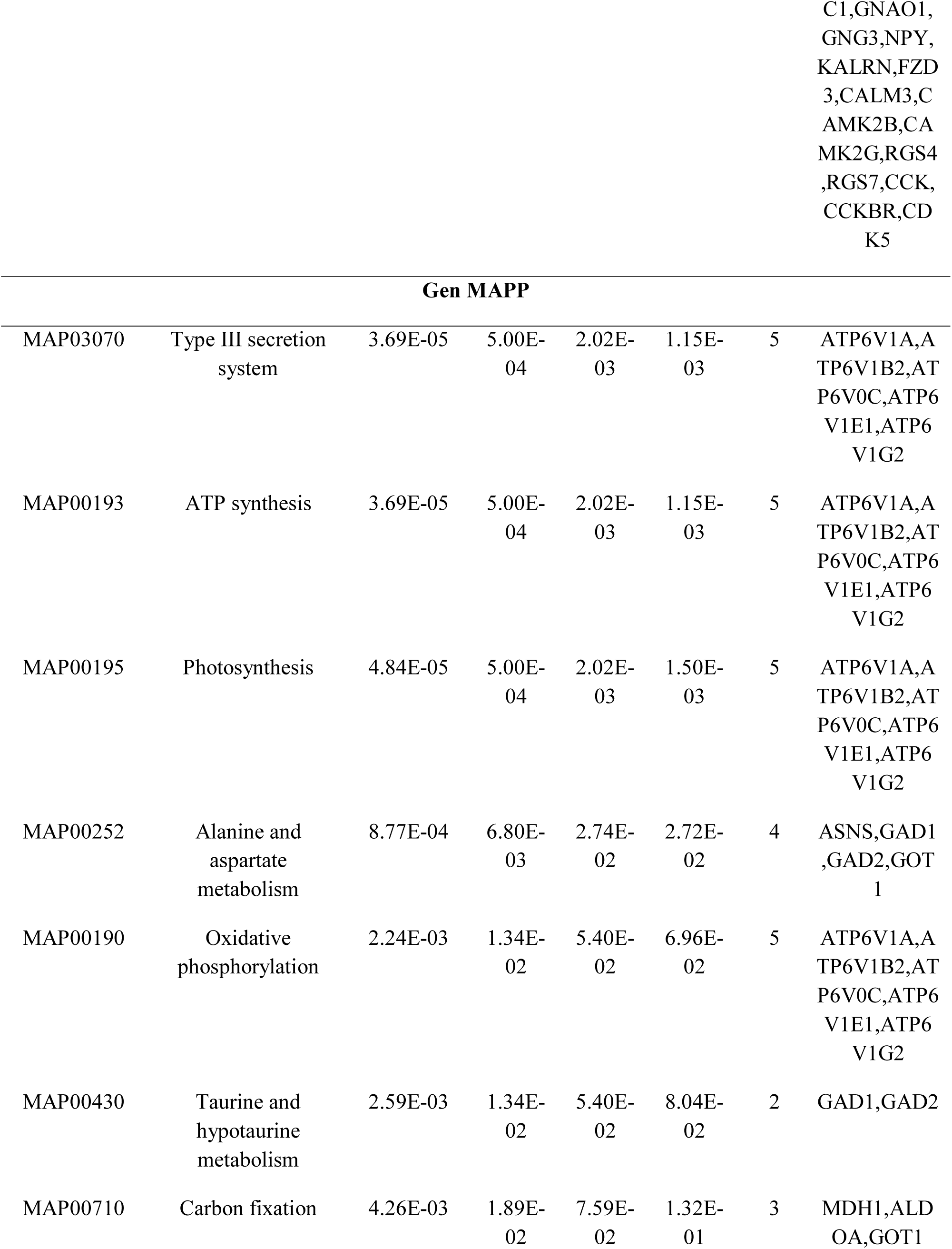
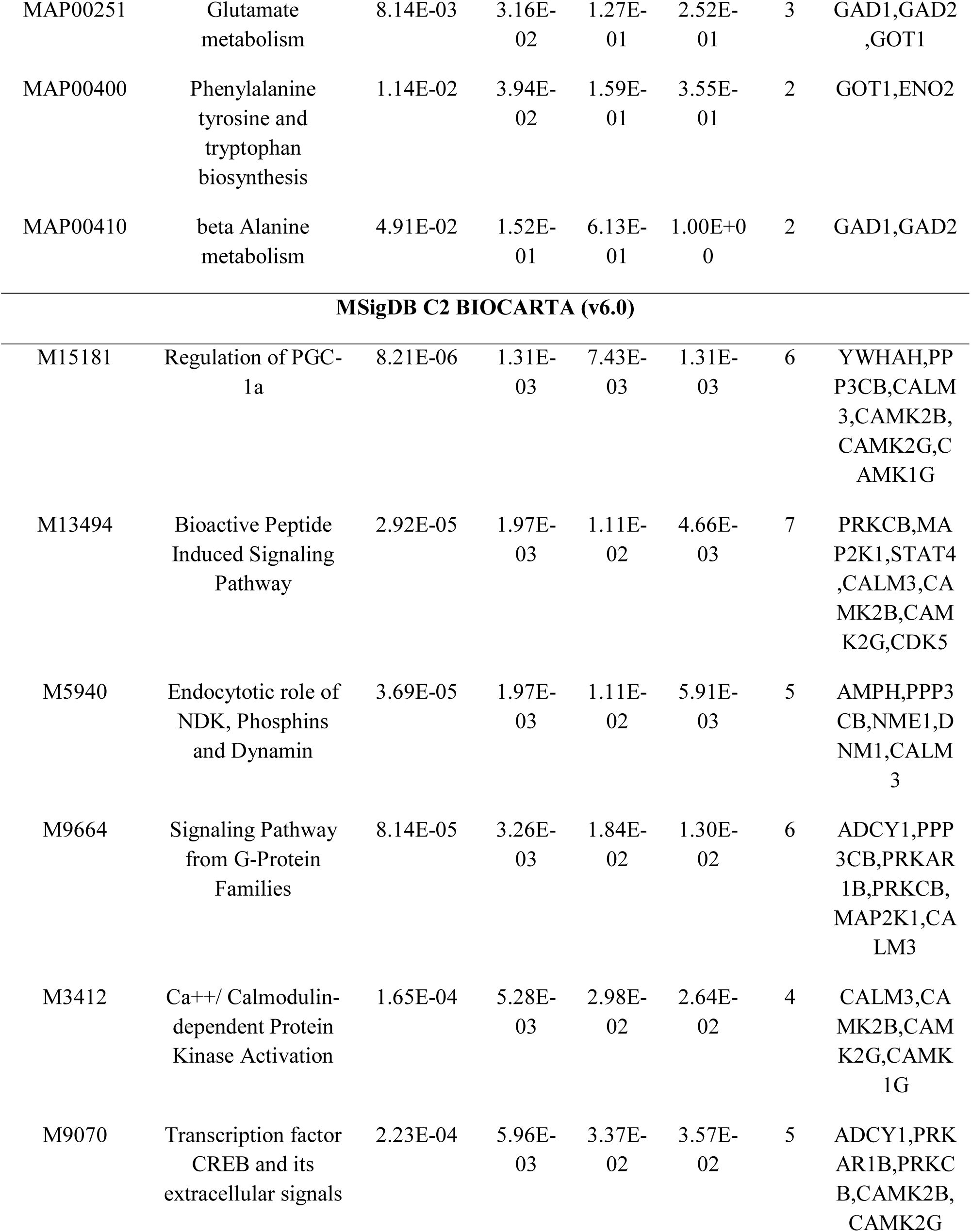
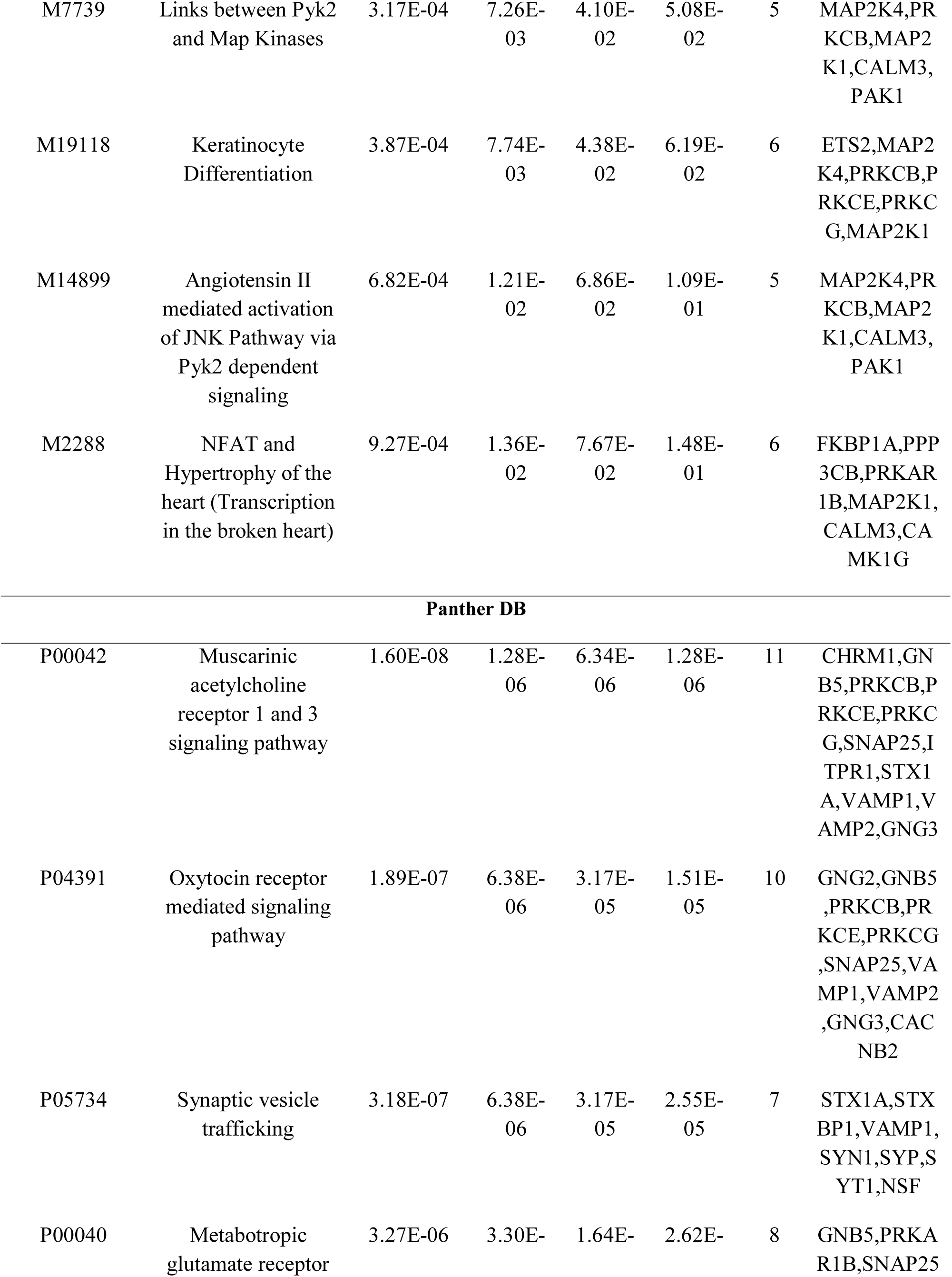
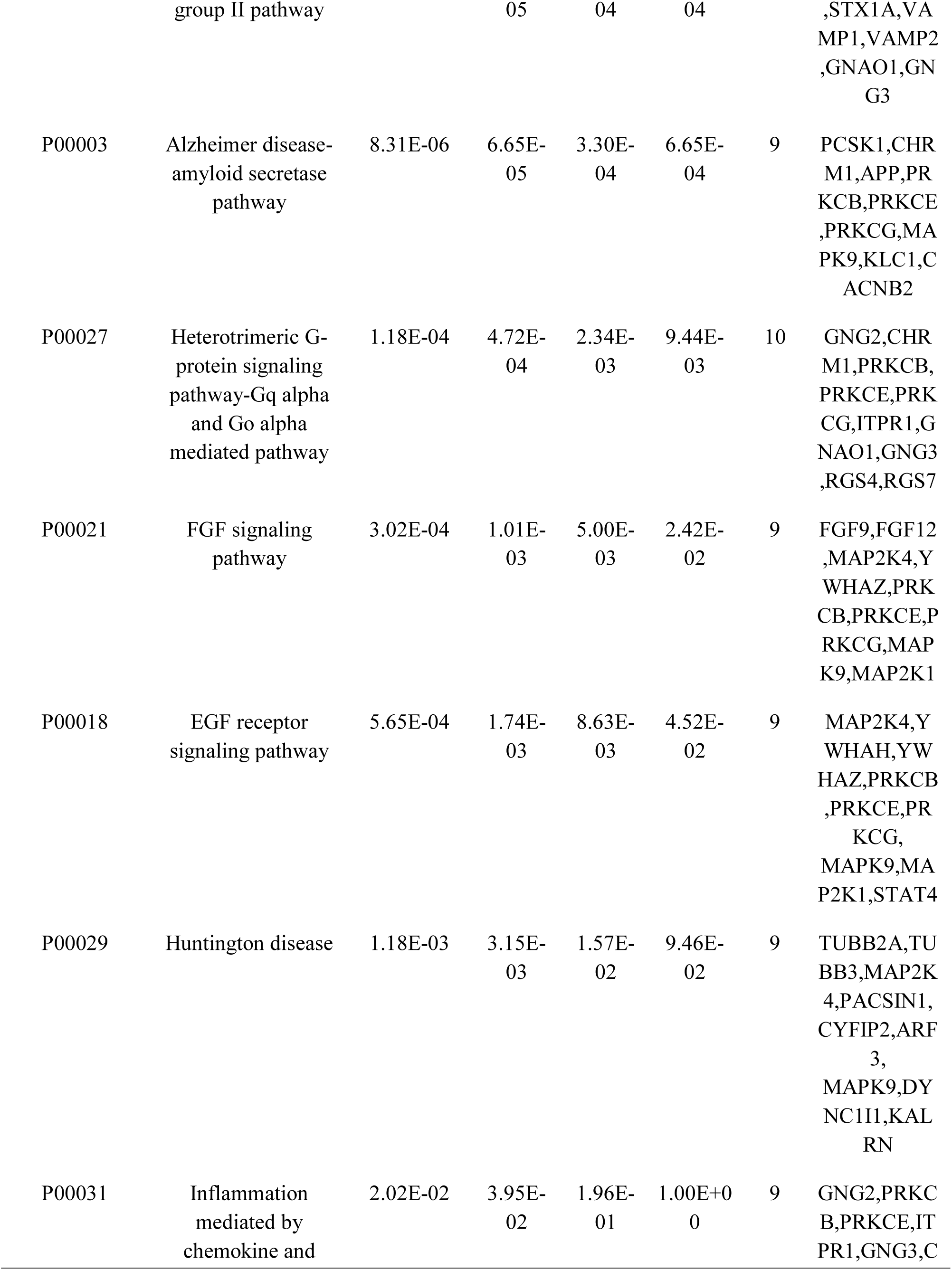
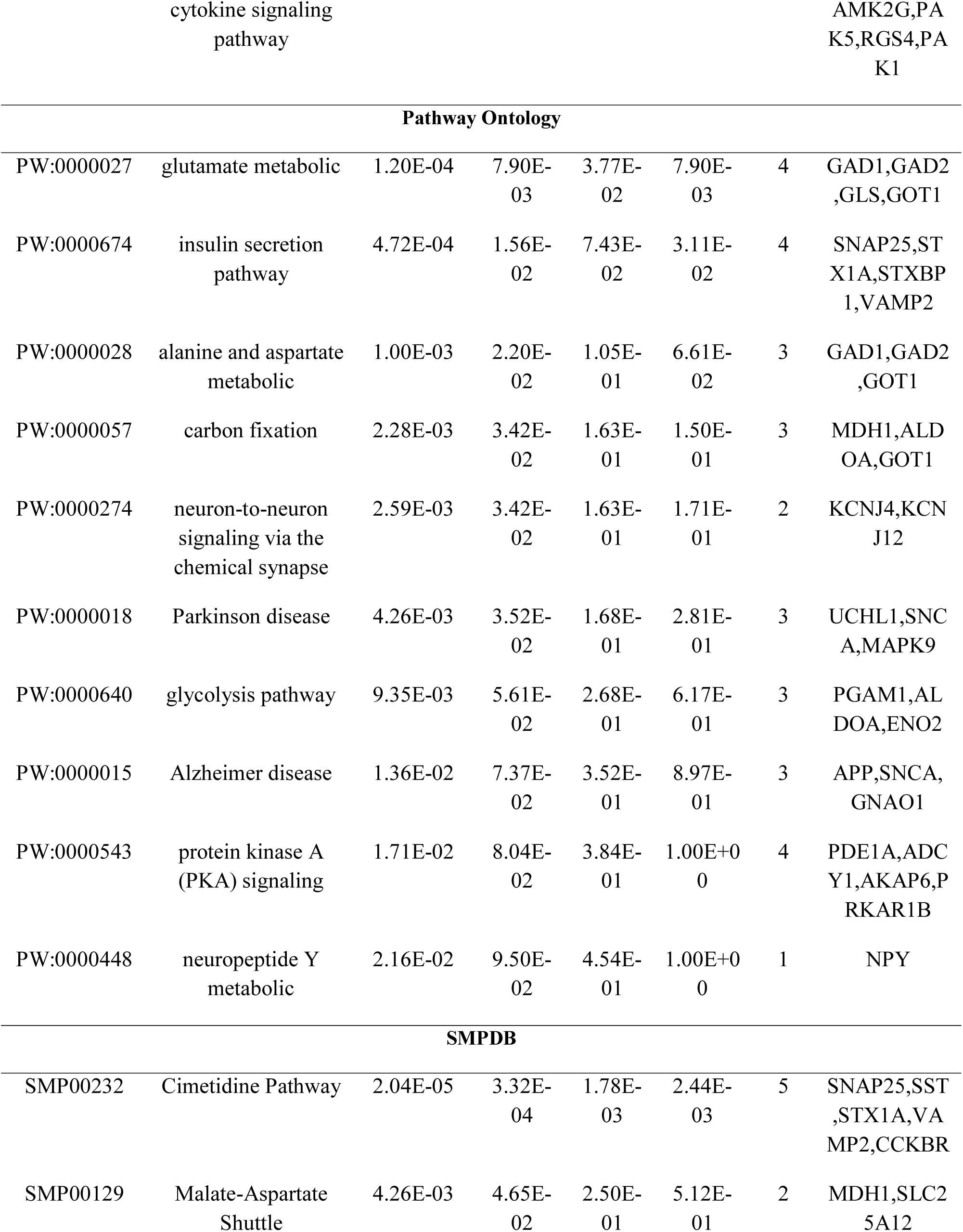
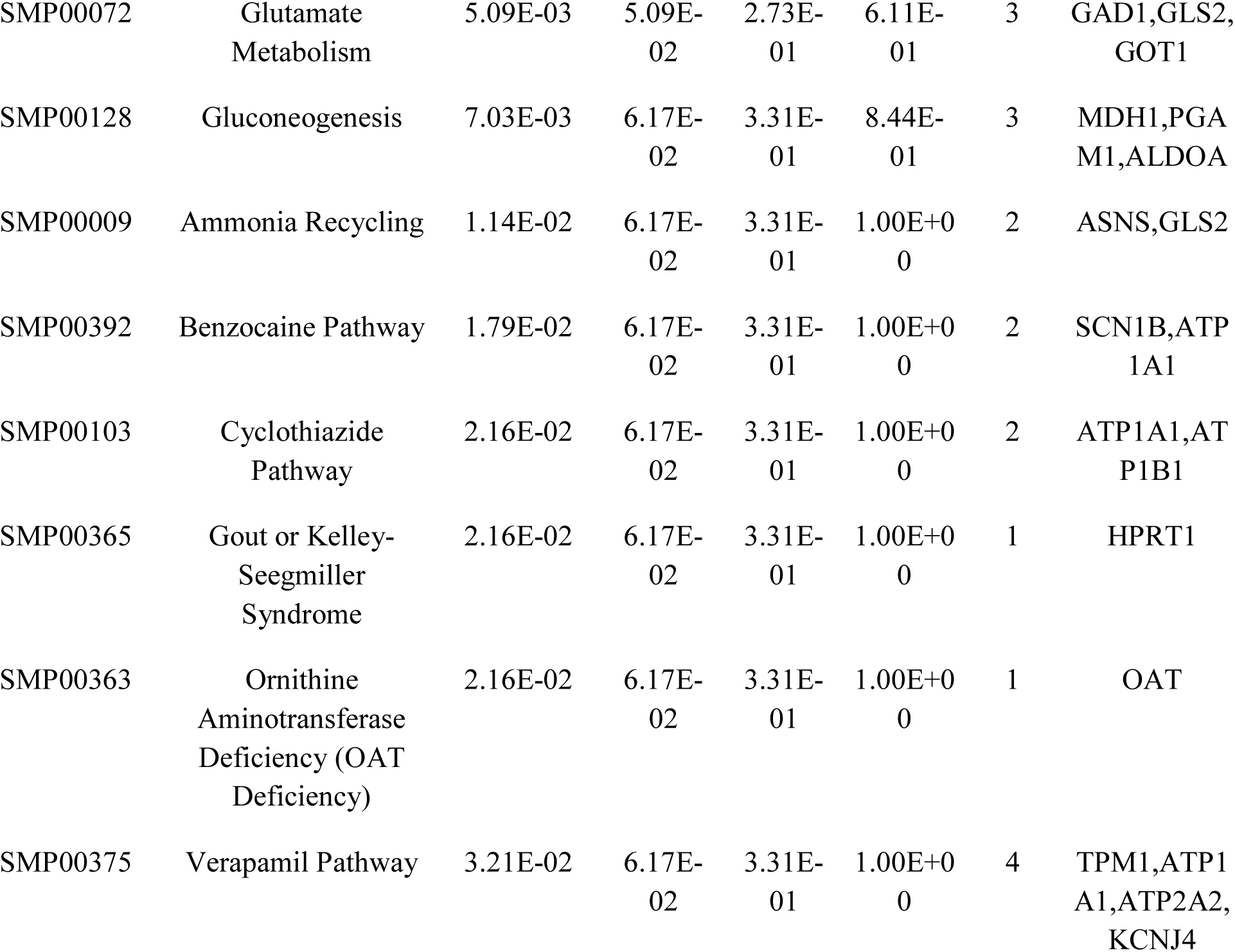
The enriched pathway terms of the up regulated differentially expressed genes

**Table 3.**
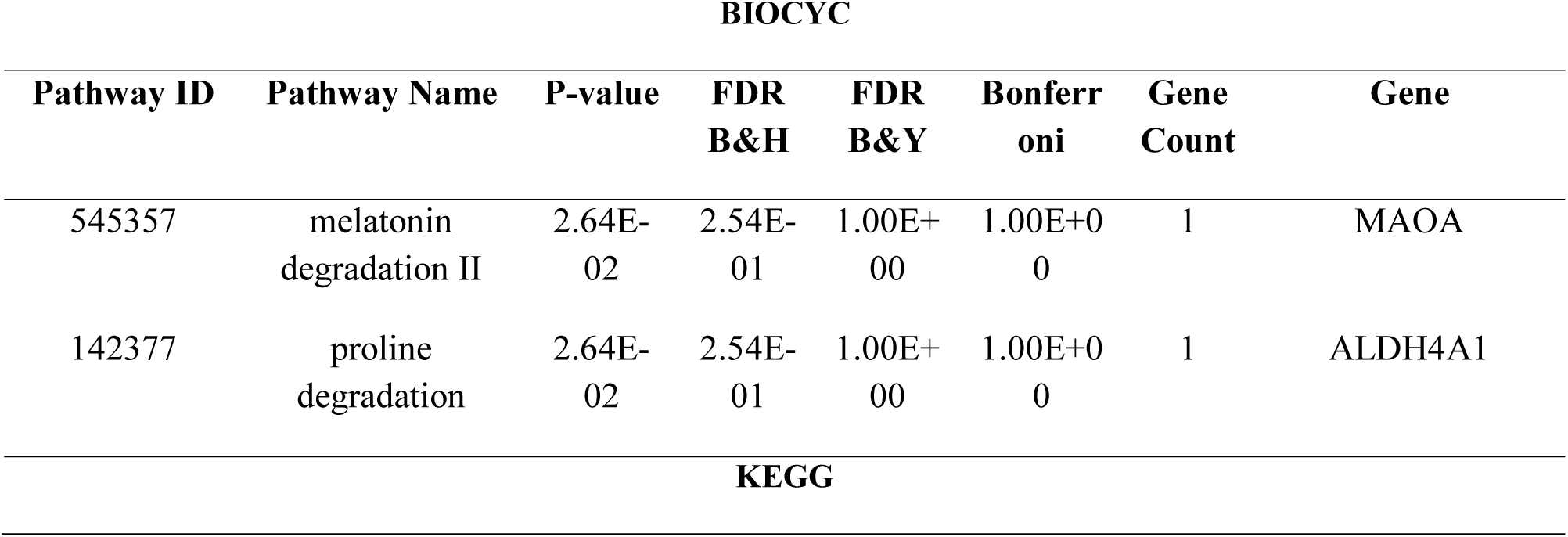
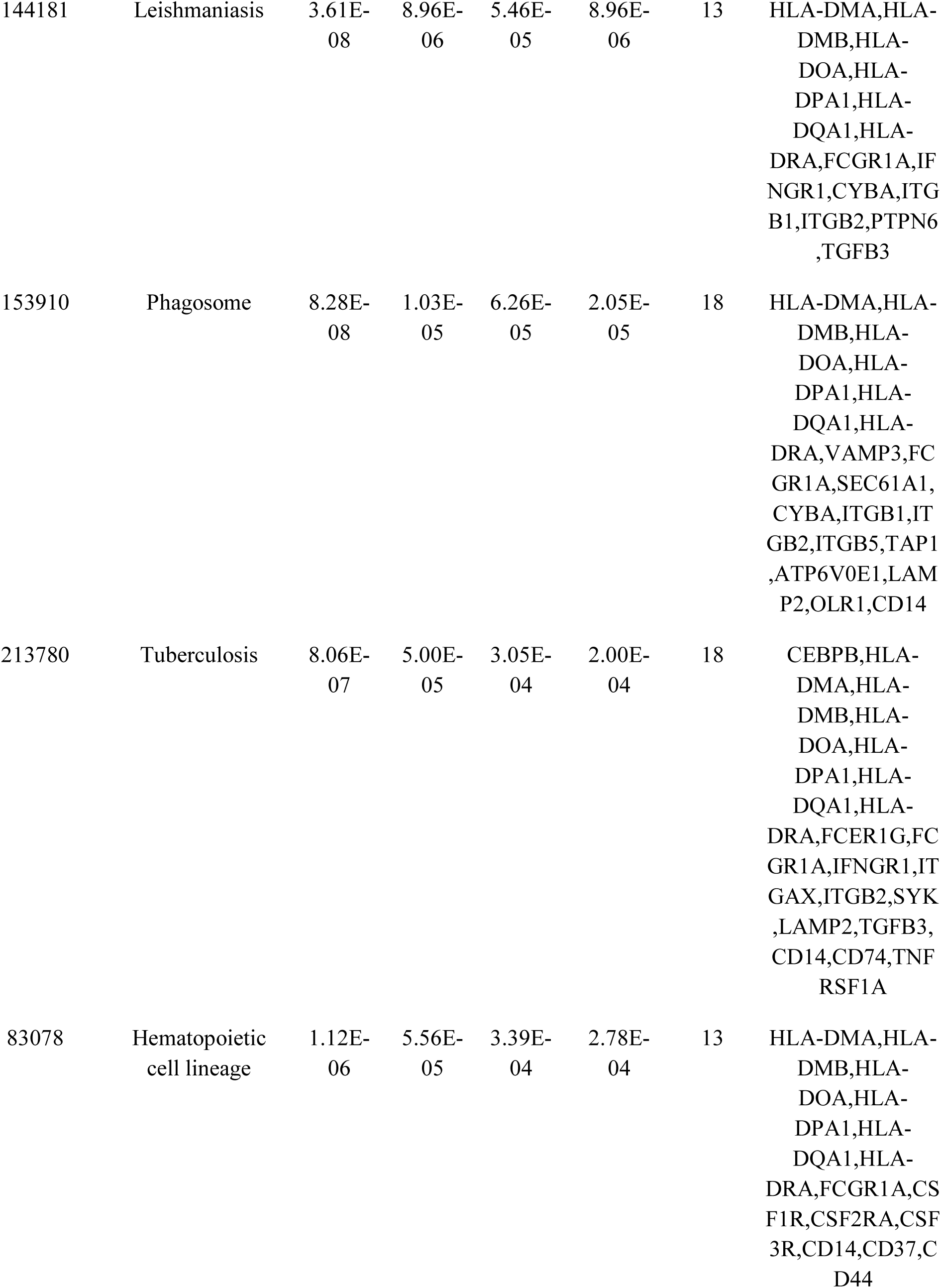
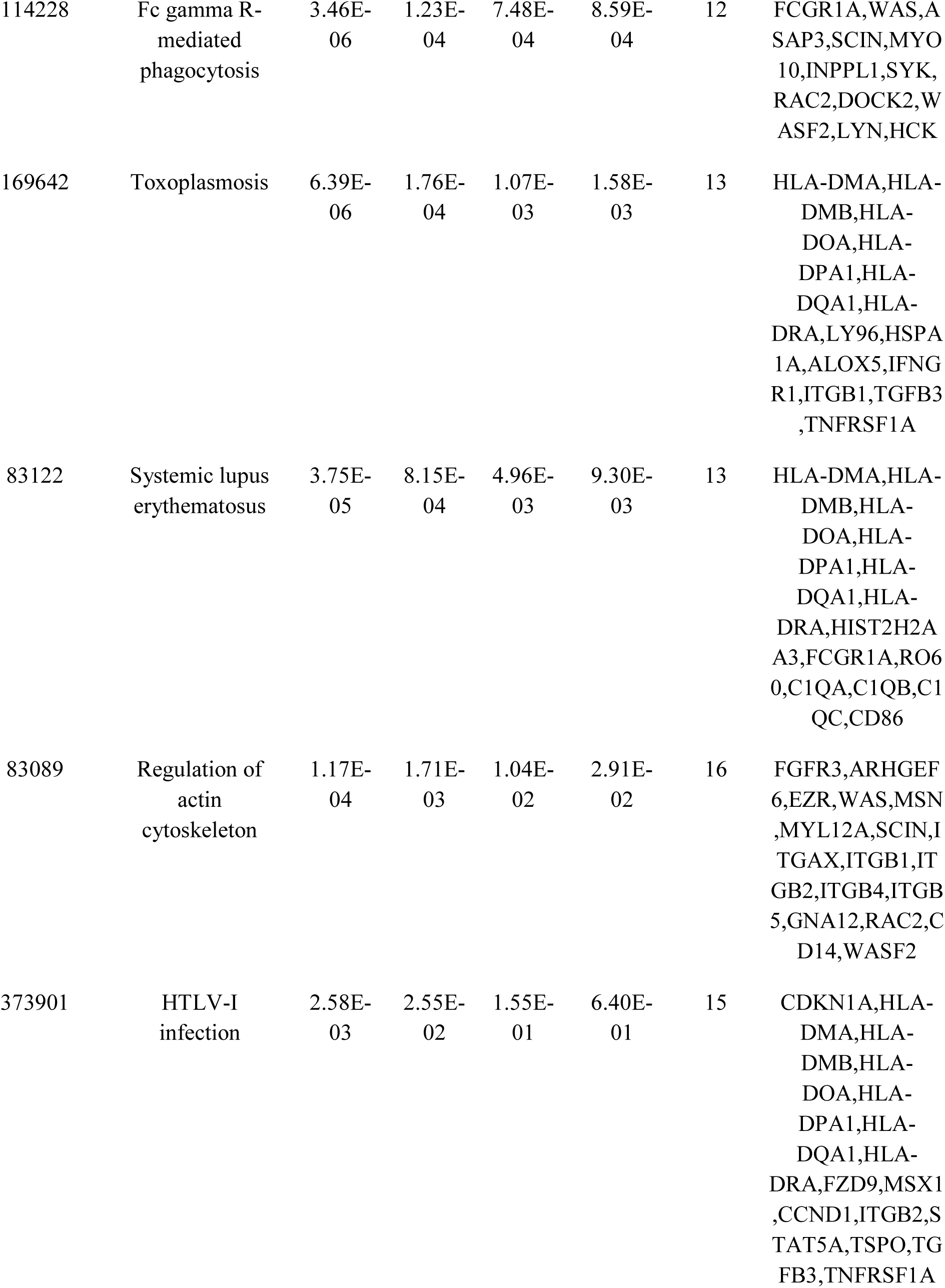
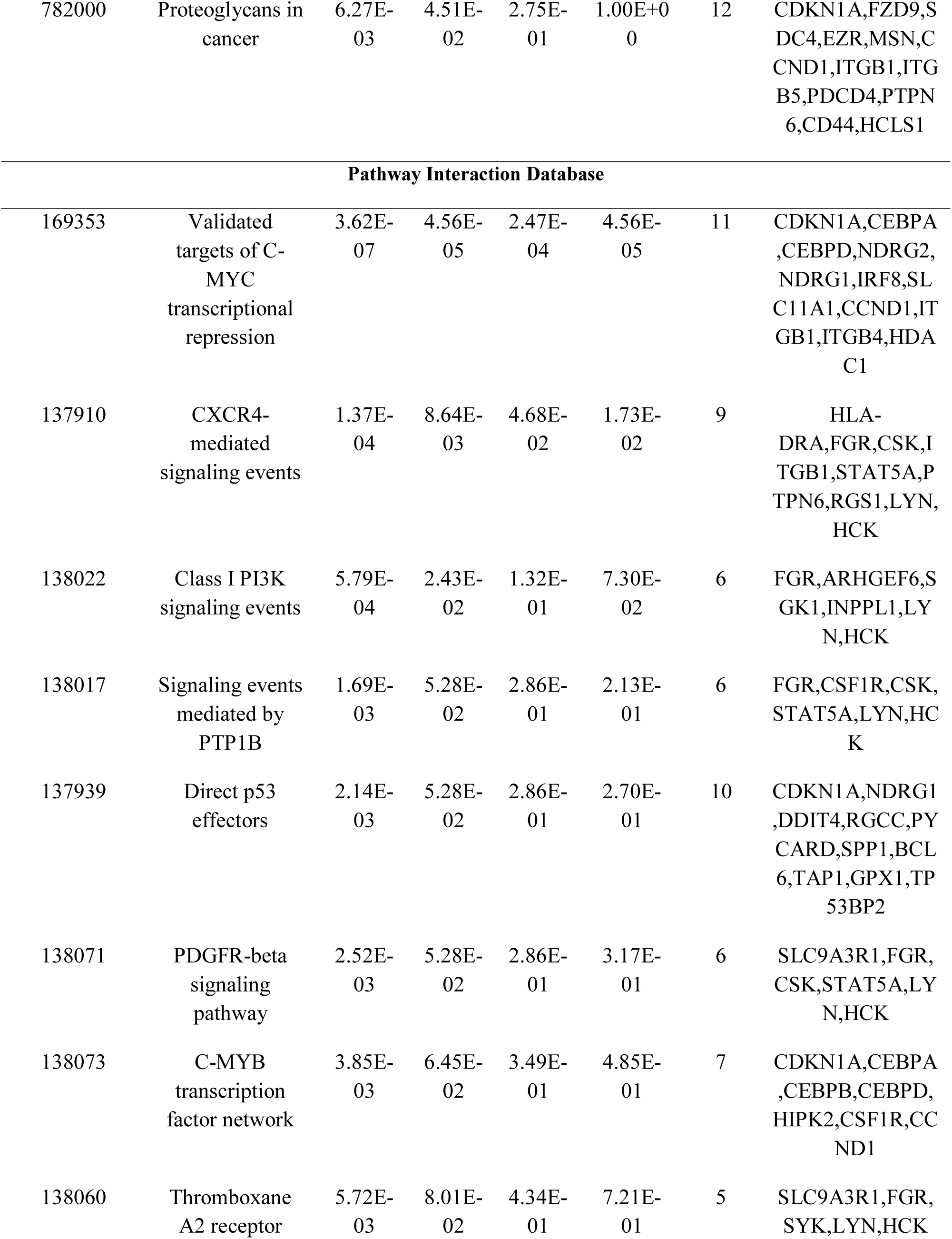
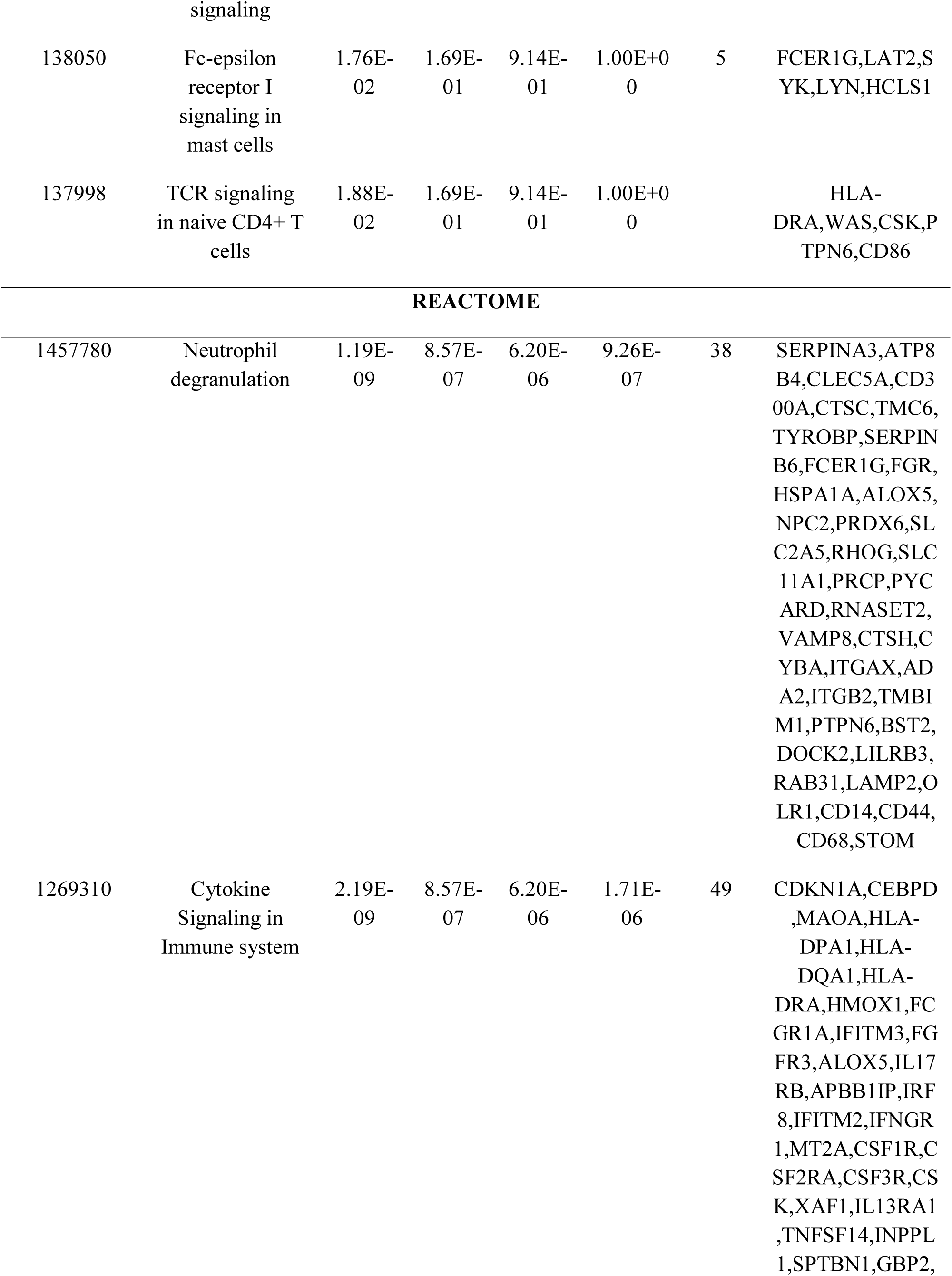
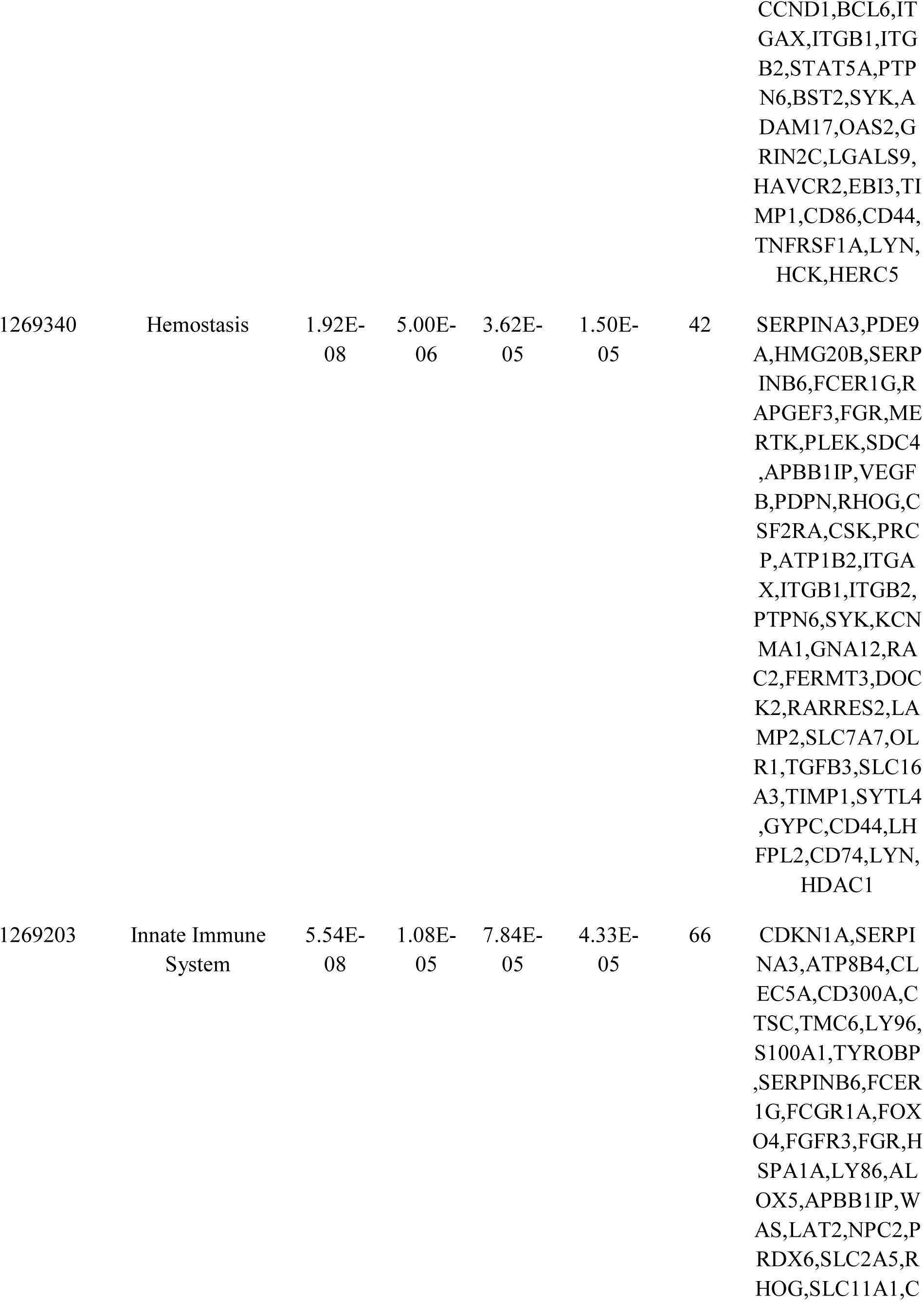
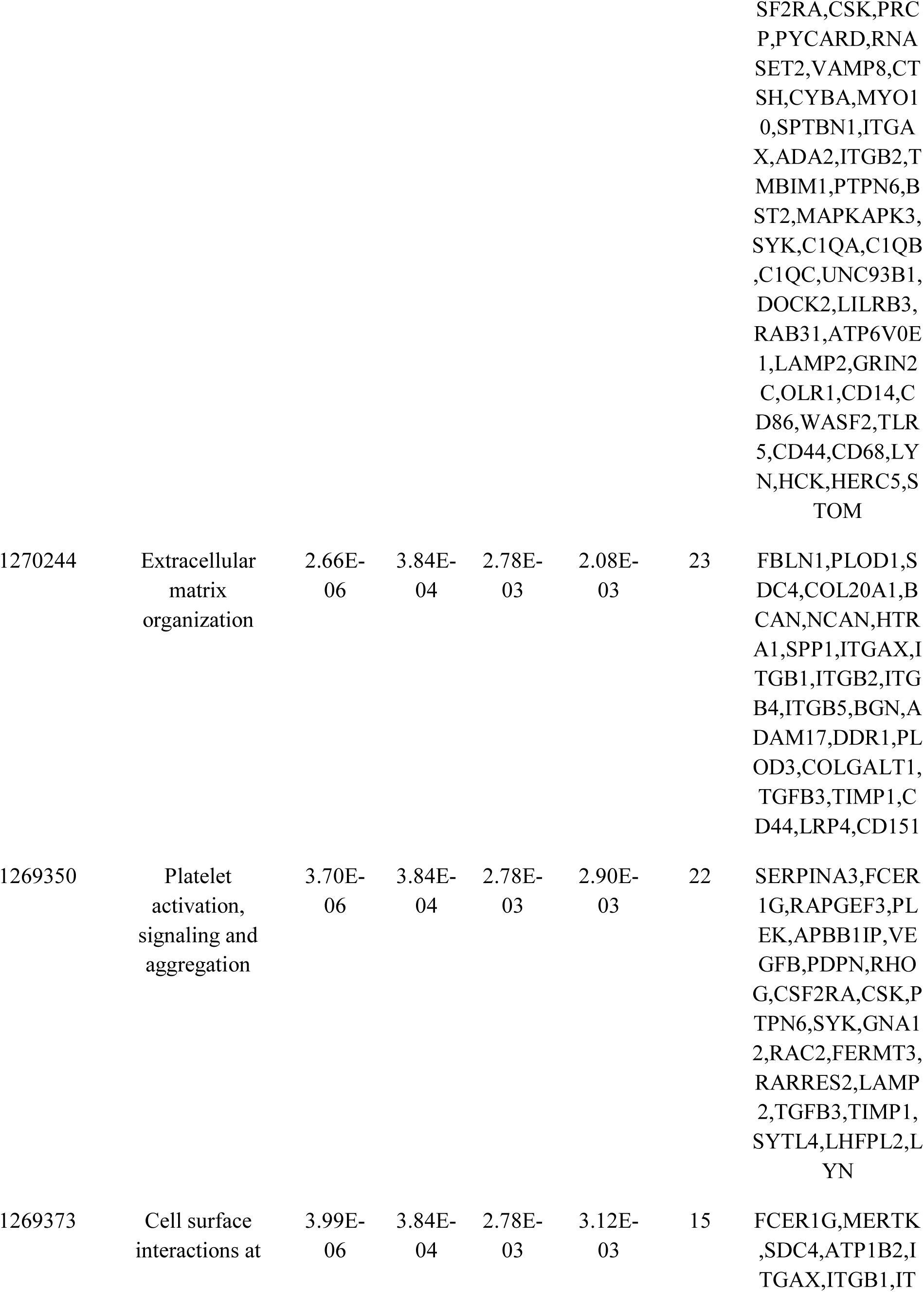
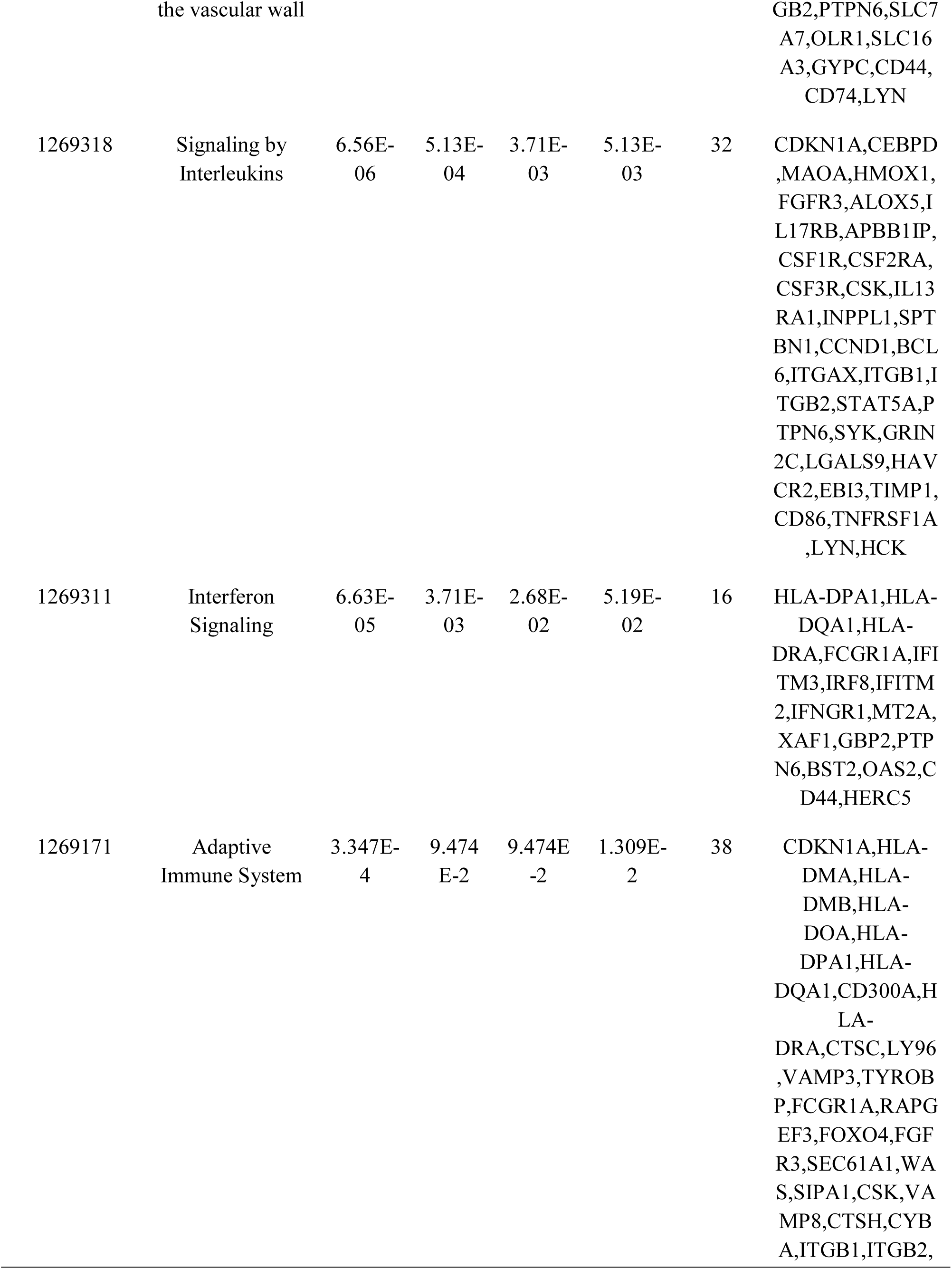
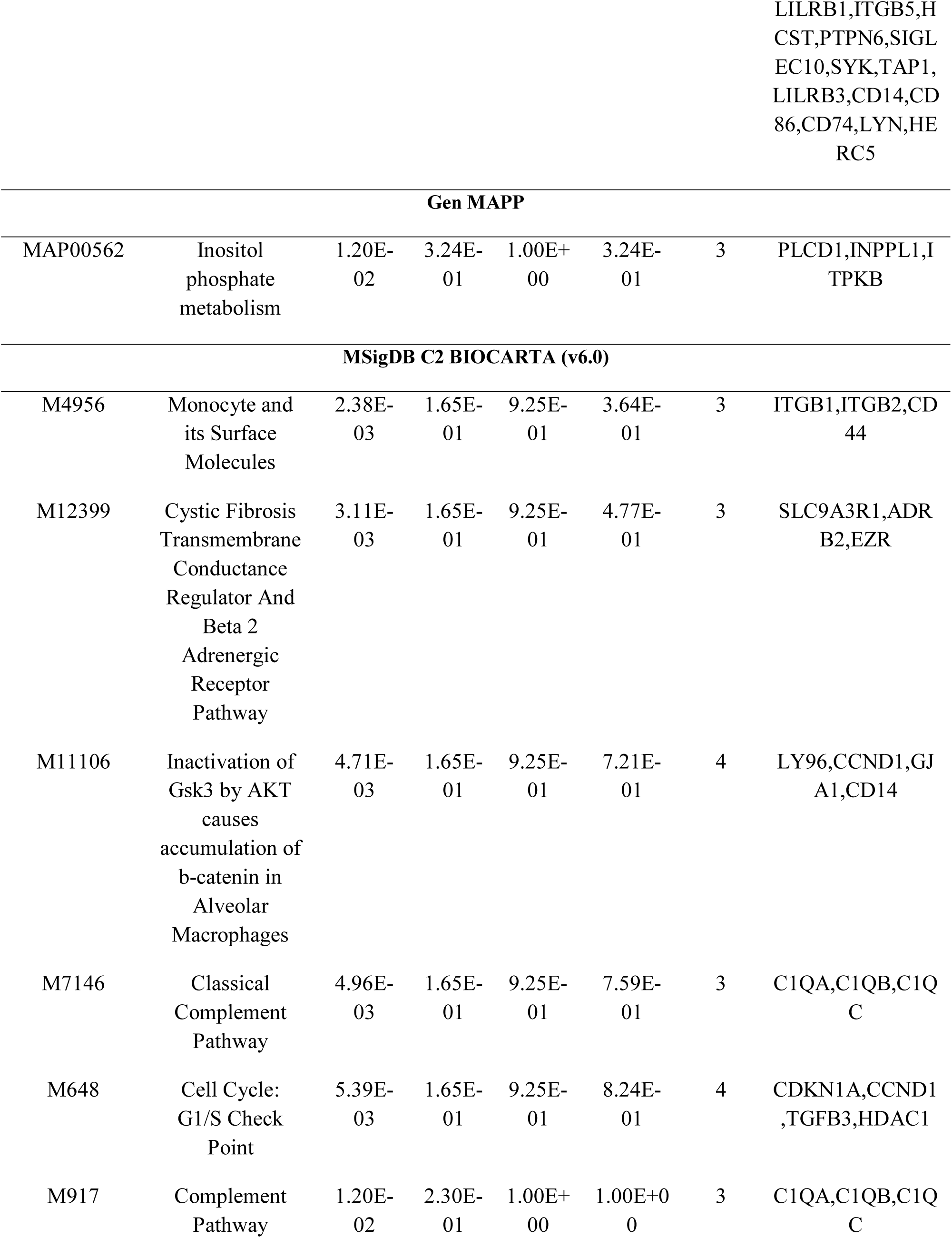
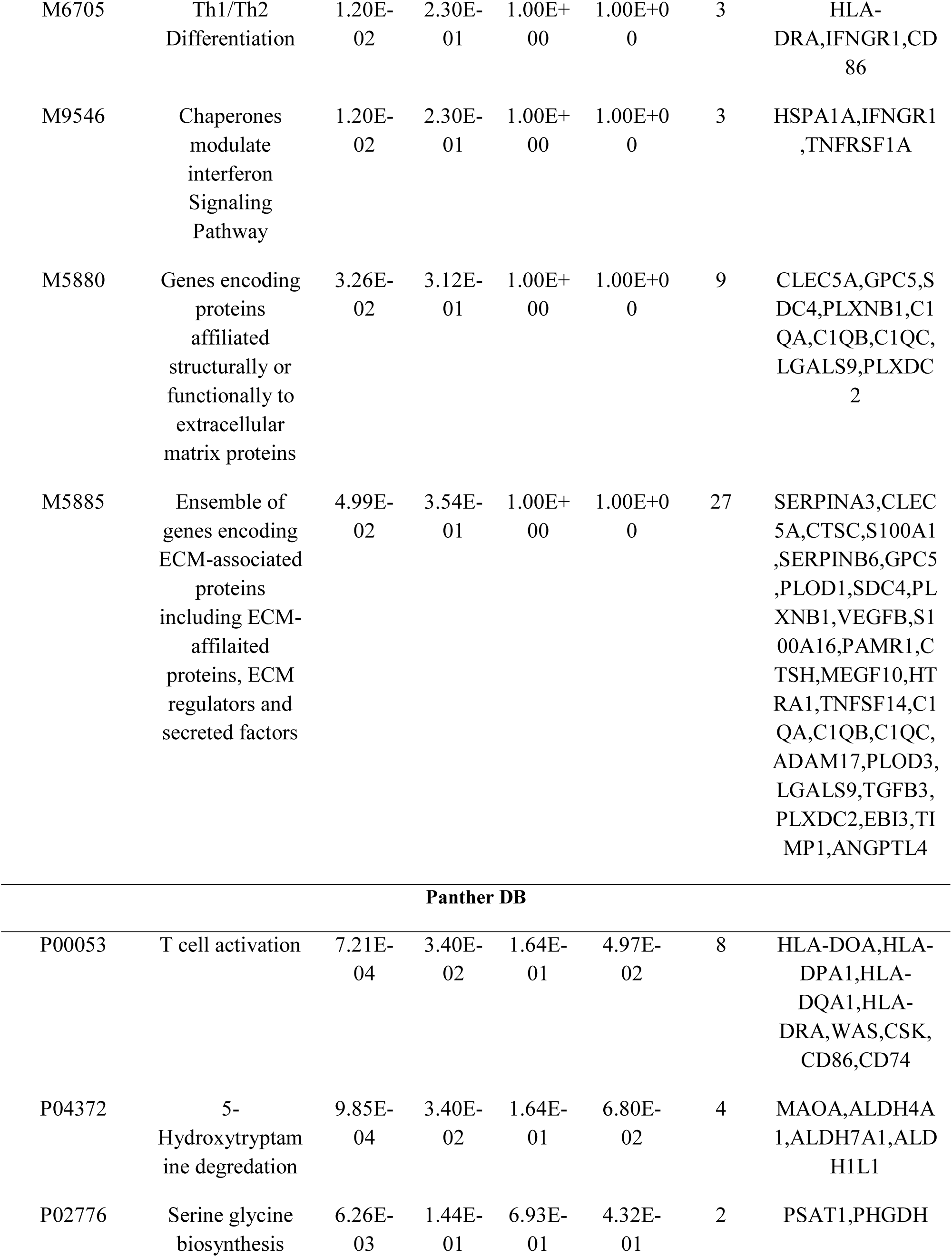
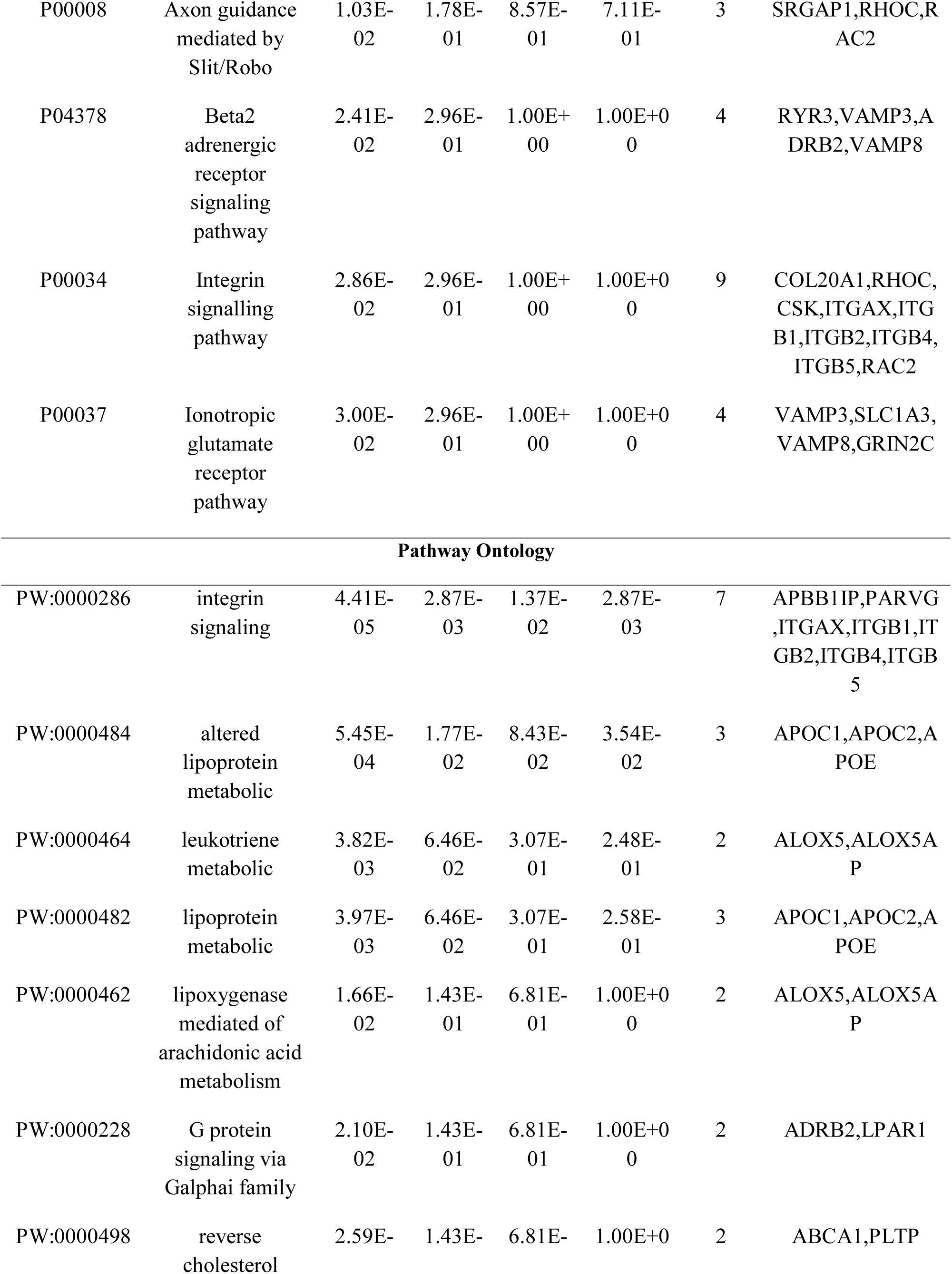
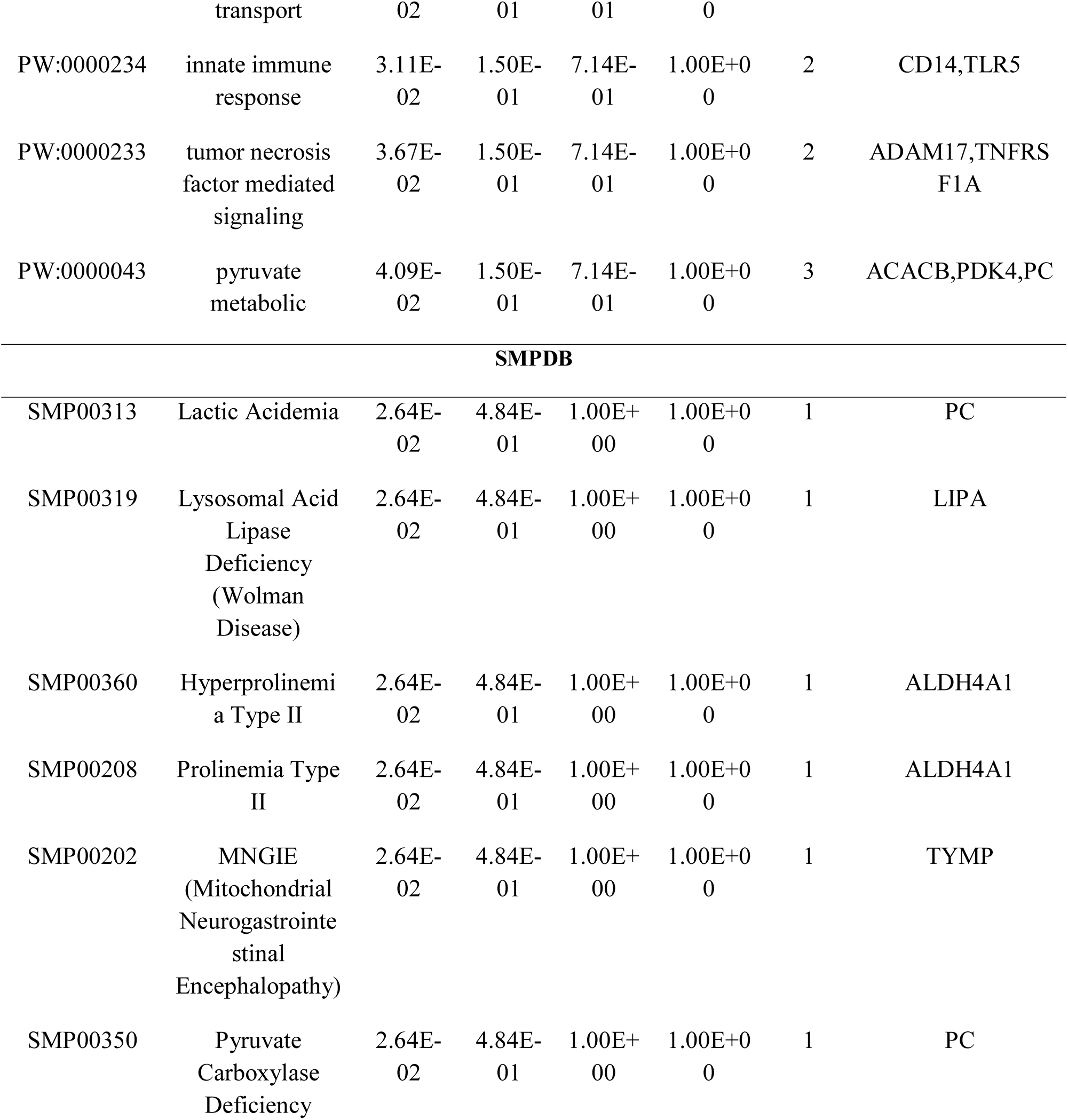
The enriched pathway terms of the down regulated differentially expressed genes

### Gene ontology (GO) enrichment analysis of DEGs

Gene ontology (GO) enrichment analysis of DEGs was performed with ToppGene, and, up and down regulated genes were classified into three functional groups such as biological processes (BP), molecular functions (MF) and cell compositions (CC). The results of Gene ontology (GO) enrichment analysis are given in Table 4 and Table 5. In the BP group, the up regulated genes were mainly enriched in chemical synaptic transmission and neuron projection development, and the down regulated genes were mainly enriched in the regulation of immune system process and cell activation. In the CC group, the up regulated genes were mainly enriched in neuron part and synapse, and the down regulated genes were mainly enriched in the membrane region and cell surface. In the MF group, the up regulated genes were mainly enriched in cytoskeletal protein binding and molecular function regulator, and the down regulated genes were mainly enriched in the enzyme binding and molecular transducer activity.

**Table 4.**
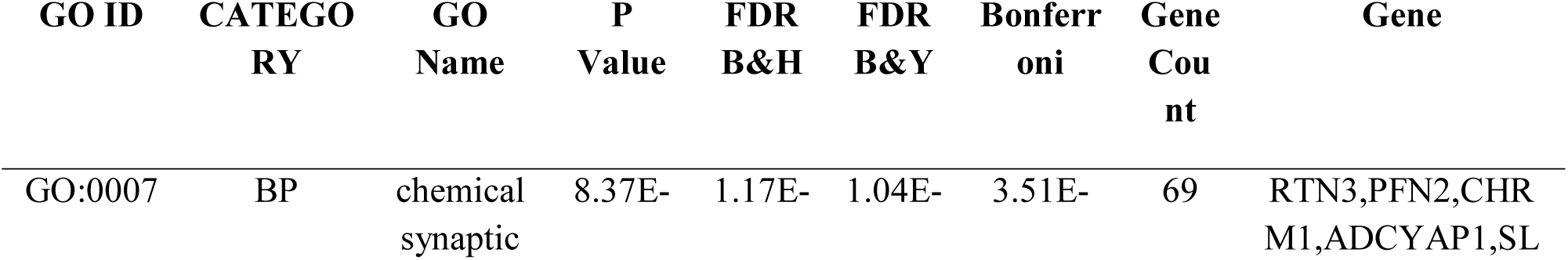
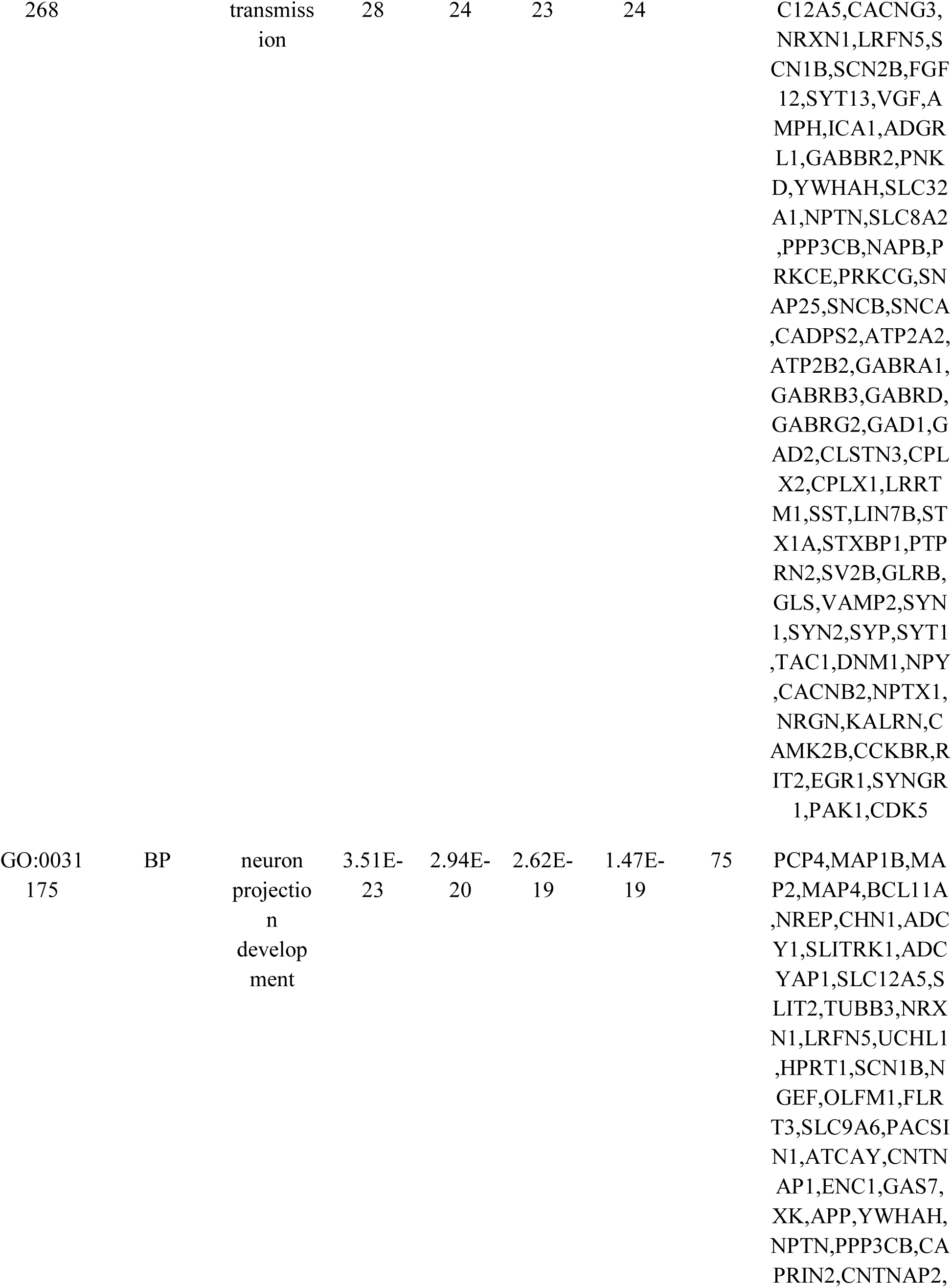
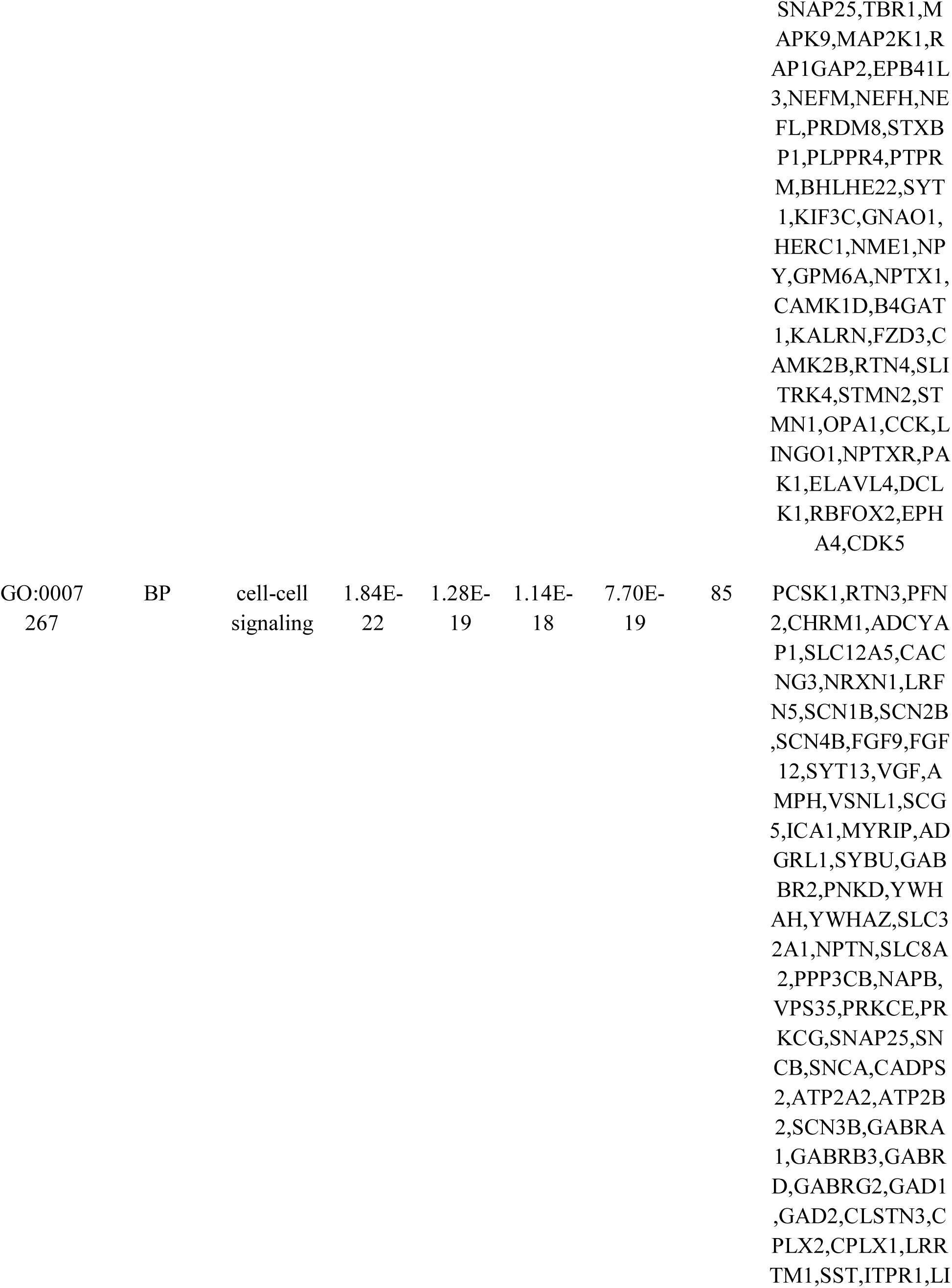
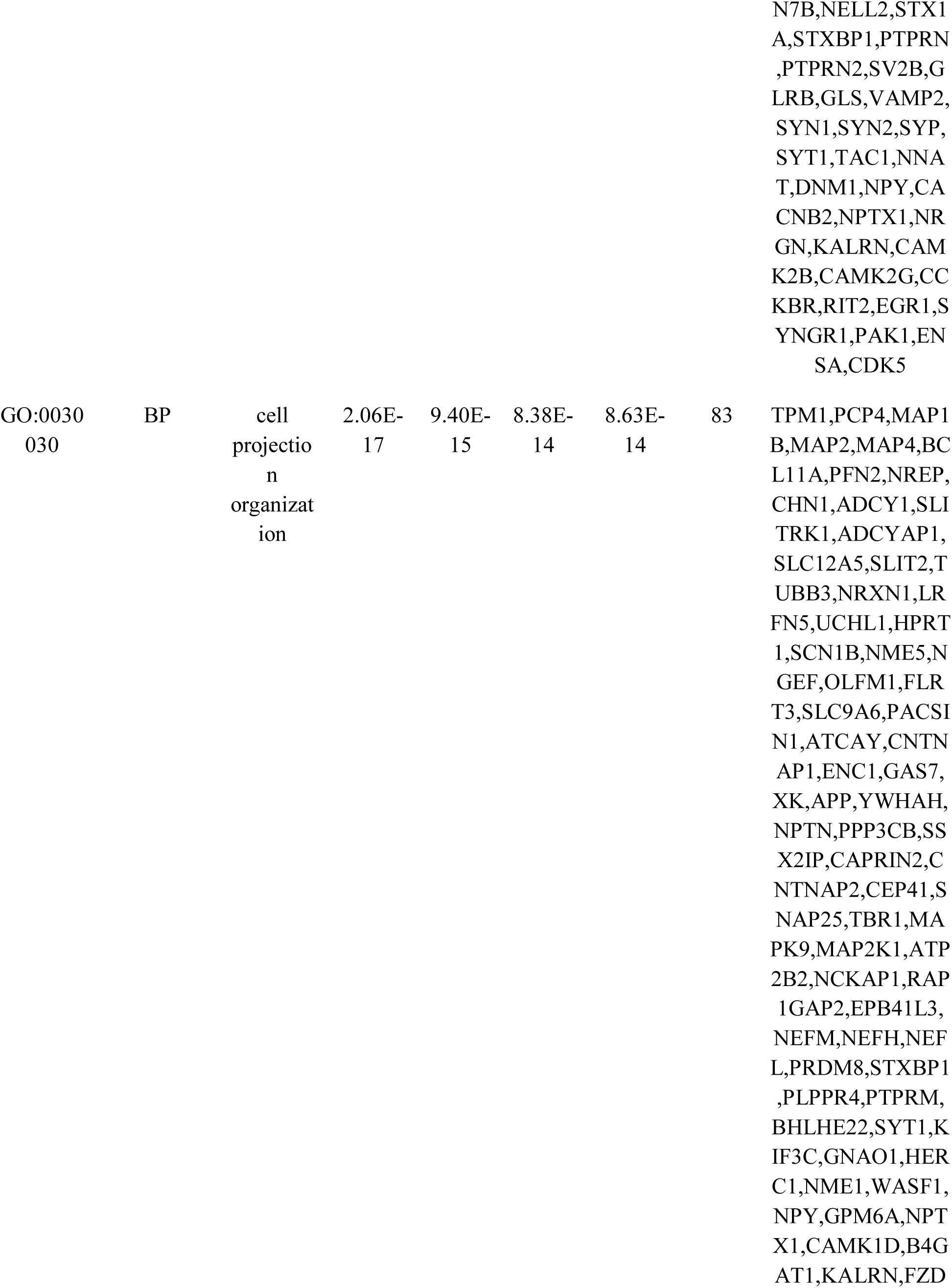
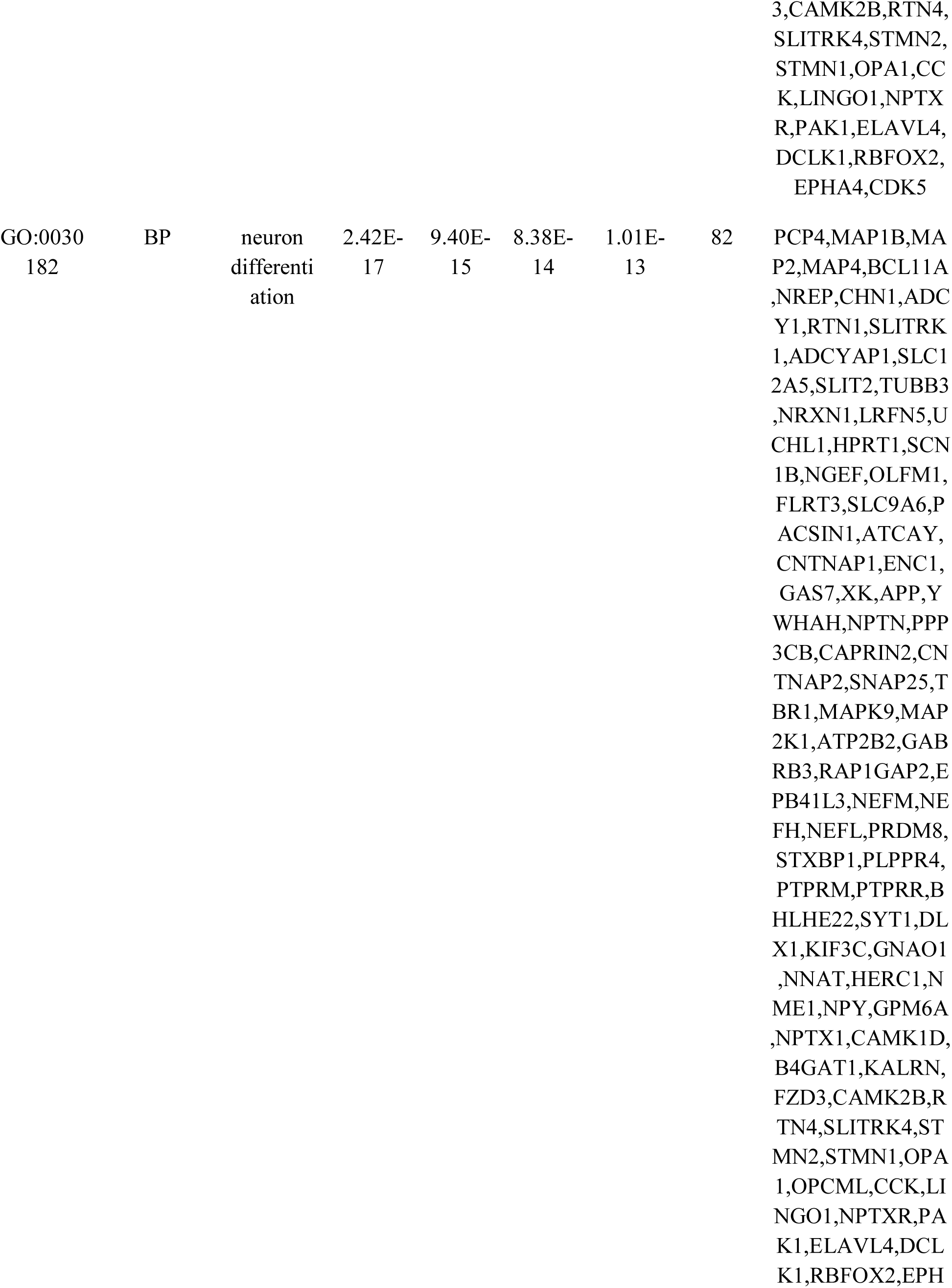
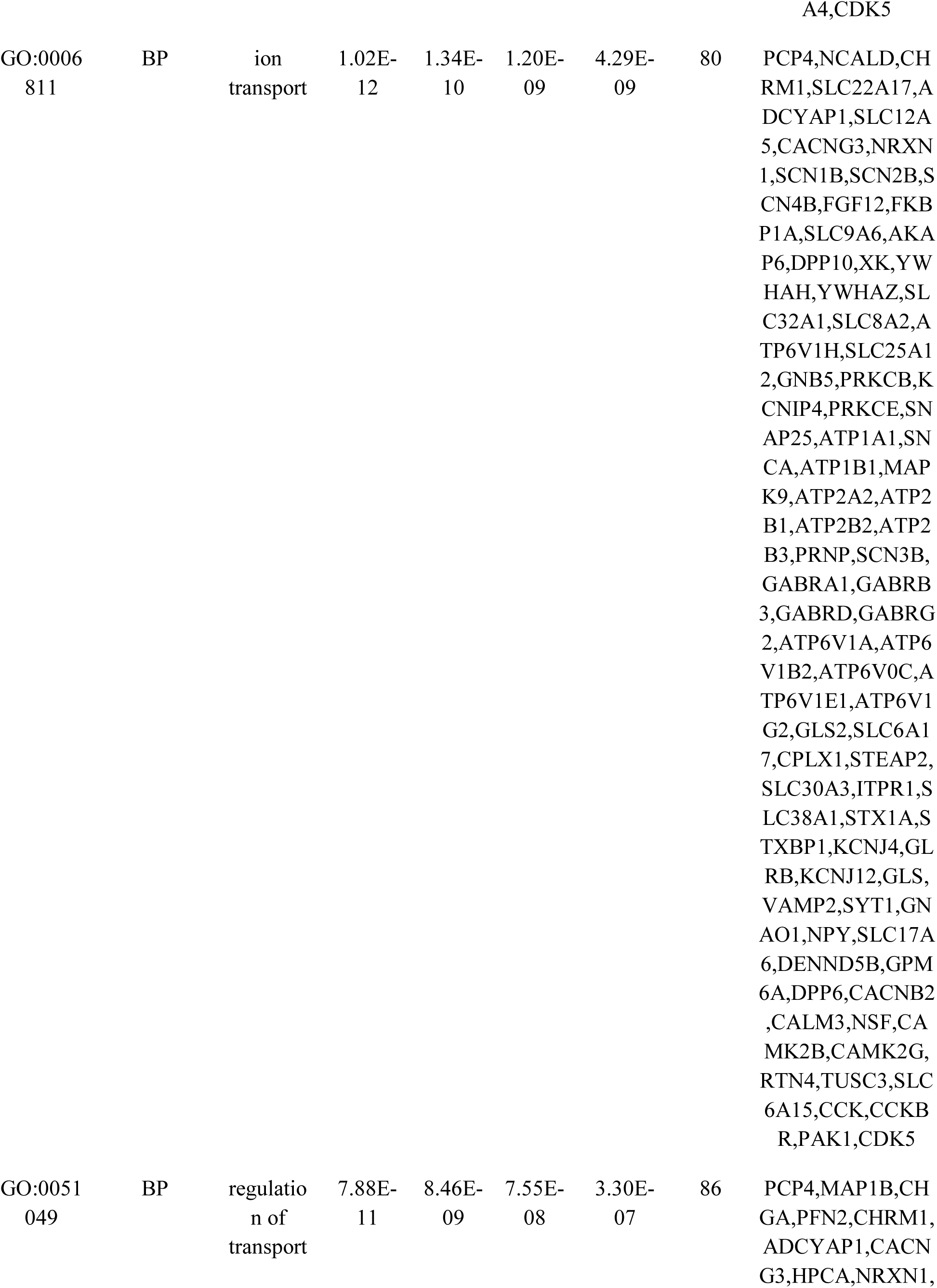
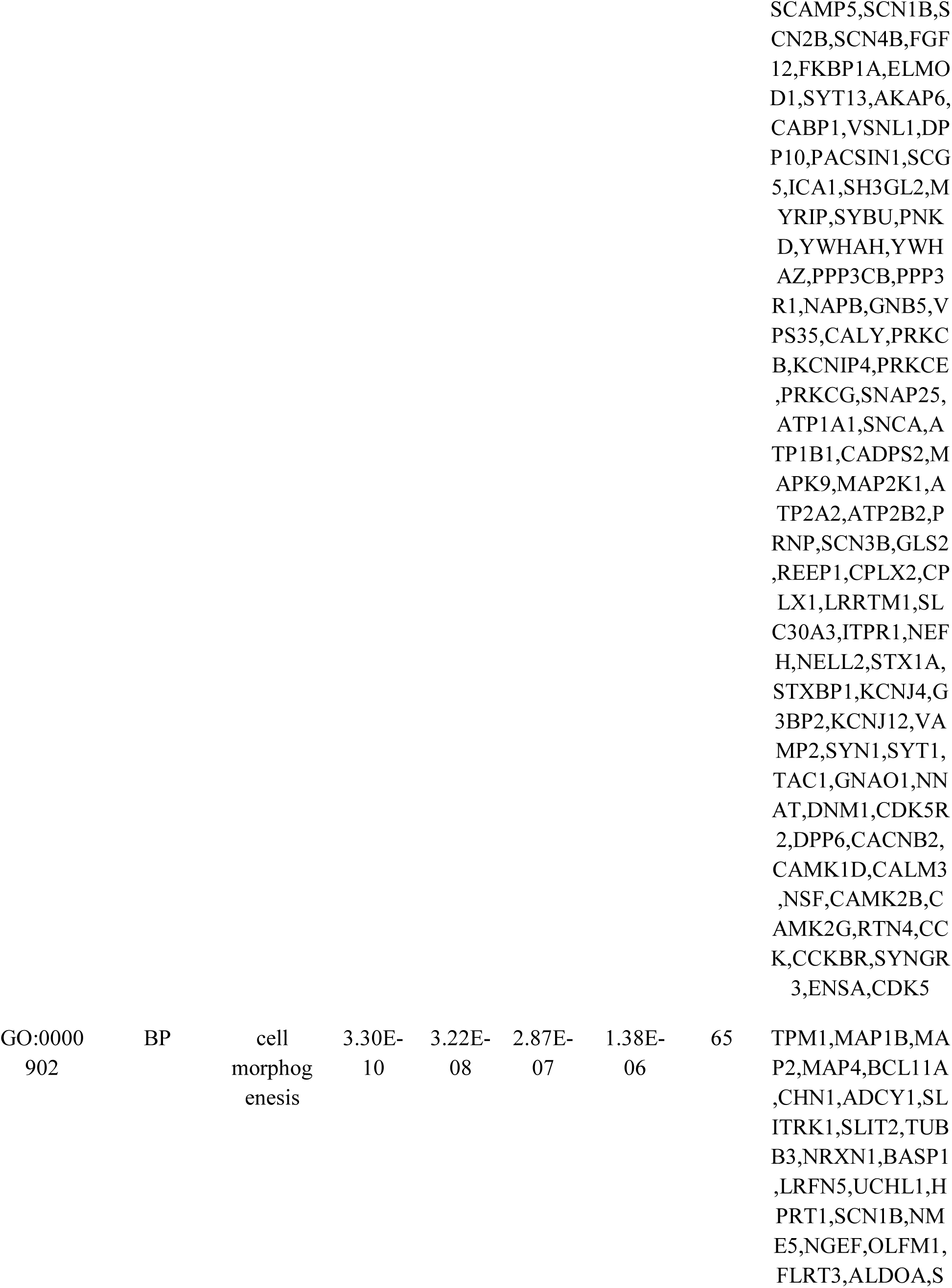
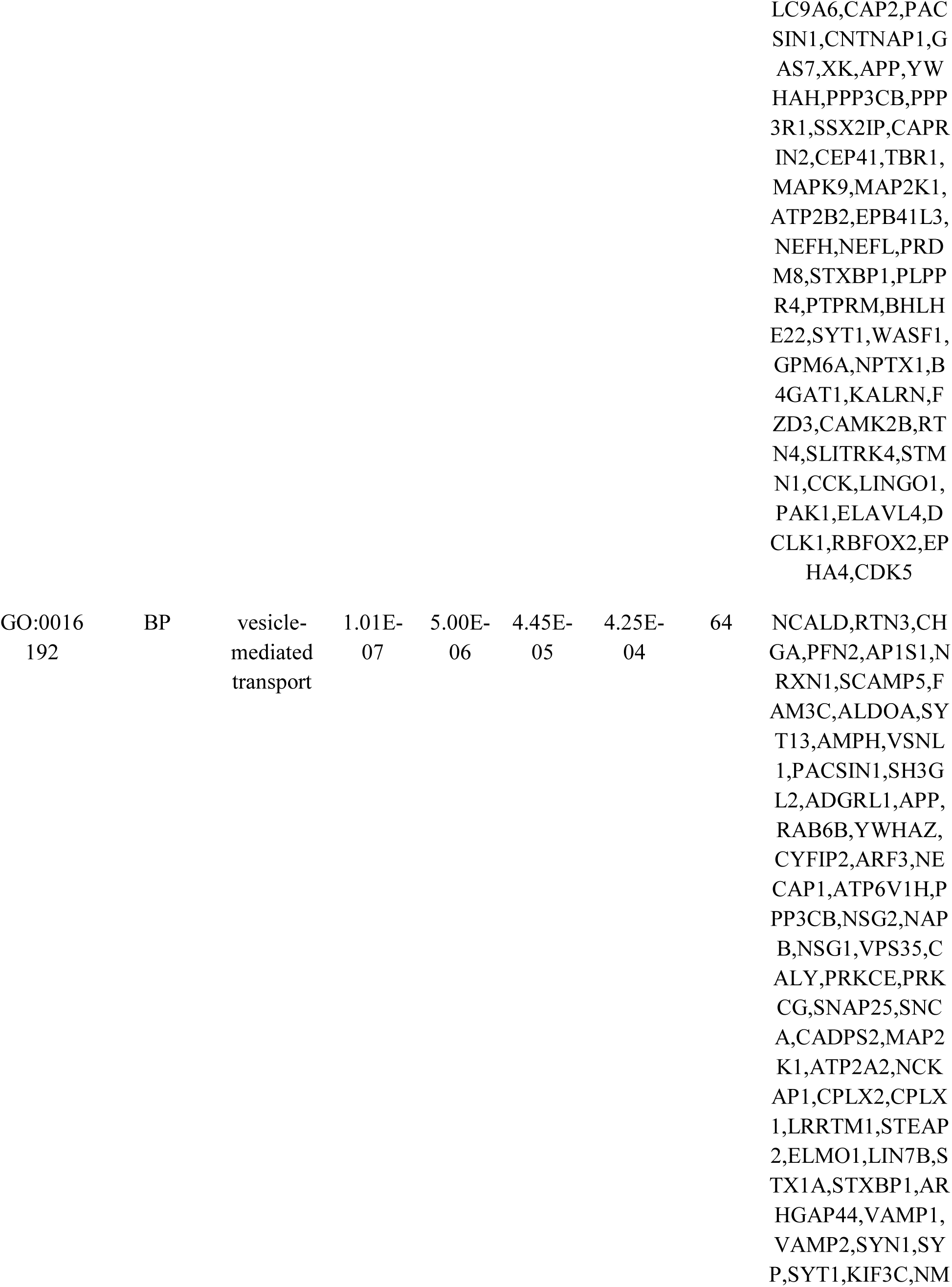
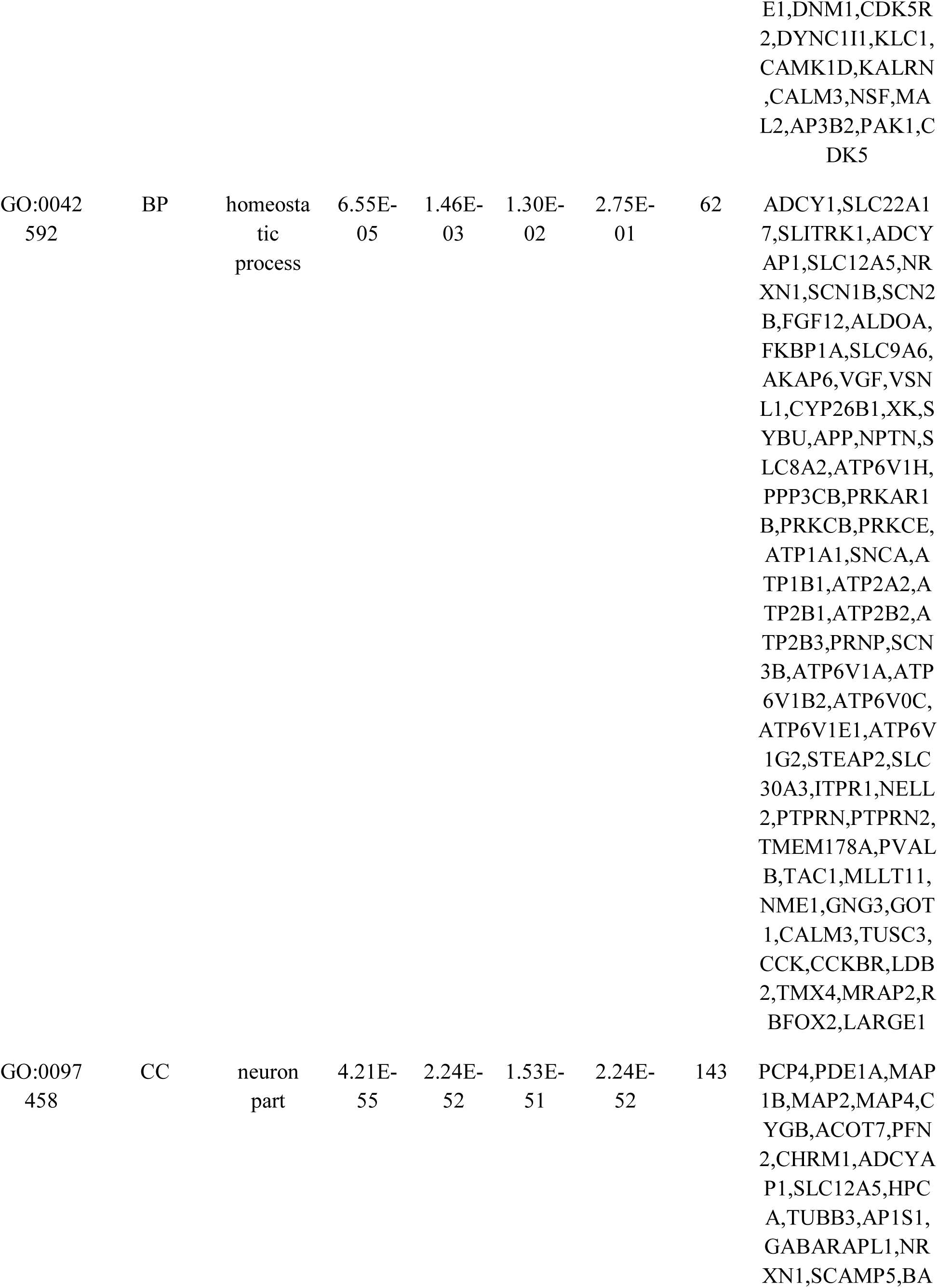
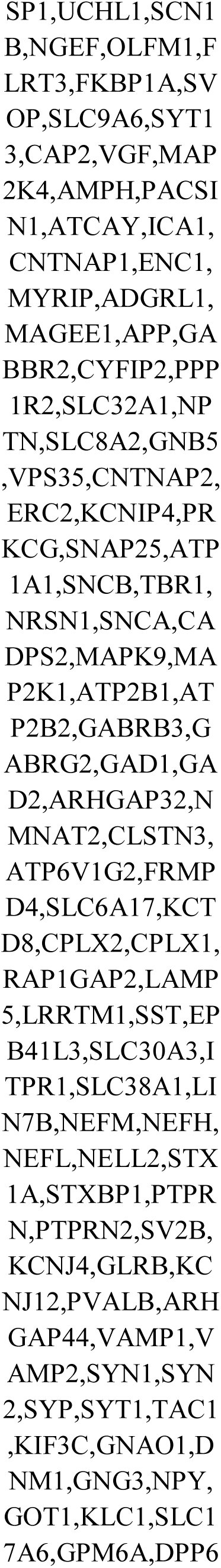
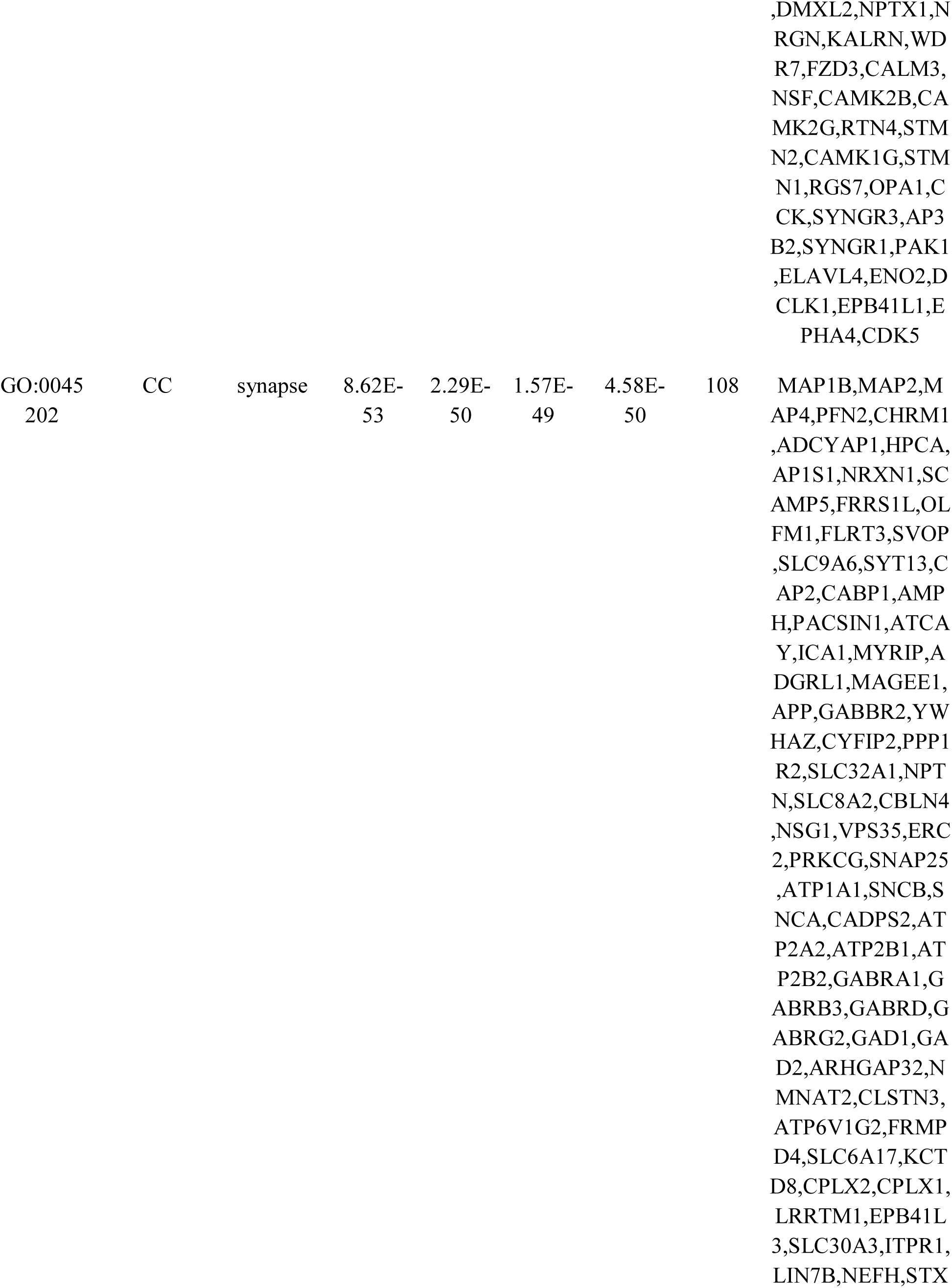
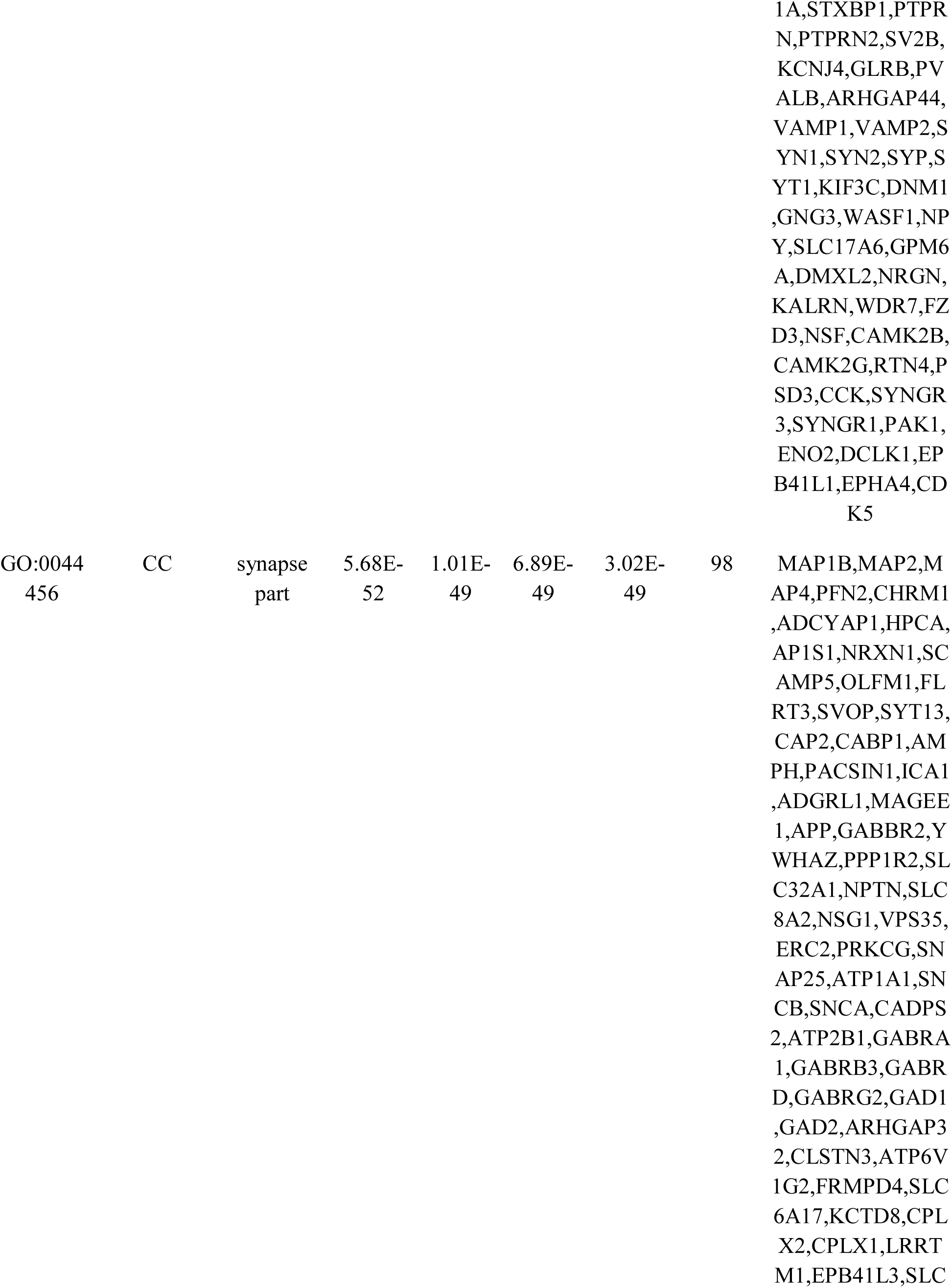
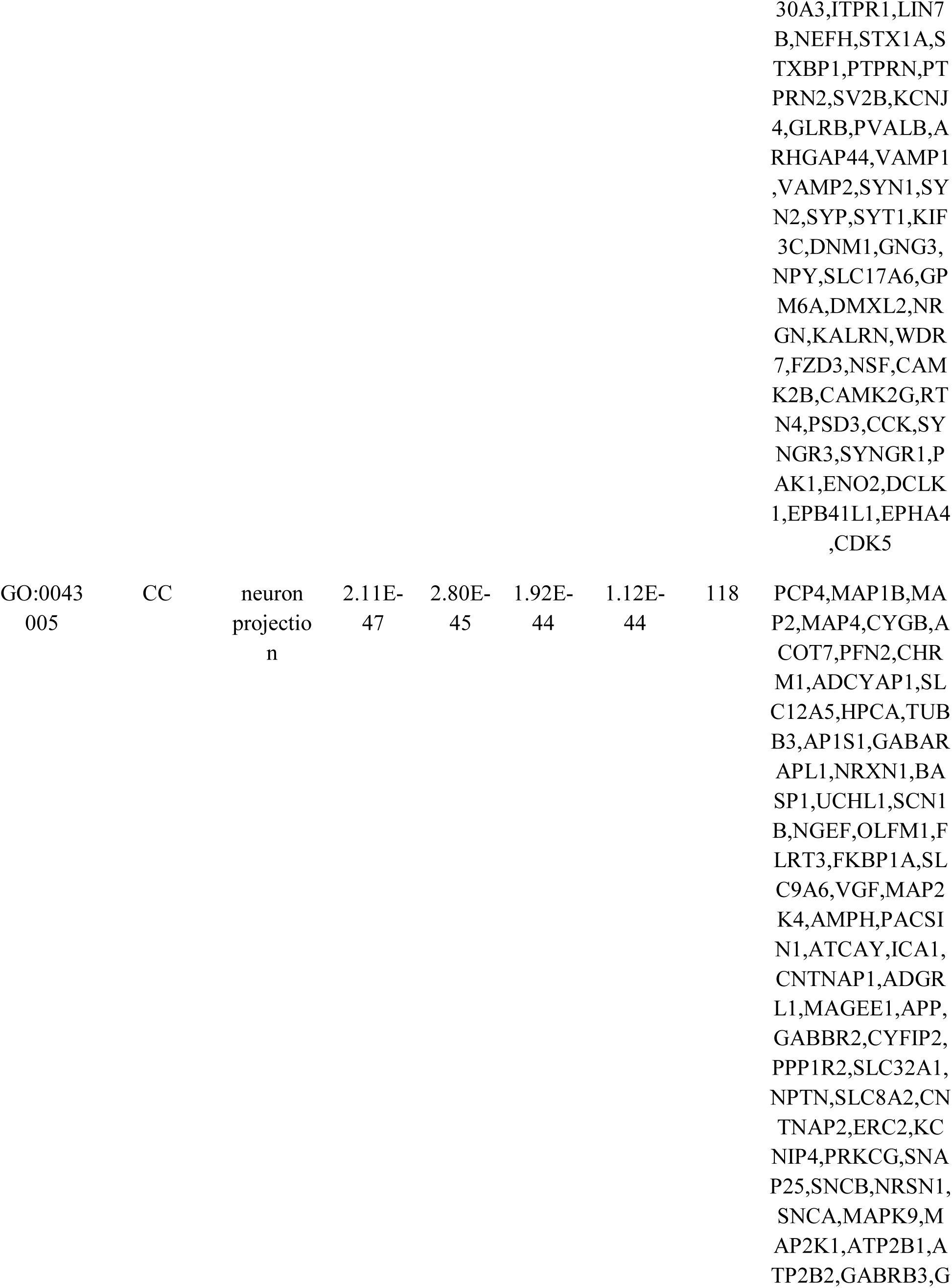
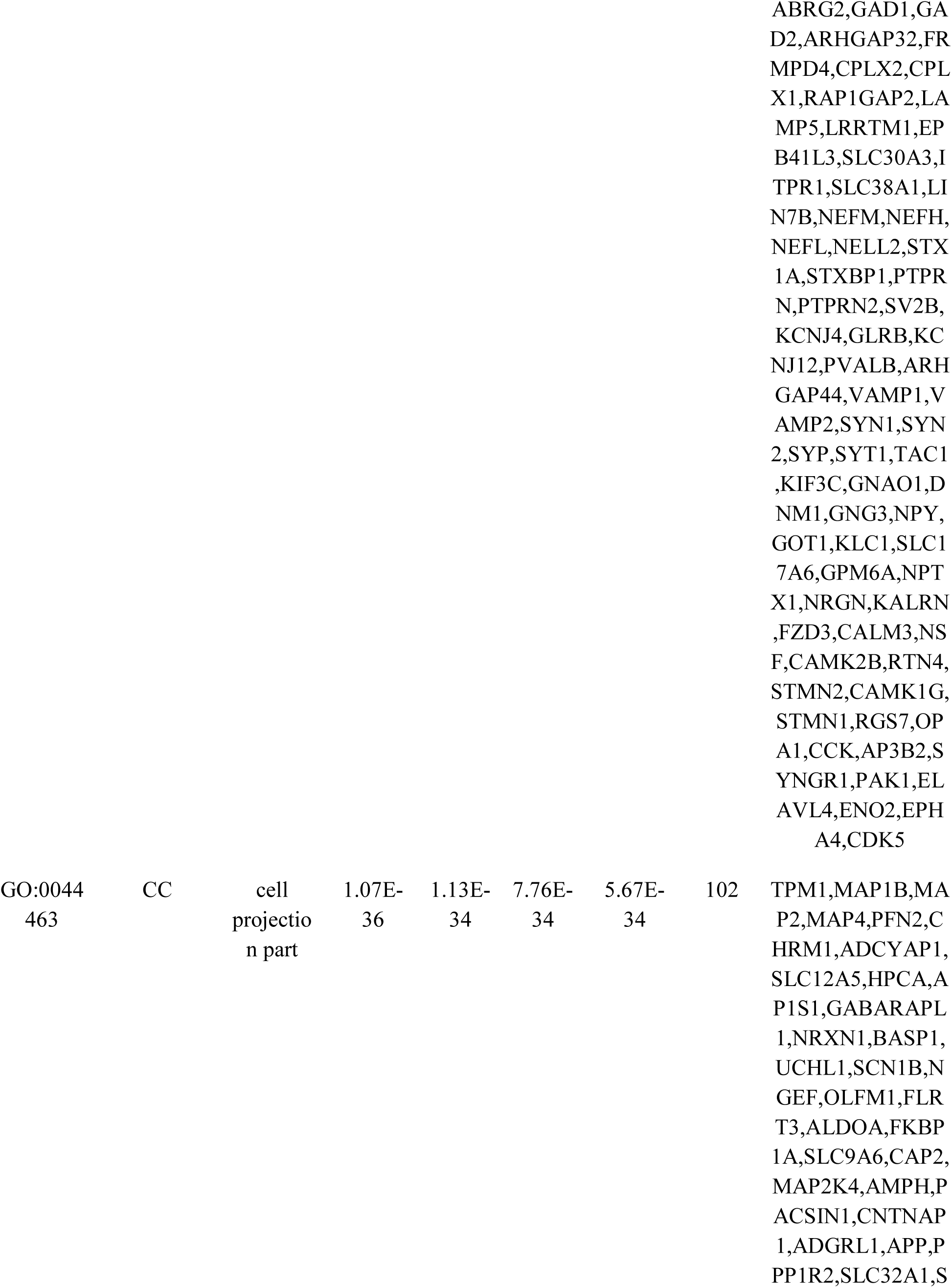
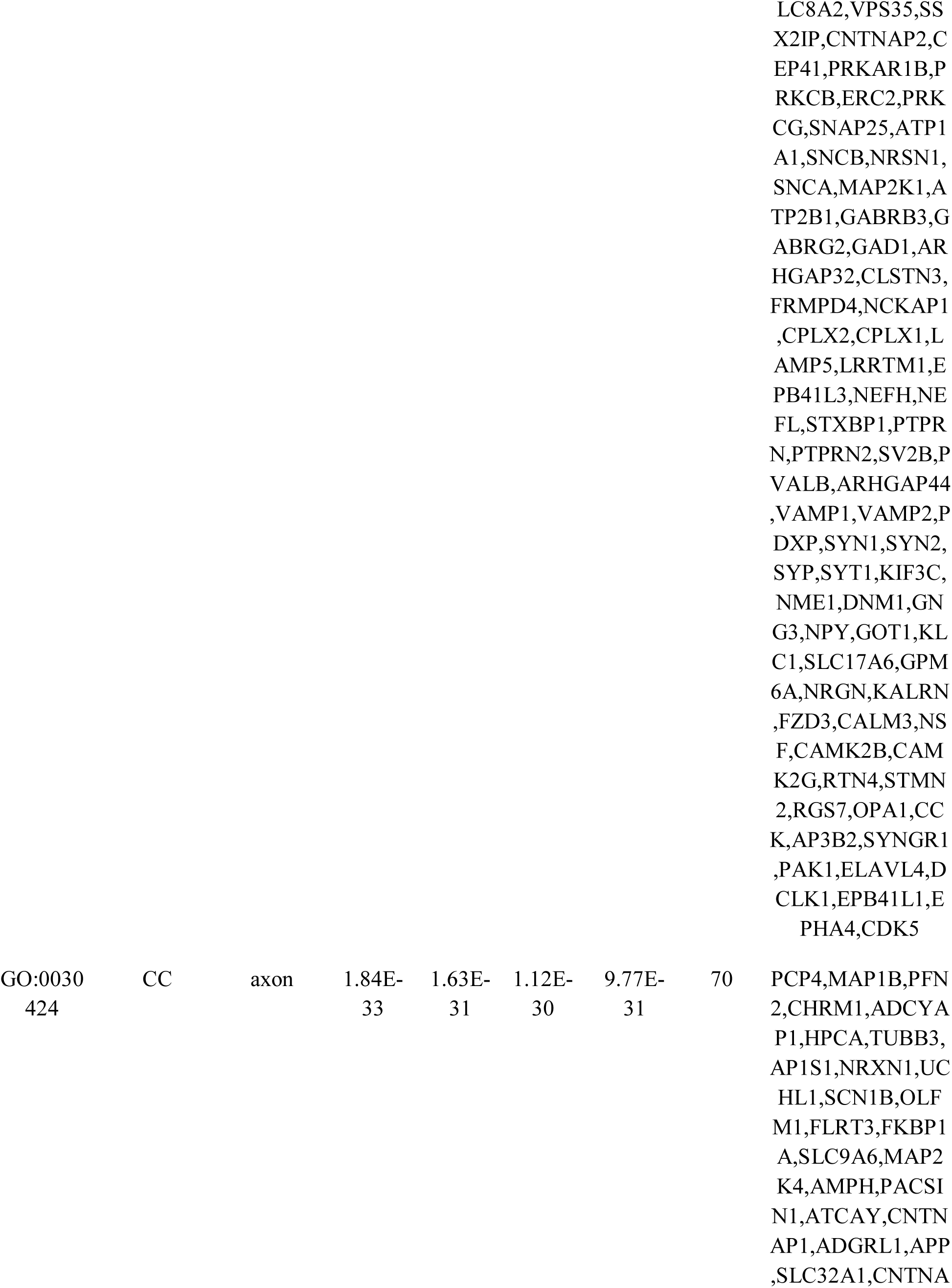
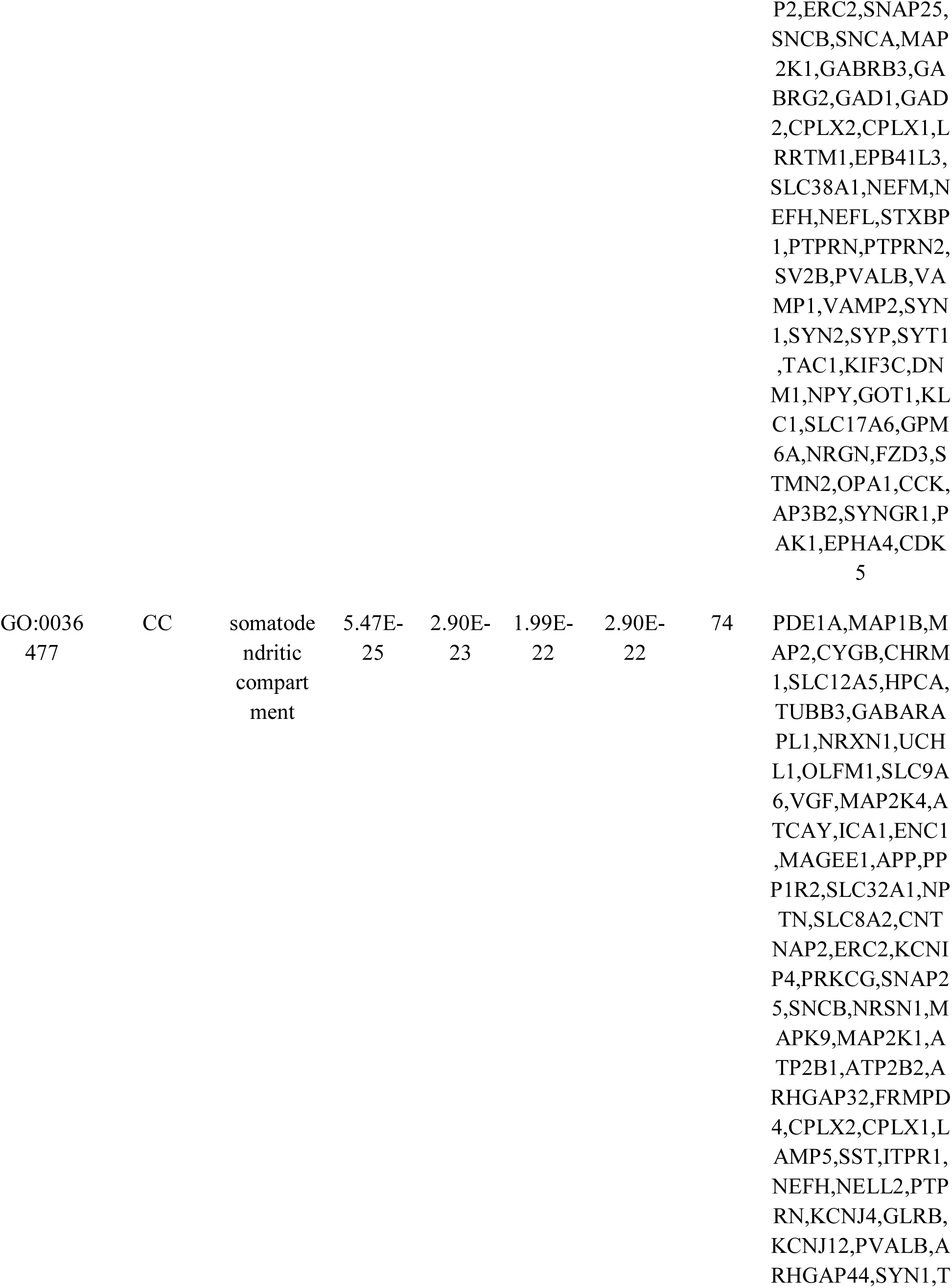
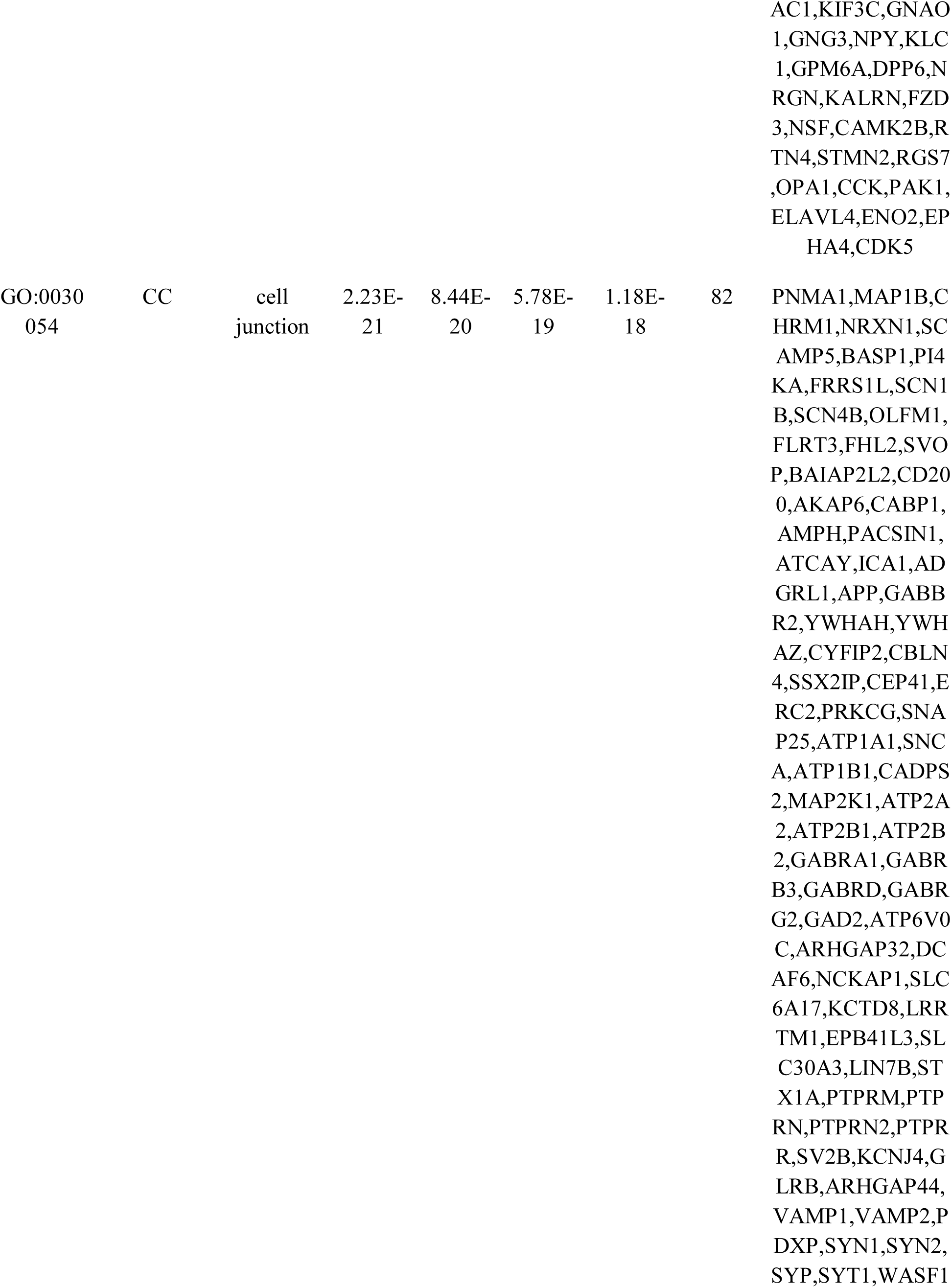
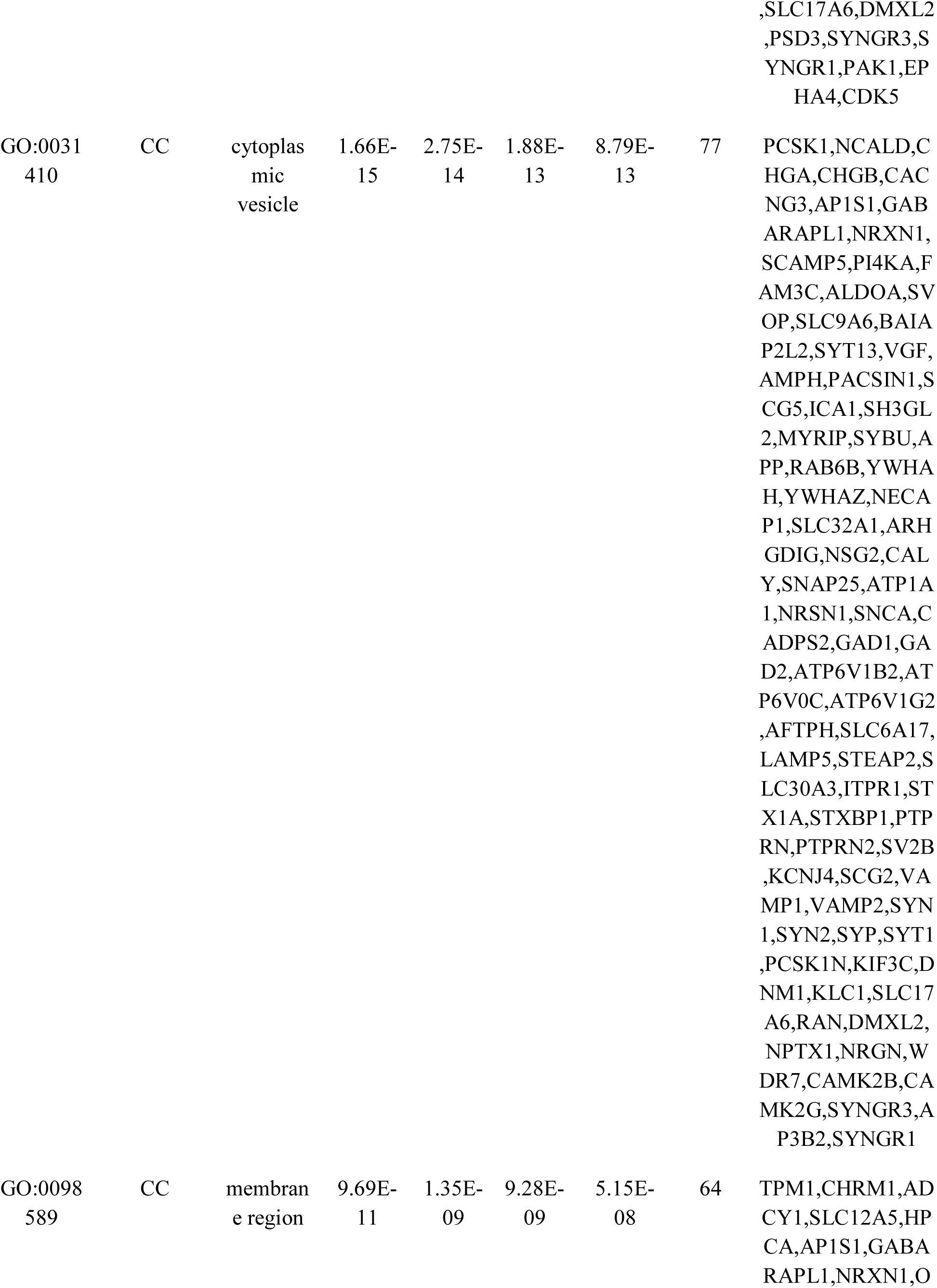
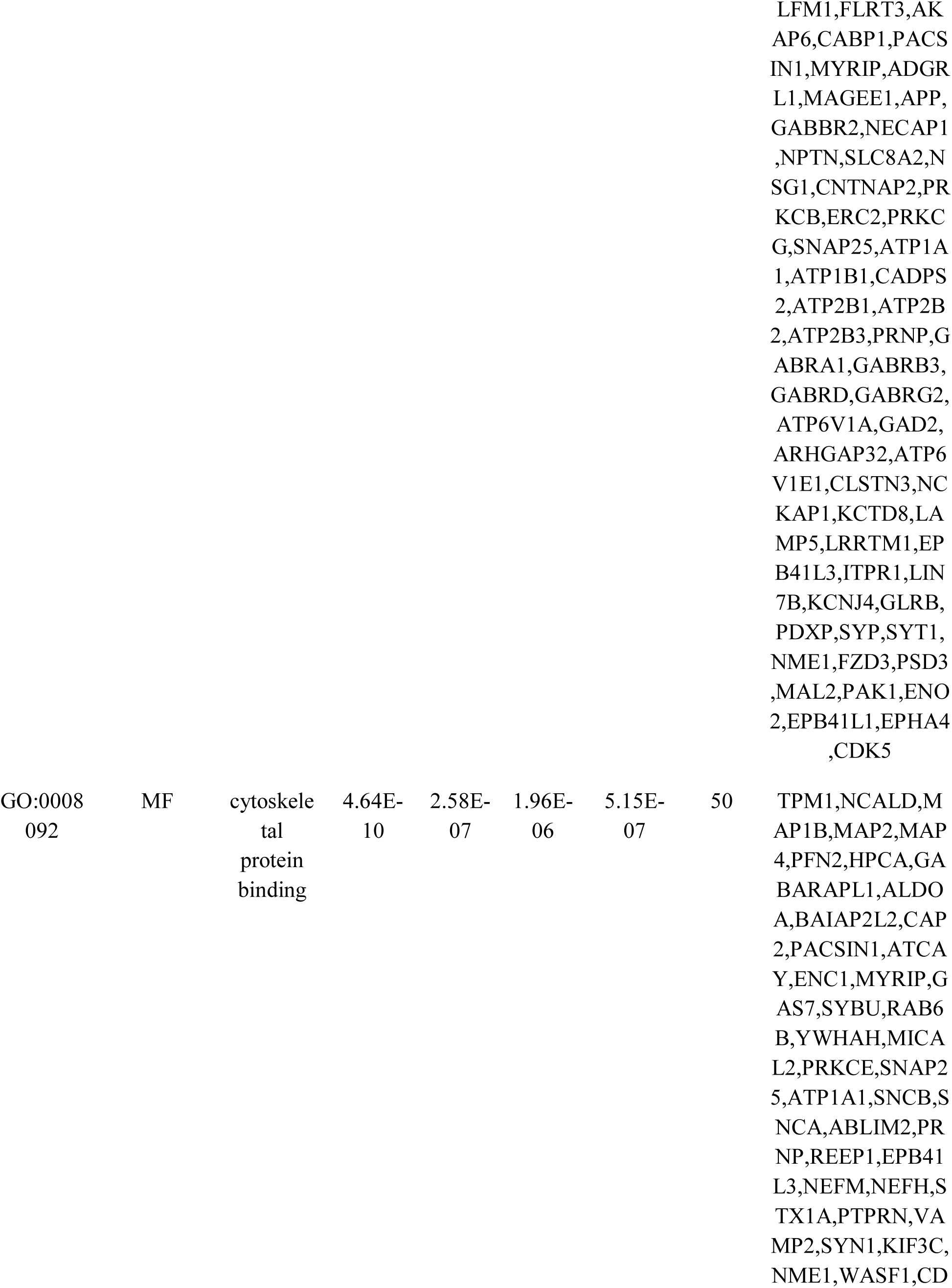
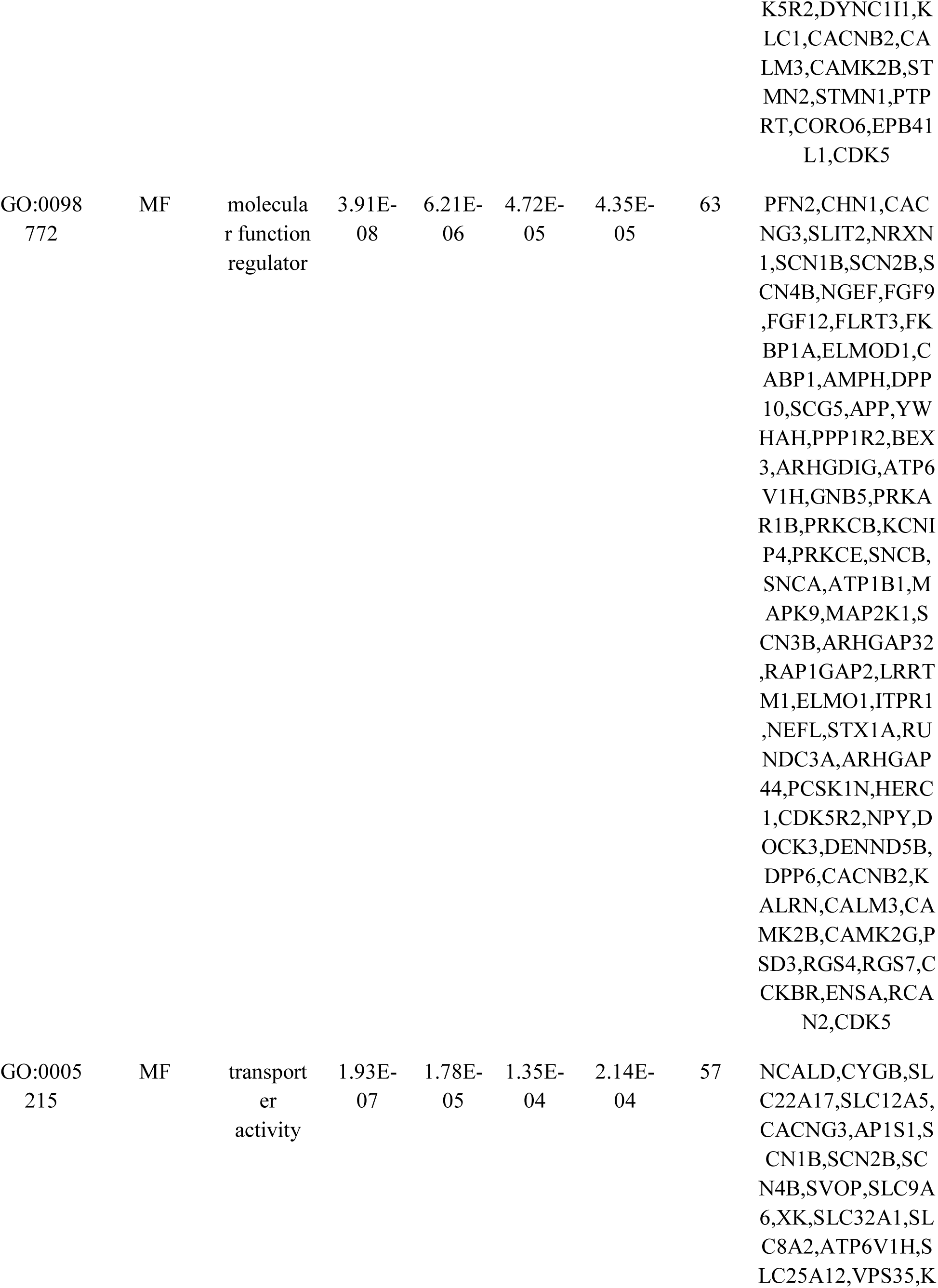
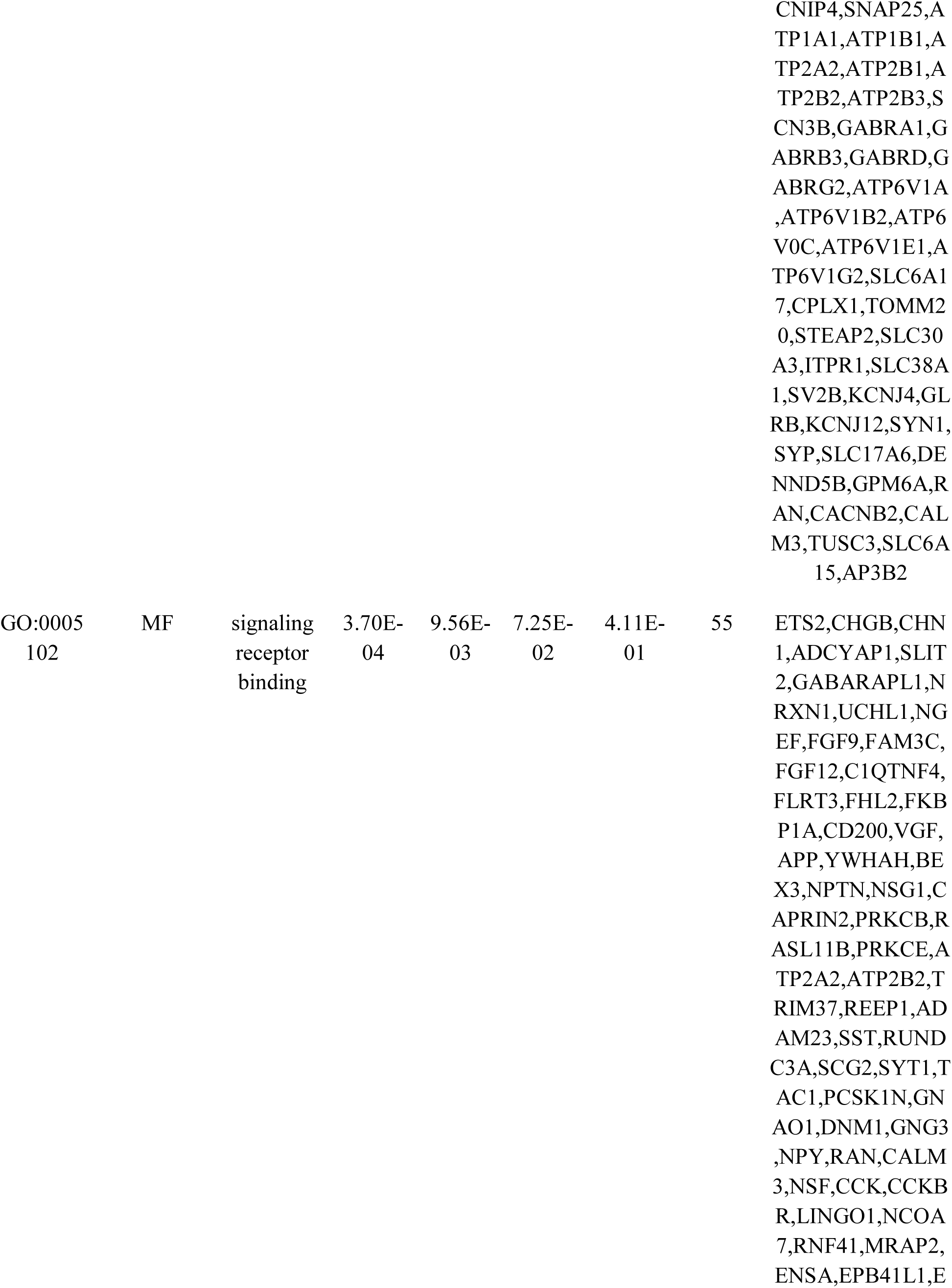
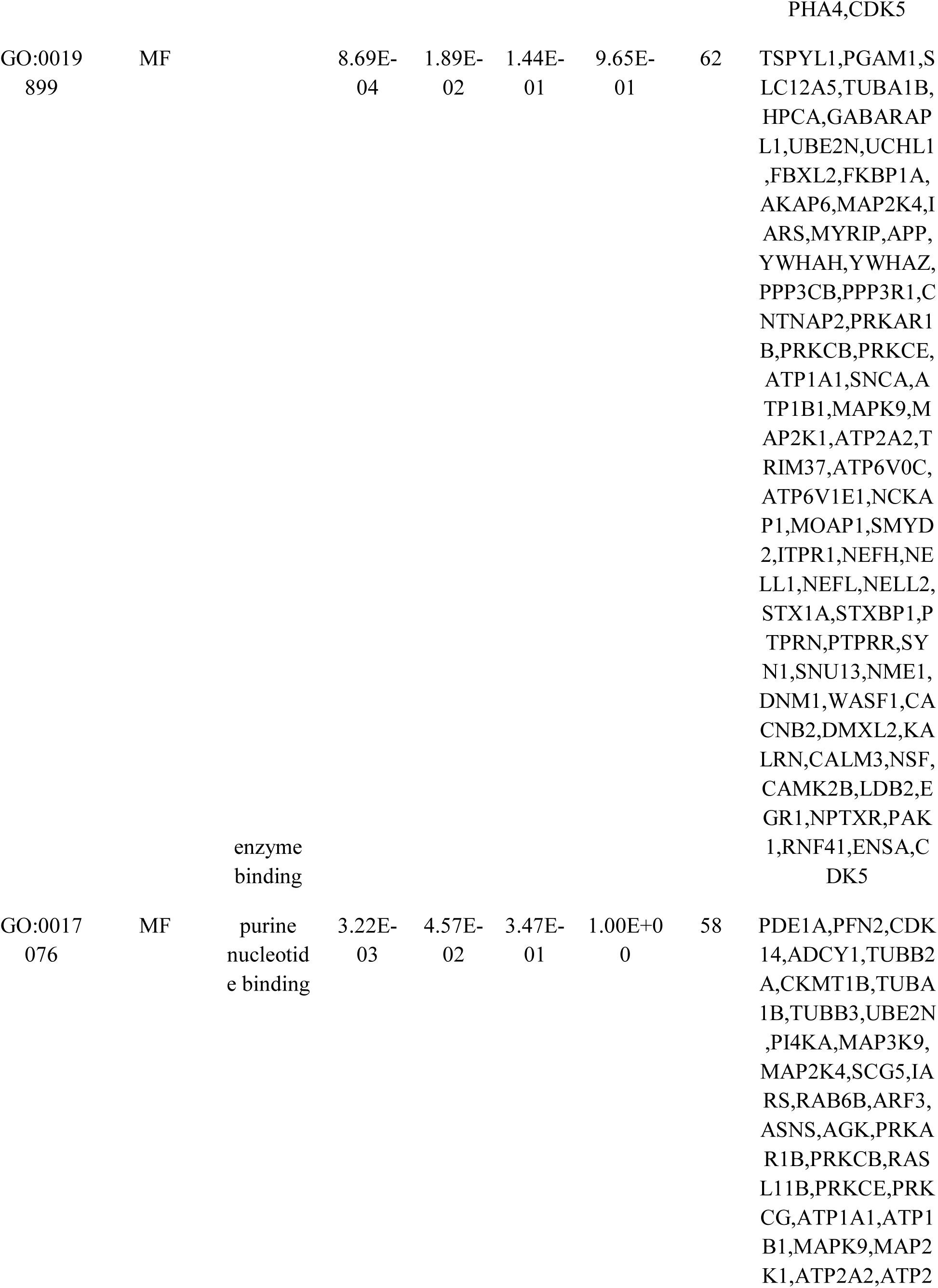
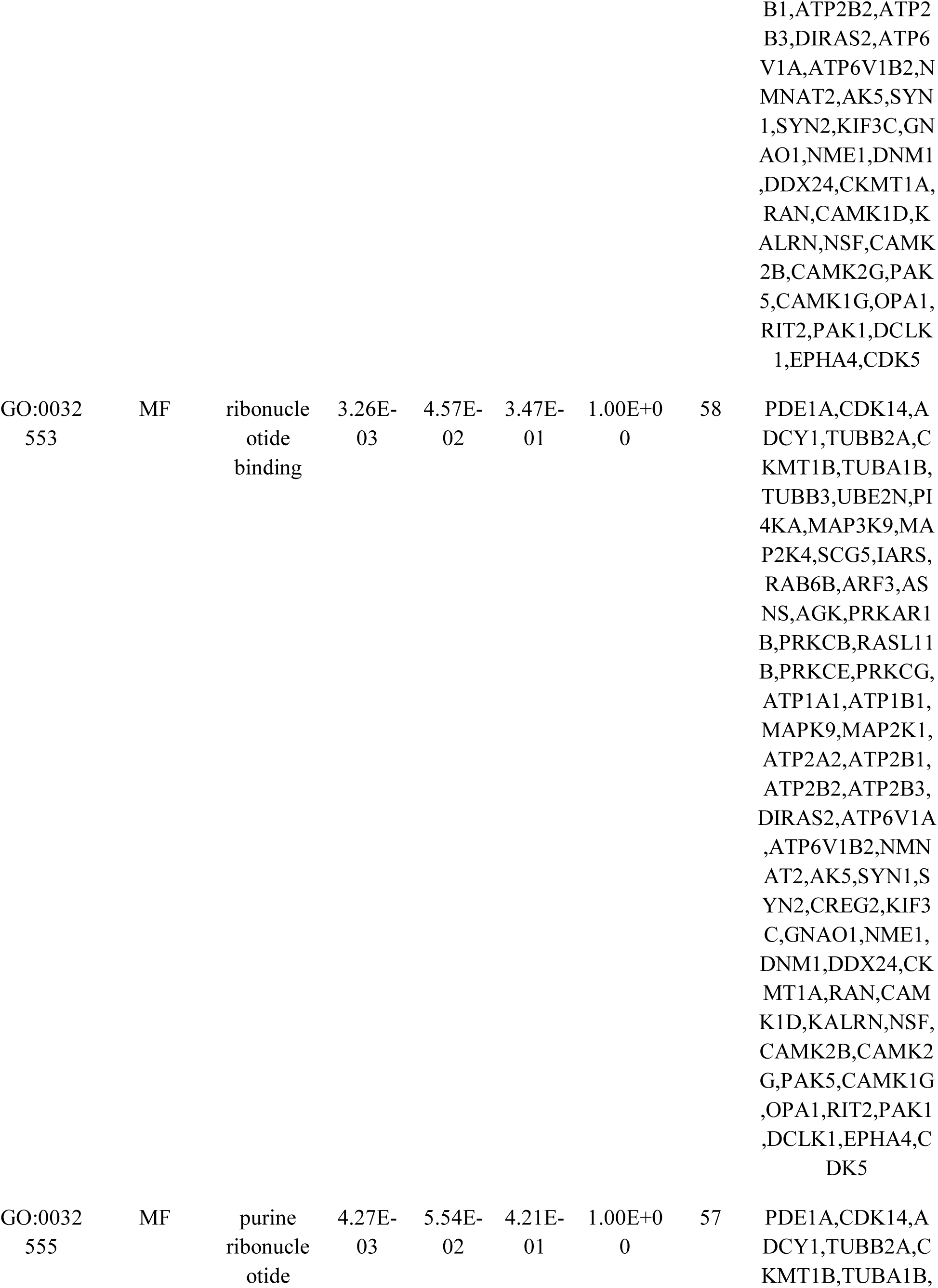
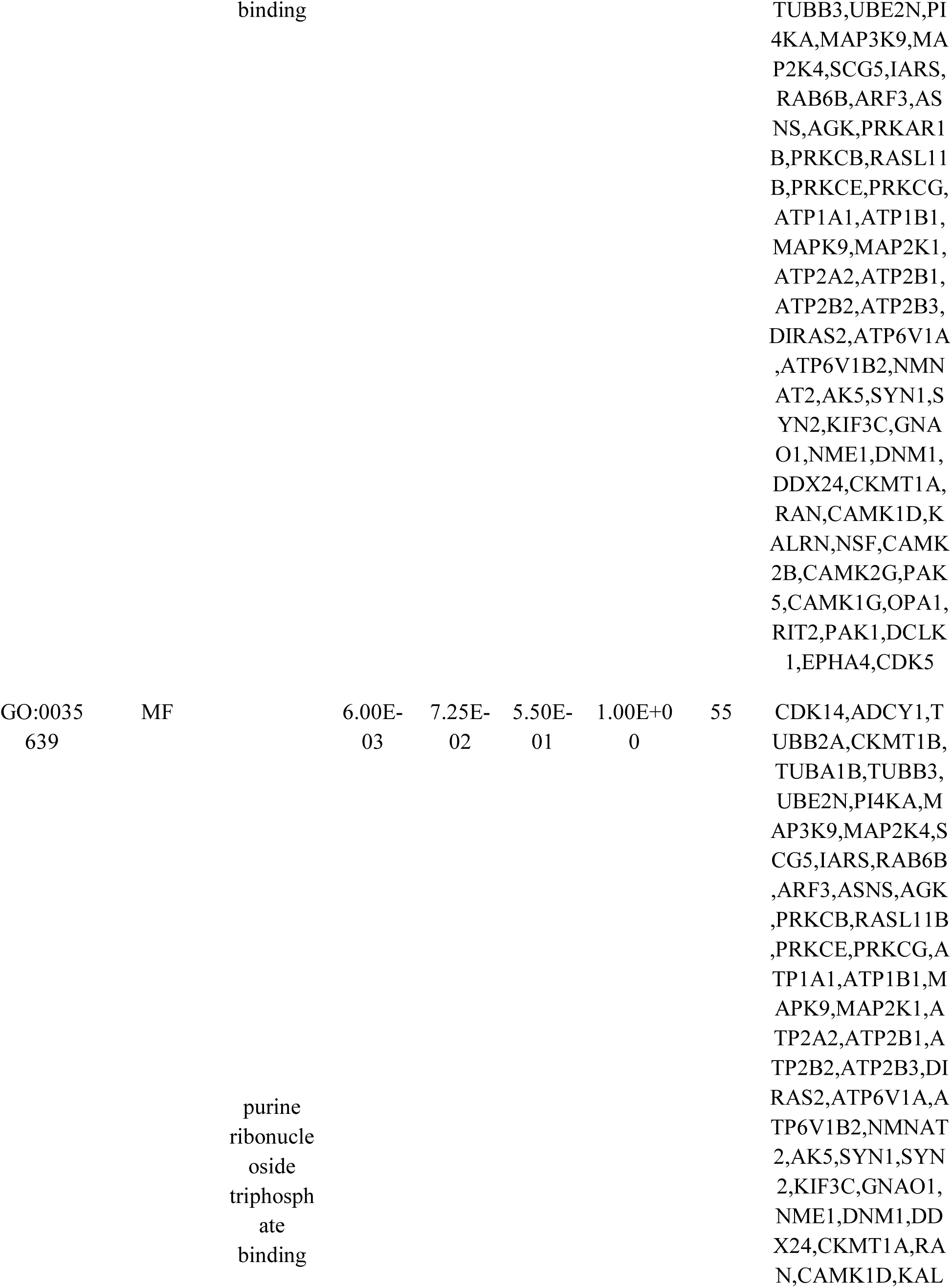
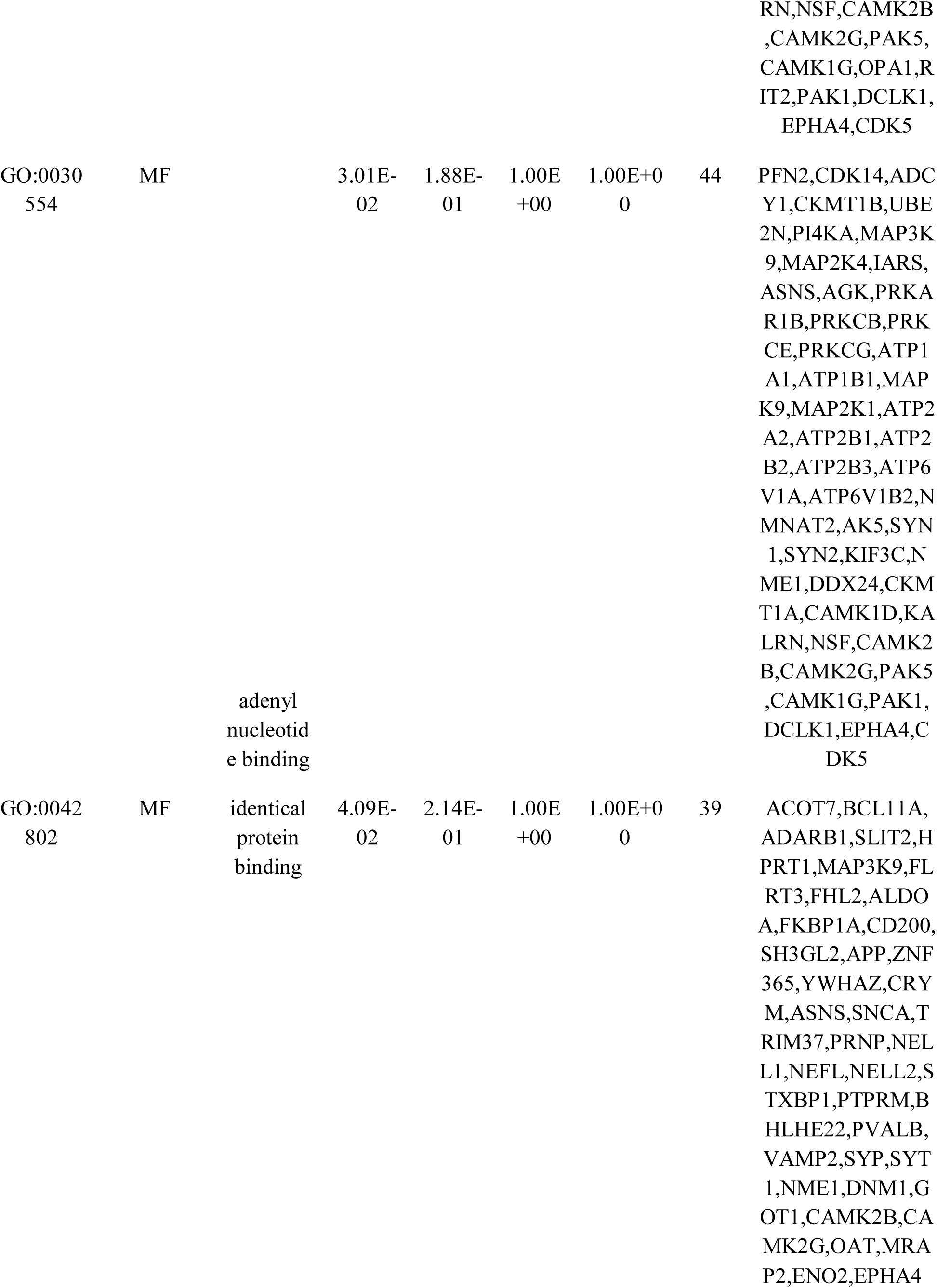
The enriched GO terms of the up regulated differentially expressed genes

**Table 5.**
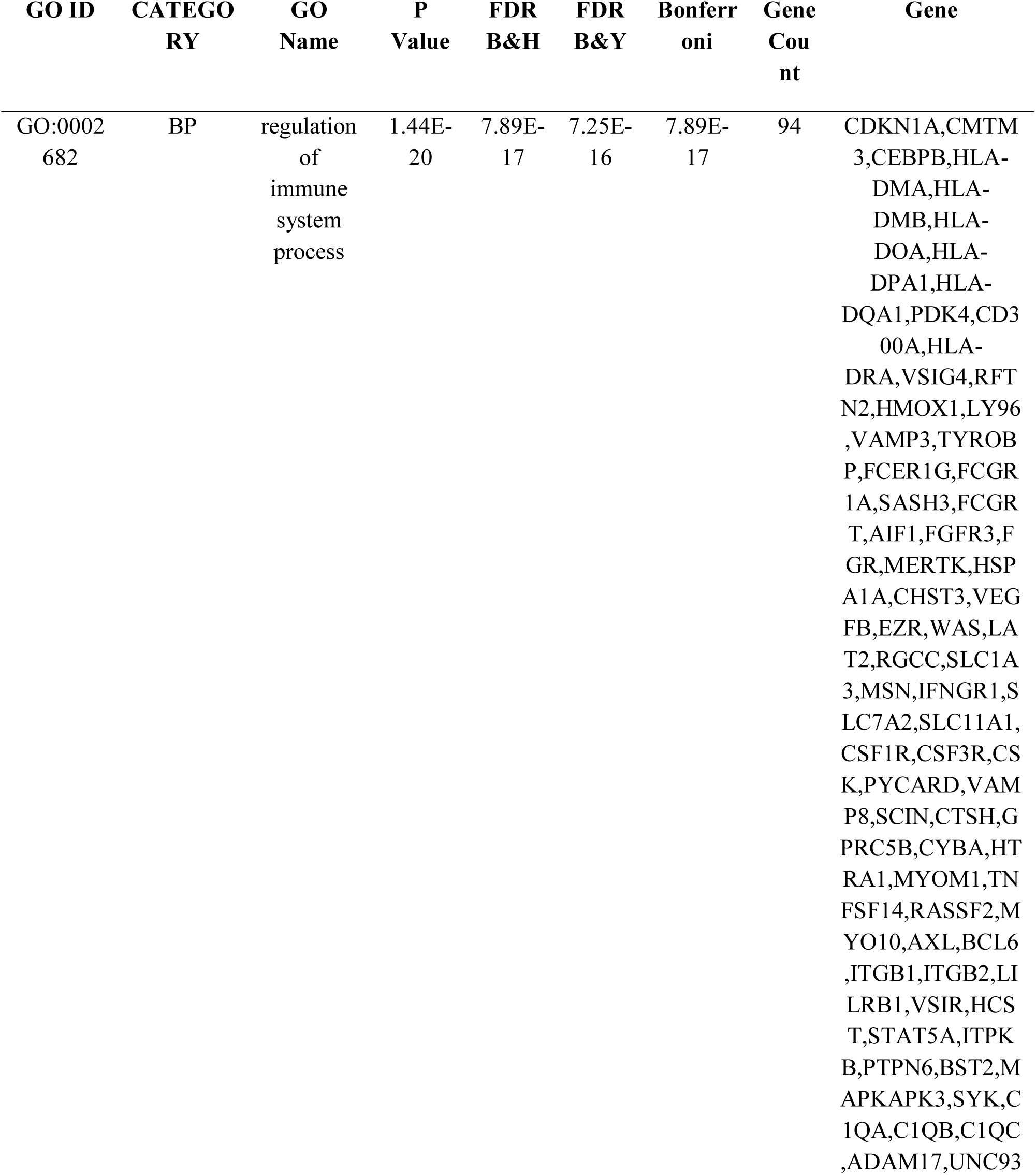
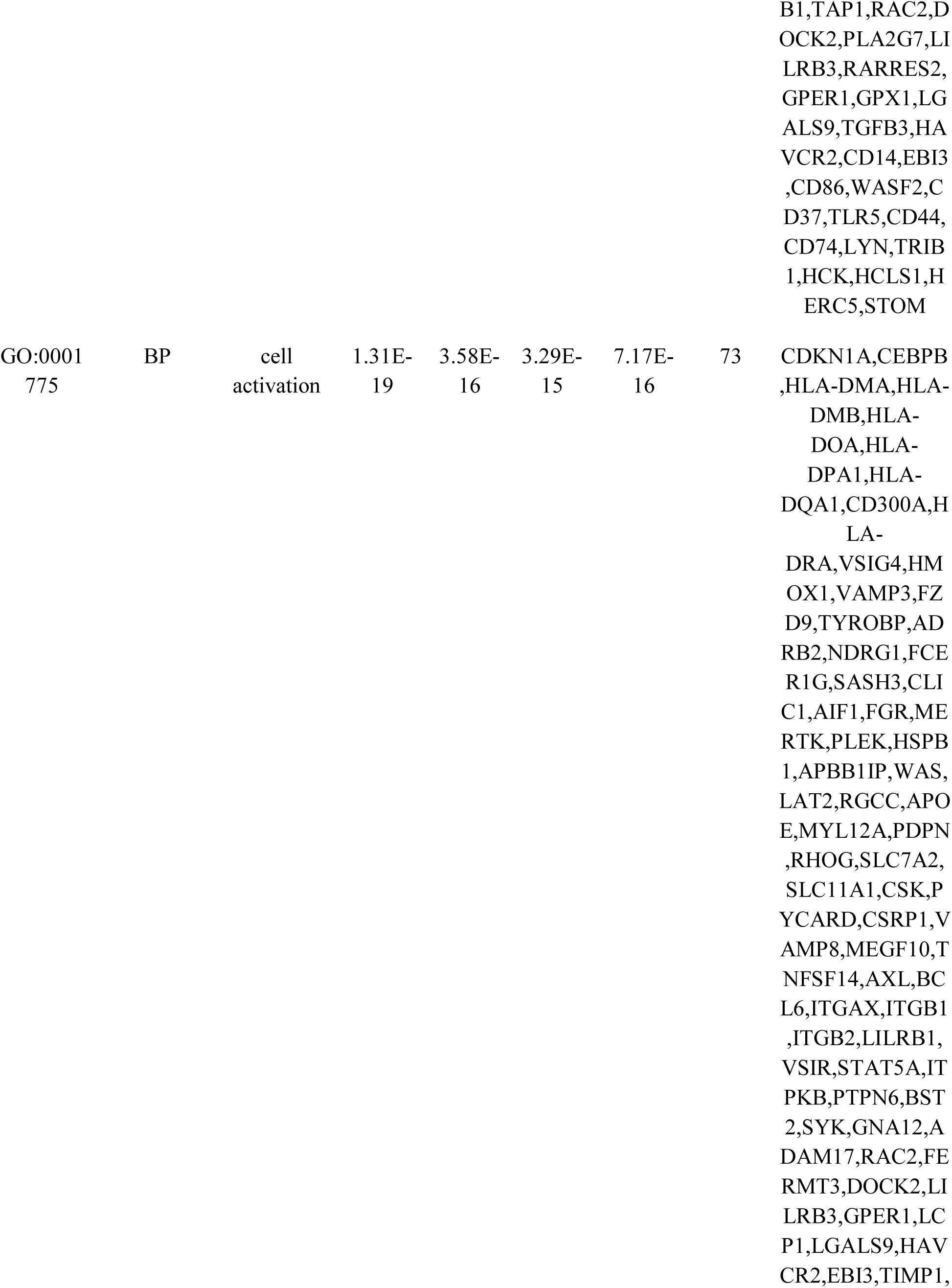
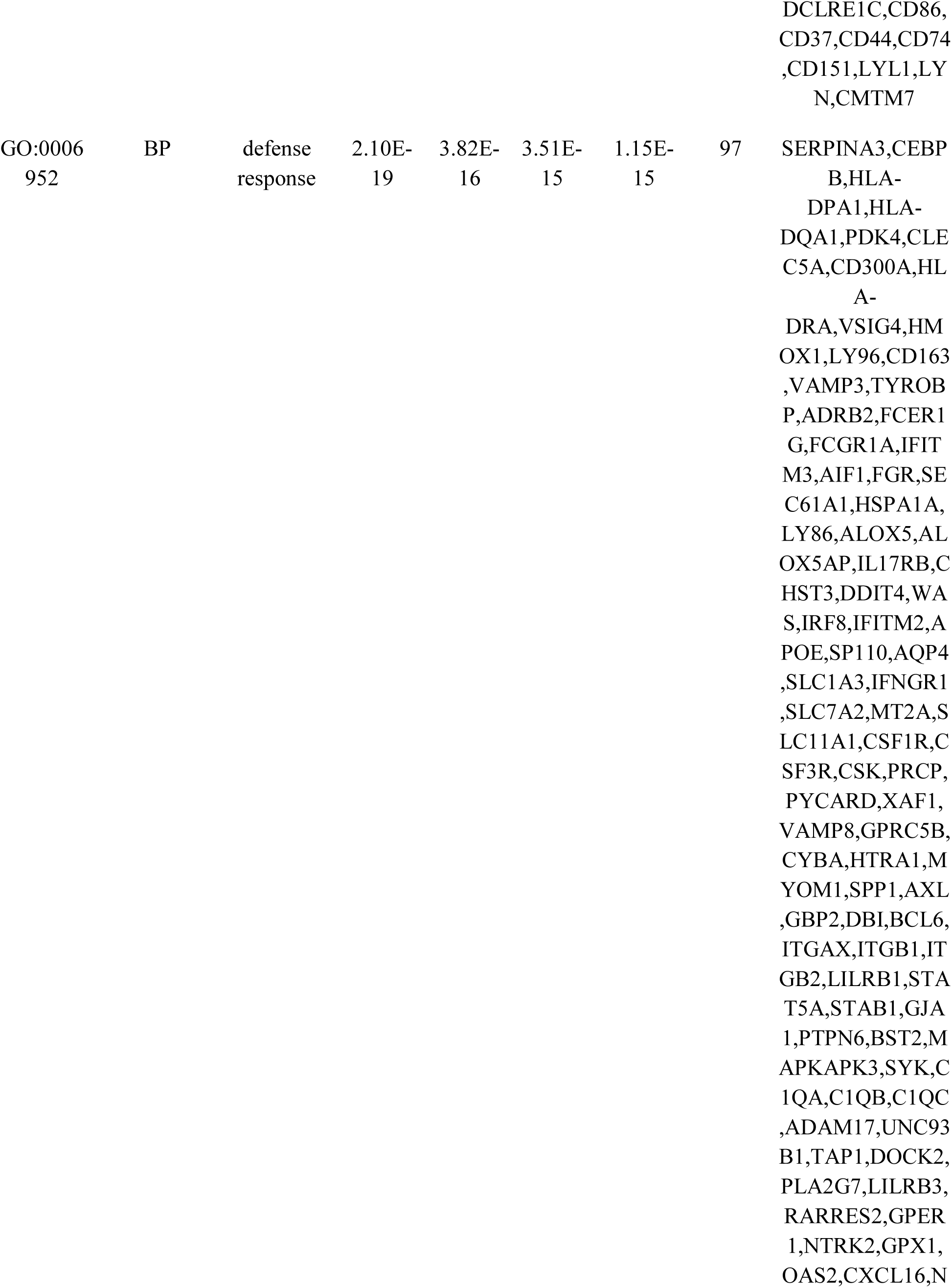
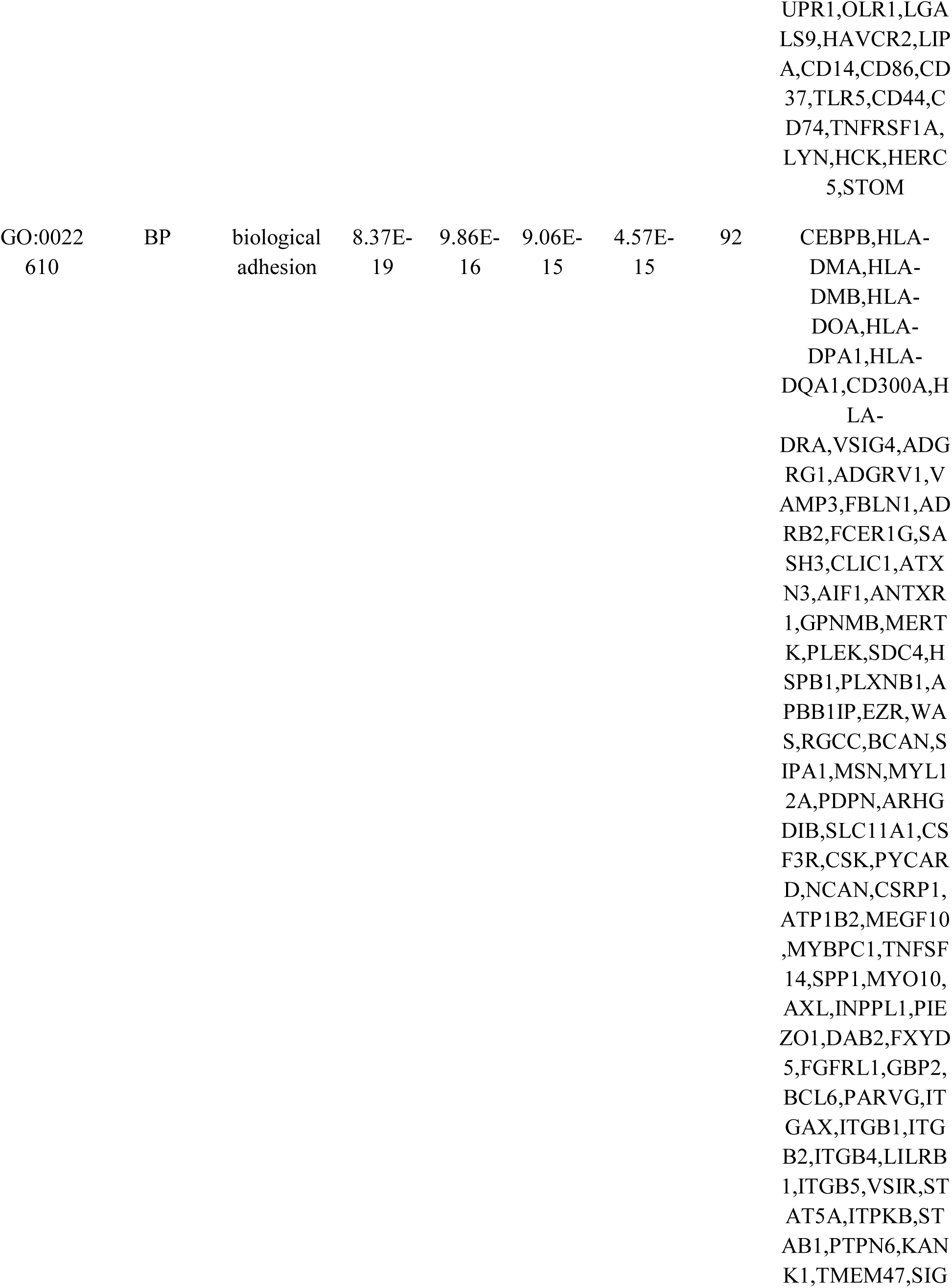
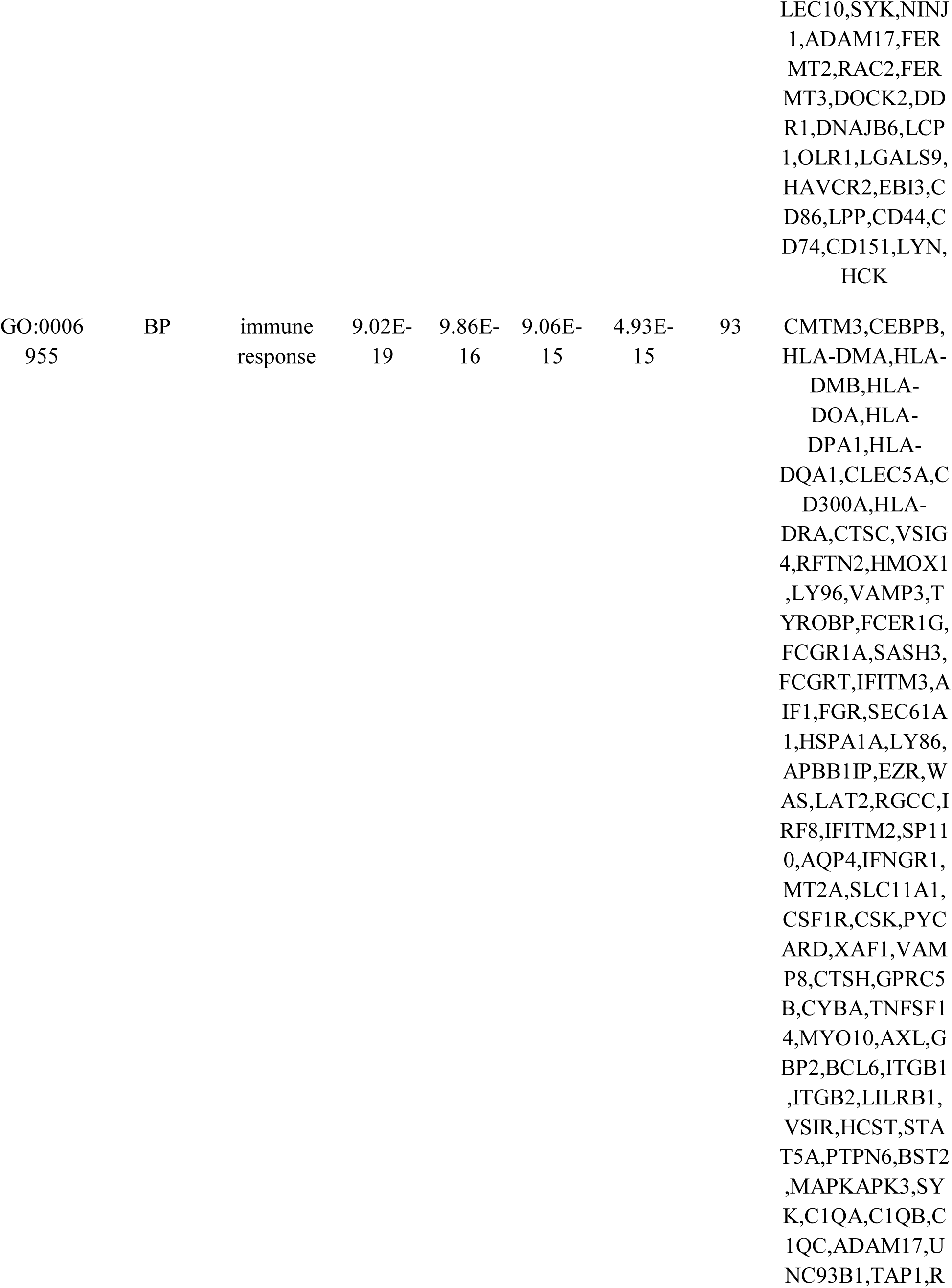
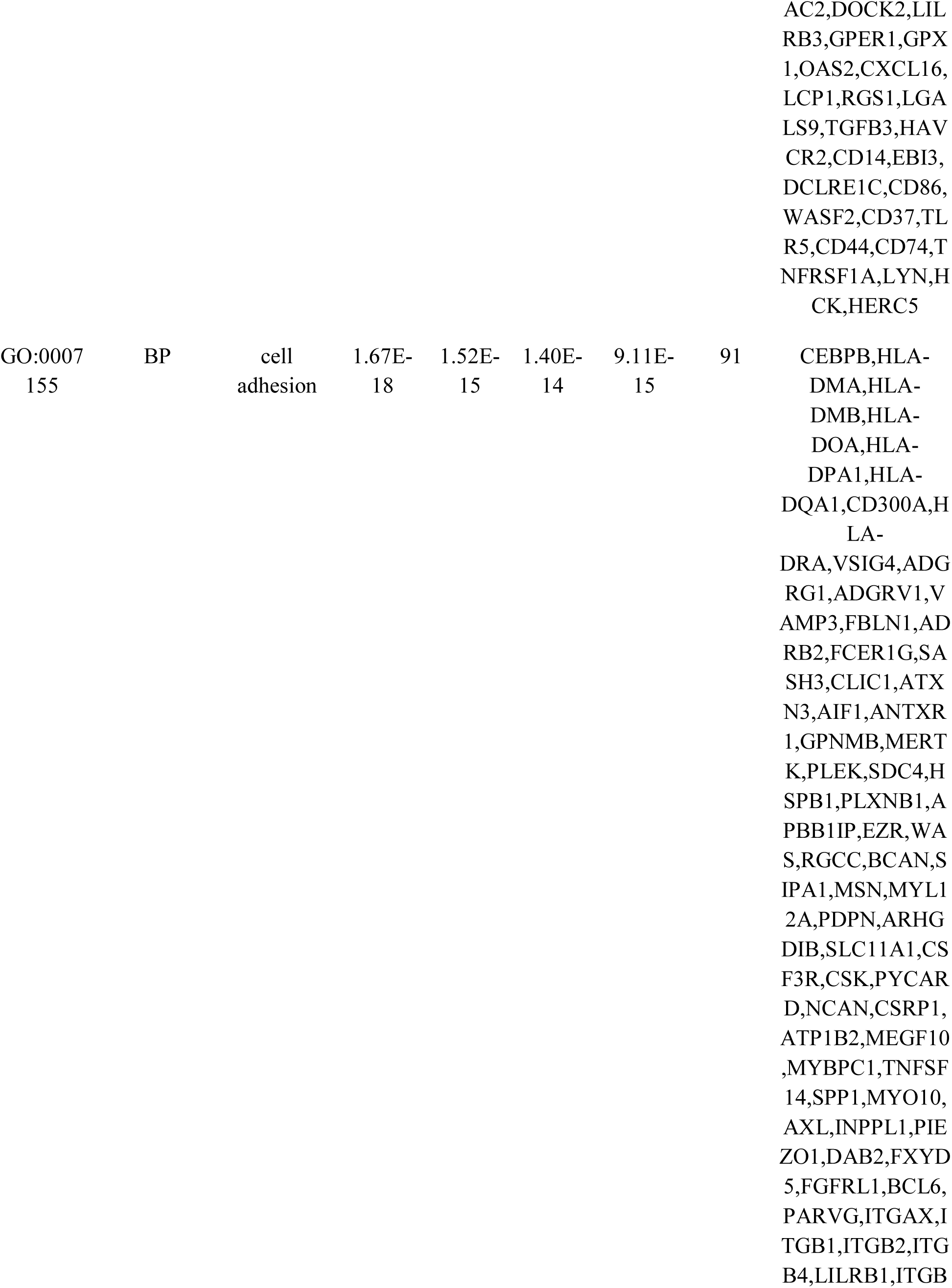
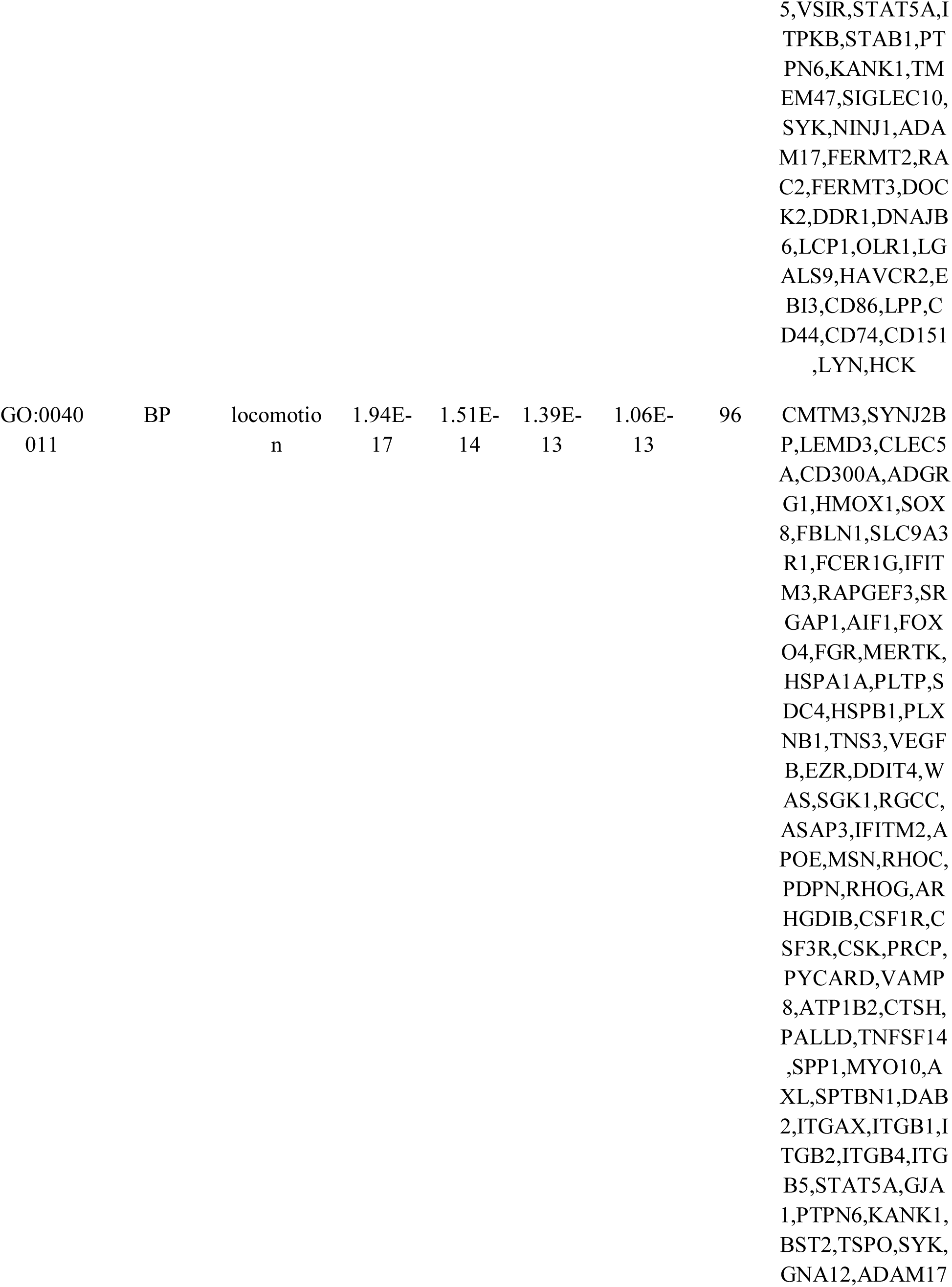
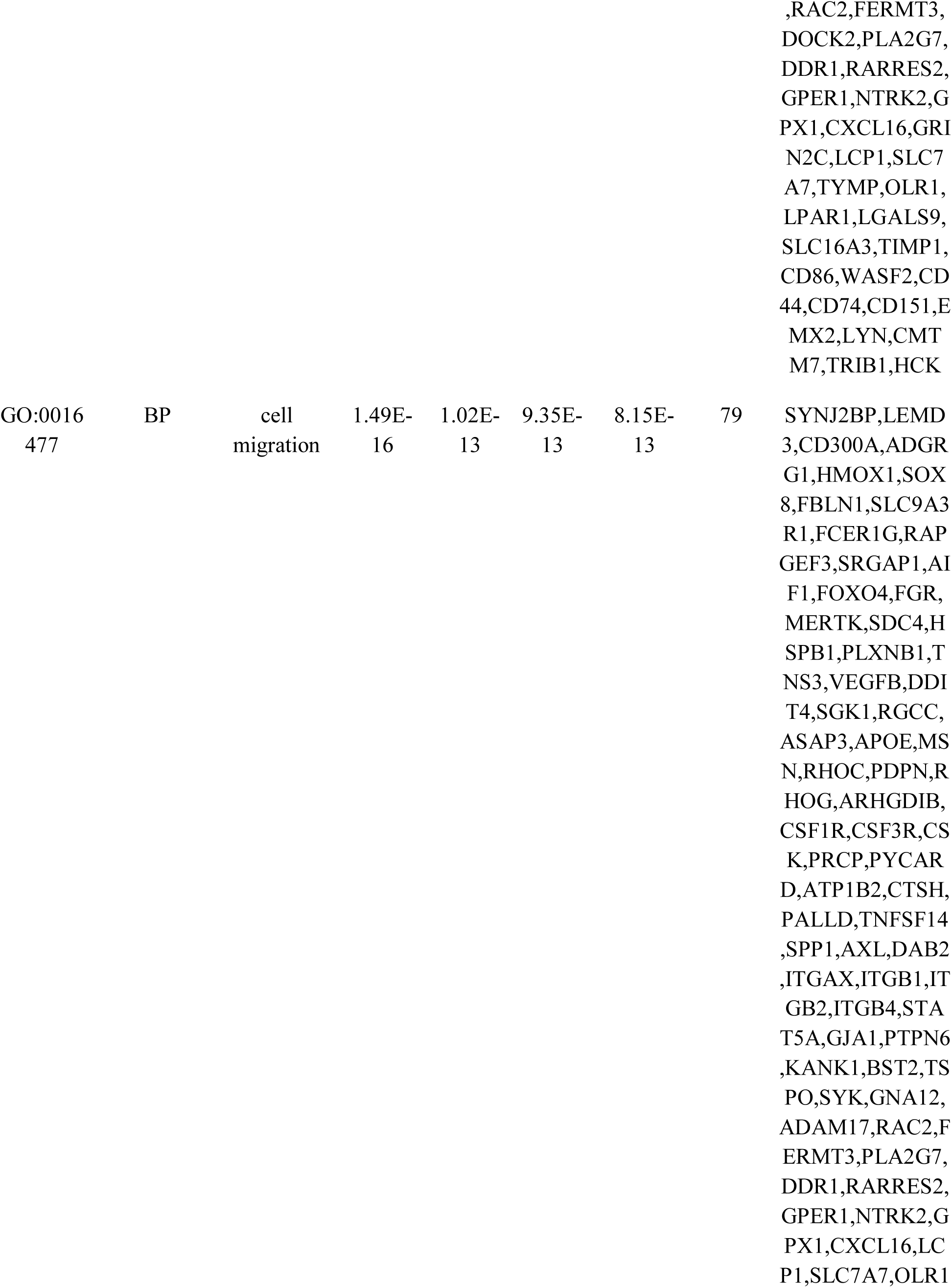
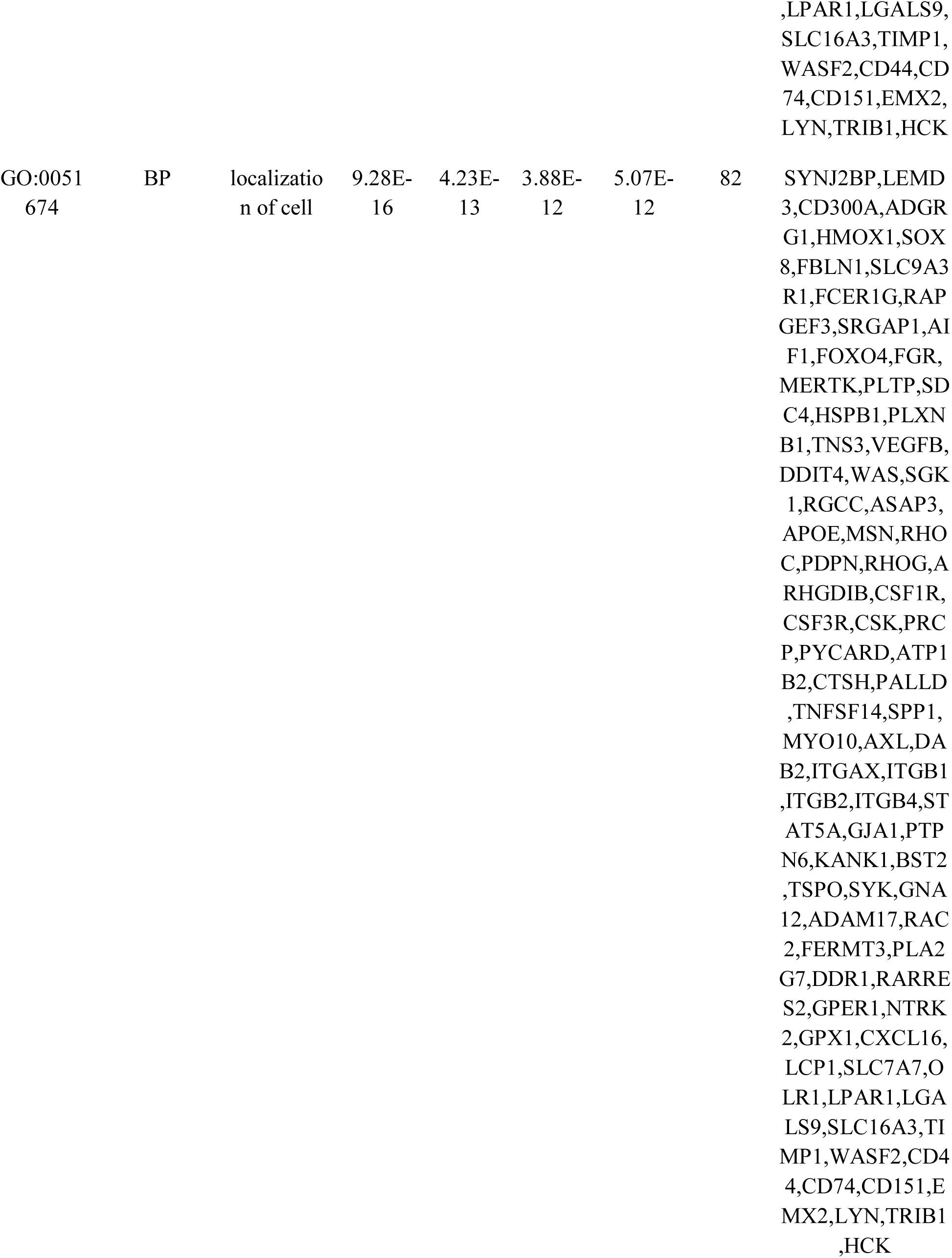
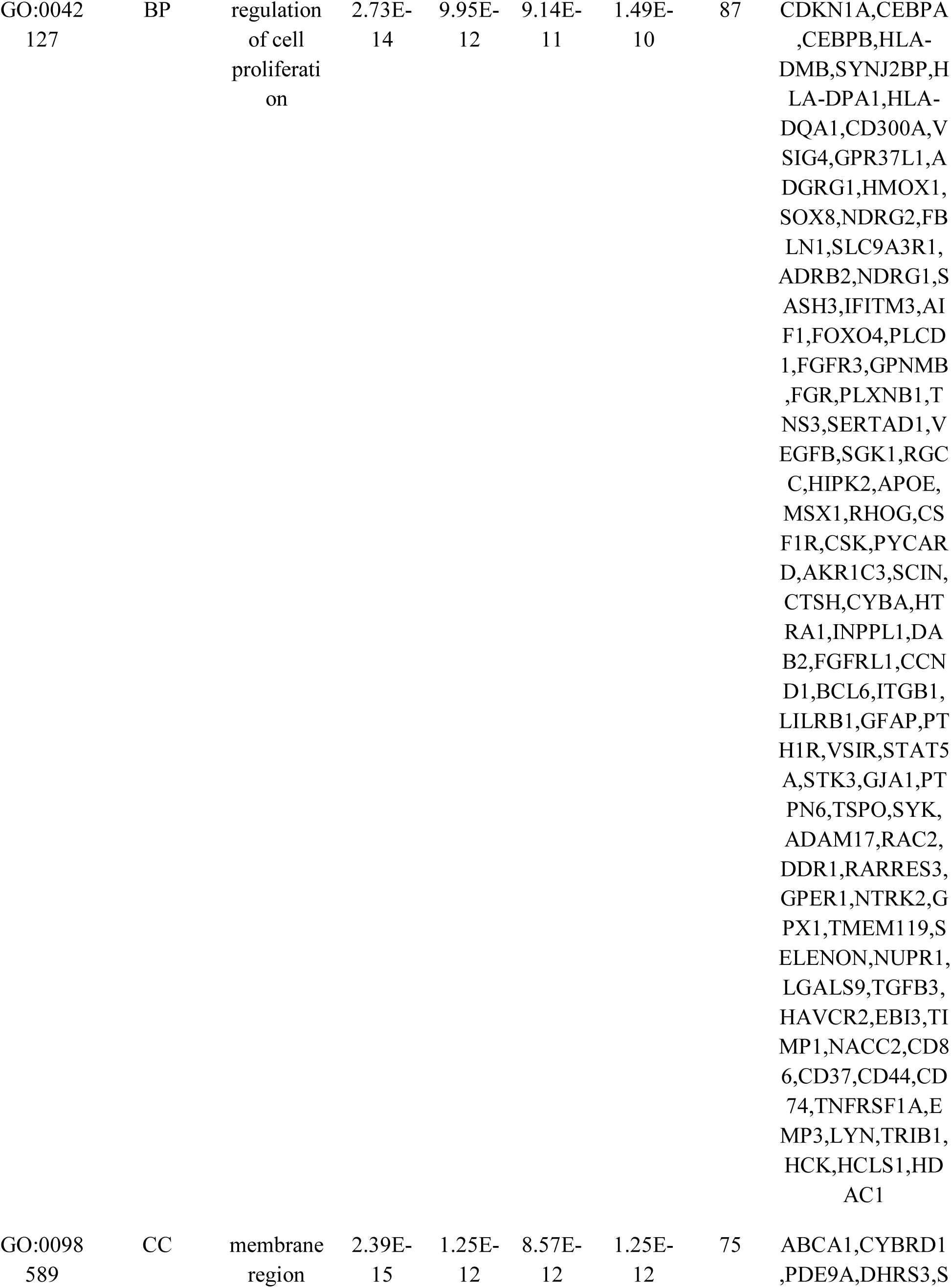
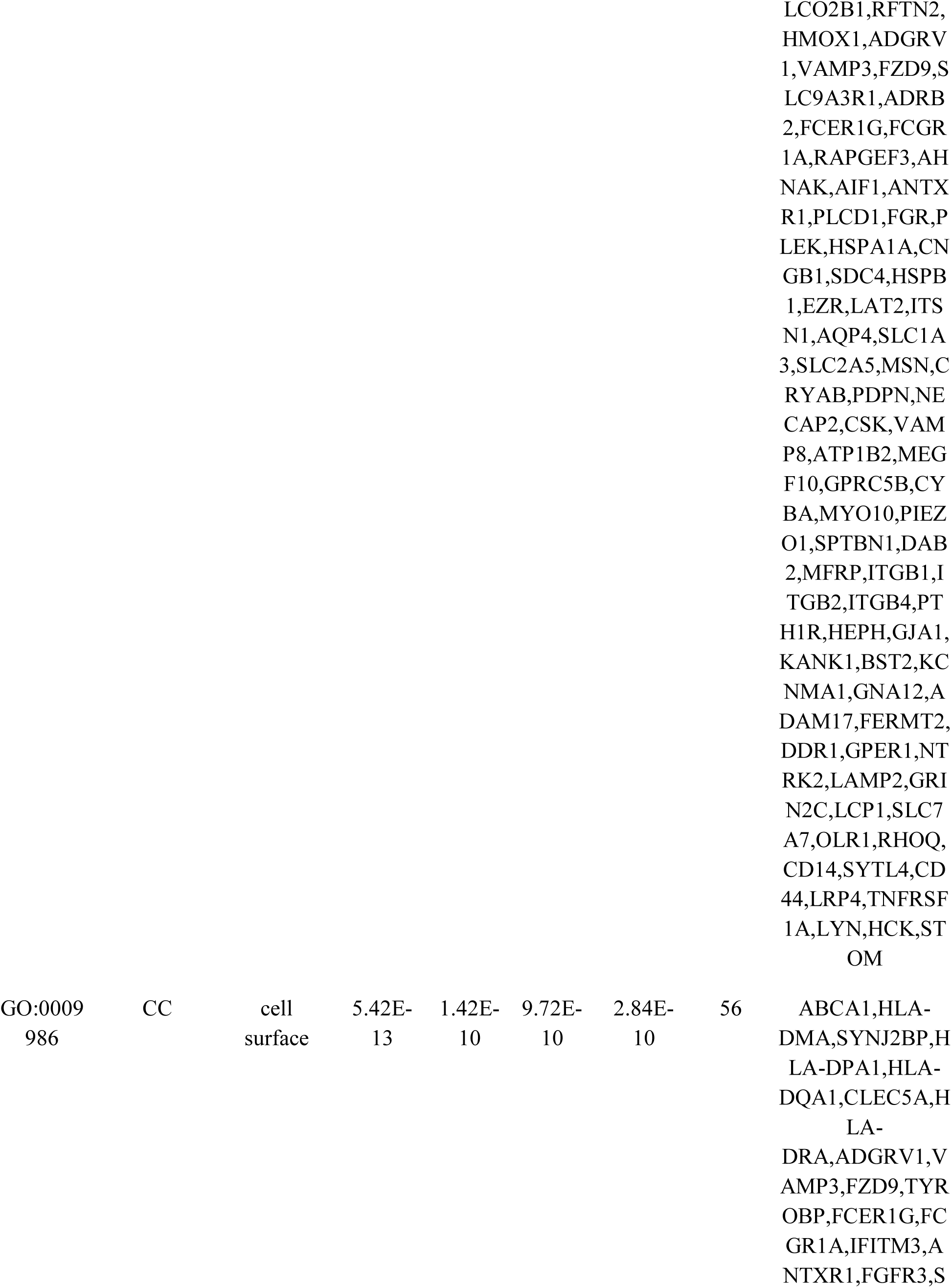
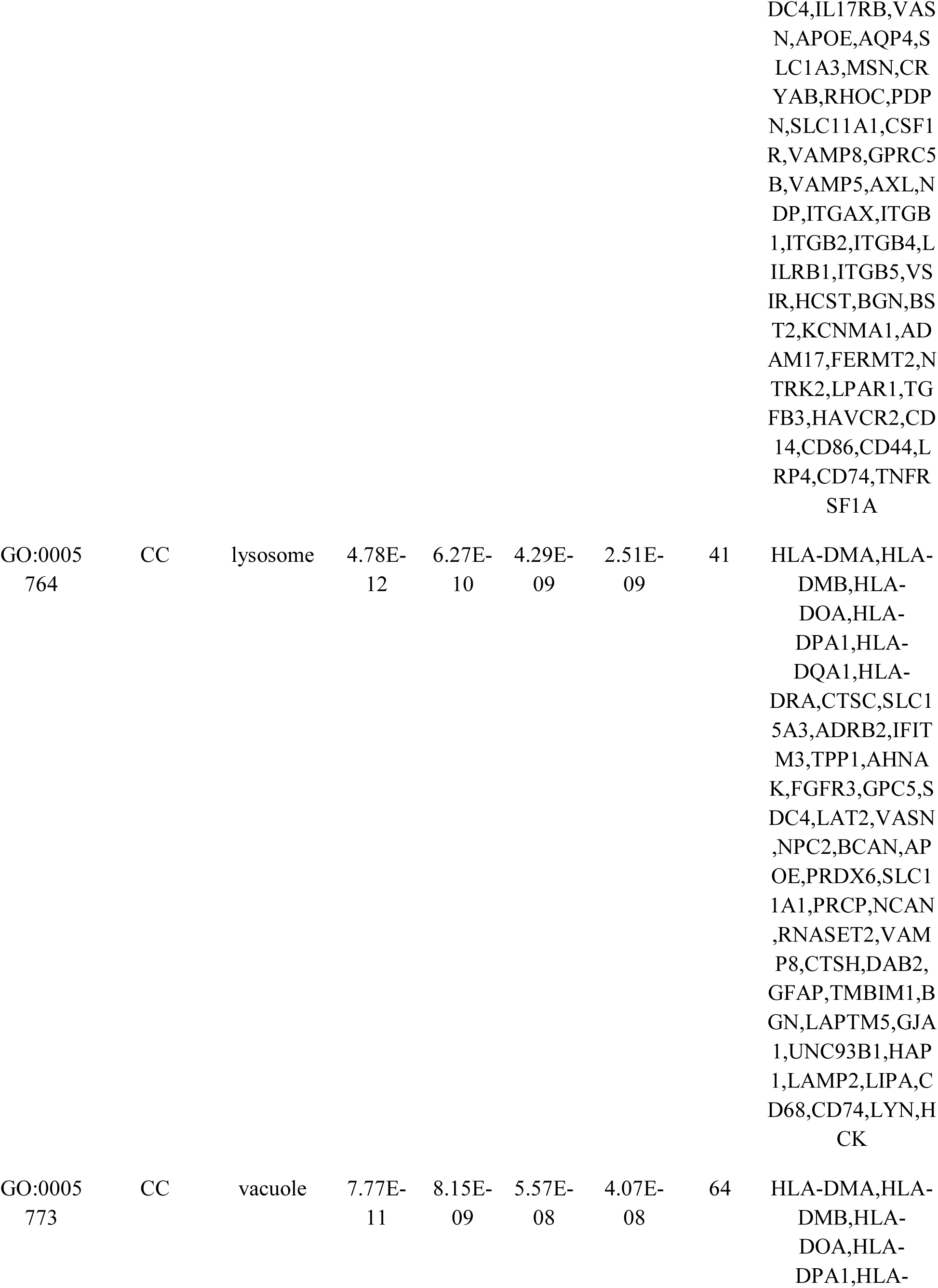
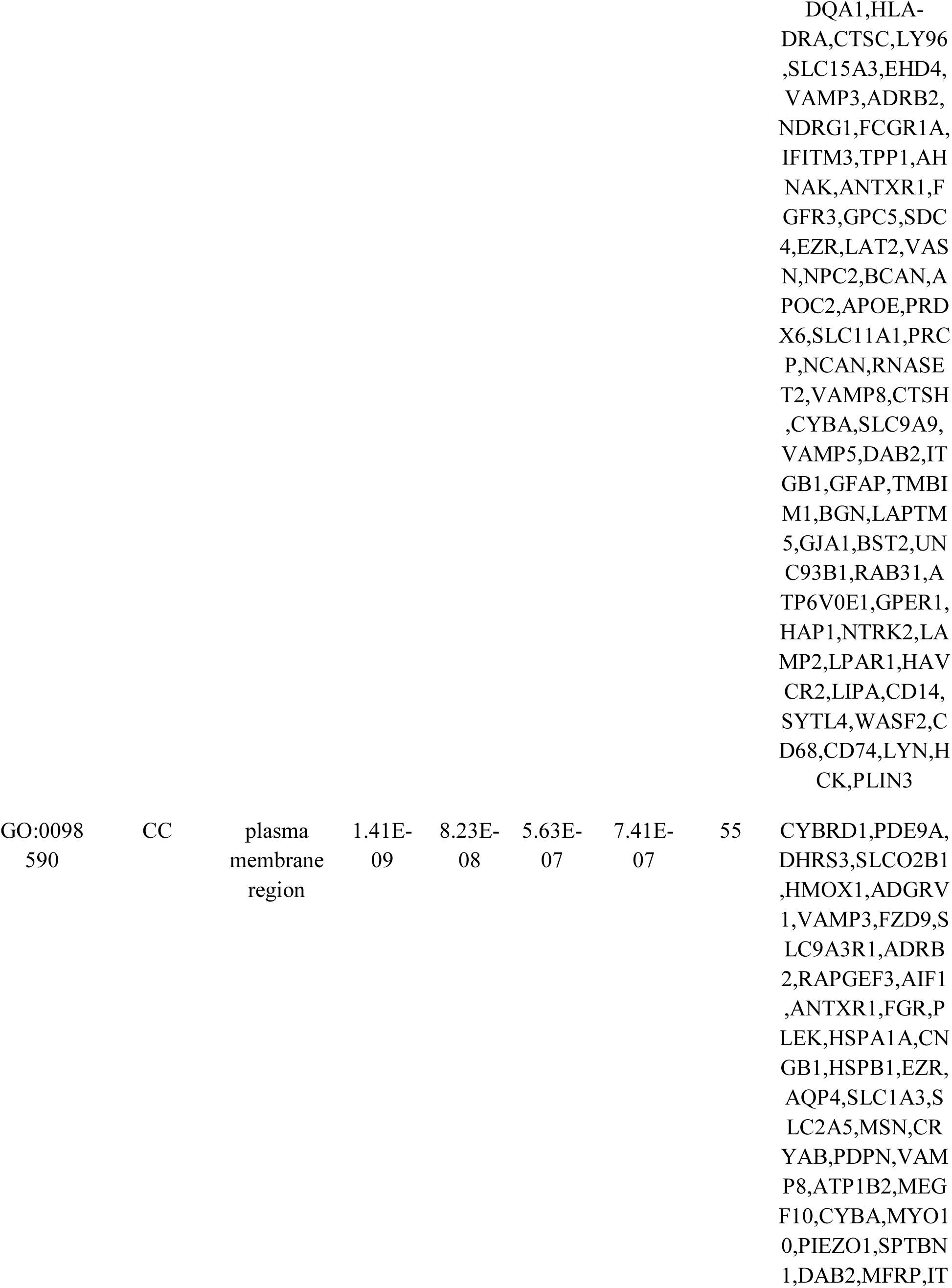
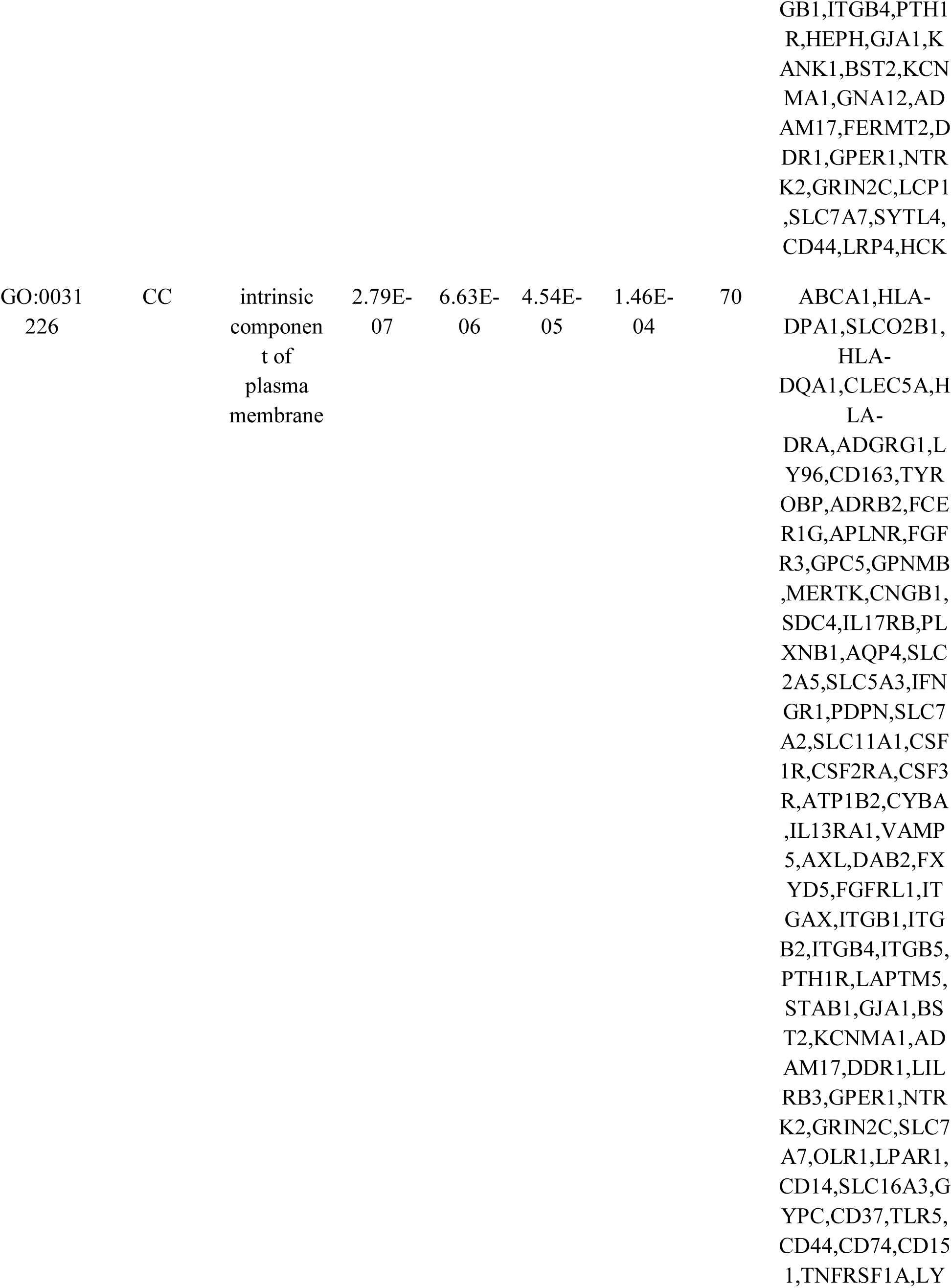
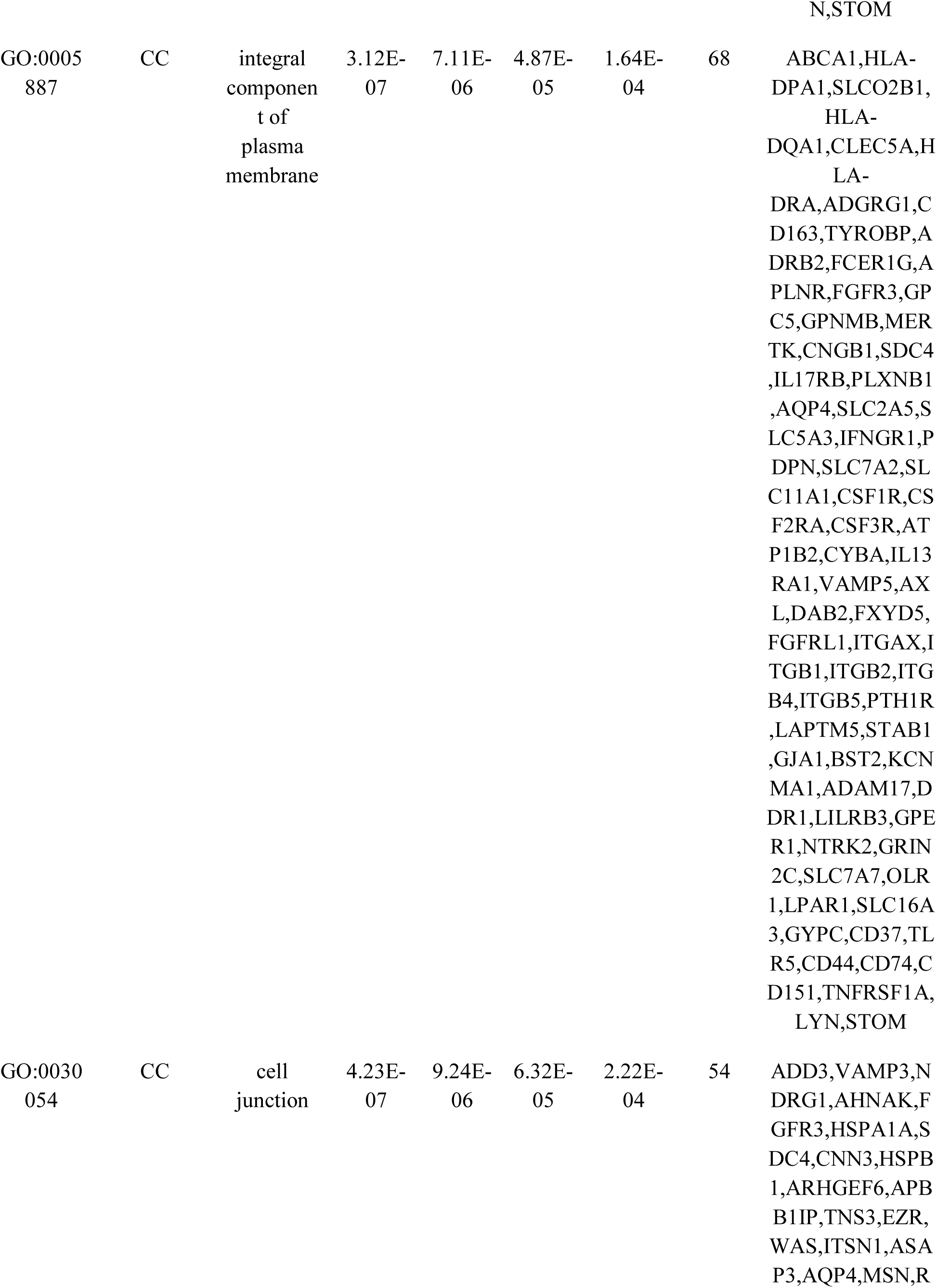
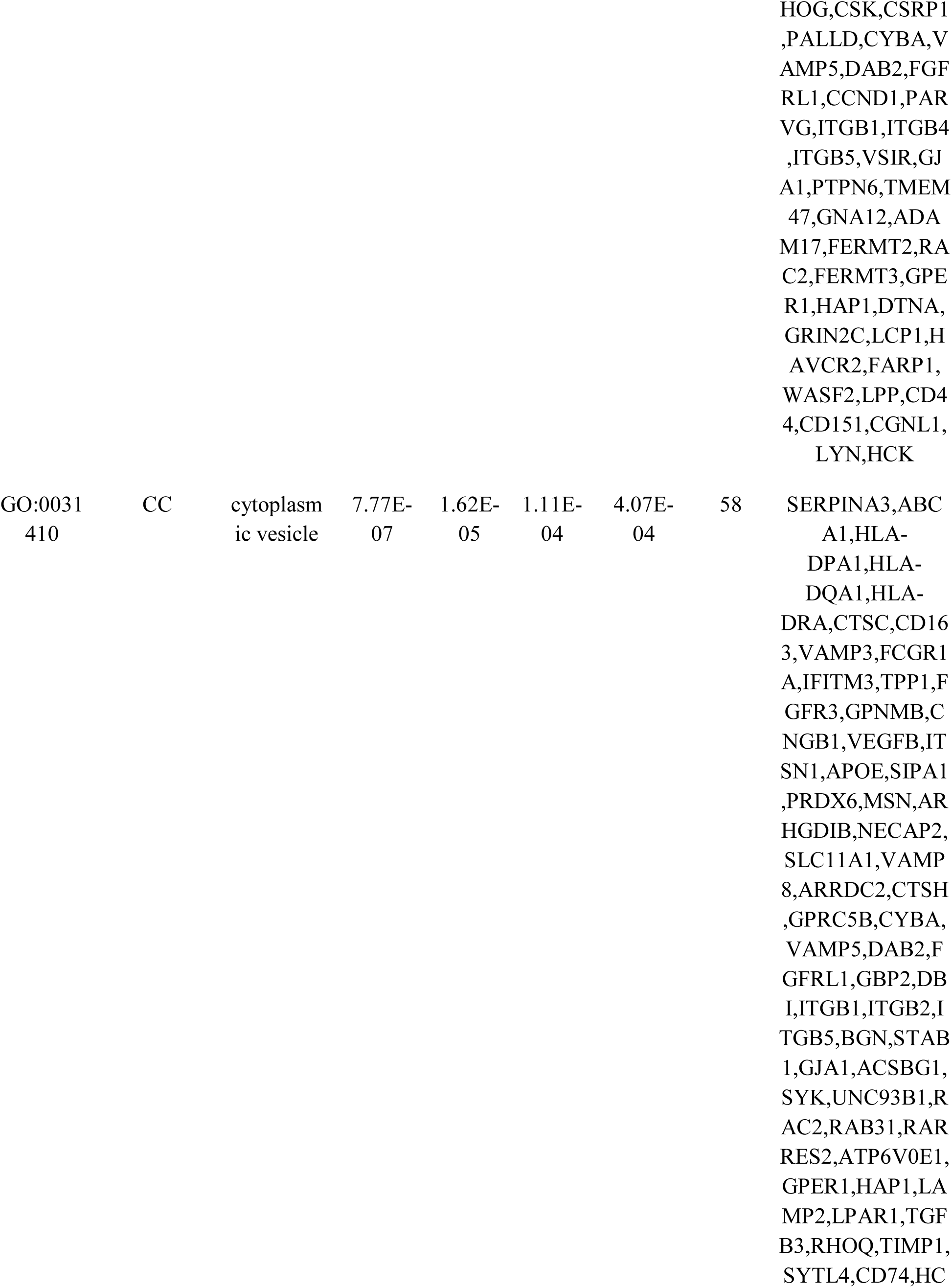
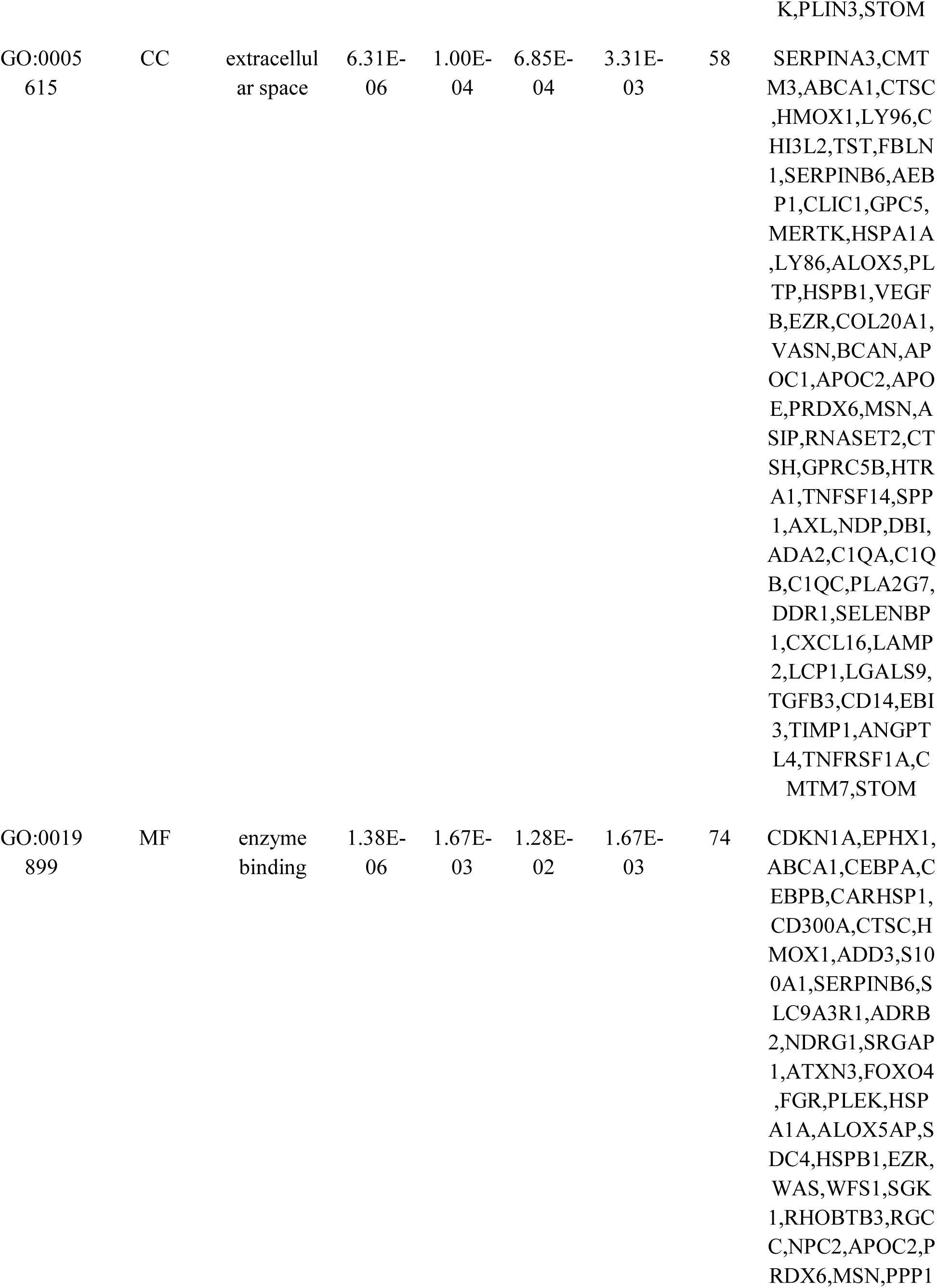
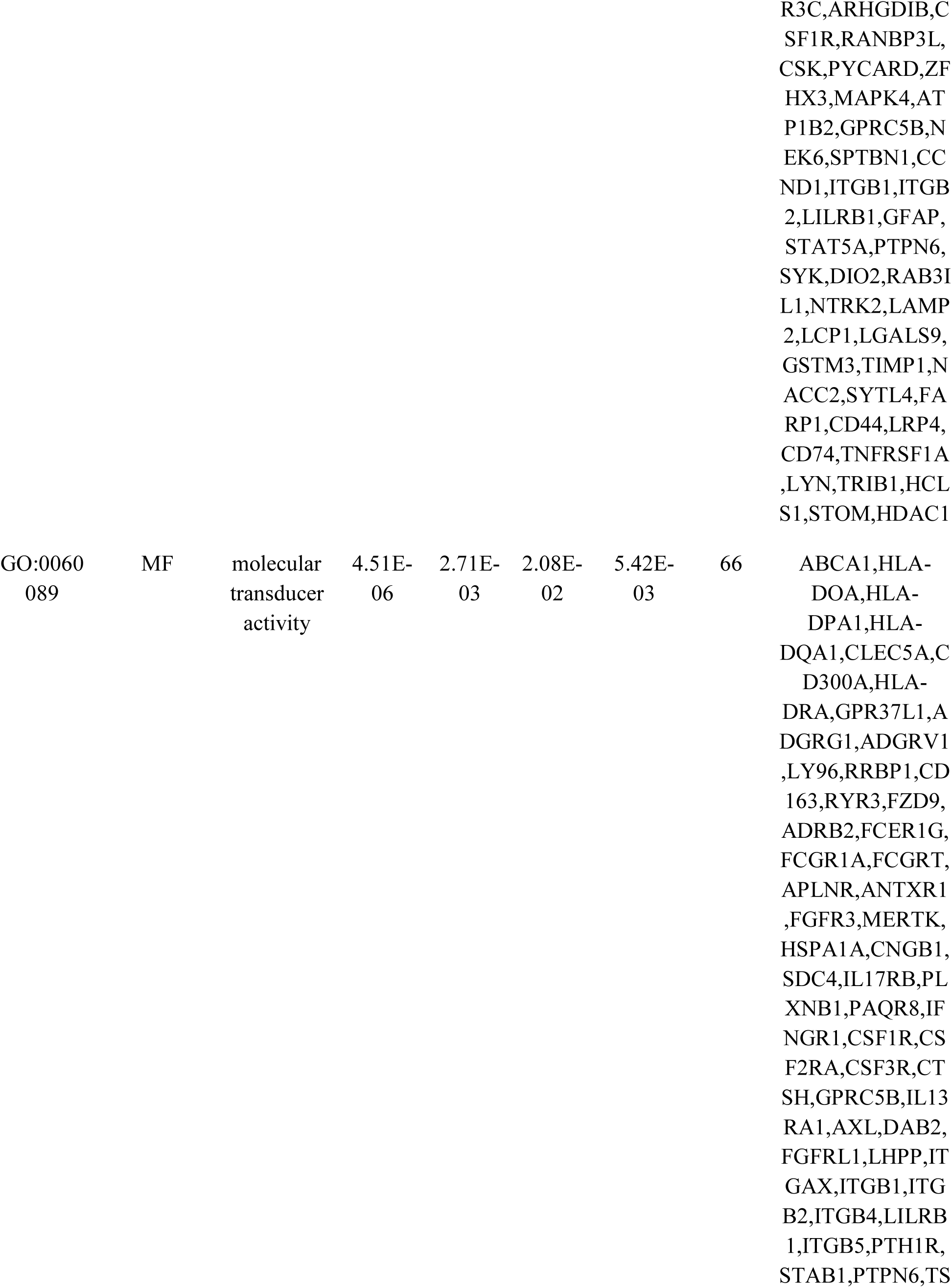
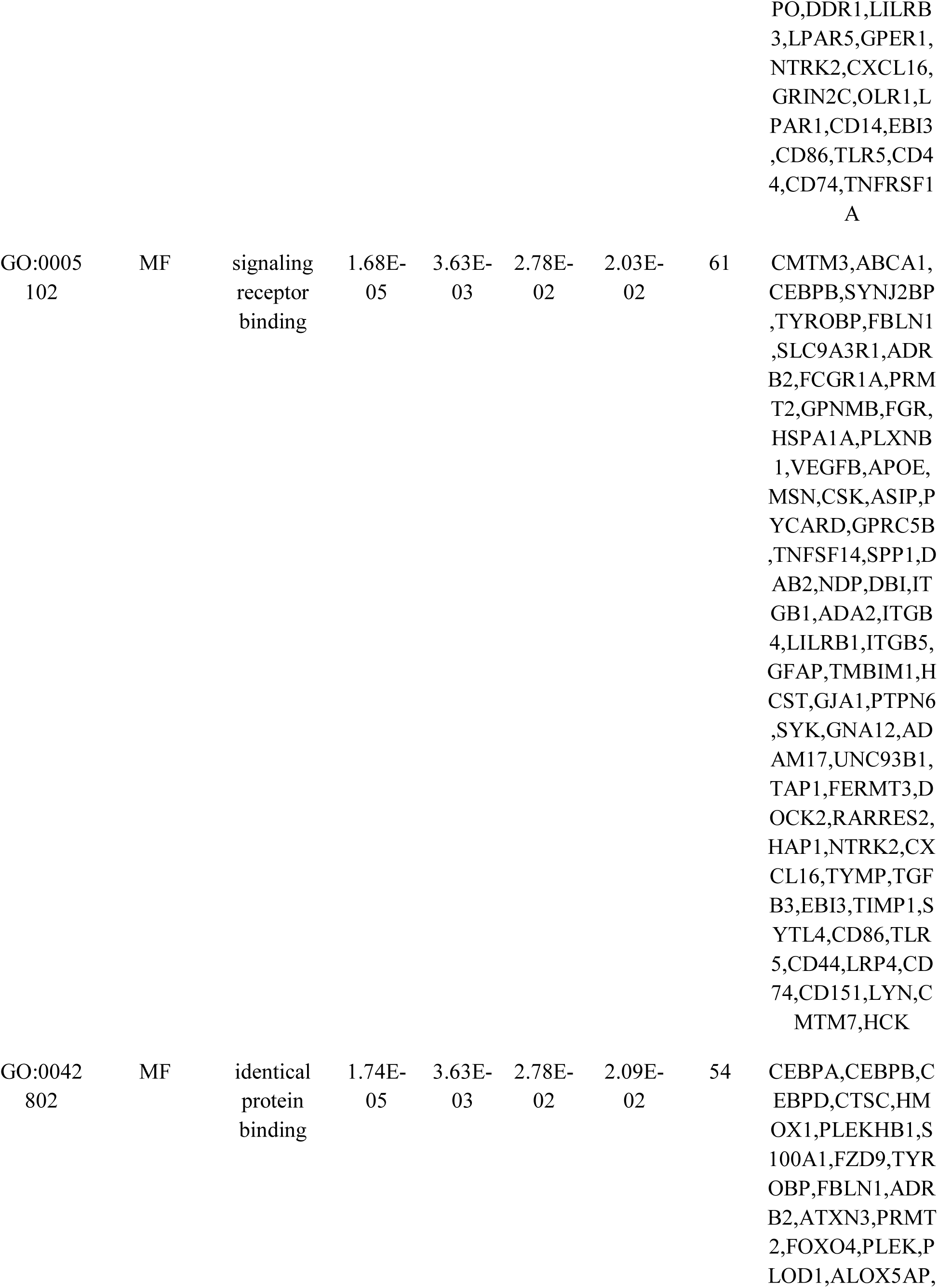
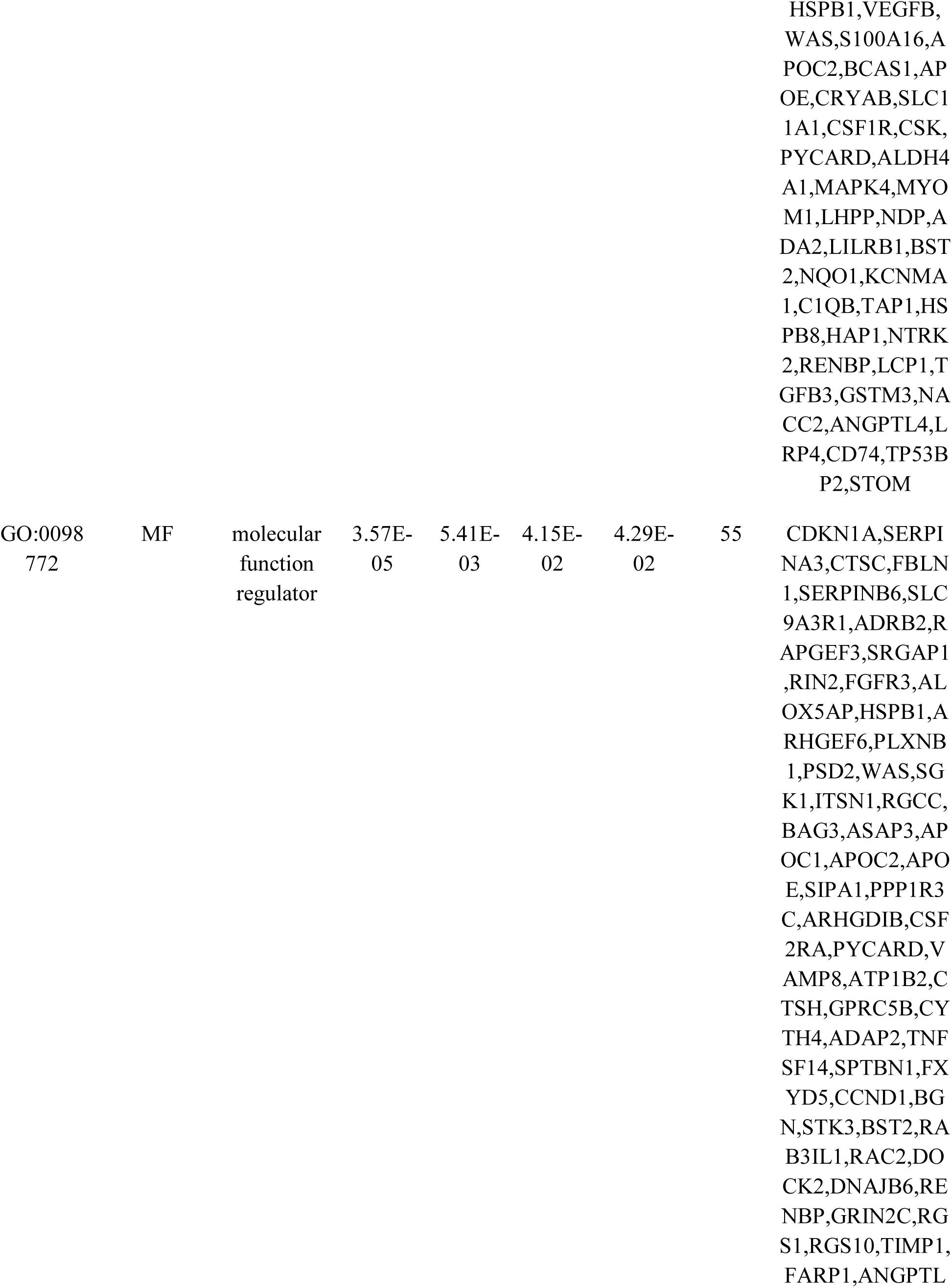
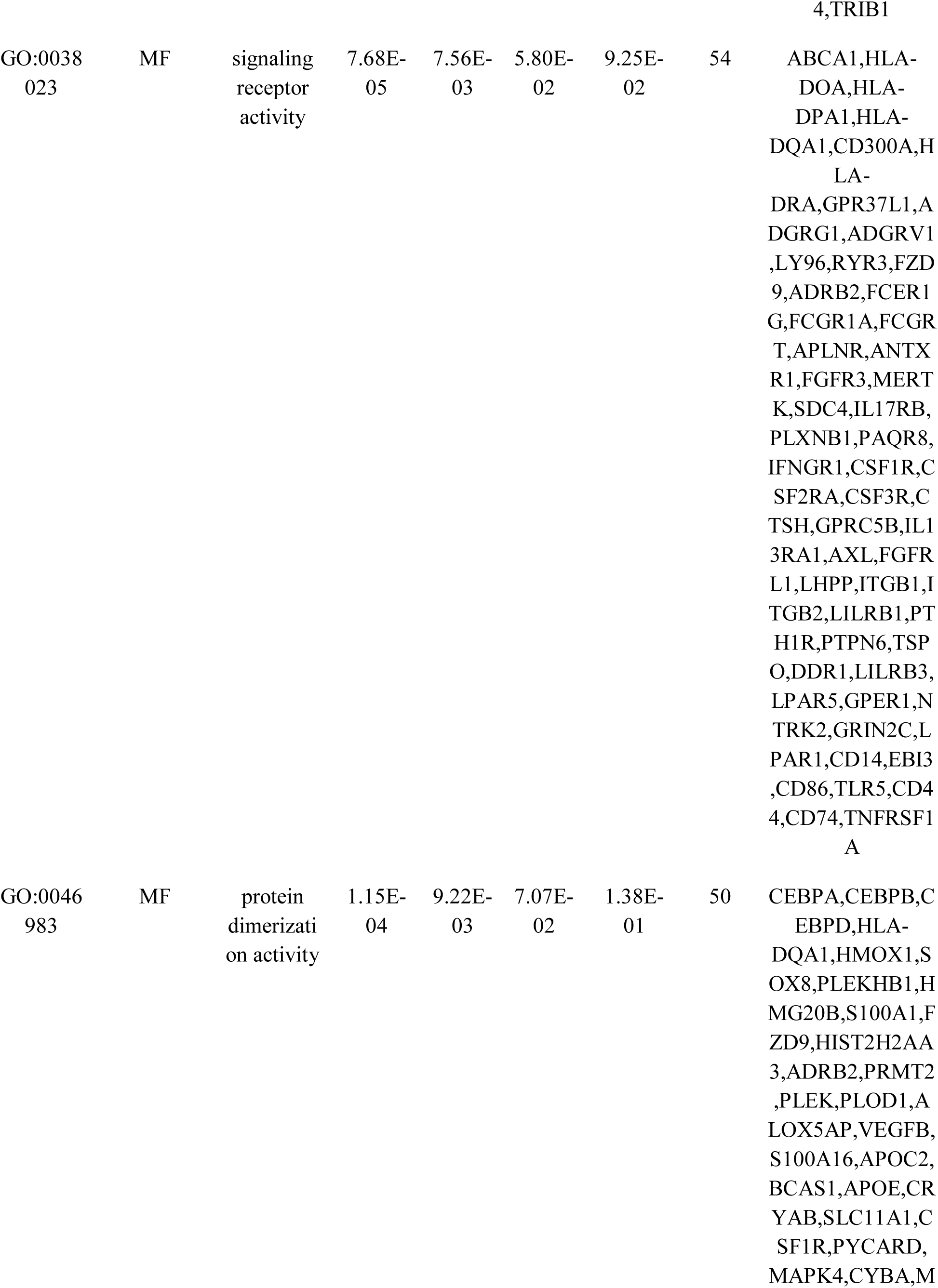
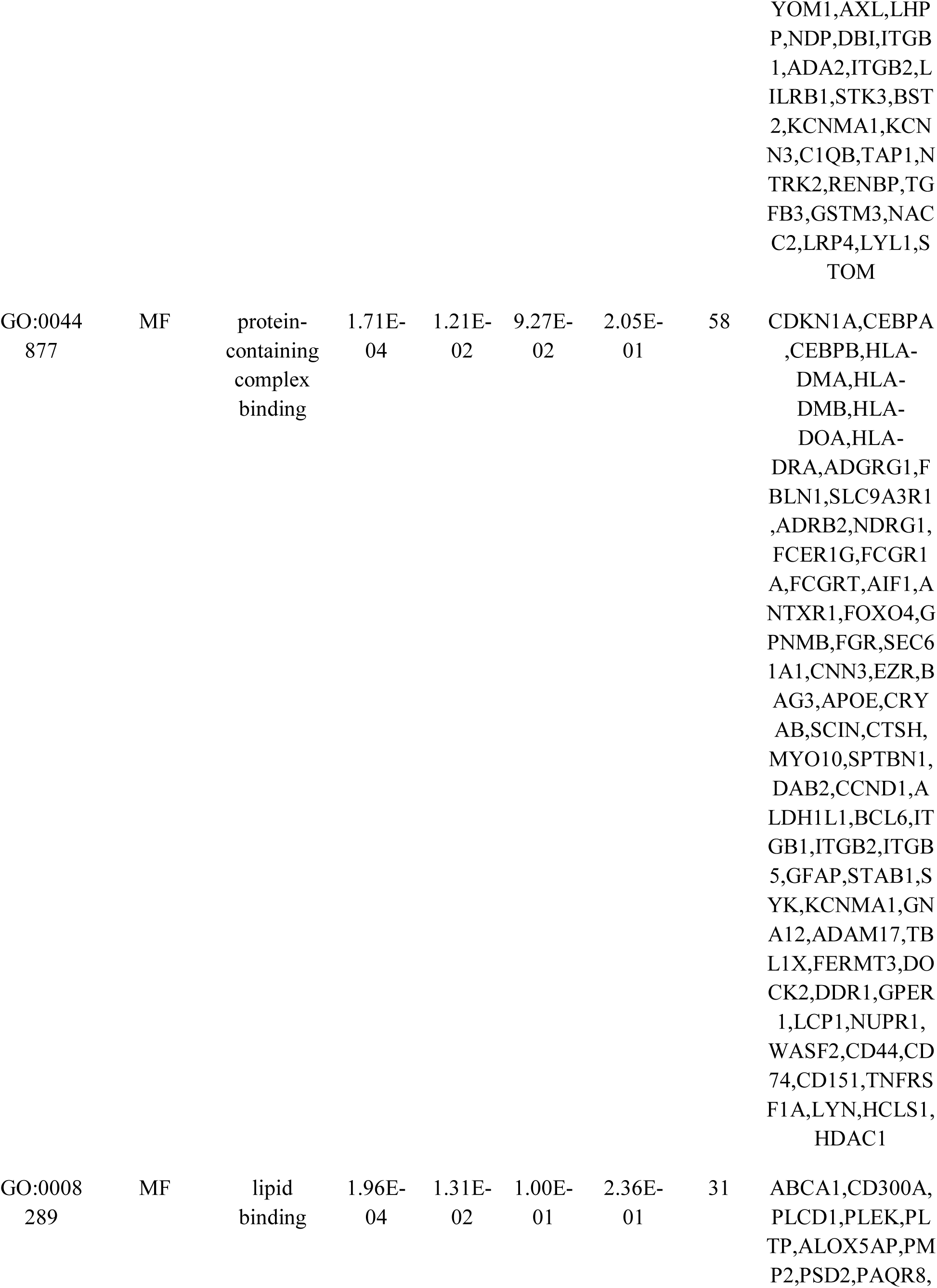
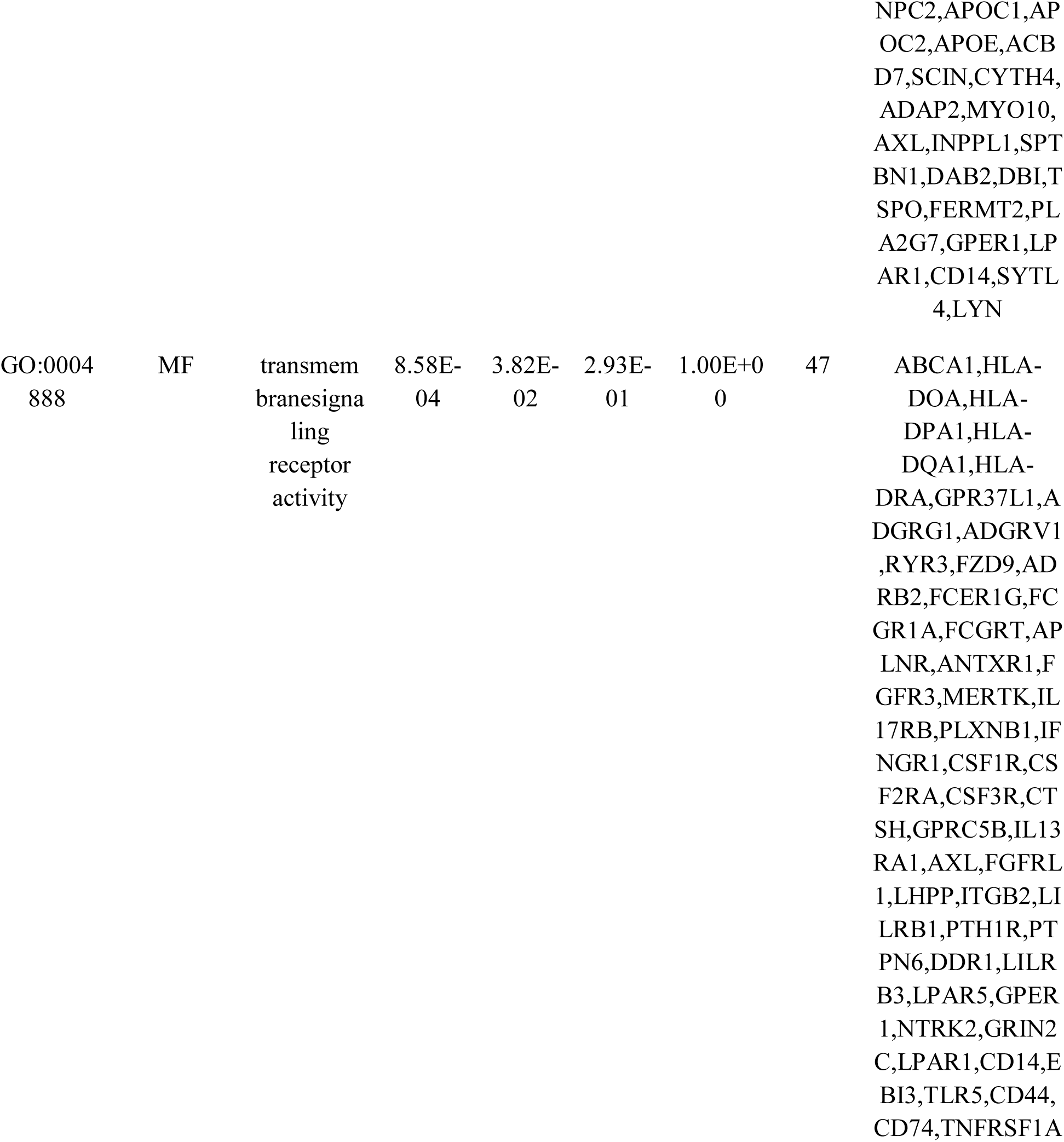
The enriched GO terms of the down regulated differentially expressed genes

### Protein–protein interaction network (PPI) and modular analysis

The potential relationships among these DEGs (up and down regulated genes) at protein levels were predicted based on the IntAct Molecular Interaction database. As shown in Fig. 5, the establishment of the PPI network for up regulated genes identified 3664 nodes and 6054 edges. The top hub genes (GABARAPL1, YWHAZ, SSX2IP, YWHAH, UBE2N, CALM3, TUBB3, NME5, CAPRIN2, BEX5, SV2B and KCNIP4) were identified based on their connectivity degree, betweenness centrality, stress centrality, closeness centrality and clustering coefficient and are listed in Table 6. Scatter plot along with statistical results for node degree, betweenness centrality, stress centrality, closeness centrality and clustering coefficient are shown in Fig. 6A - 6E. Pathways and GO enrichment analysis revealed that these hub genes were markedly enriched in GABAergic synapse, role of calcineurin-dependent NFAT signaling in lymphocytes, cell projection organization, chemical synaptic transmission, enzyme binding, oxytocin signaling pathway, Huntington disease, cell morphogenesis, signaling receptor binding, cell-cell signaling and ion transport. As shown in Fig. 7, the establishment of the PPI network for down regulated genes identified 3095 nodes and 4747 edges. The top hub genes (HSPB1, HDAC1, CDKN1A, TNFRSF1A, FKBP5, BCL6, MYL12A, SPTBN1, SFMBT2, NUPR1, TRIM47, RAPGEF3 and MS4A7) were identified based on their connectivity degree, betweenness centrality, stress centrality, closeness centrality and clustering coefficient and are listed in Table 6. Scatter plot along with statistical results for node degree, betweenness centrality, stress centrality, closeness centrality and clustering coefficient are shown in Fig. 8A - 8E. Pathways and GO enrichment analysis revealed that these hub genes were markedly enriched in cell activation, validated targets of C-MYC transcriptional repression, HTLV-I infection, defense response, direct p53 effectors, regulation of actin cytoskeleton, neutrophil degranulation, regulation of cell proliferation and hemostasis,

**Fig. 5.**
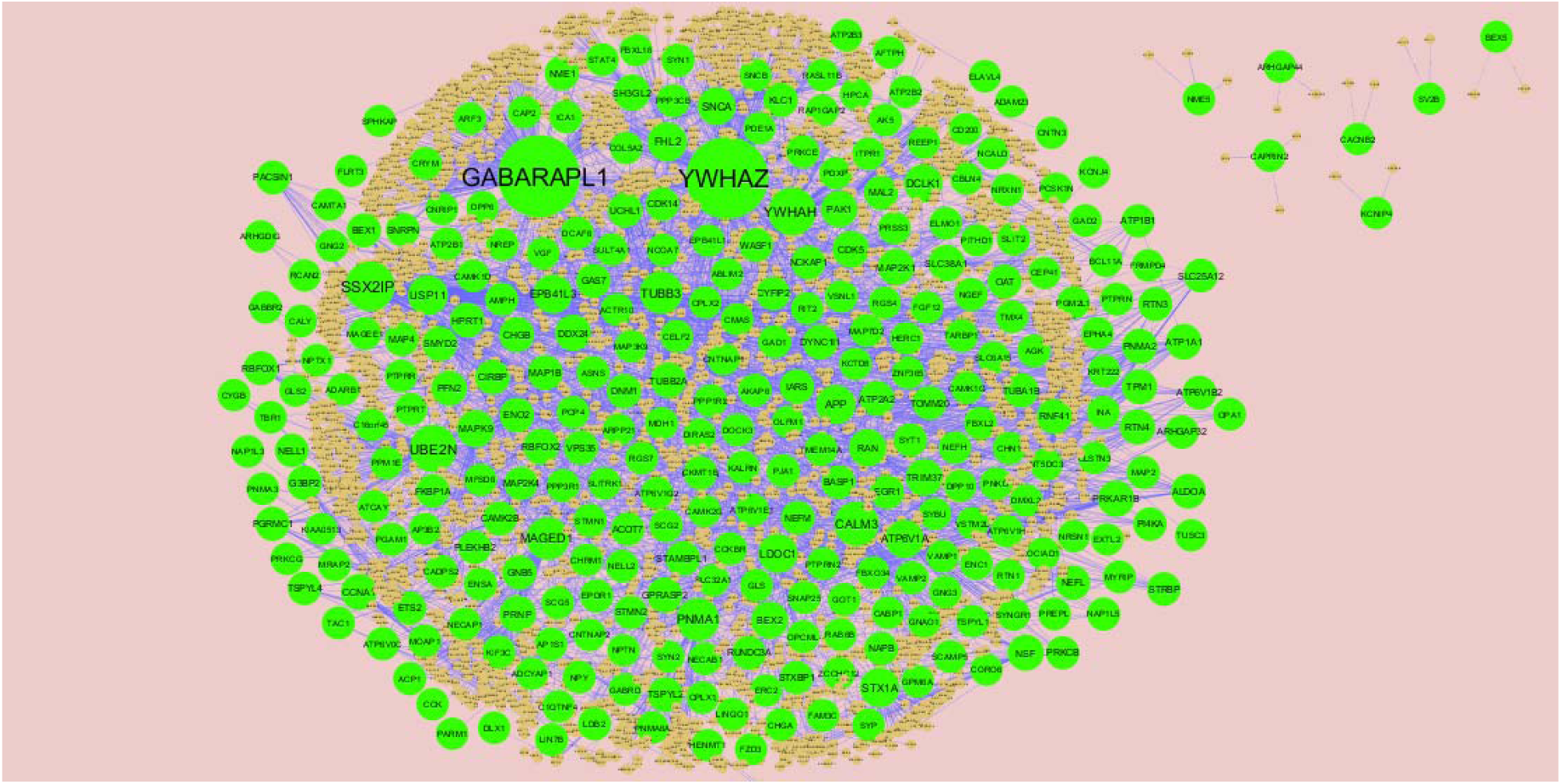
Protein–protein interaction network of differentially expressed genes (DEGs). Green nodes denotes up regulated genes.

**Fig. 6.**
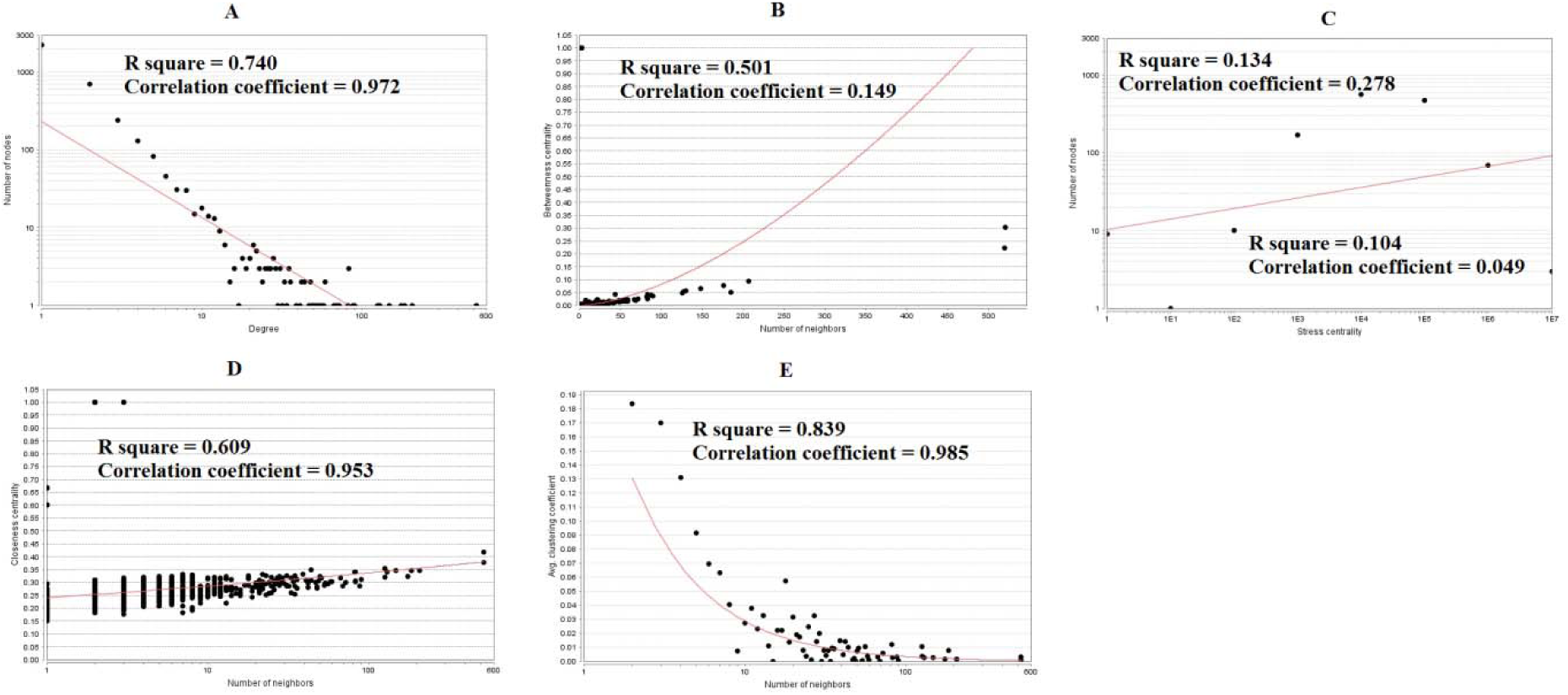
Scatter plot for up regulated genes. (A- Node degree; B- Betweenness centrality; C- Stress centrality; D- Closeness centrality; E- Clustering coefficient)

**Fig. 7.**
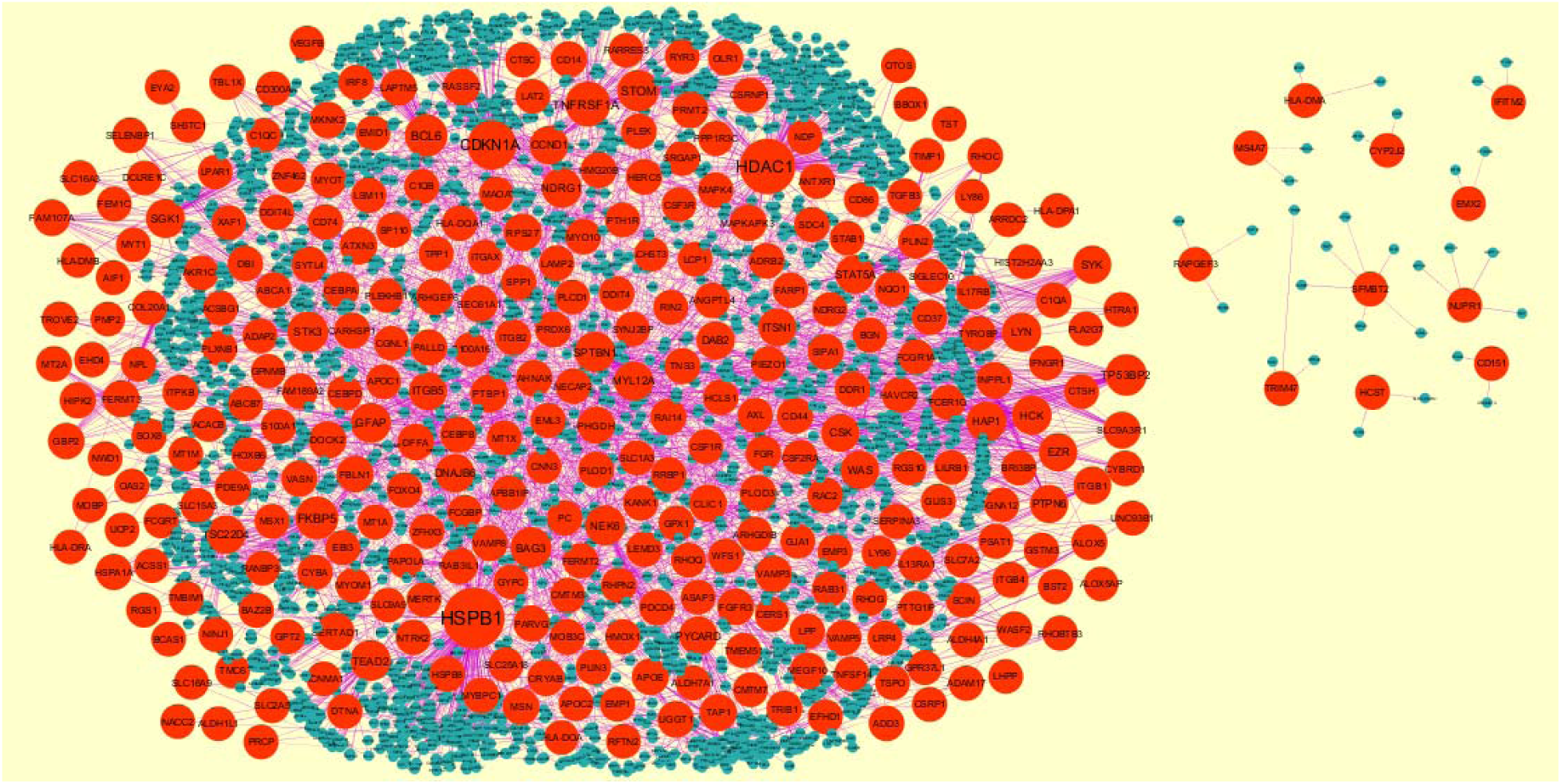
Protein–protein interaction network of differentially expressed genes (DEGs). Red nodes denotes down regulated genes.

**Fig. 8.**
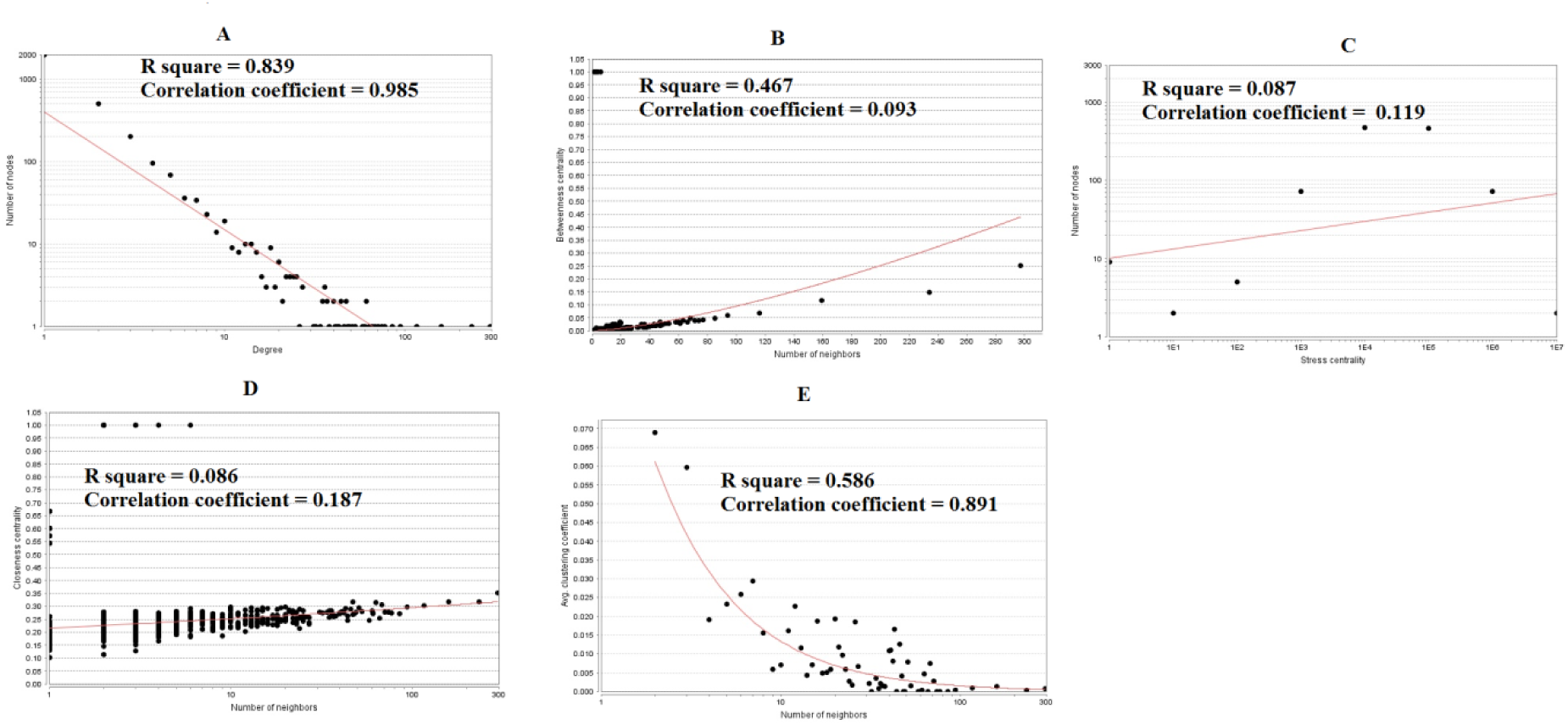
Scatter plot for down regulated genes. (A - Node degree; B - Betweenness centrality; C- Stress centrality; D- Closeness centrality; E - Clustering coefficient)

**Table 6.**
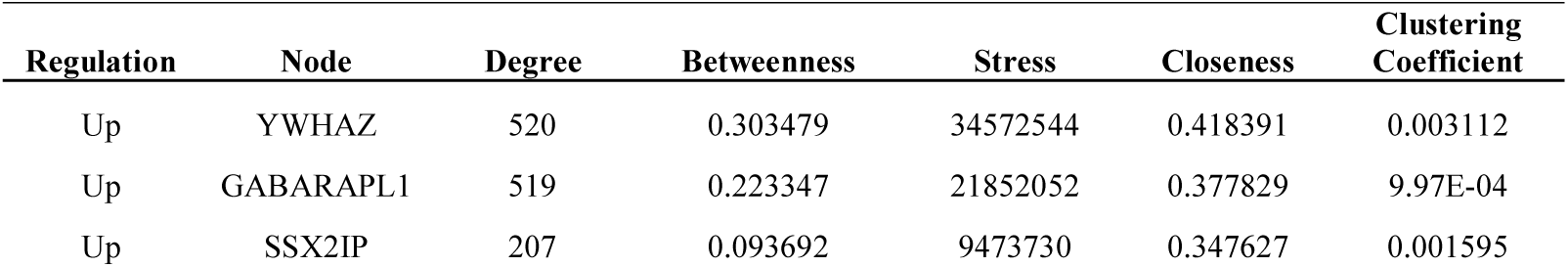
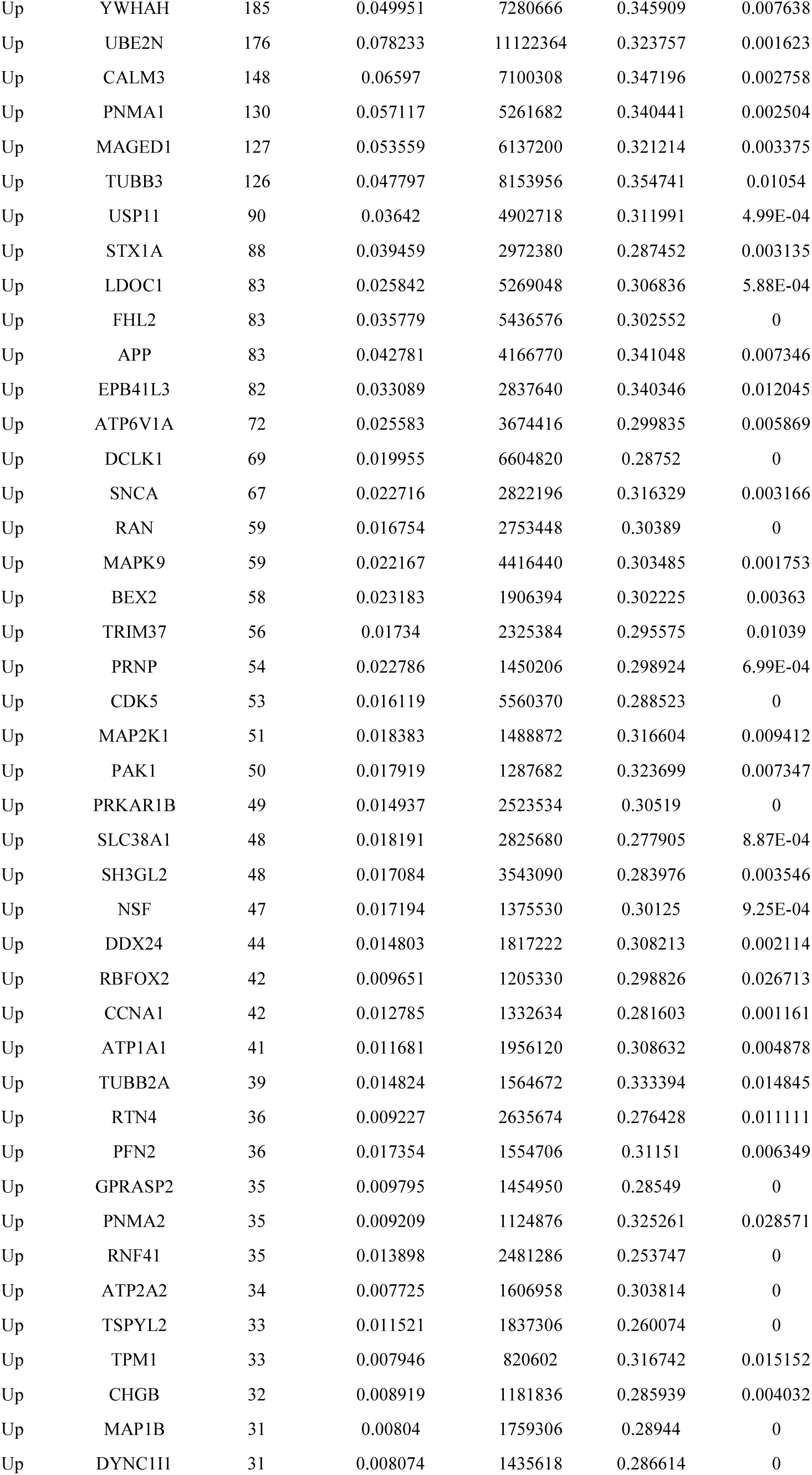
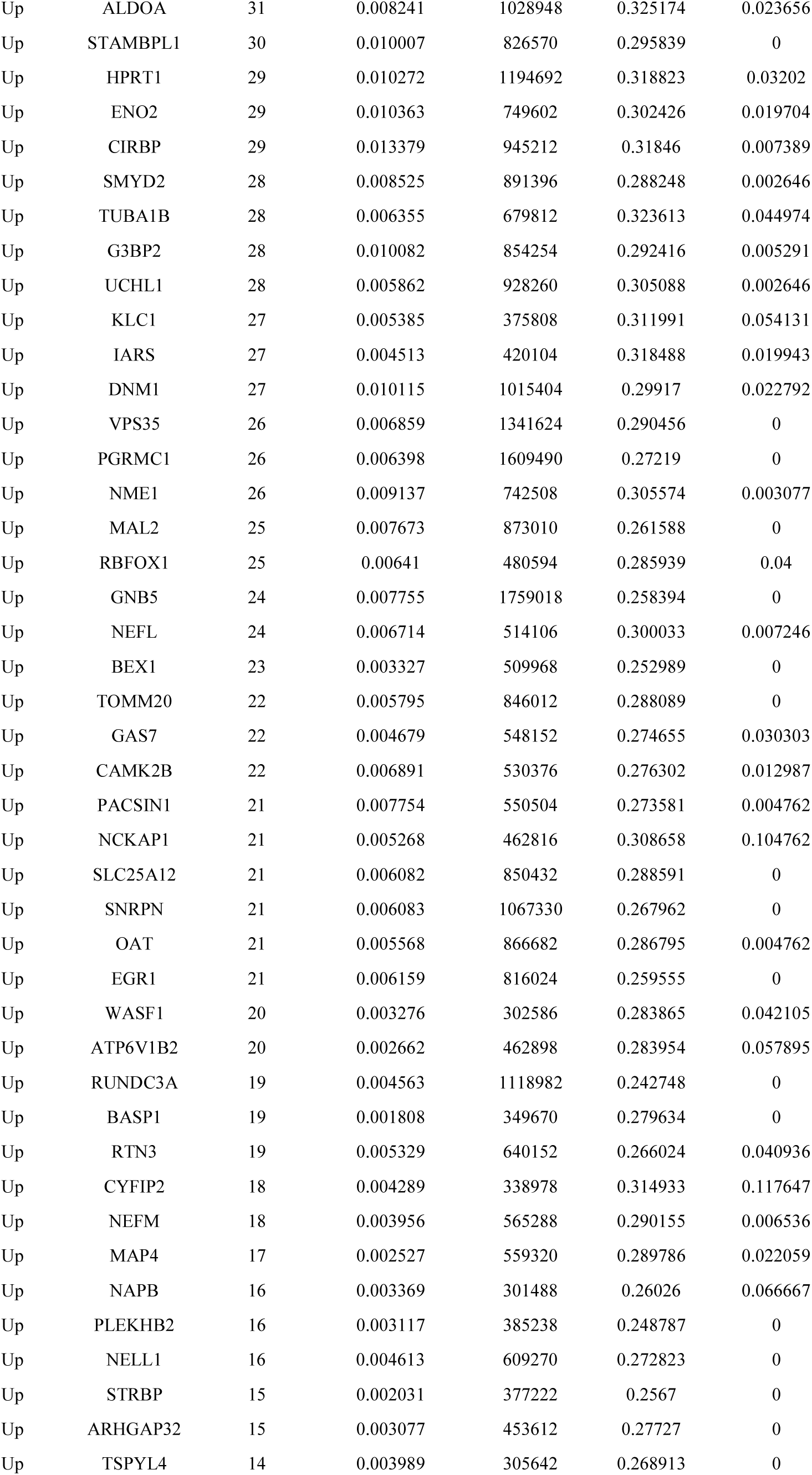
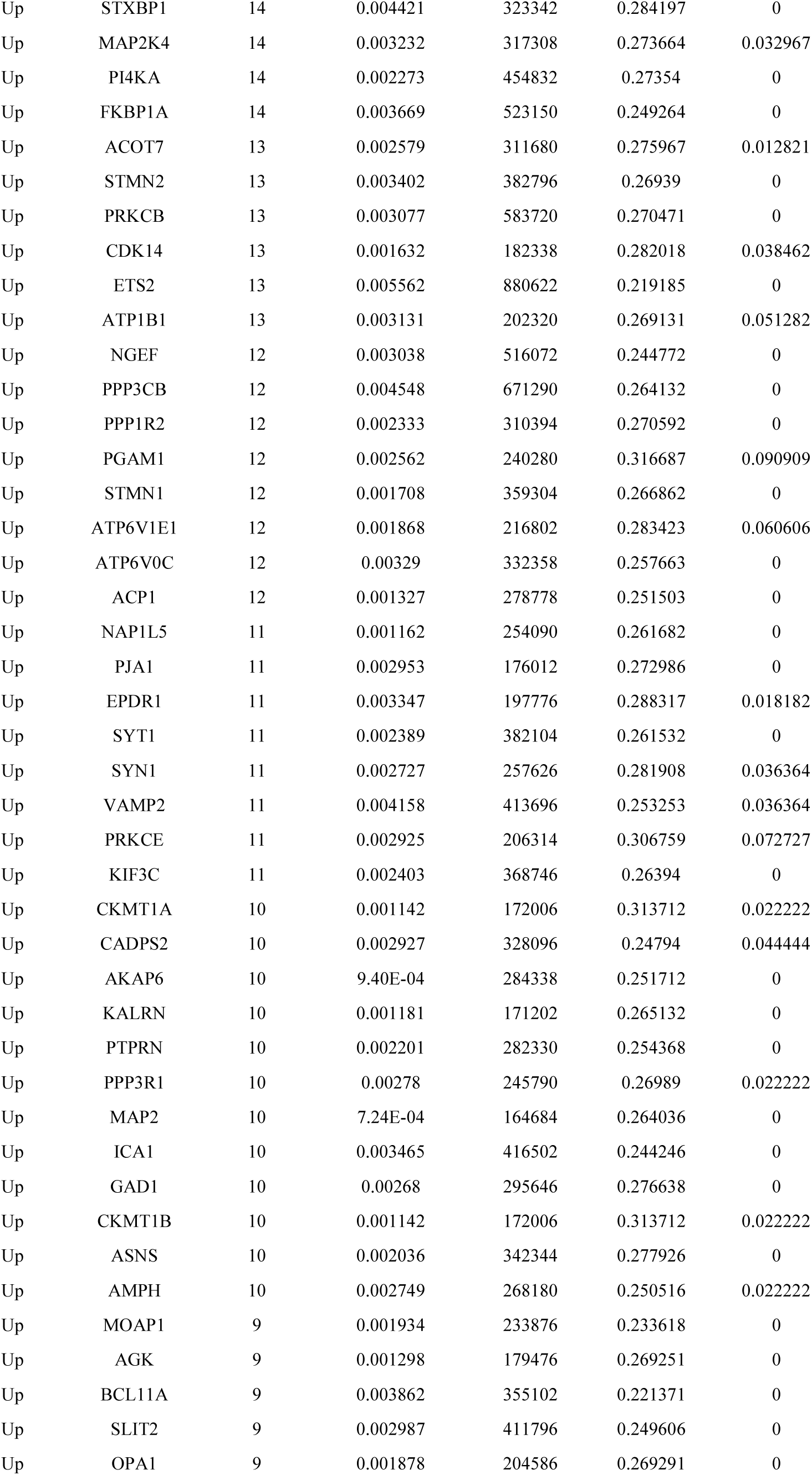
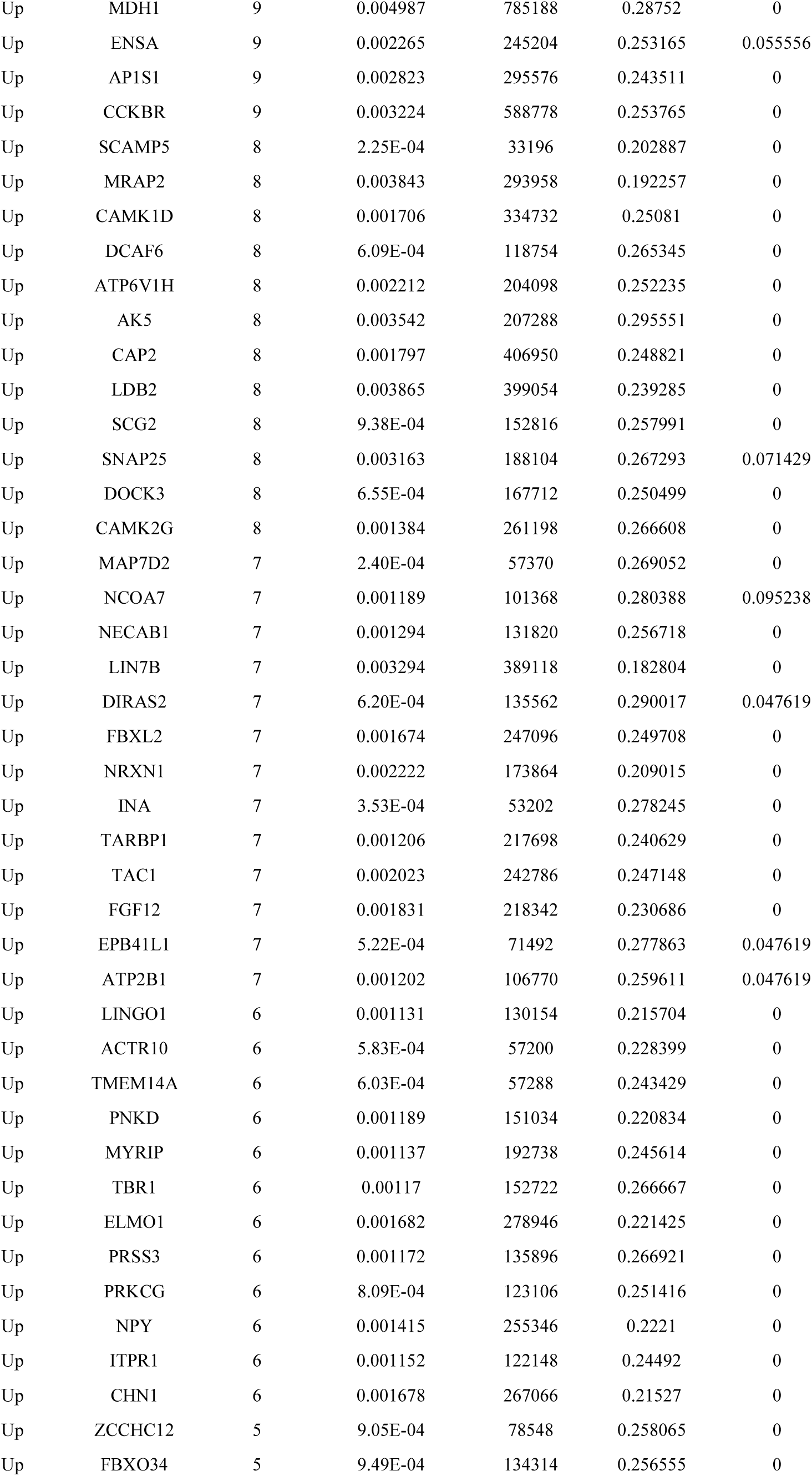
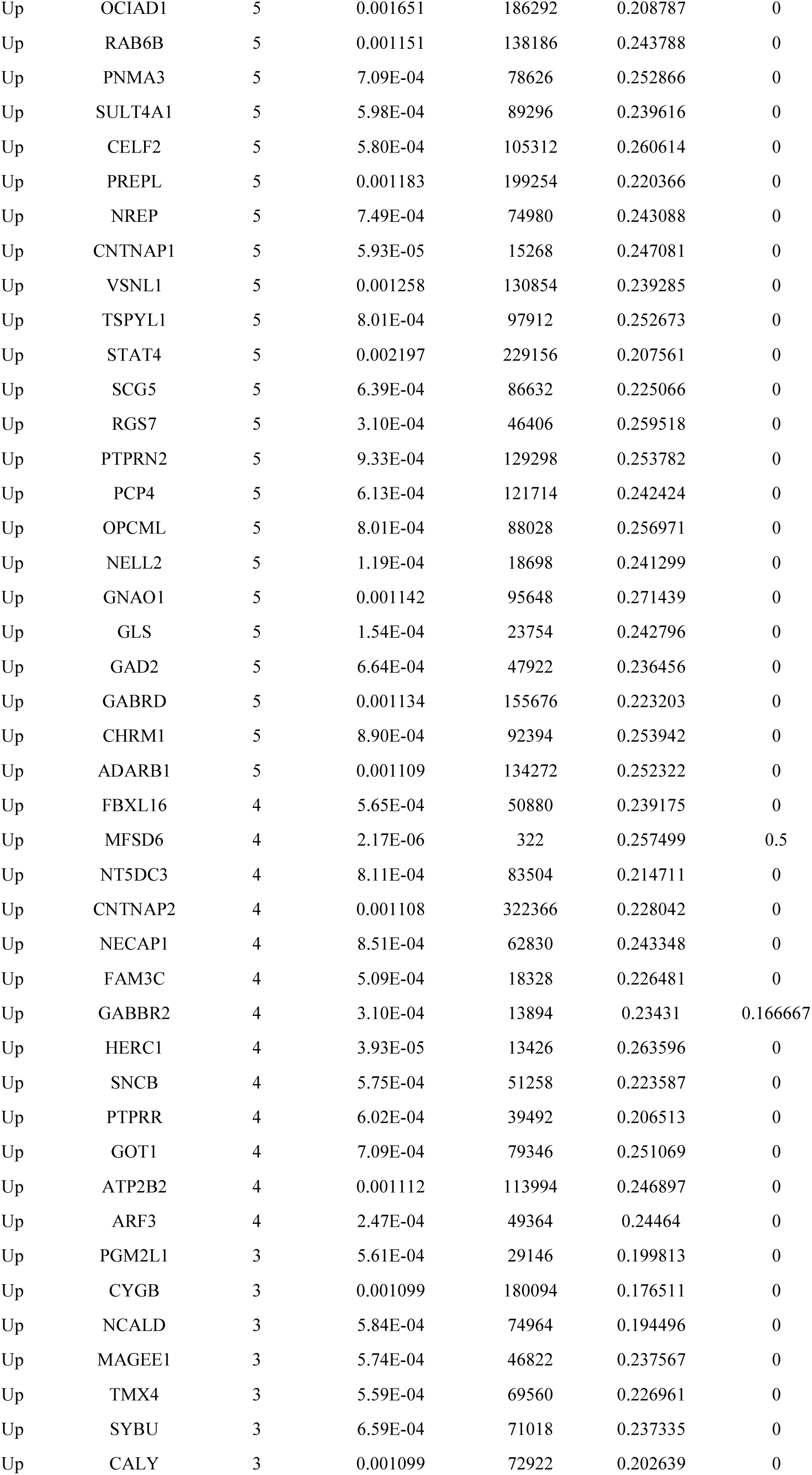
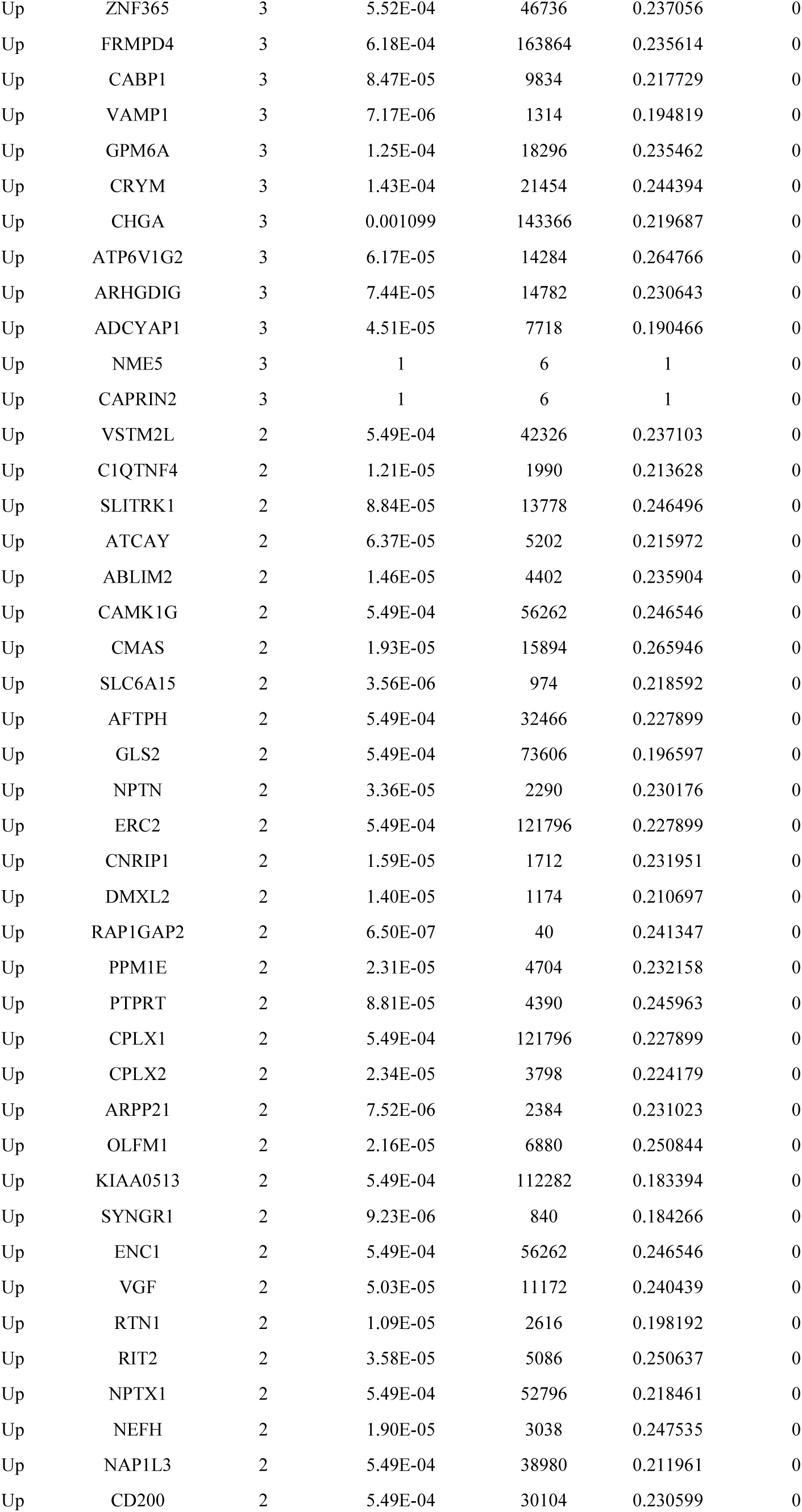
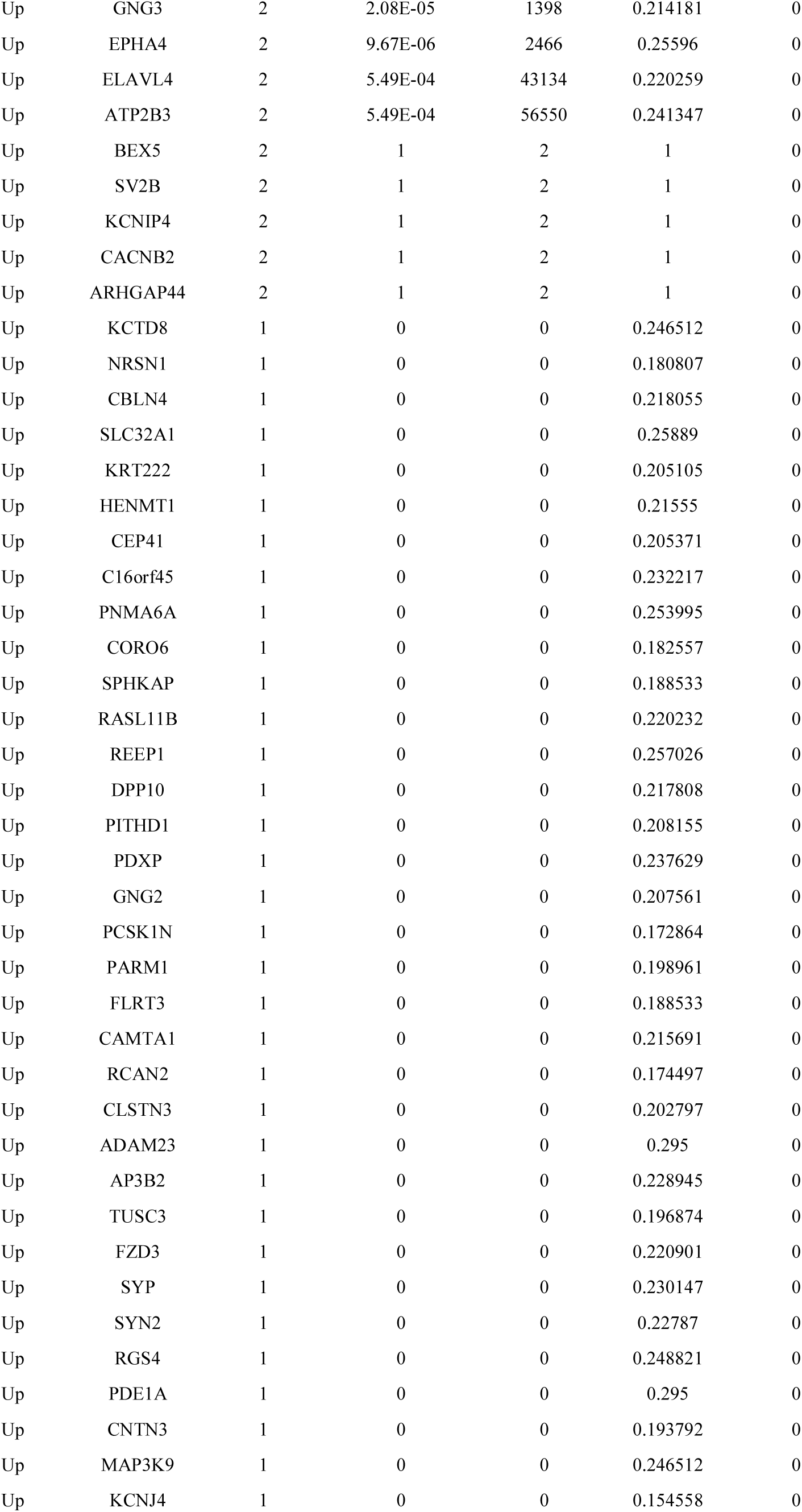
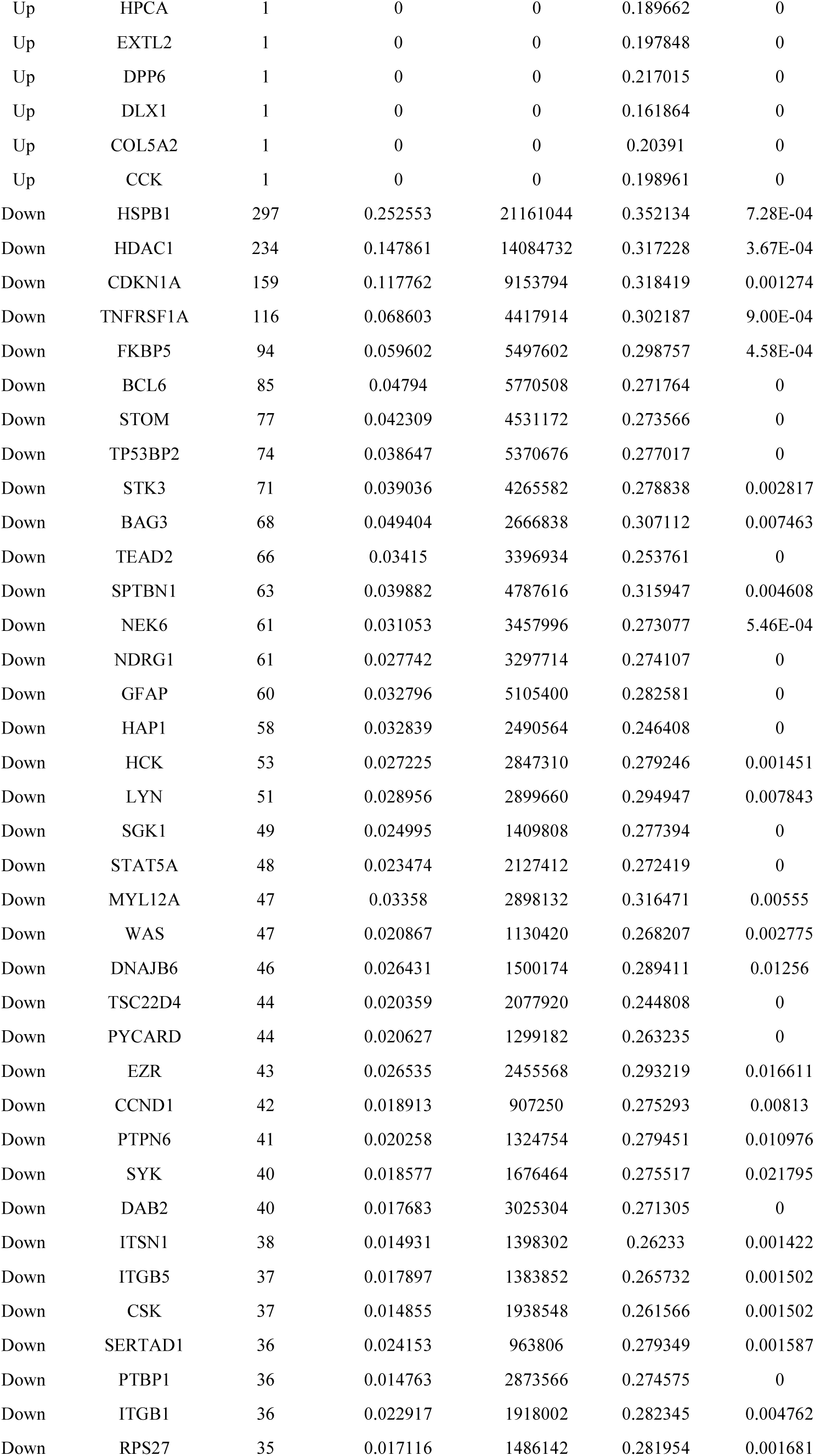
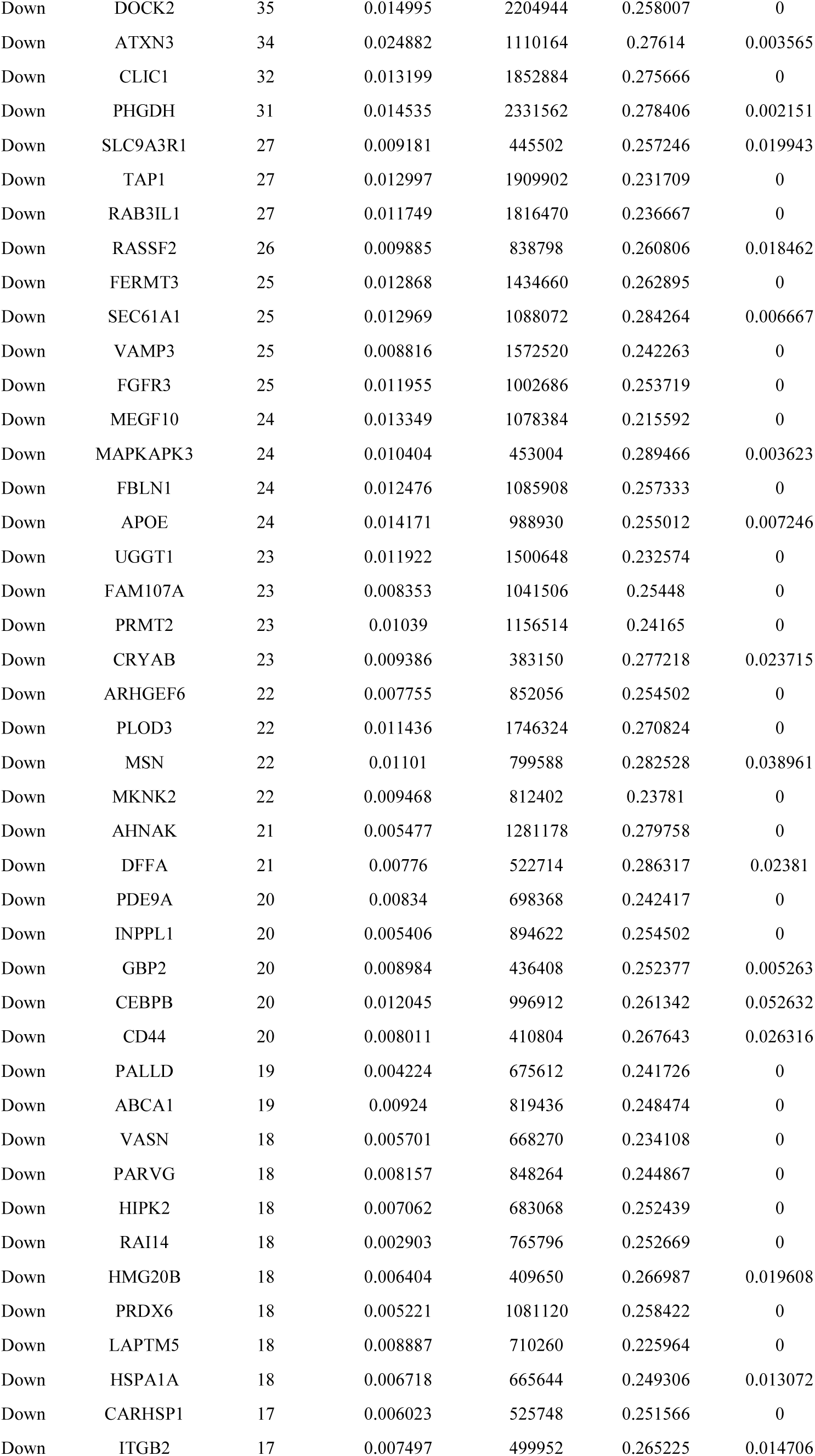
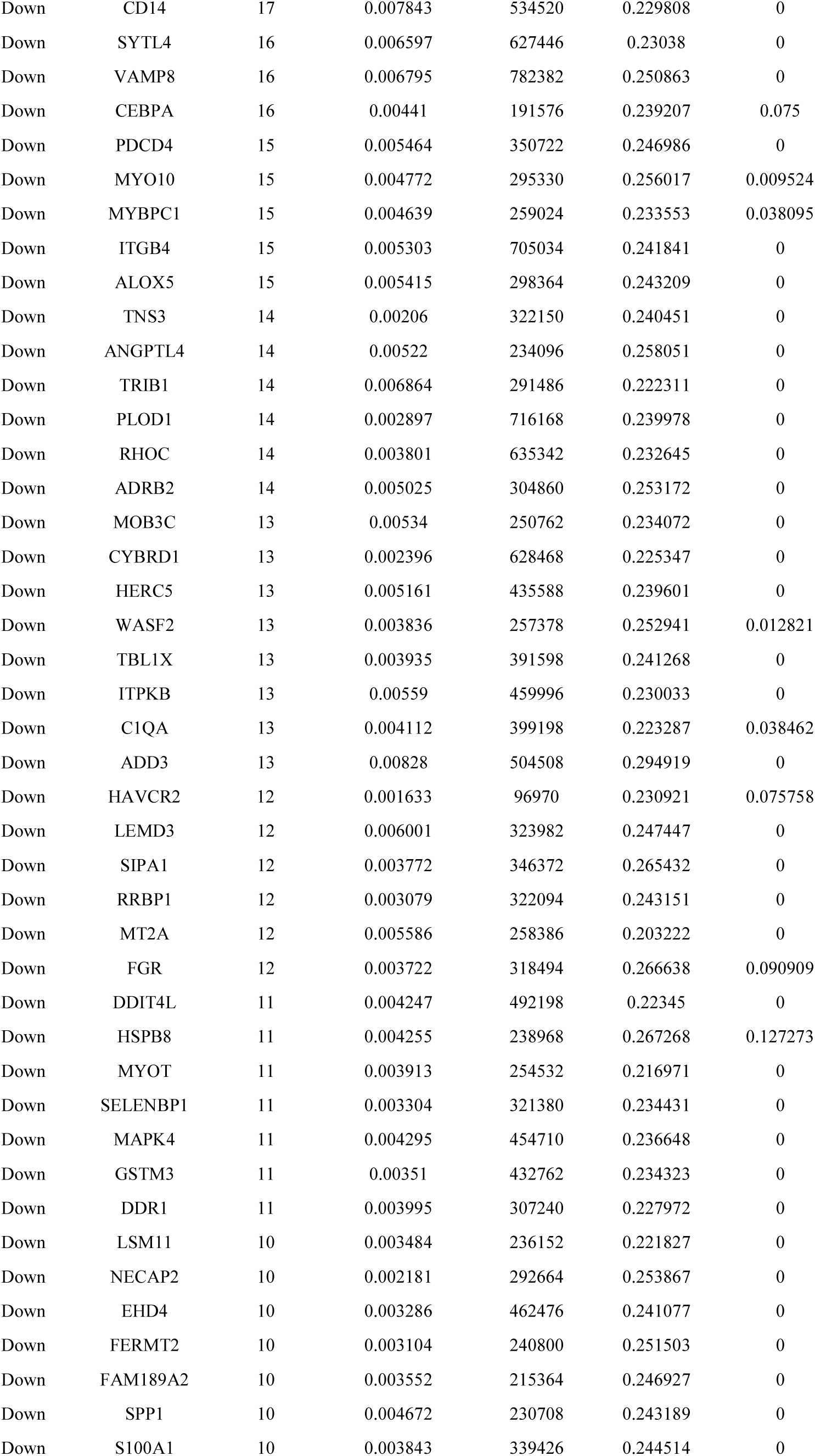
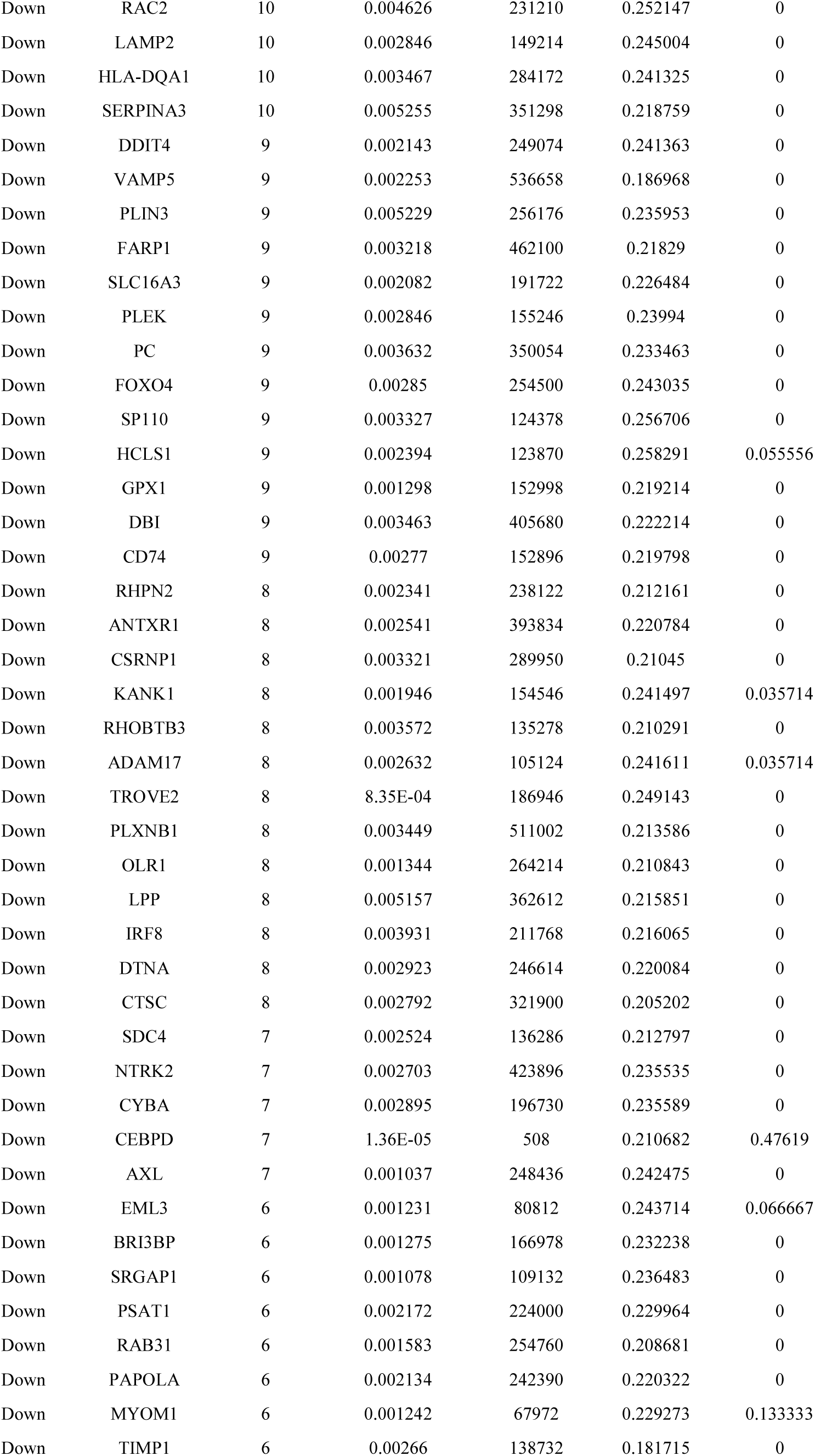
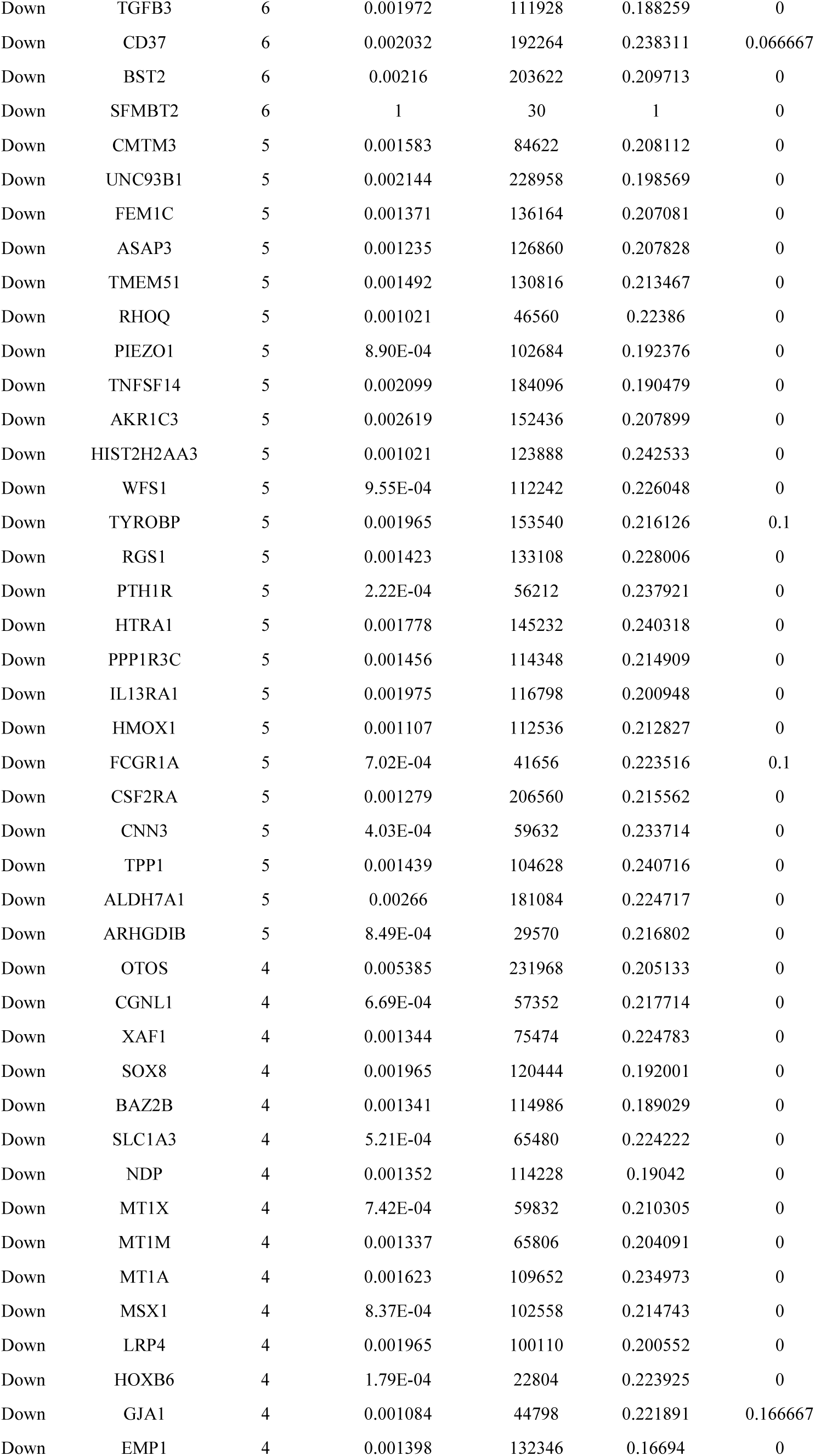
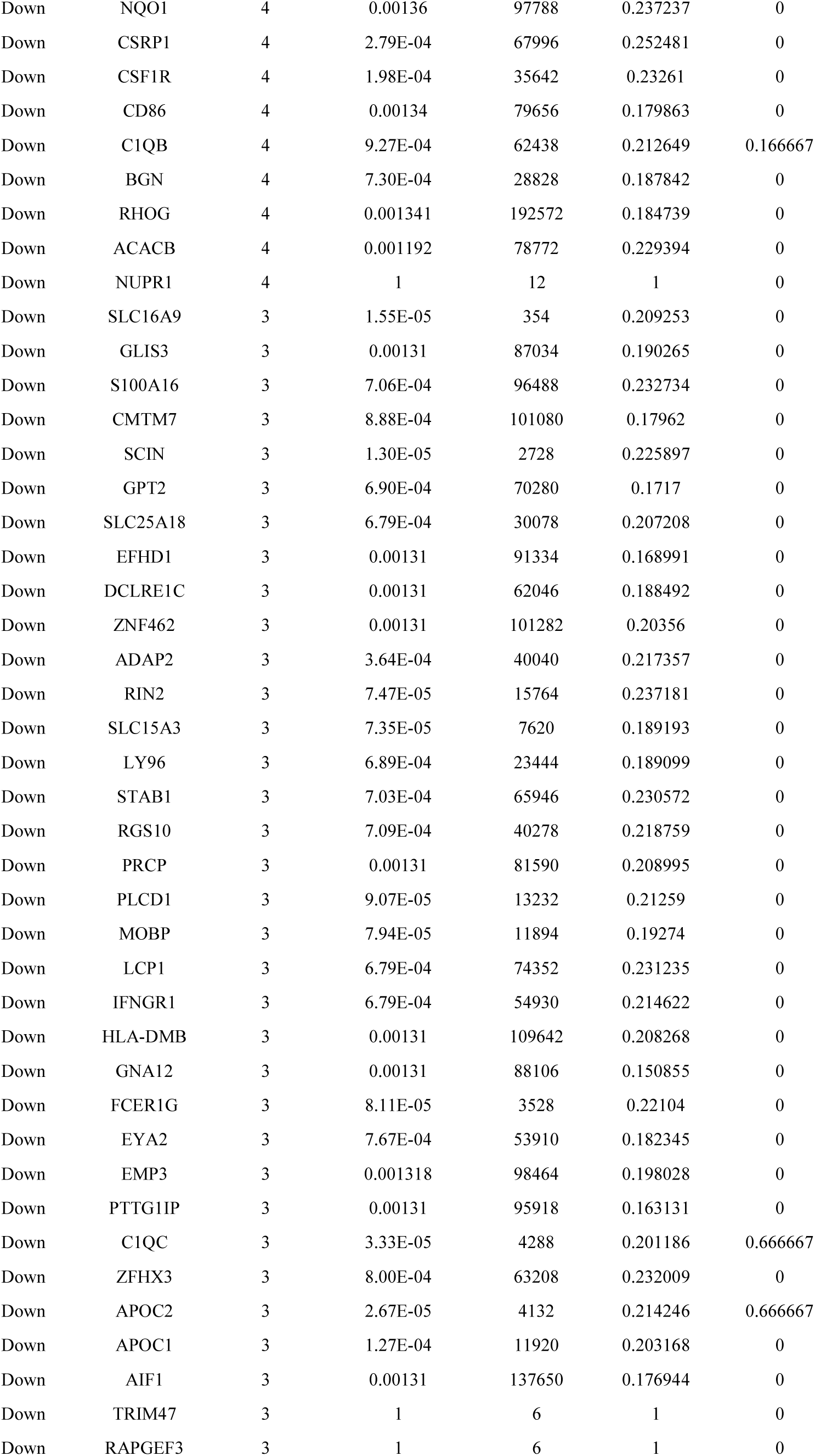
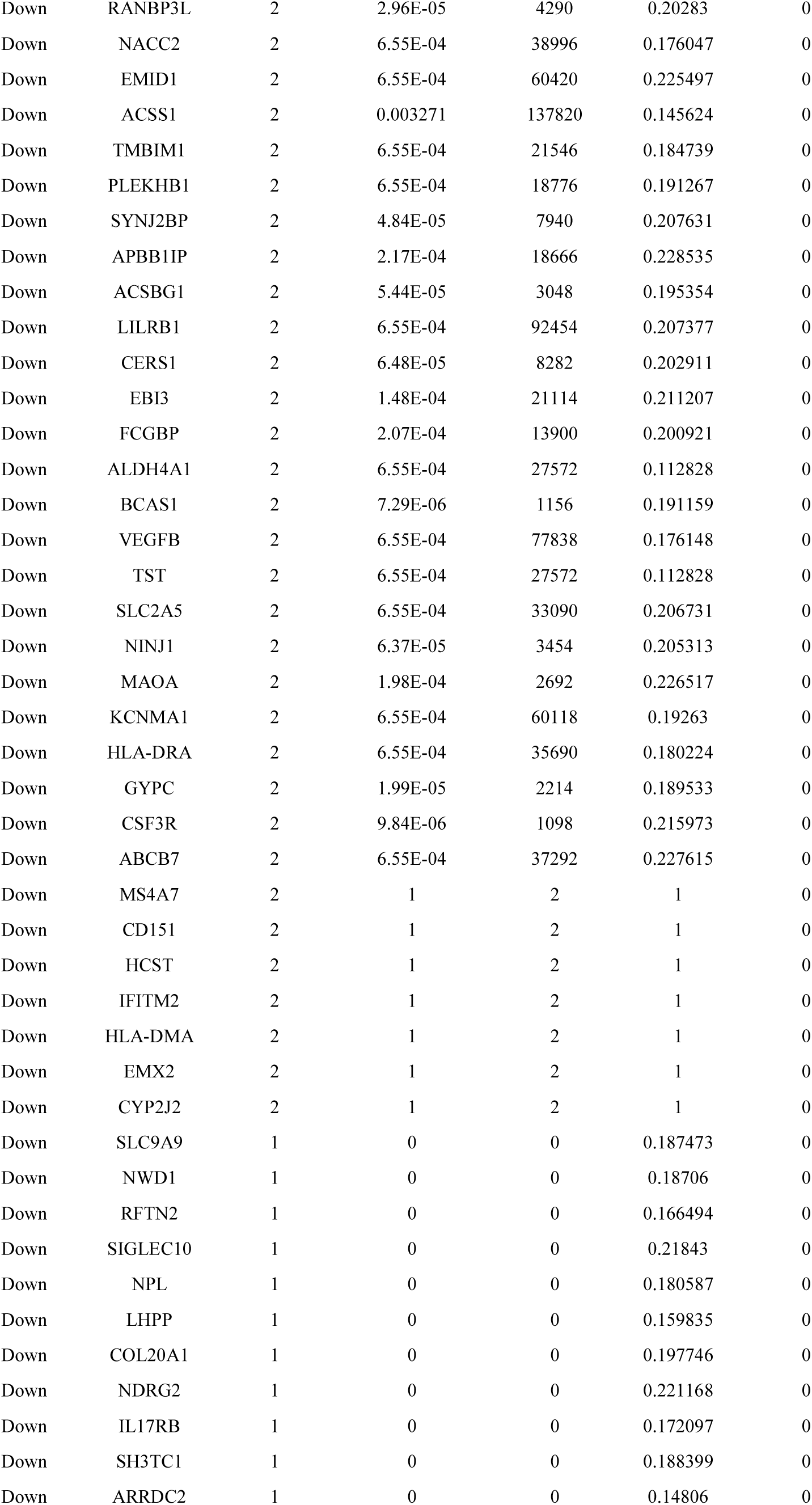
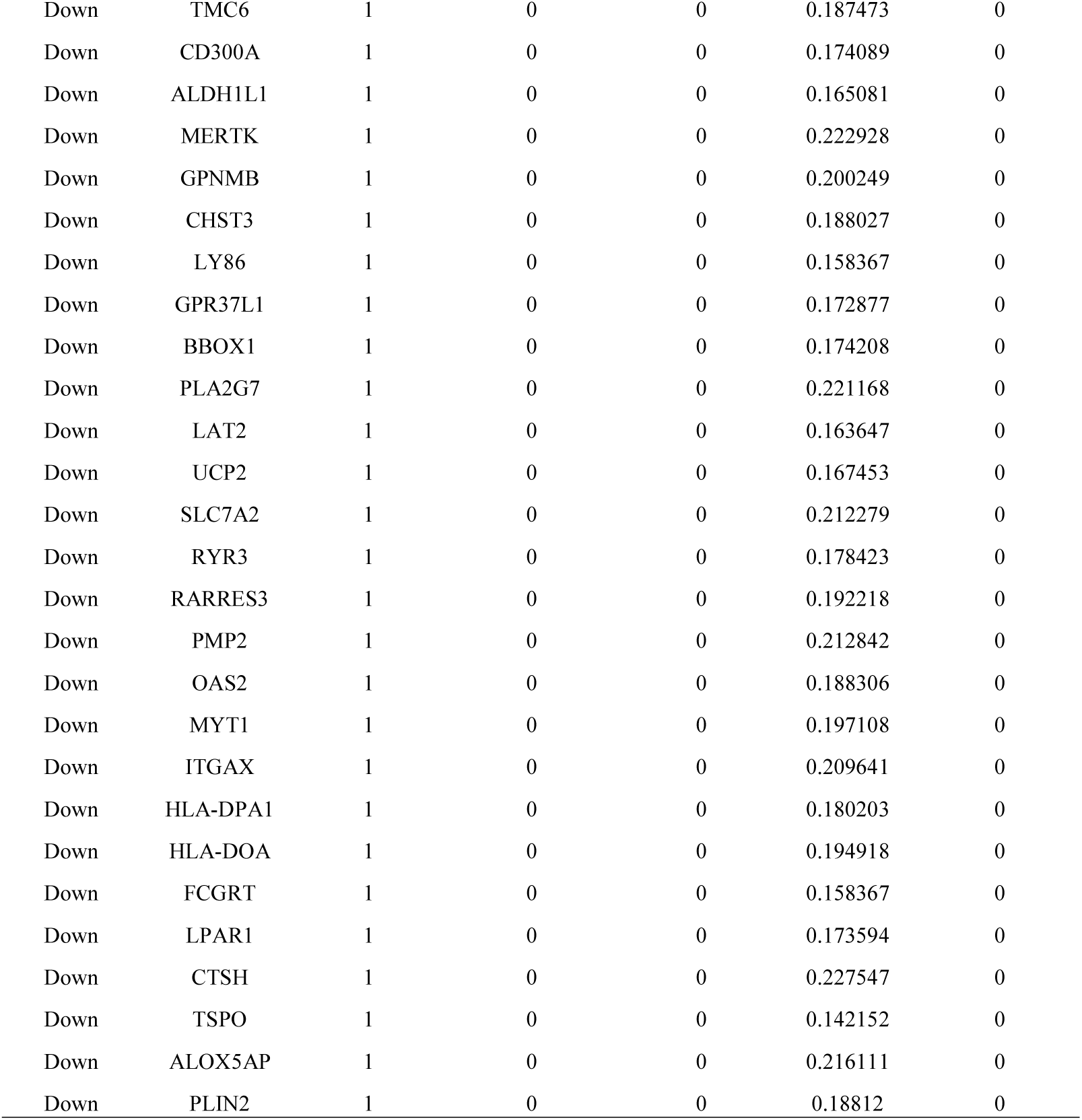
Topology table for up and down regulated genes

To explore the significance of the molecules of the sCJD related PPI network, the module analysis was performed by PEWCC1. The four most significant modules of the PPI network for up regulated genes were shown in Fig 9, in which GABARAPL1, YWHAZ, YWHAH, EPB41L3, KLC1, CYFIP2, NCKAP1, MAGED1, RBFOX1, RBFOX2, SSX2IP, TUBB2A, USP11, PFN2, HPRT1, NCOA and ENO2 were the top hub genes. Pathway and GO enrichment analyses of these hub genes associated in these module were performed using ToppGene. Results showed that the genes in these significant modules were predominantly enriched for terms associated with GABAergic synapse, neurotrophic factor-mediated Trk receptor signaling, Huntington disease, superpathway of purine nucleotide salvage, cell-cell signaling, neuron projection development, cell projection organization and chemical synaptic transmission. The four most significant modules of the PPI network for down regulated genes were shown in Fig 10, in which PTPN6, CD37, EZR, MSN, SLC9A3R1, CD44, CEBPA, CEBPB, CEBPD, HSPB1, LYN, HSPB8, BAG3 and CRYAB were the top hub genes. Pathway and GO enrichment analyses of these hub genes associated in these module were performed using ToppGene. Results showed that the genes in these significant modules were predominantly enriched for terms associated with leishmaniasis, regulation of actin cytoskeleton, validated targets of C-MYC transcriptional repression, regulation of immune system process, biological adhesion, cell activation, cell adhesion and identical protein binding.

**Fig. 9.**
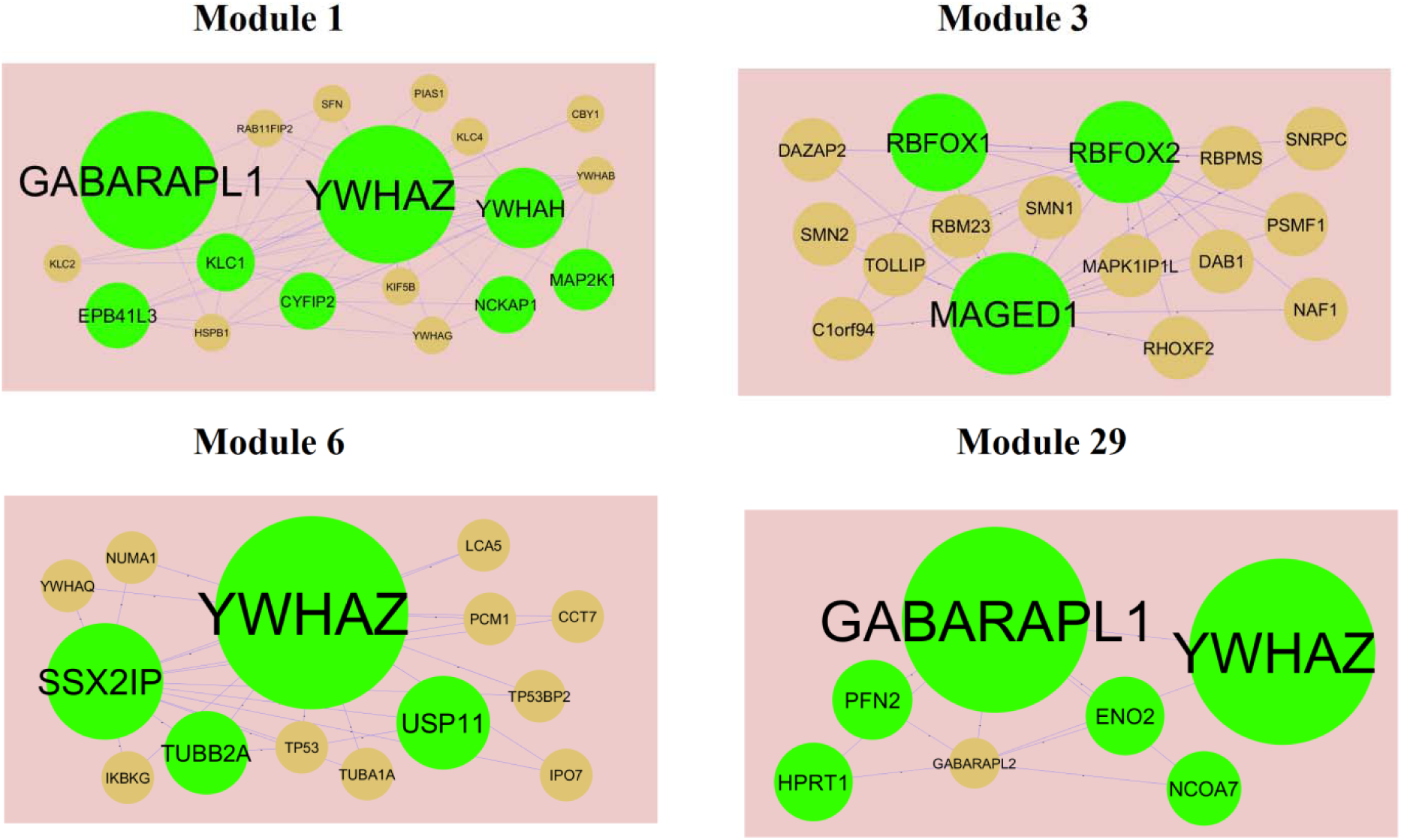
Modules in PPI network. The green nodes denote the up regulated genes

**Fig. 10.**
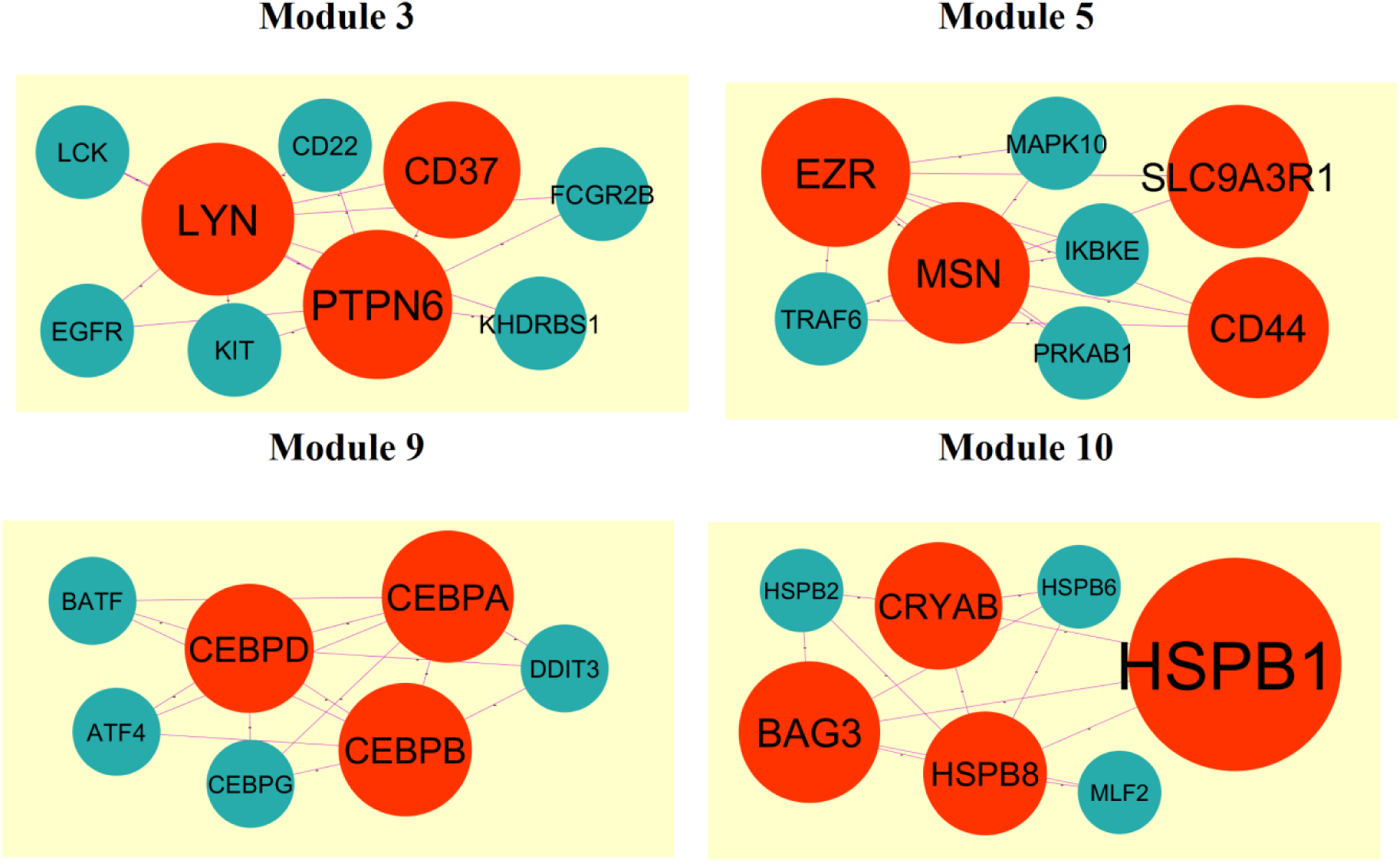
Modules in PPI network. The red nodes denote the down regulated genes.

### Construction of target gene - miRNA regulatory network

The target gene - miRNA regulatory network for up regulated genes is shown in Fig. 11. Top five up regulated target genes such as TUBB2A interacts with 183 miRNAs (ex, hsa-mir-5681a), YWHAZ interacts with 156 miRNAs (ex, hsa-mir-3189-5p), MAP3K9 interacts with 121 miRNAs (ex, hsa-mir-6791-3p), PGM2L1 interacts with 117 miRNAs (ex, hsa-mir-6764-3p) and ALDOA interacts with 112 miRNAs (ex, hsa-mir-4476) are listed in Table 7. Pathways and GO enrichment analysis revealed that these target genes were markedly enriched in Huntington disease, role of calcineurin-dependent NFAT signaling in lymphocytes, purine nucleotide binding and identical protein binding. Similarly, target gene - miRNA regulatory network for down regulated genes is shown in Fig. 12. Top five down regulated target genes such as CCND1 interacts with 197 miRNAs (ex, hsa-mir-3973), MKNK2 interacts with 153 miRNAs (ex, hsa-mir-6515-5p), CDKN1A interacts with 131 miRNAs (ex, hsa-mir-6886-3p), SYNJ2BP interacts with 120 miRNAs (ex, hsa-mir-4659a-3p) and BRI3BP interacts with 119 miRNAs (ex, hsa-mir-3689b-3p) are listed in Table 7. Pathways and GO enrichment analysis revealed that these target genes were markedly enriched in HTLV-I infection, regulation of immune system process and locomotion.

**Fig. 11.**
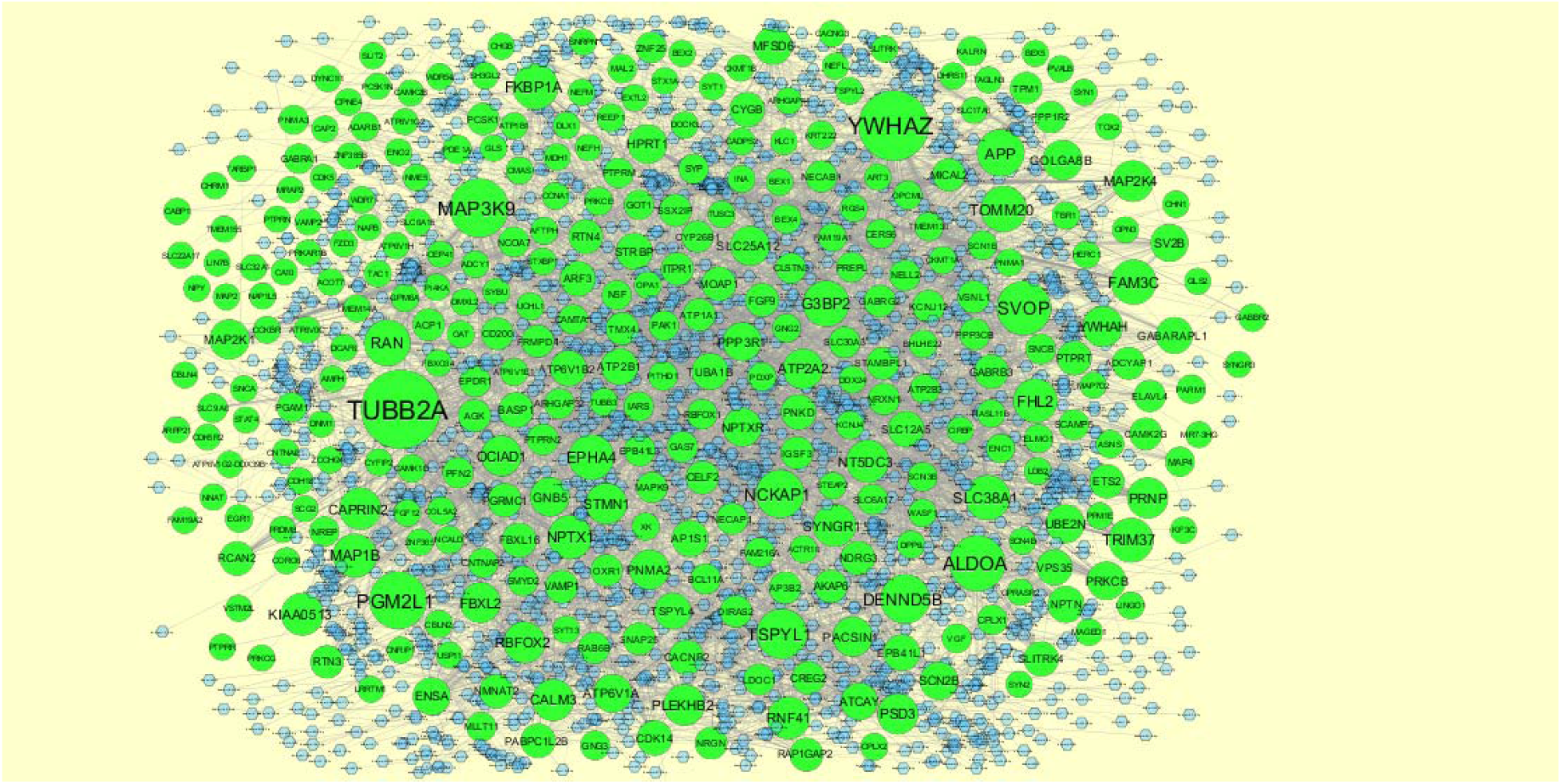
The network of up regulated DEGs and their related miRNAs. The green circles nodes are the up regulated DEGs, and blue diamond nodes are the miRNAs

**Fig. 12.**
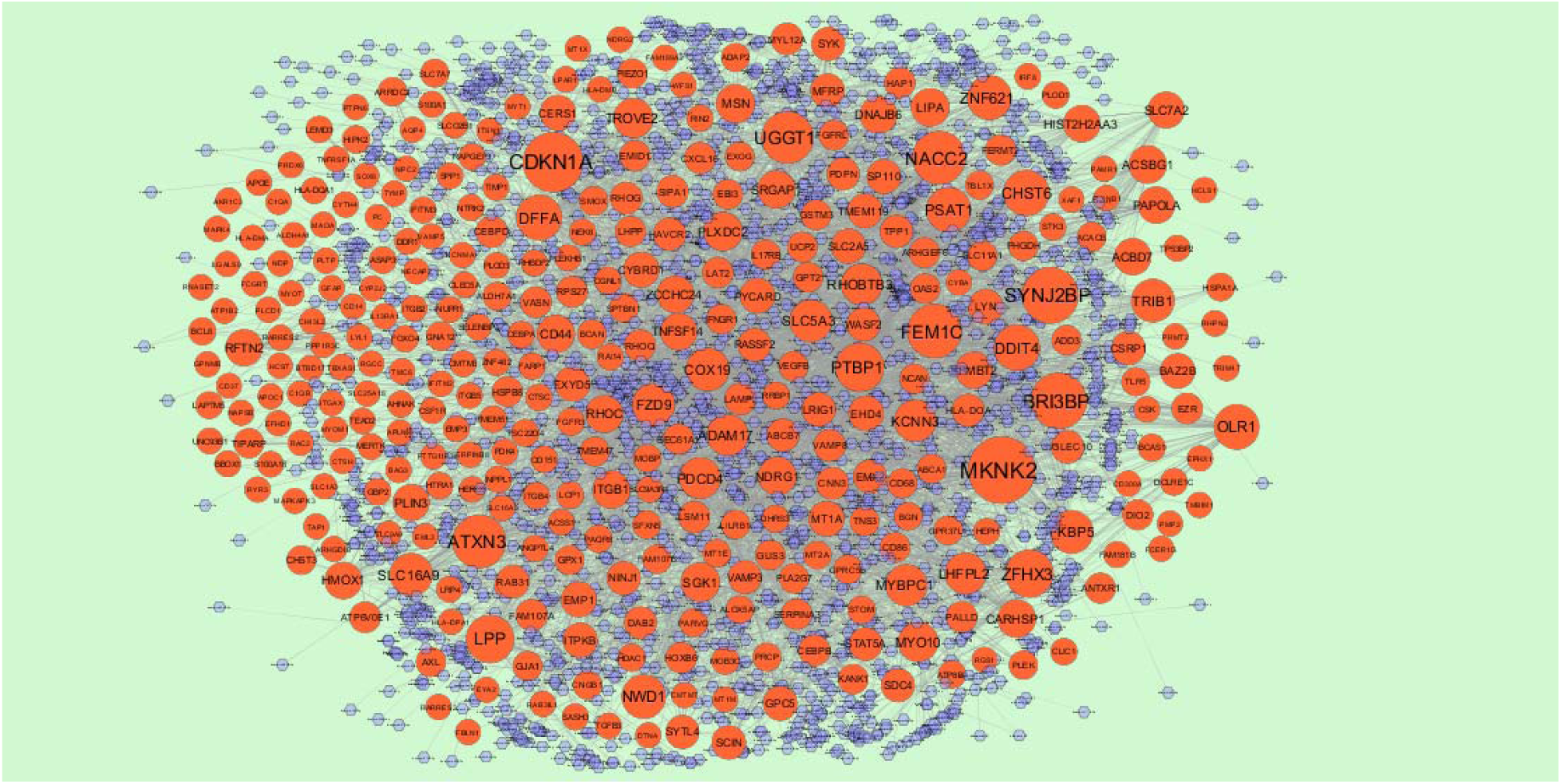
The network of down regulated DEGs and their related miRNAs. The red circles nodes are the down-regulated DEGs, and lavender diamond nodes are the miRNAs

**Table 7.**
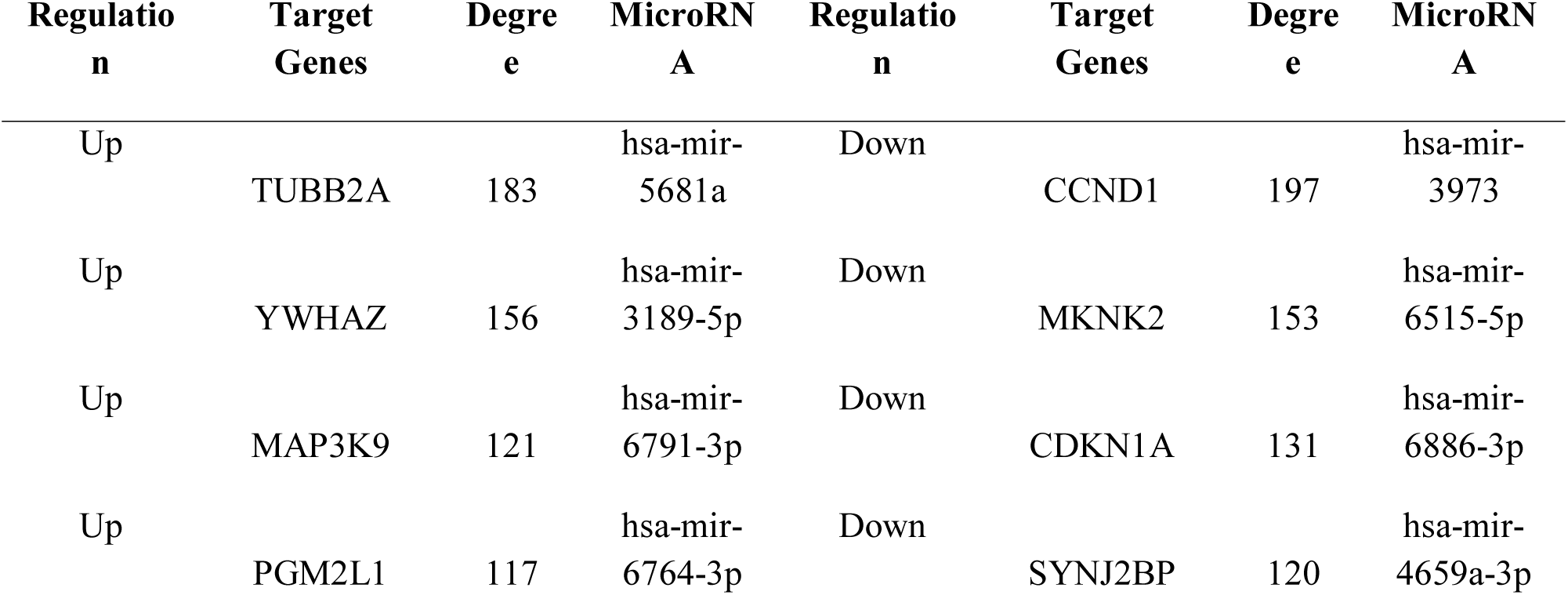
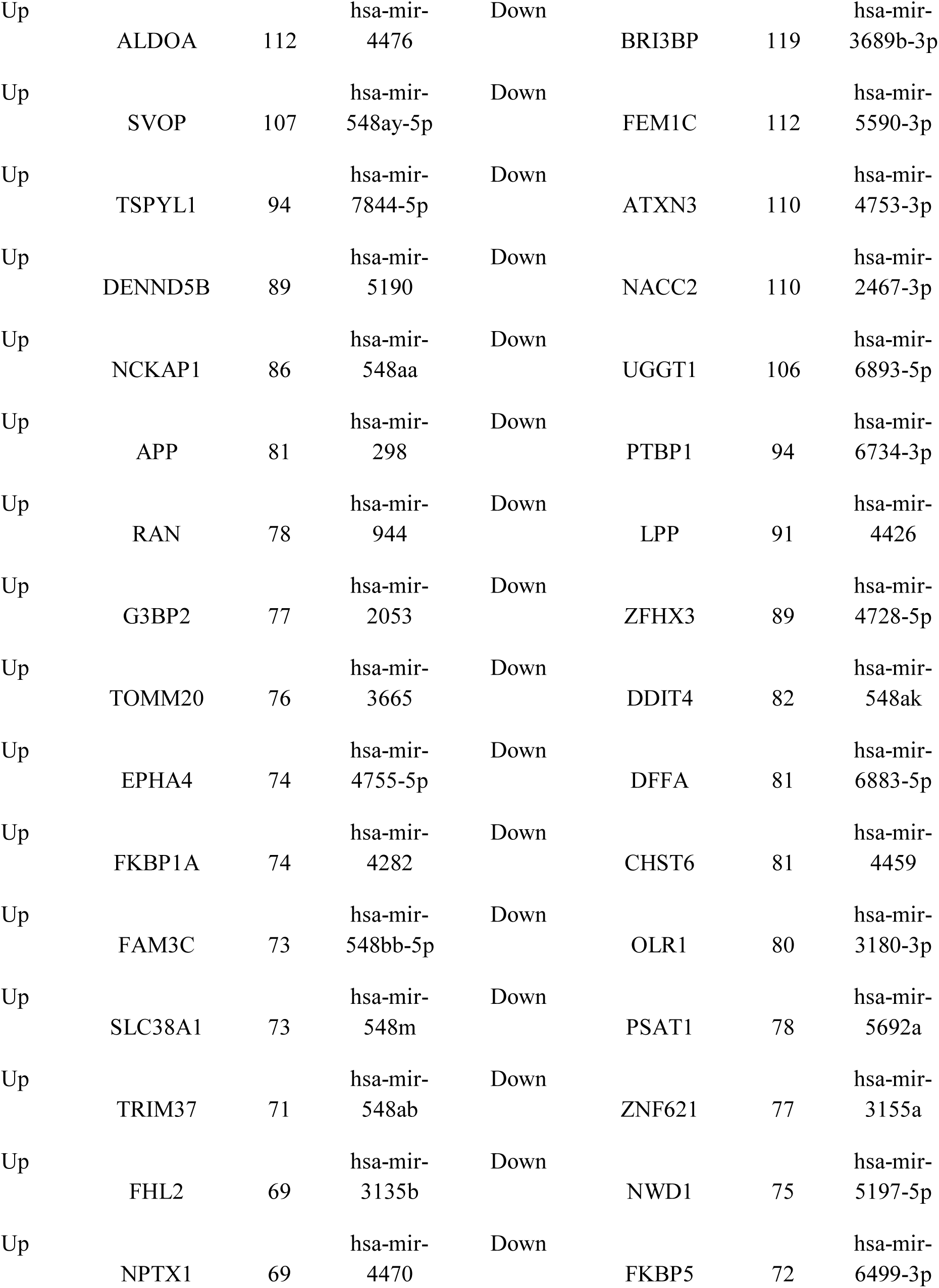
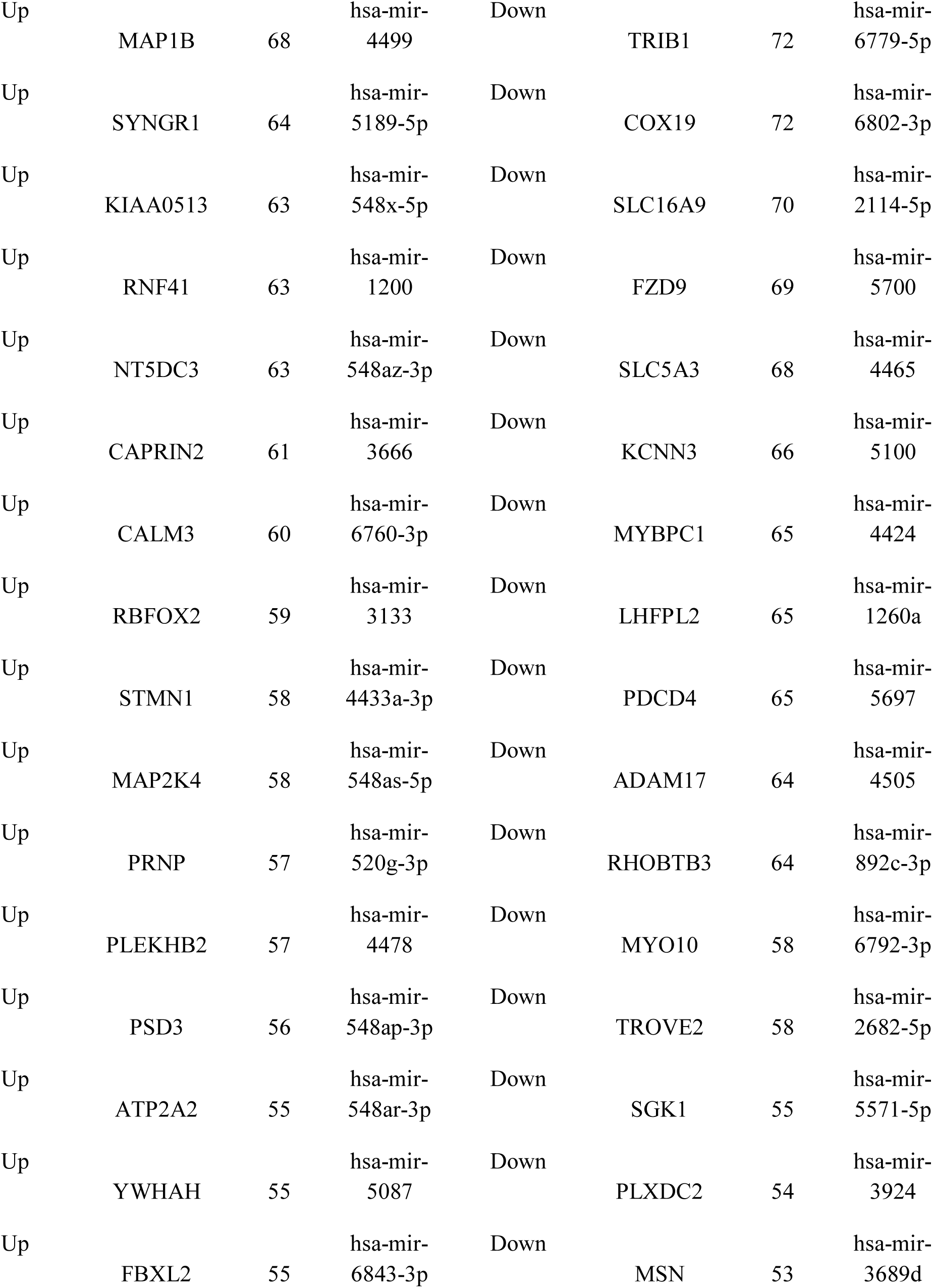
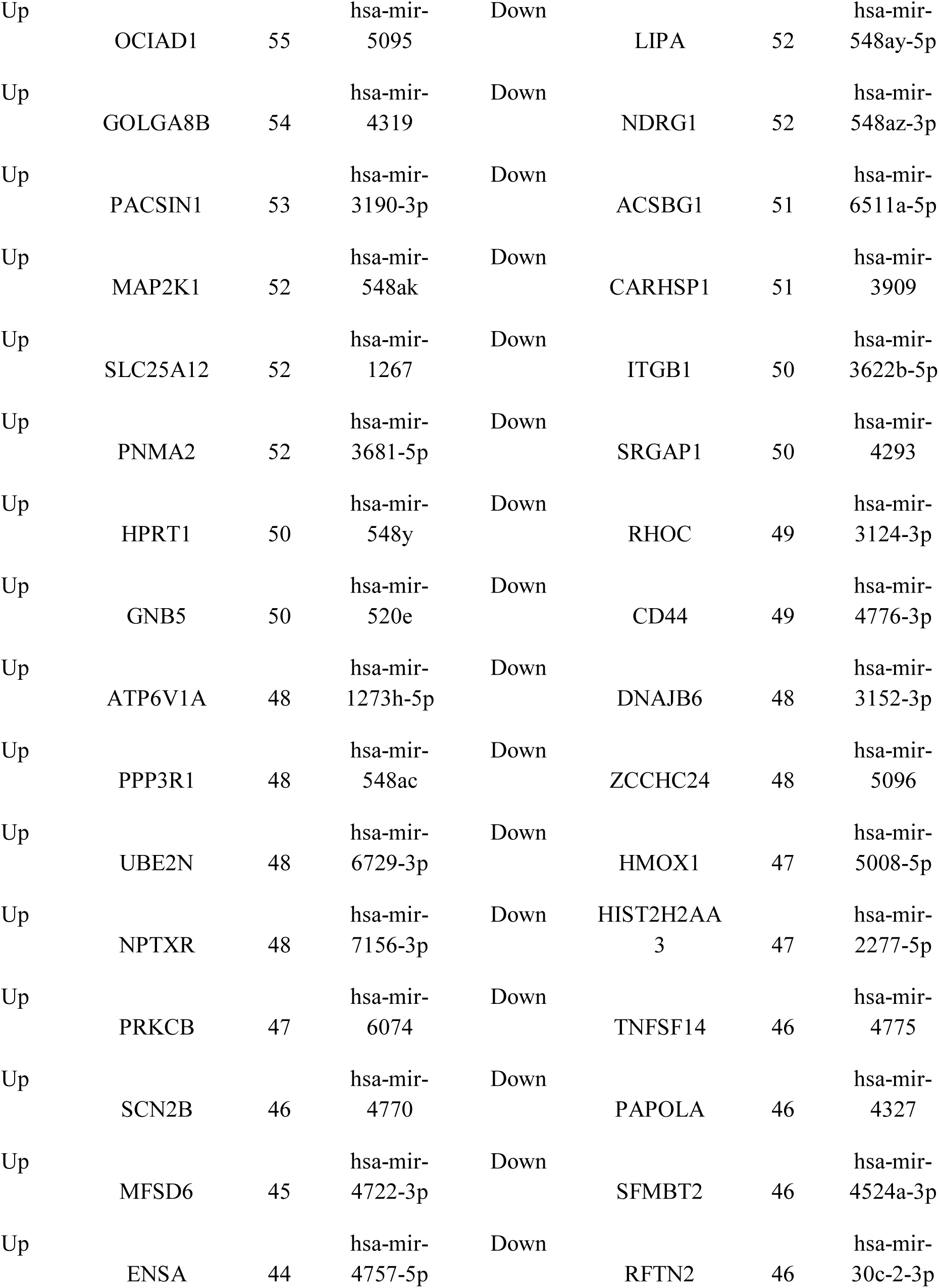
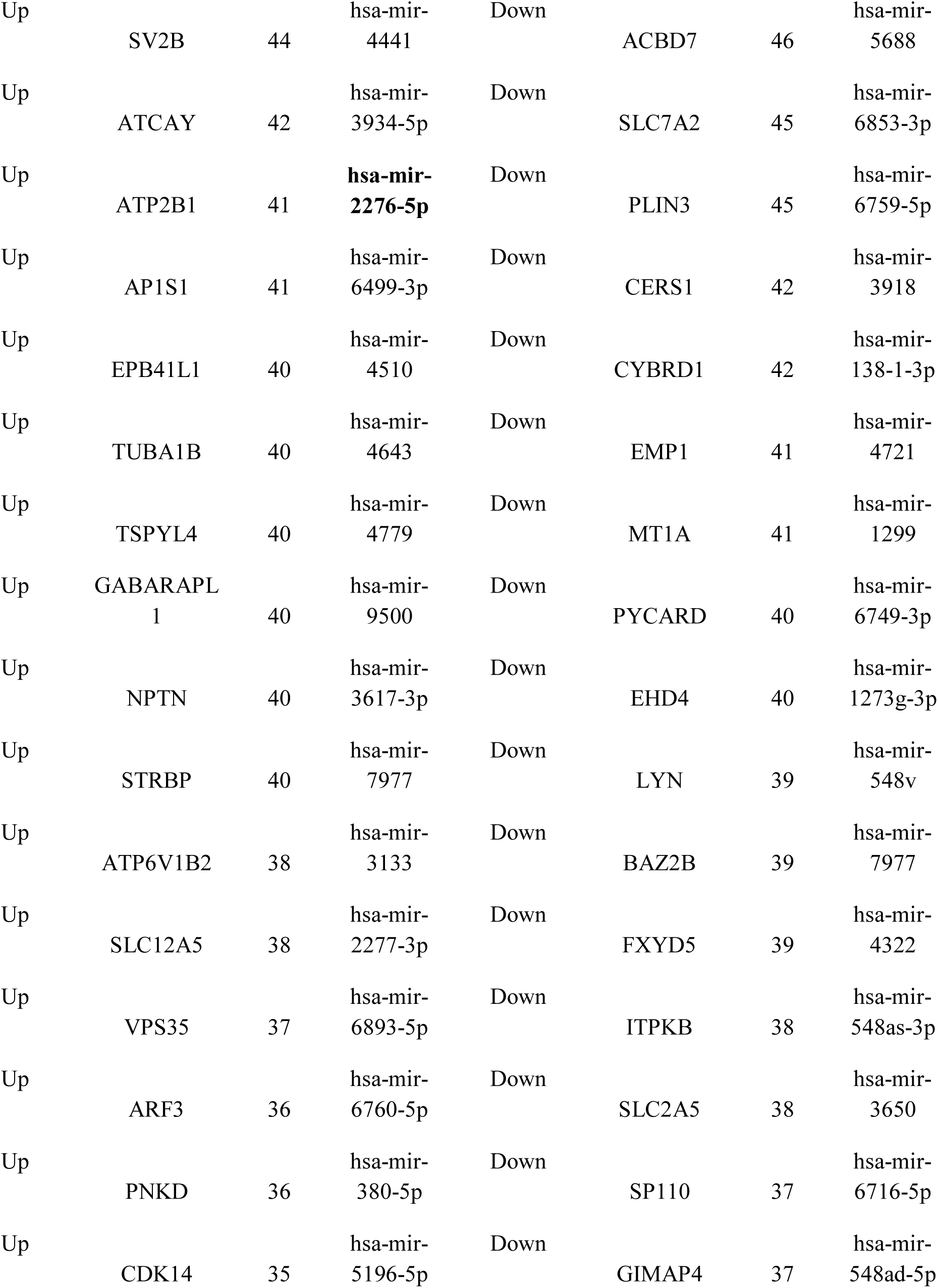
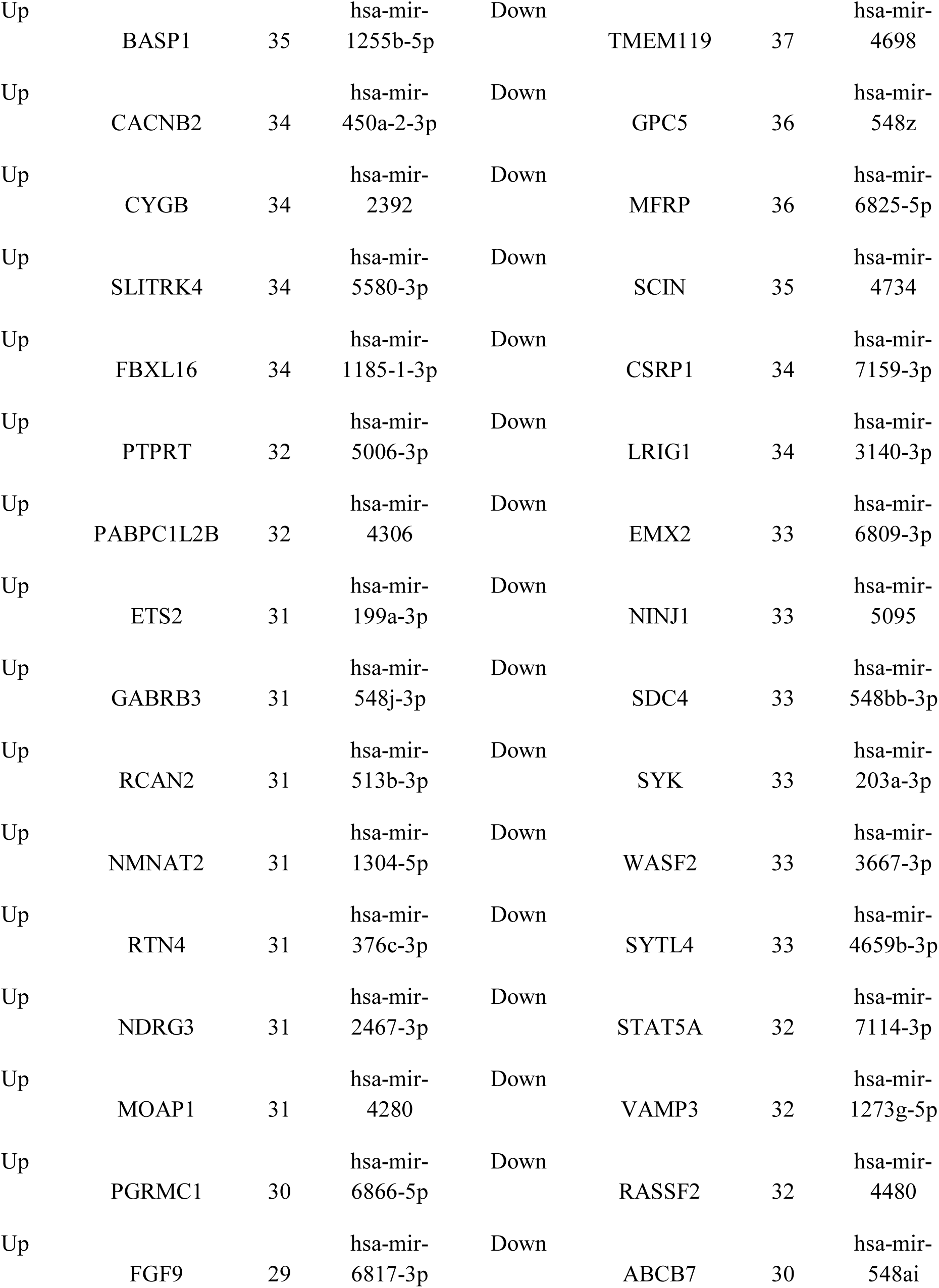
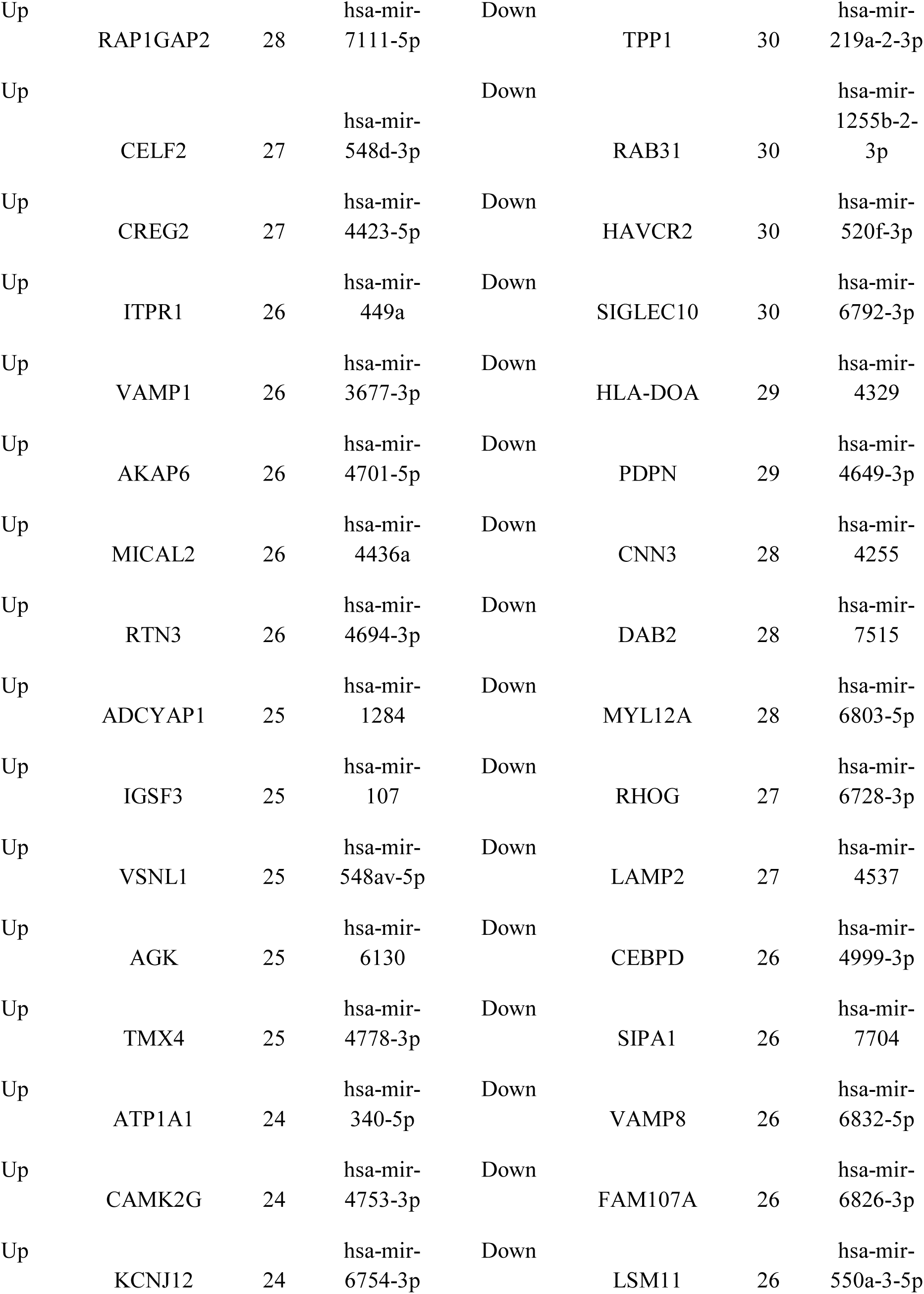
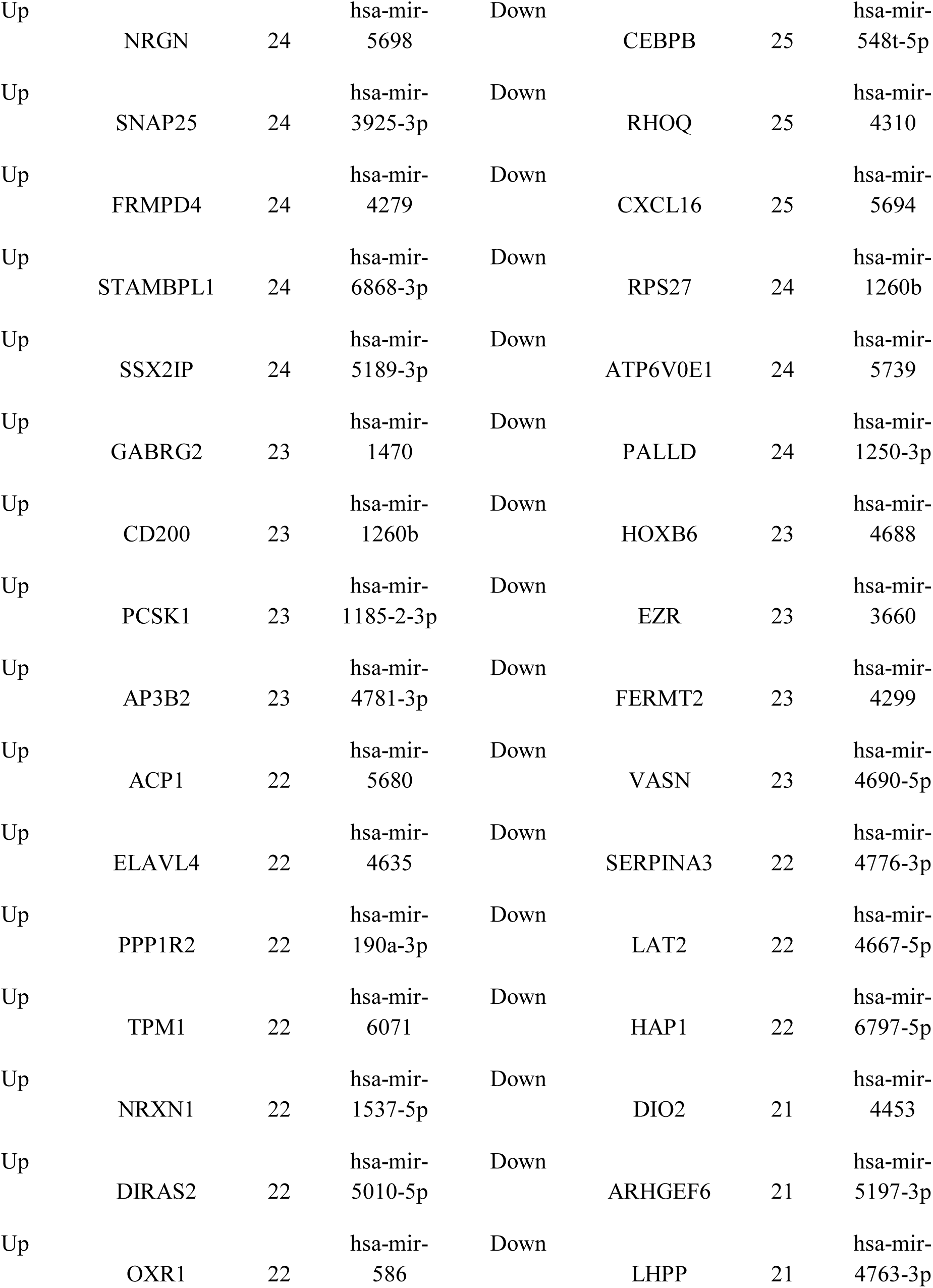
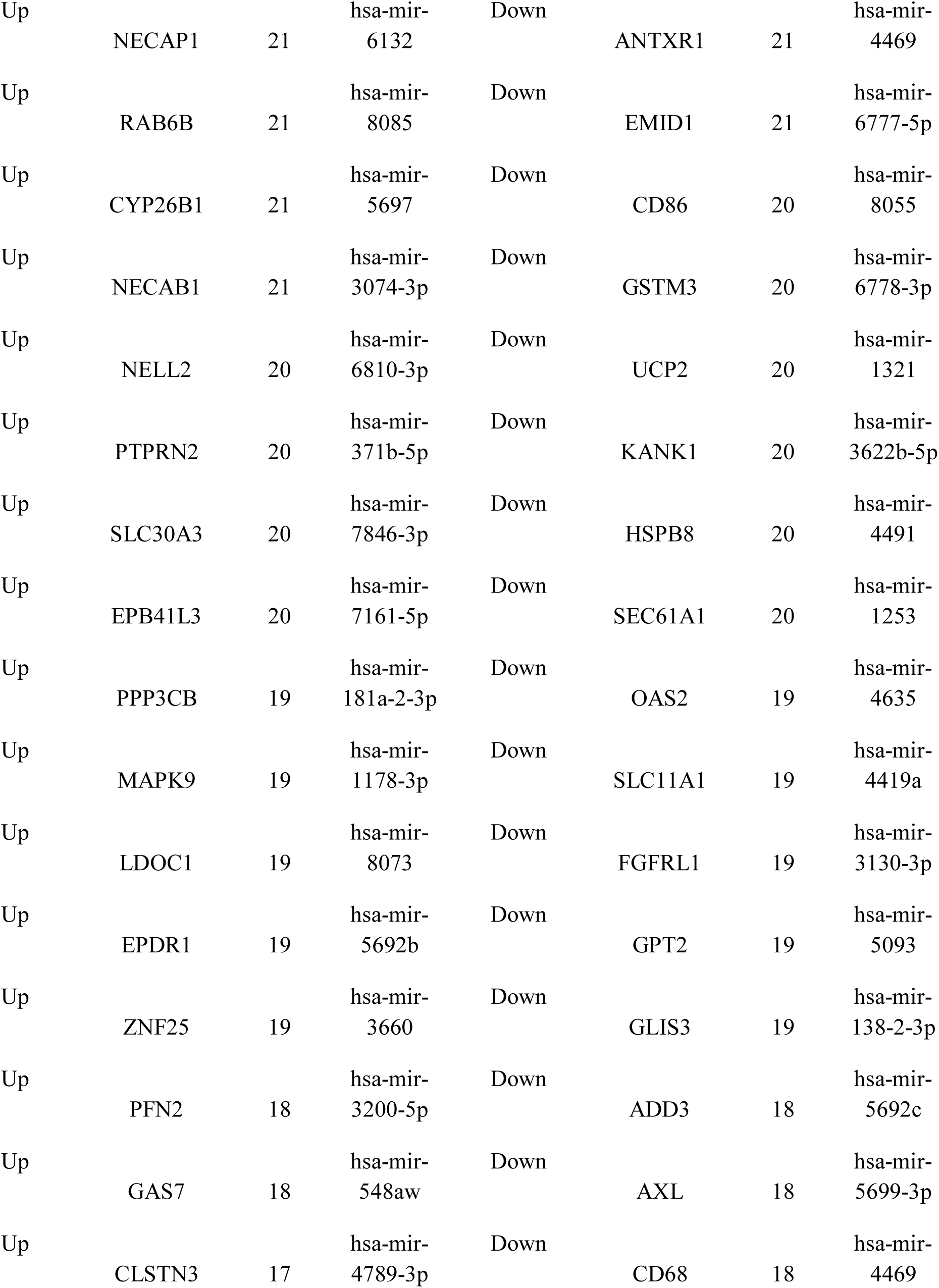
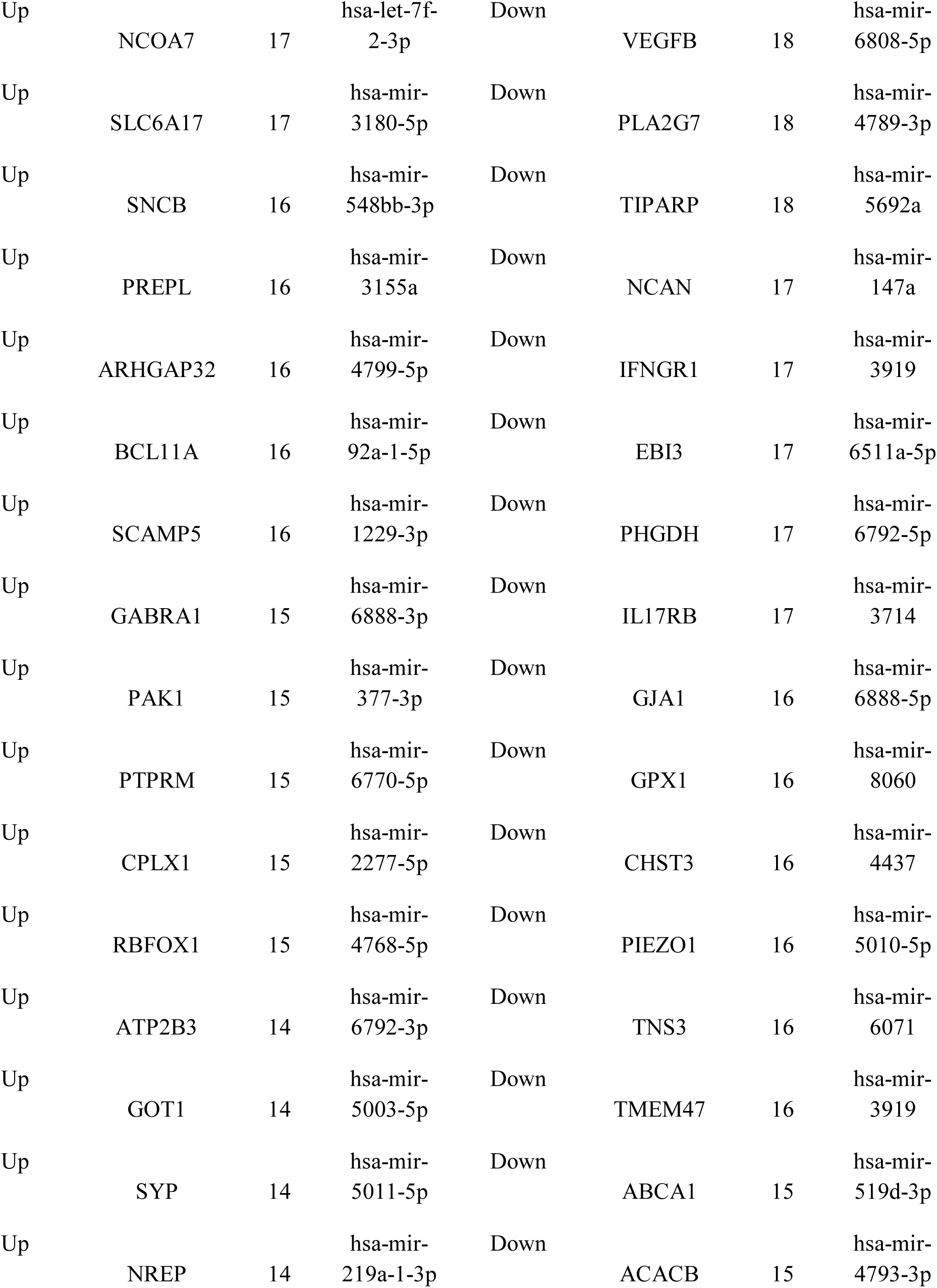
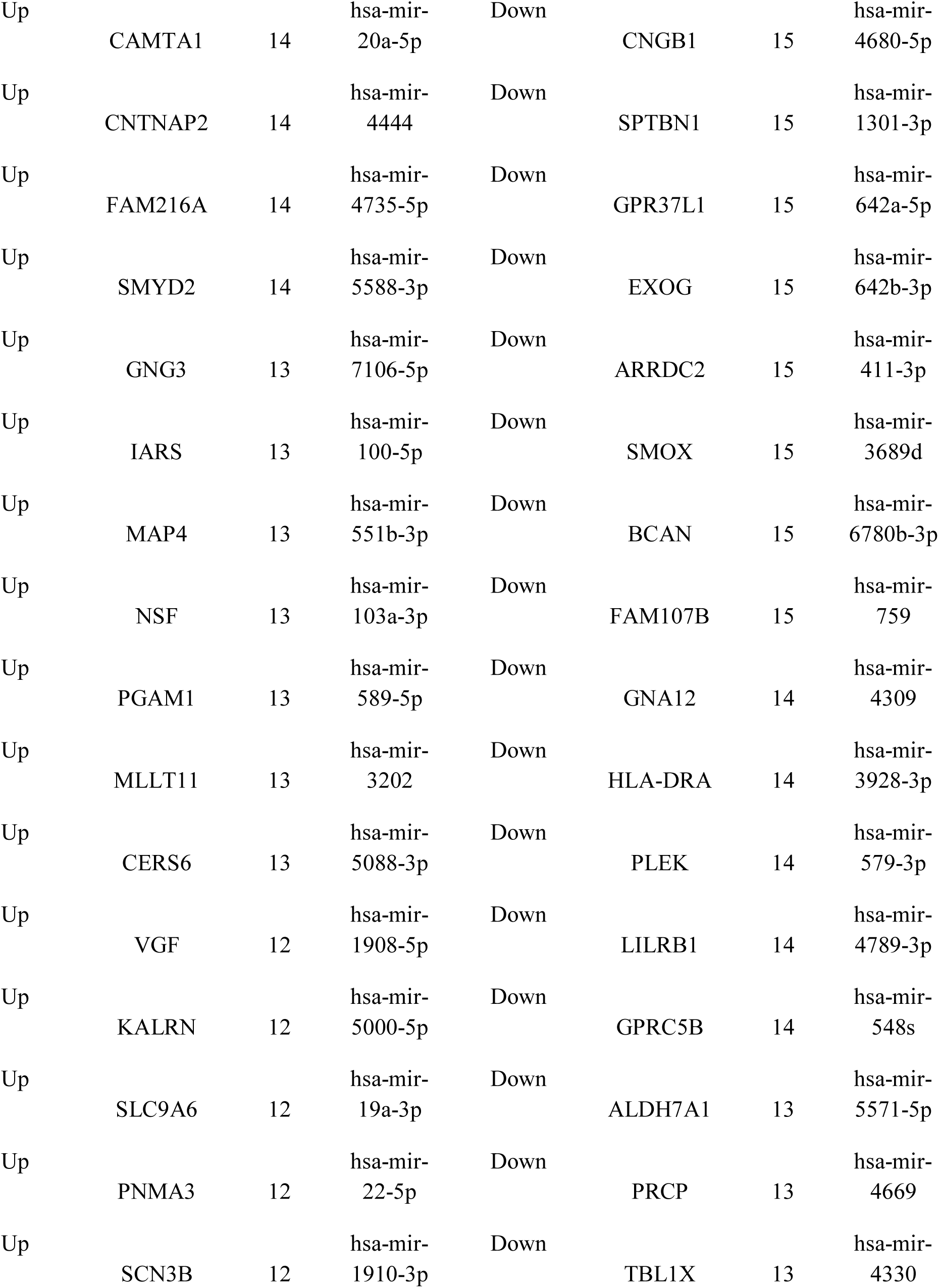
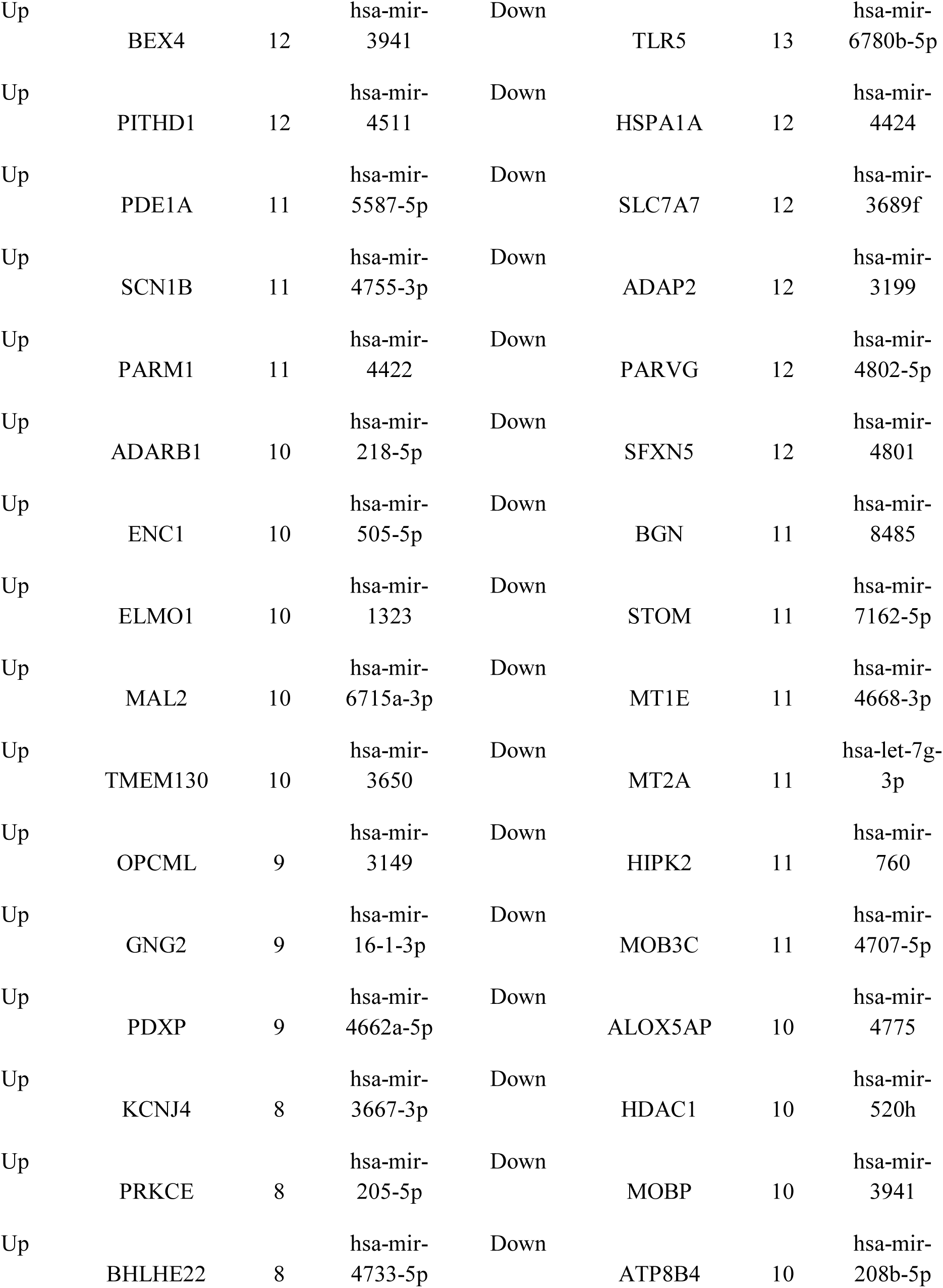
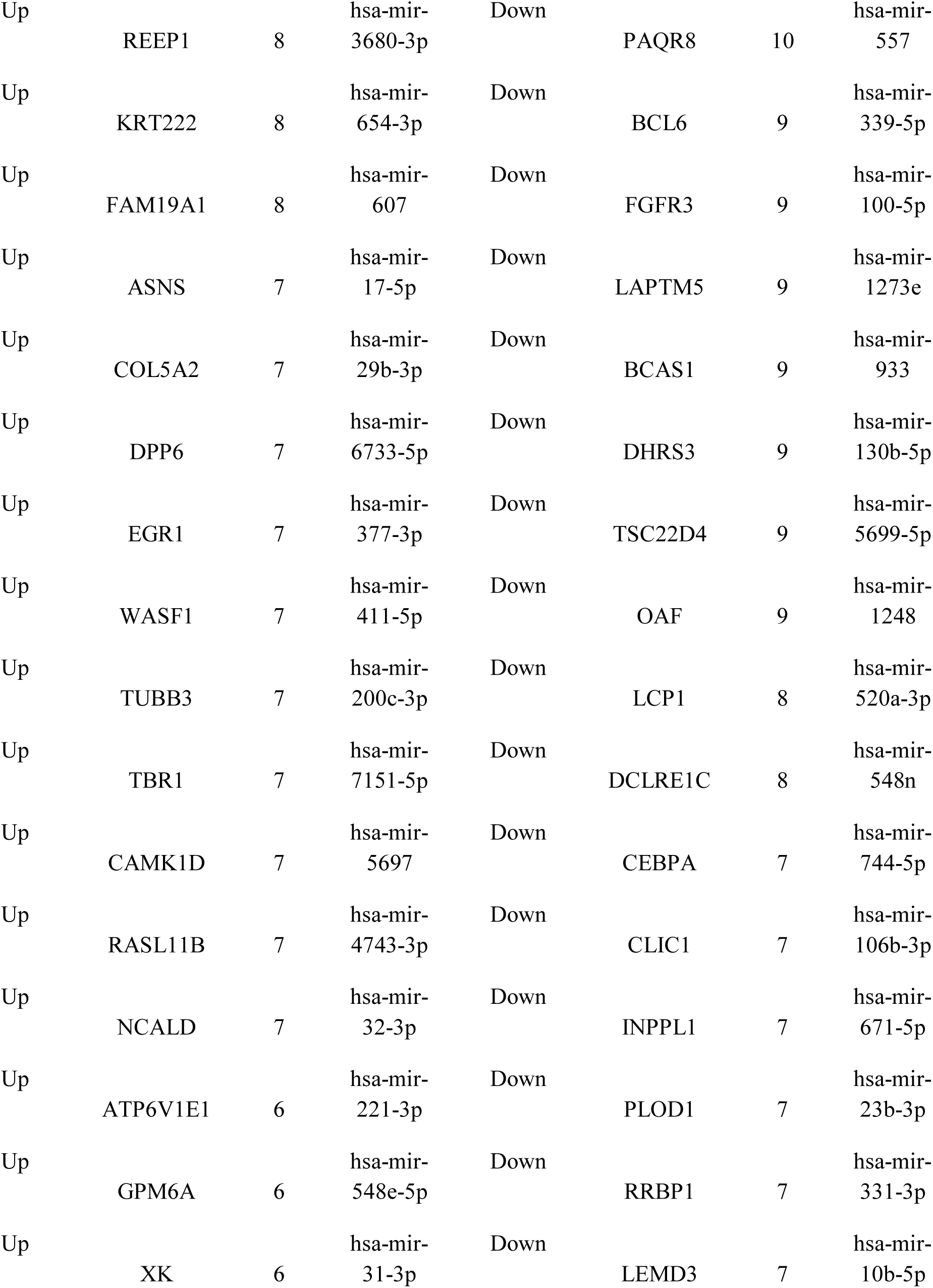
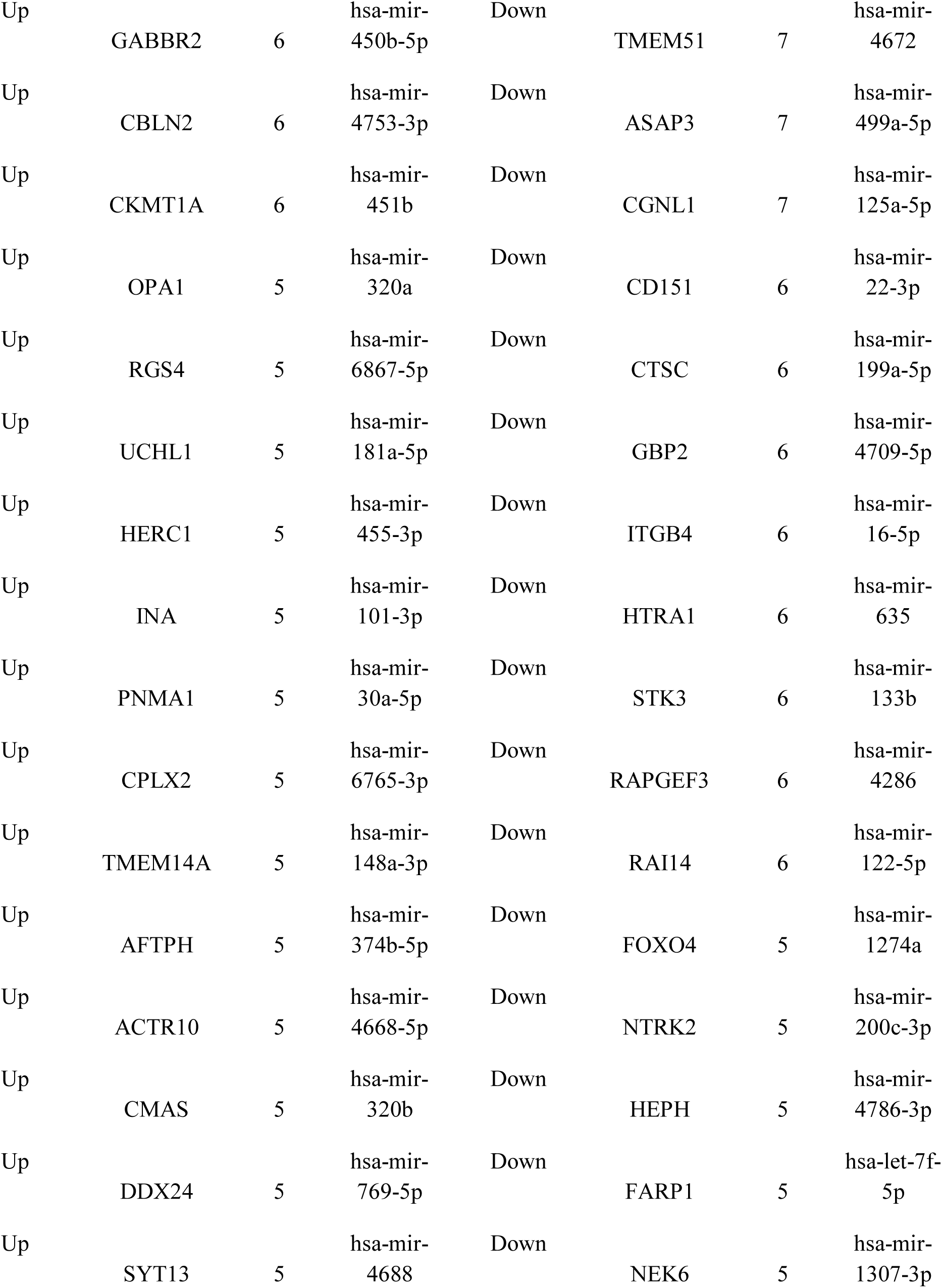
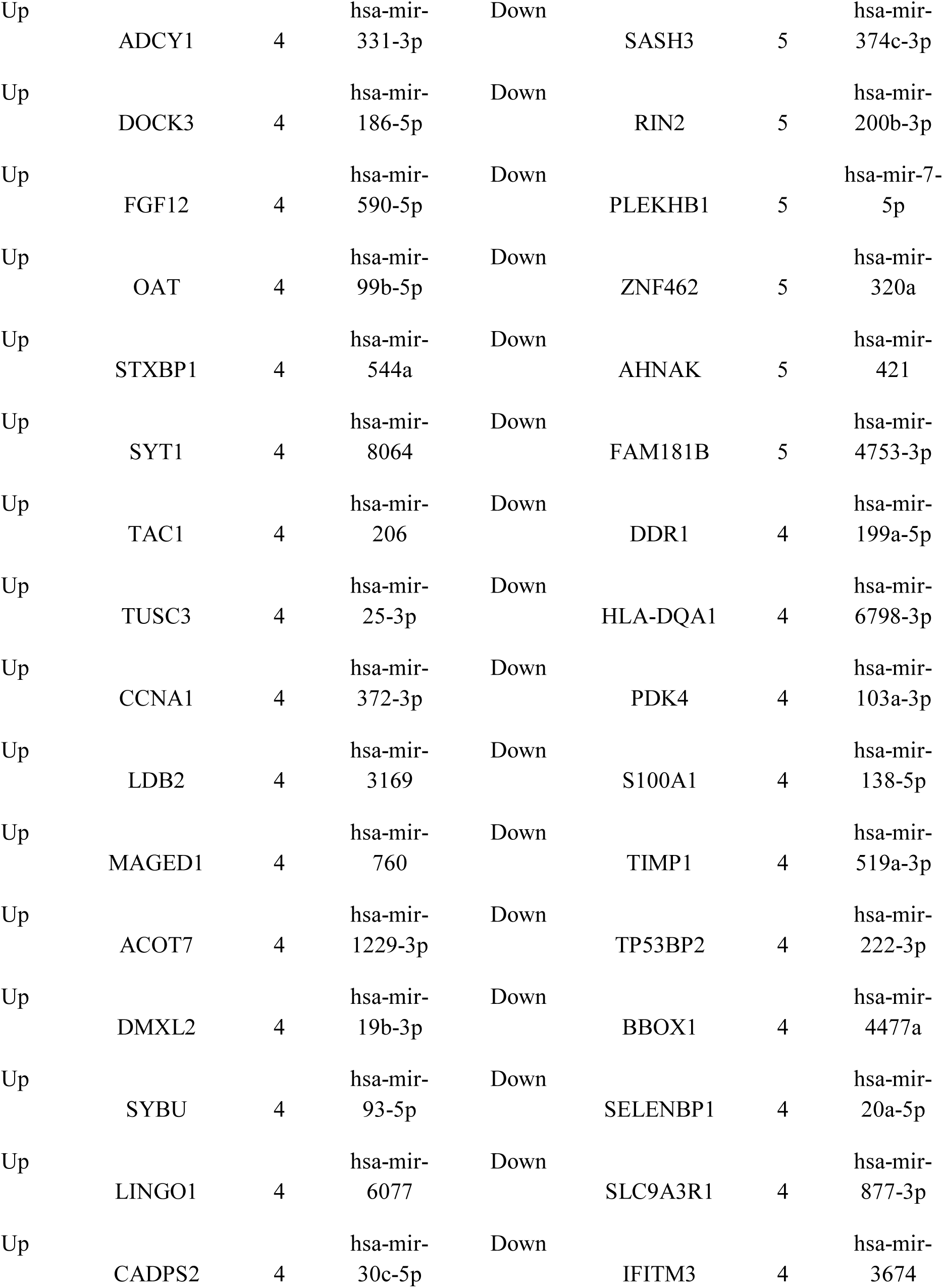
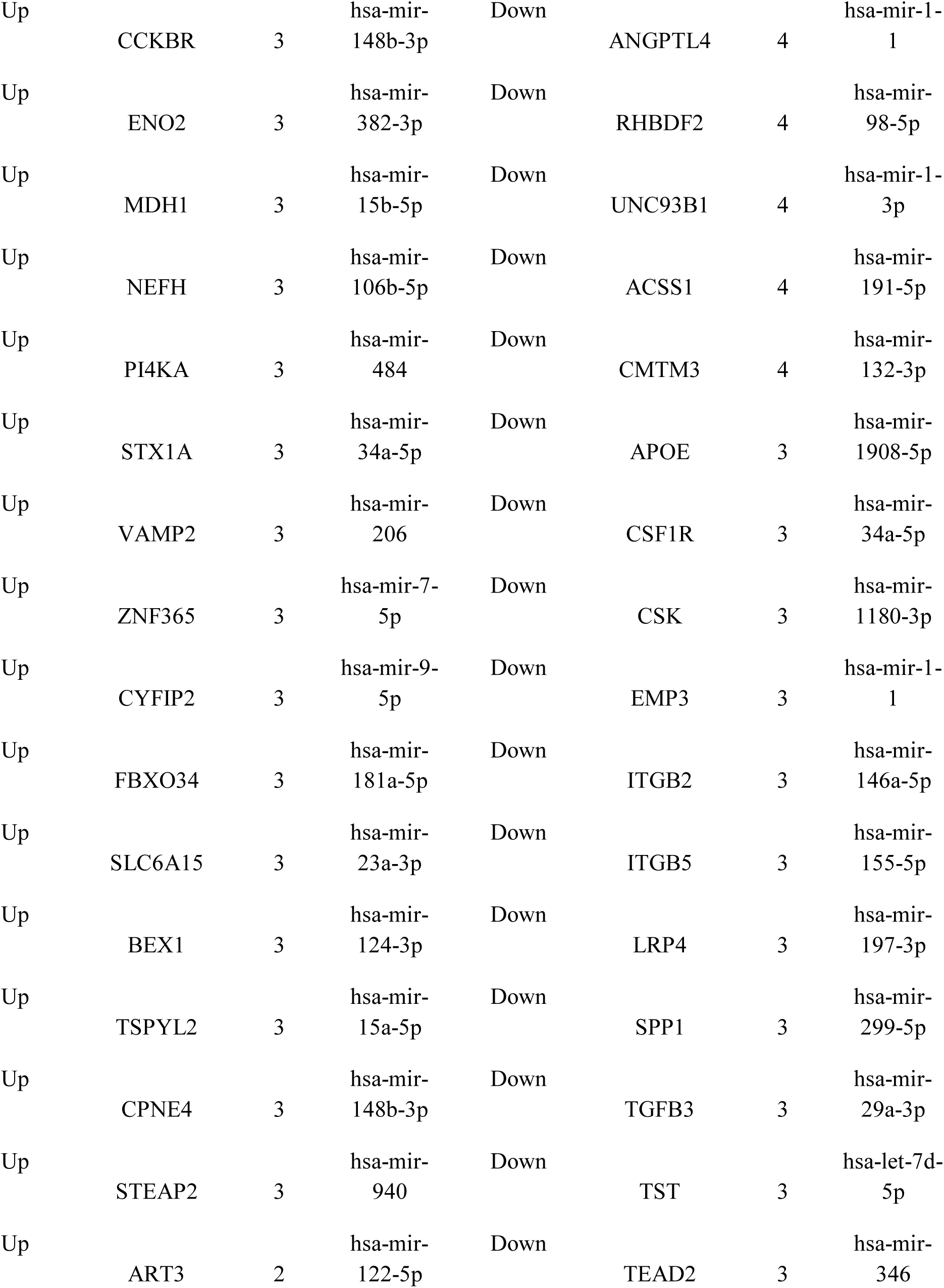
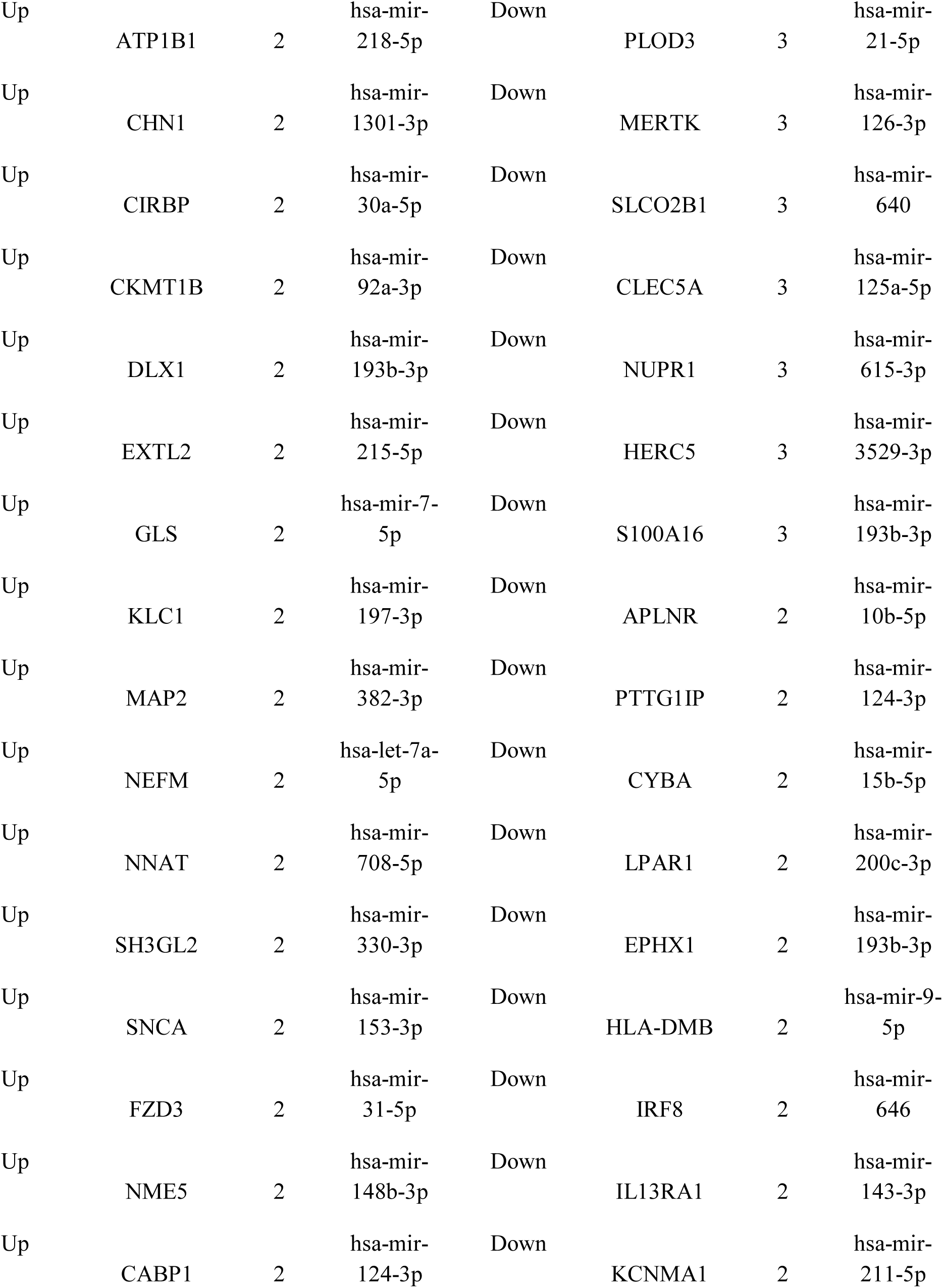
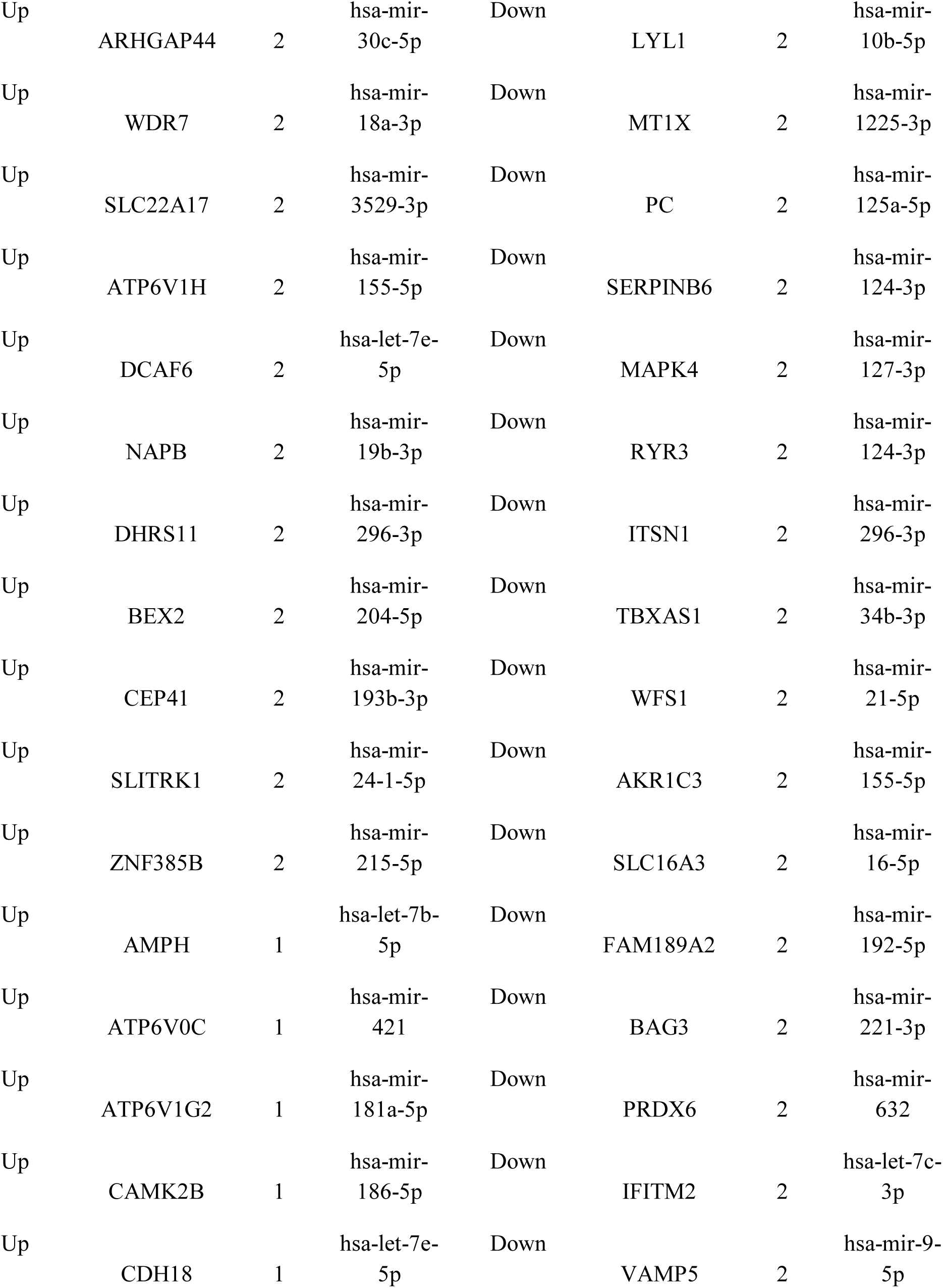
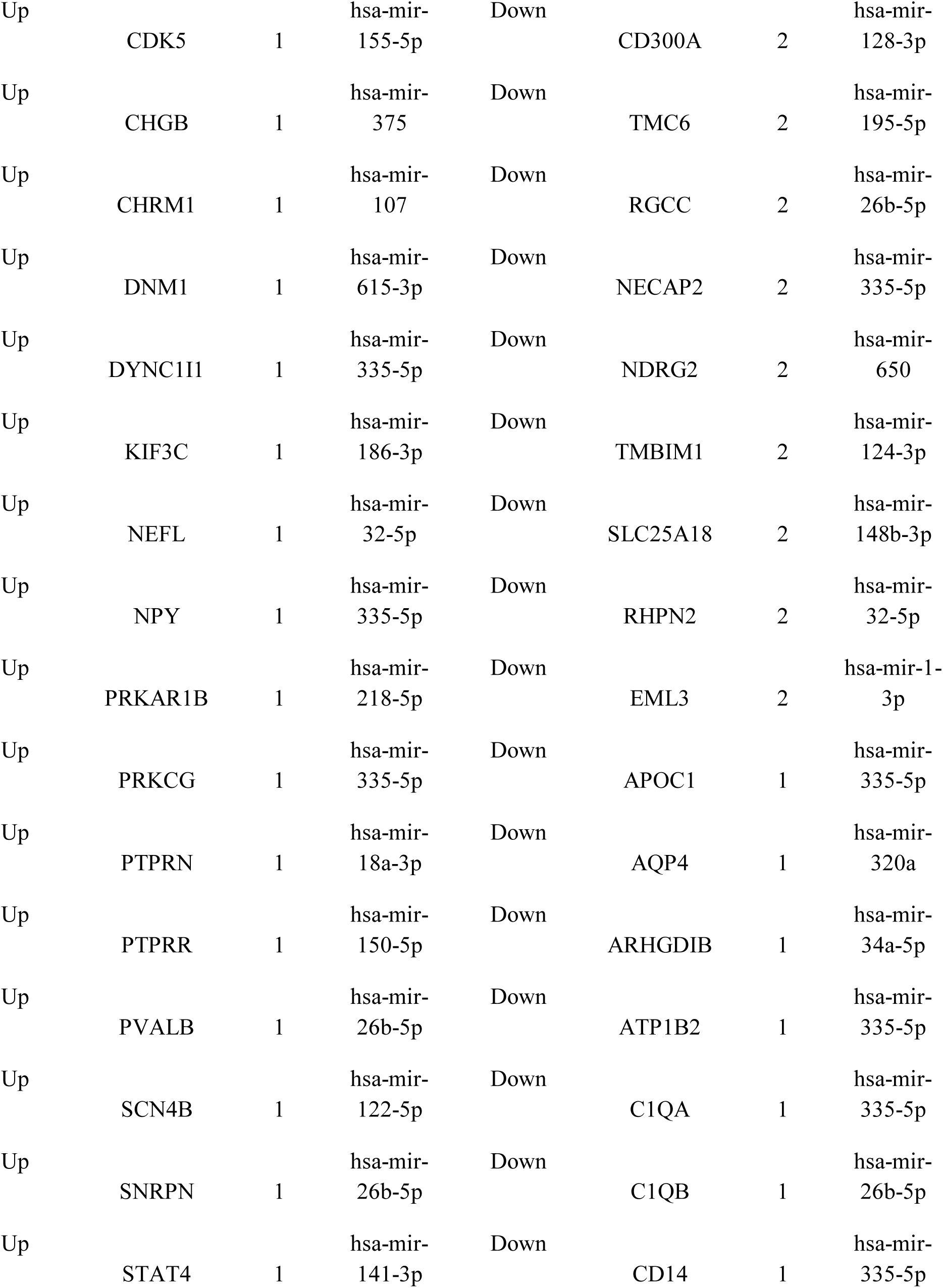
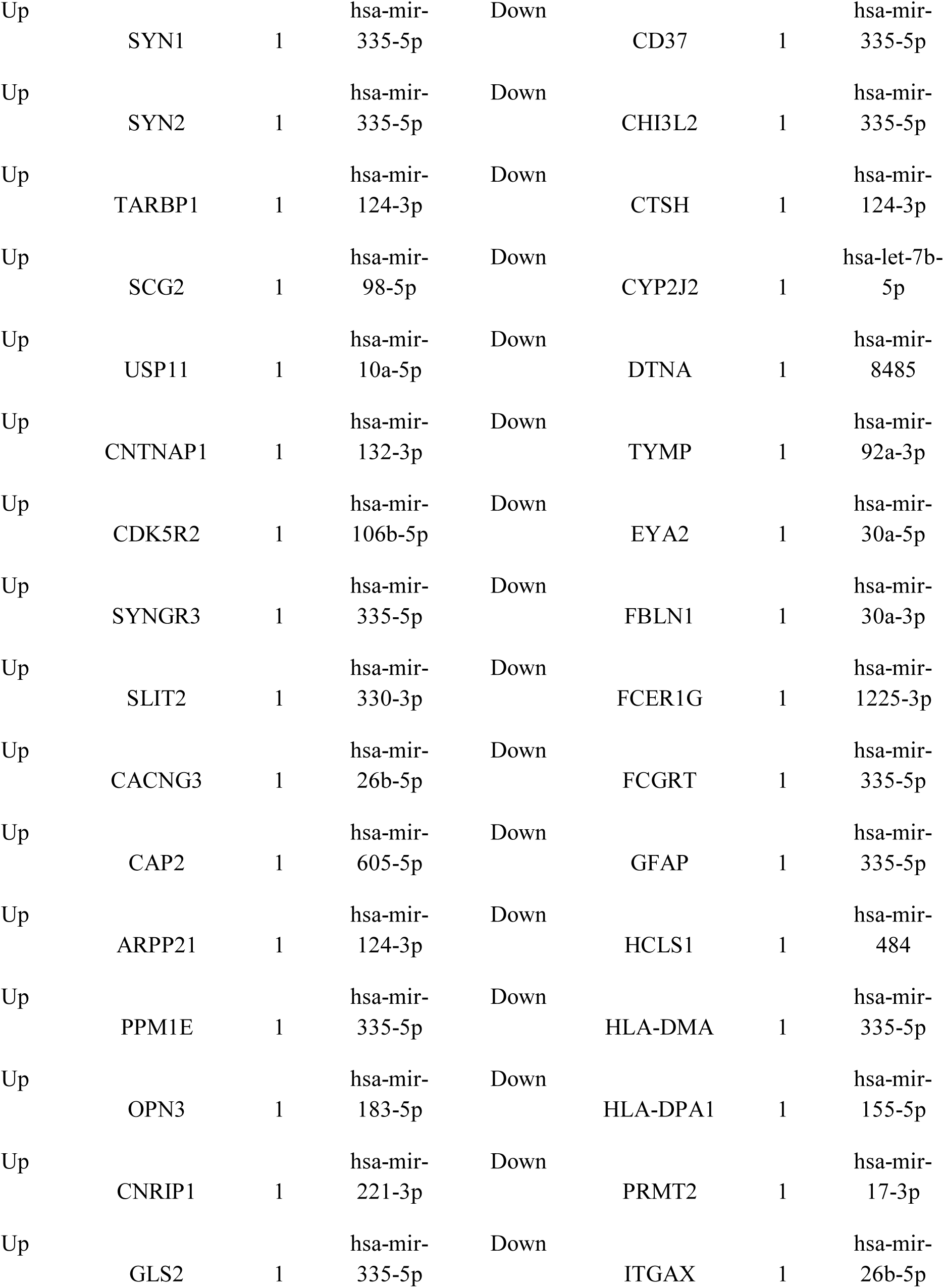
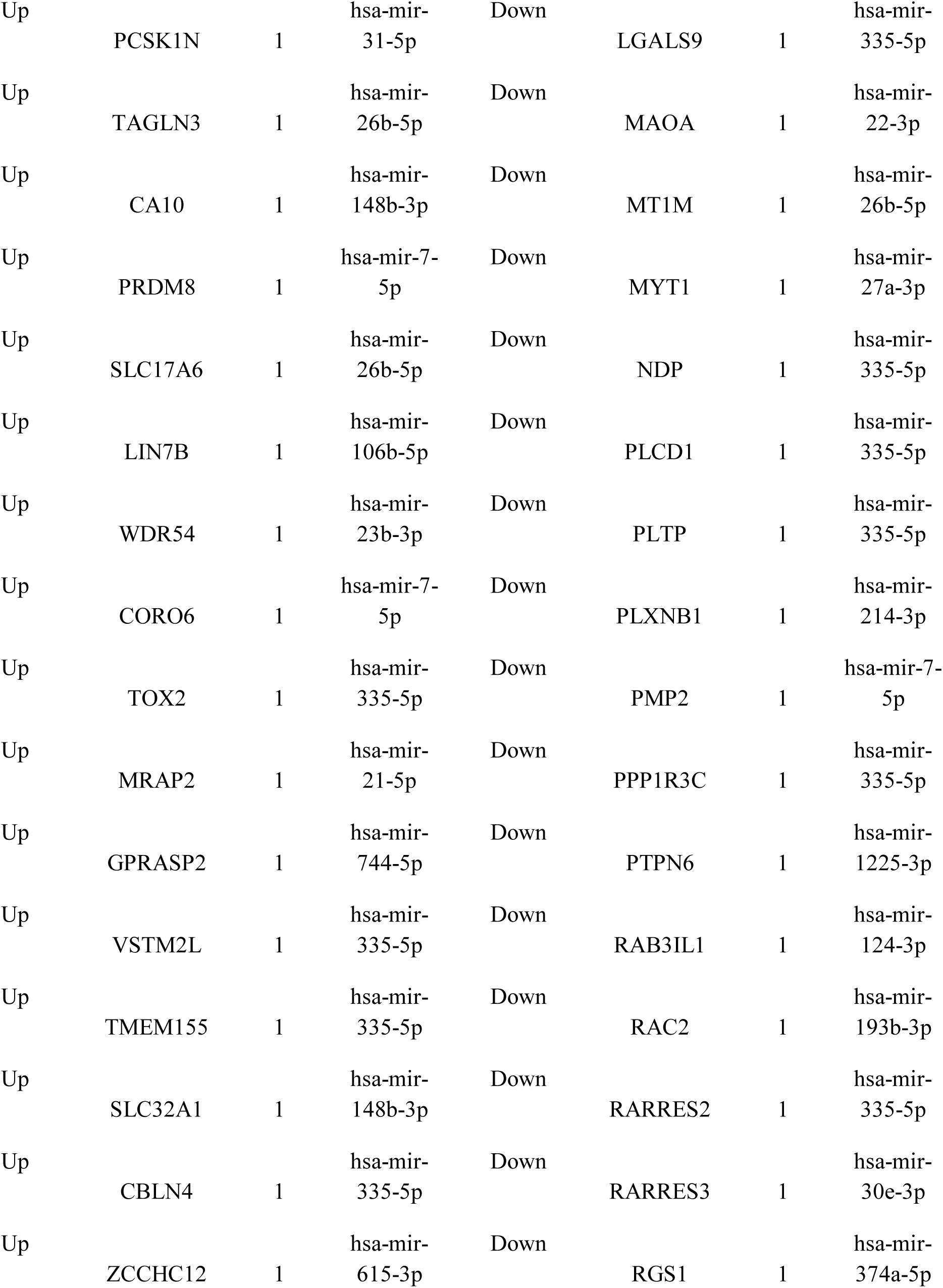
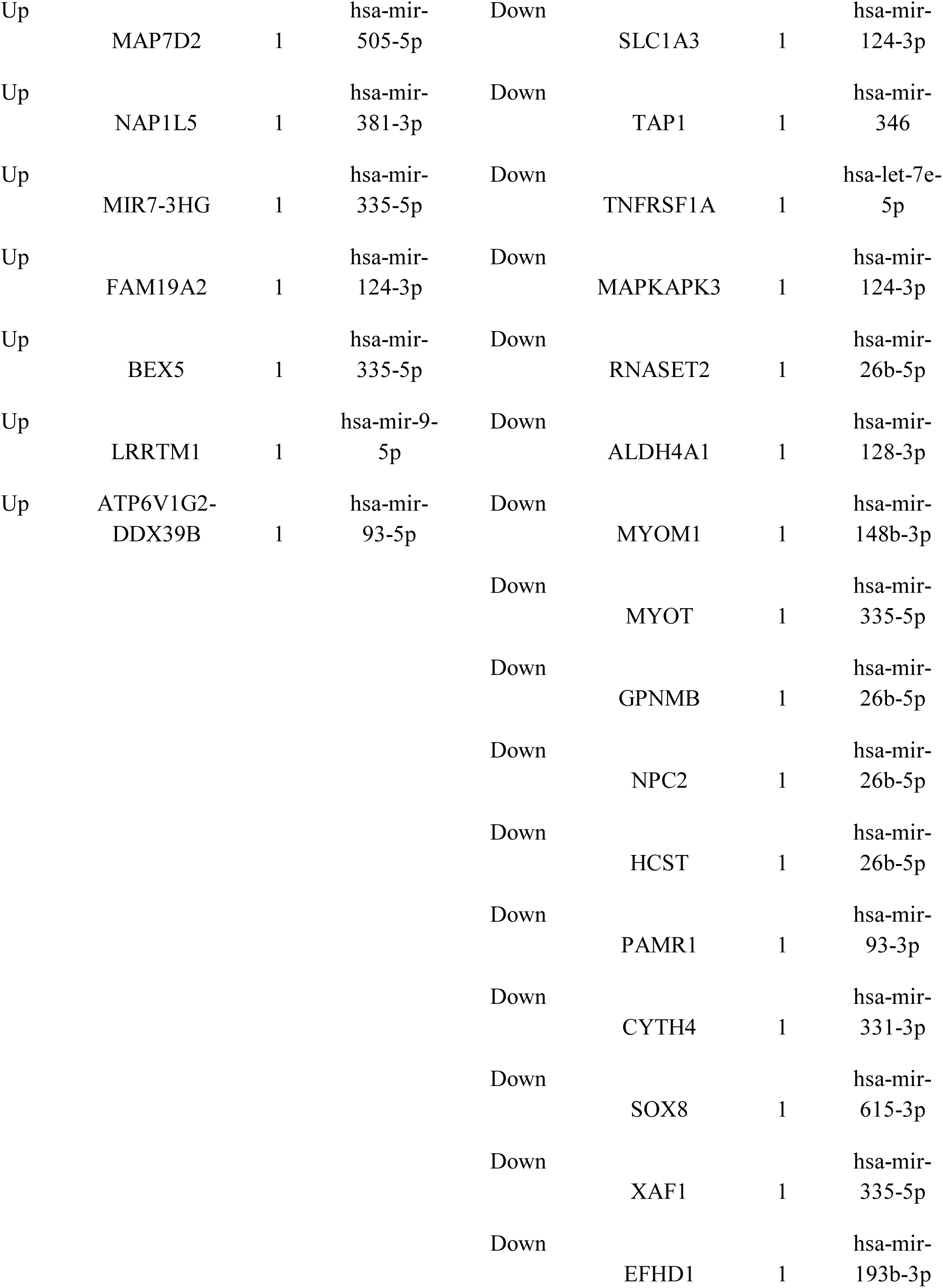
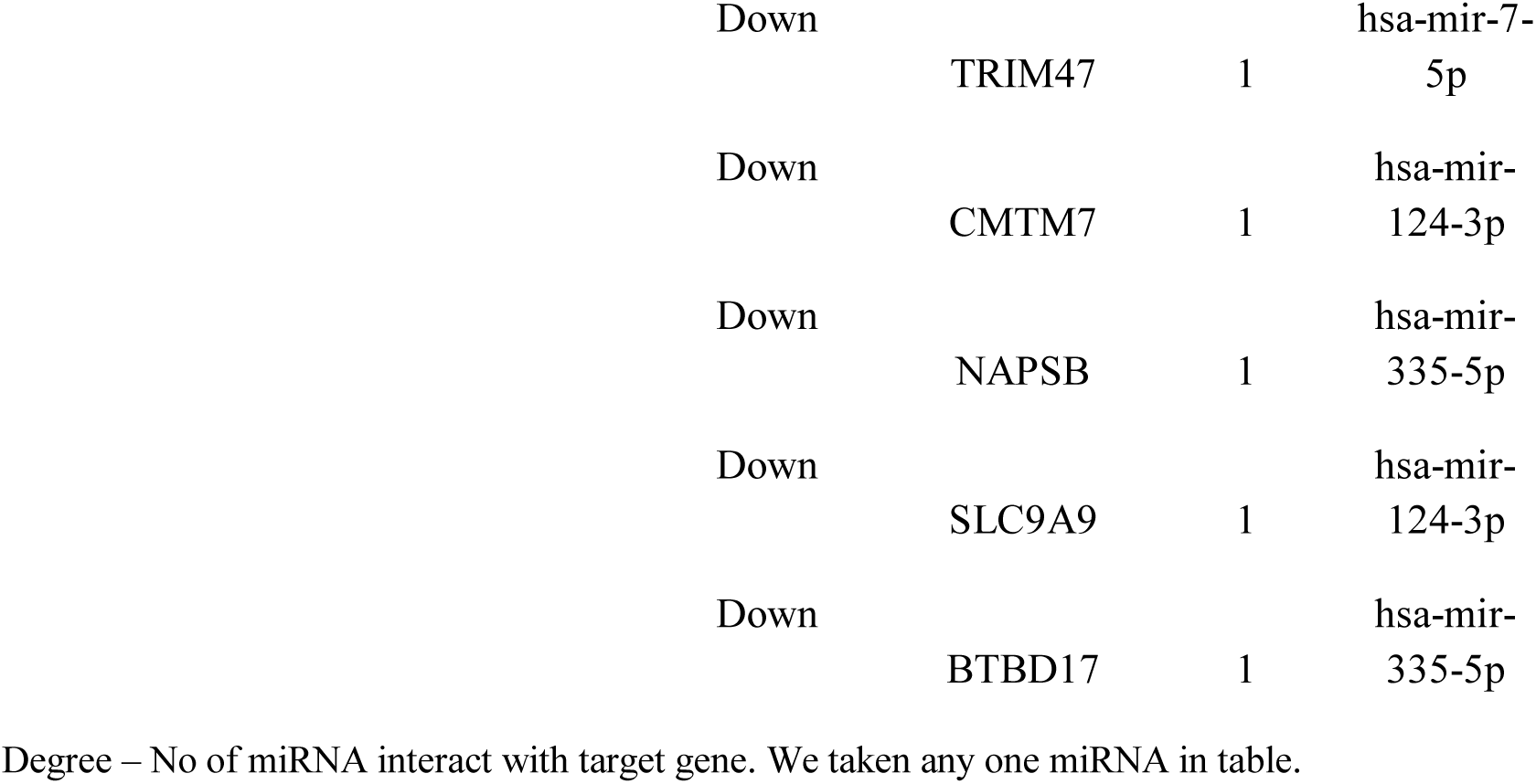
miRNA - target gene interaction table

### Construction of target genes - TF regulatory network

The target gene - TF regulatory network for up regulated genes is shown in Fig.13. Top five up regulated target genes such as CDK14 interacts with 235 TFs (ex, SOX2), YWHAH interacts with 193 TFs (ex, MYC), SYN2 interacts with 182 TFs (ex, REST), SYT1 interacts with 179 TFs (ex, SUZ12) and MICAL2 interacts with 169 TFs (ex, AR) are listed in Table 8. Pathways and GO enrichment analysis revealed that these target genes were markedly enriched in purine nucleotide binding, role of calcineurin-dependent NFAT signaling in lymphocytes, transmission across chemical synapses, synaptic vesicle cycle and cytoskeletal protein binding. Similarly, target gene - miRNA regulatory network for down regulated genes is shown in Fig.14. Top five down regulated target genes such as HSPA1A interacts with 224 TFs (ex, SOX2), CD300A interacts with 212 TFs (ex, SPI1), HLA-DOA interacts with 197 TFs (ex, EGR1), TST interacts with 184 TFs (ex, HNF4A) and HSPB1 interacts with 179 TFs (ex, MYC) are listed in Table 8. Pathways and GO enrichment analysis revealed that these target genes were markedly enriched in toxoplasmosis, neutrophil degranulation, extracellular space and cell activation.

**Fig. 13.**
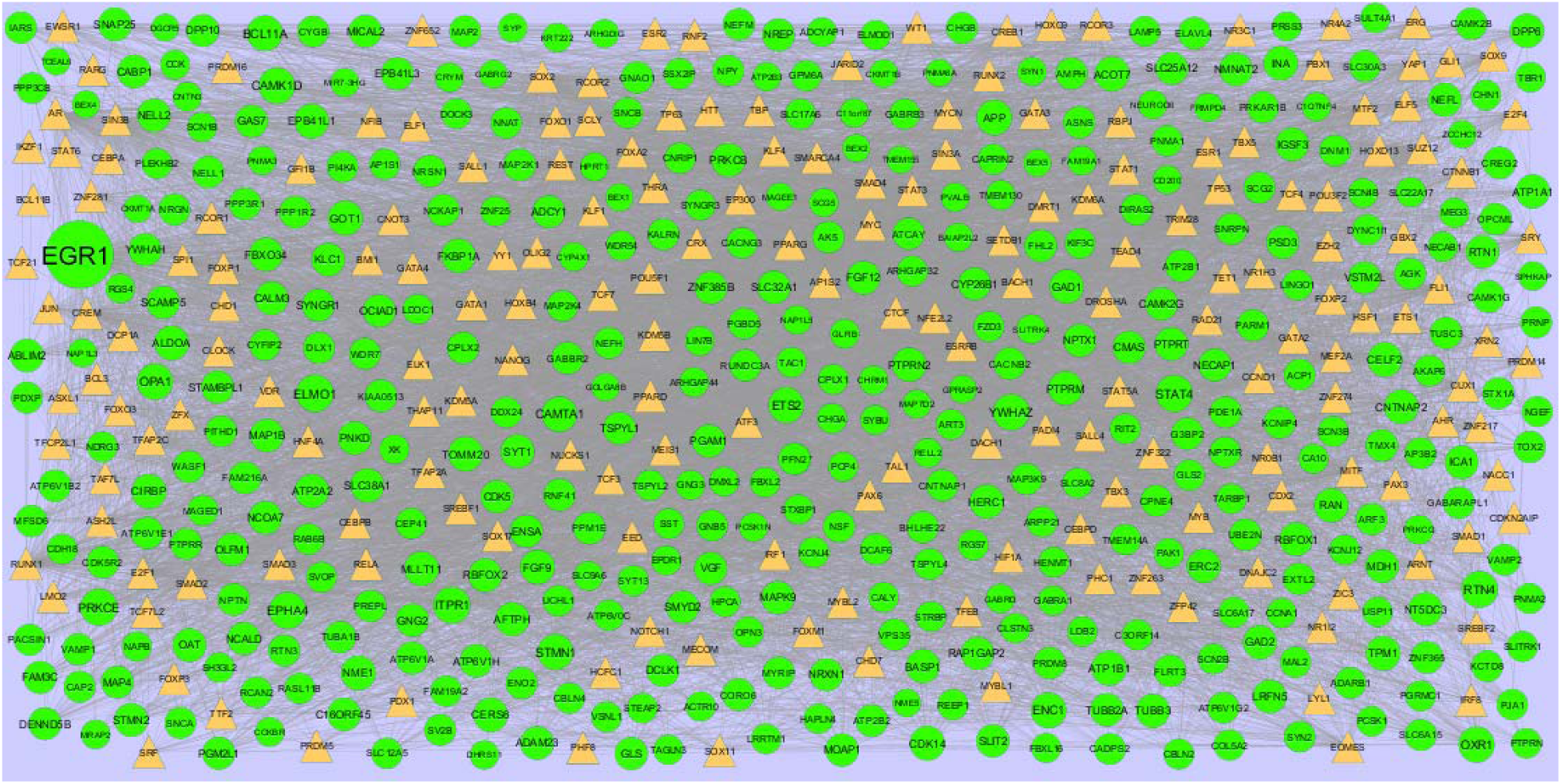
TF-gene network of predicted target up regulated genes. (Orange triangle - TFs and green circles-target up regulated genes)

**Fig. 14.**
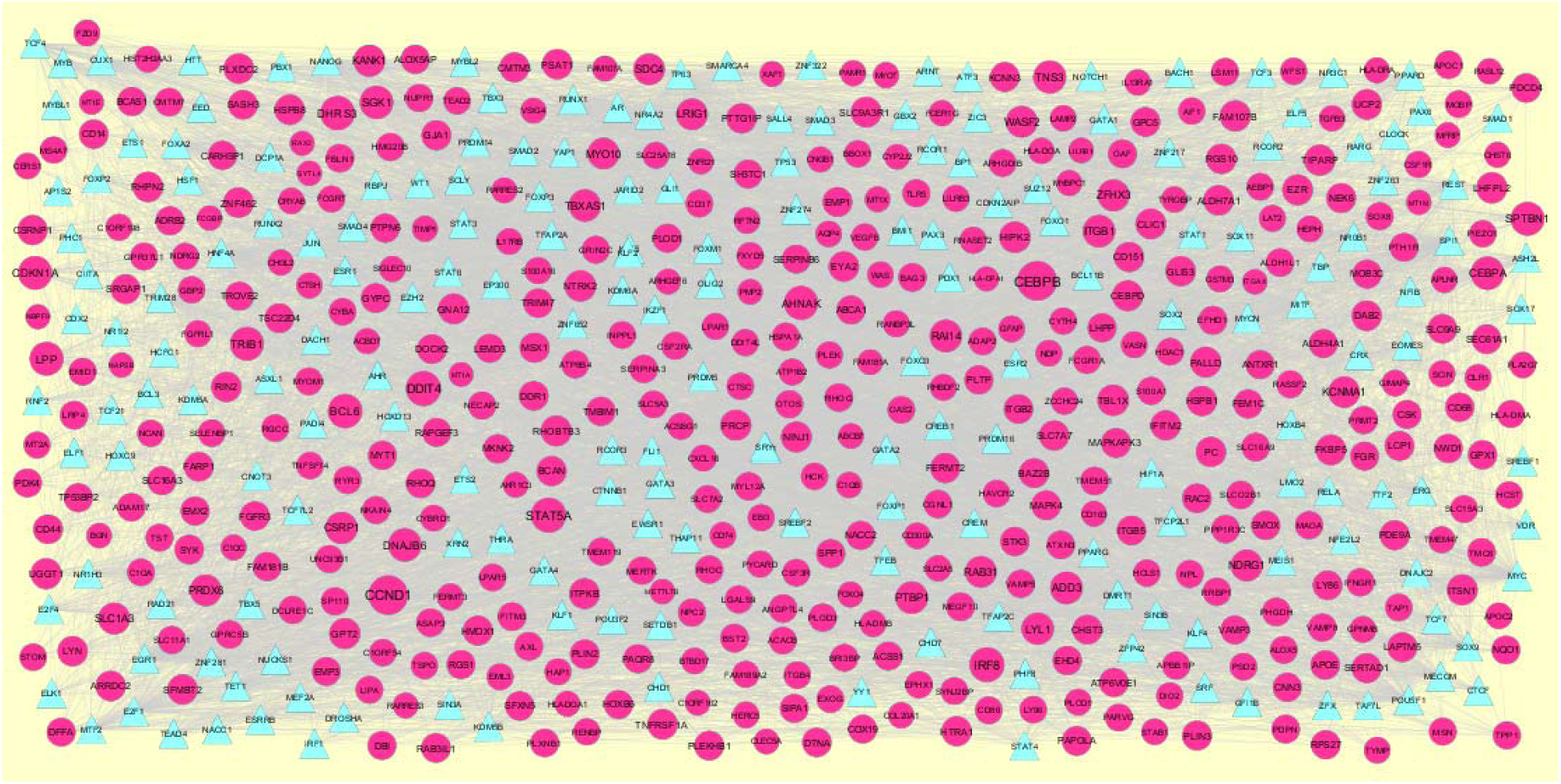
TF-gene network of predicted target down regulated genes. (Blue triangle - TFs and pink circles-target up regulated genes)

**Table 8.**
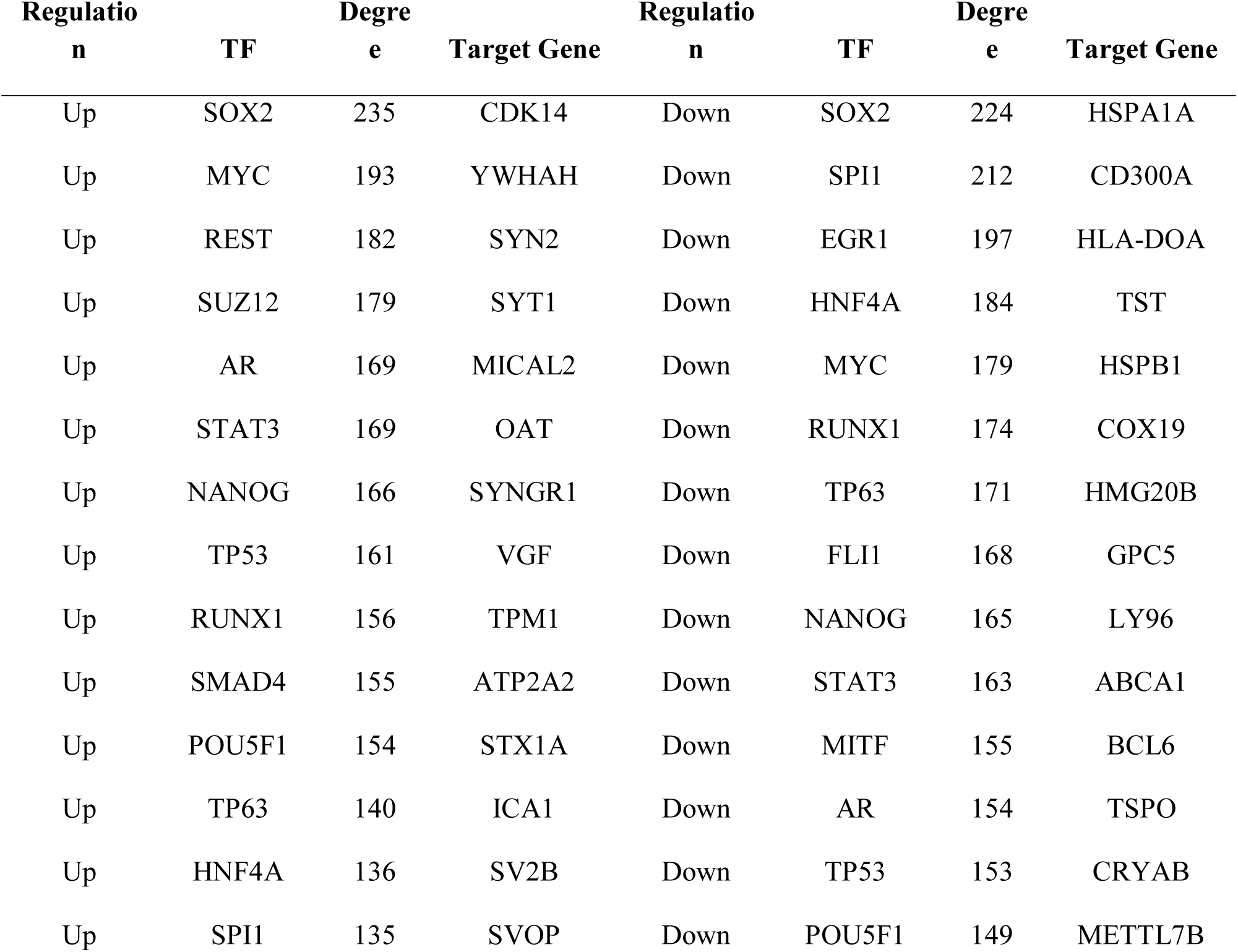
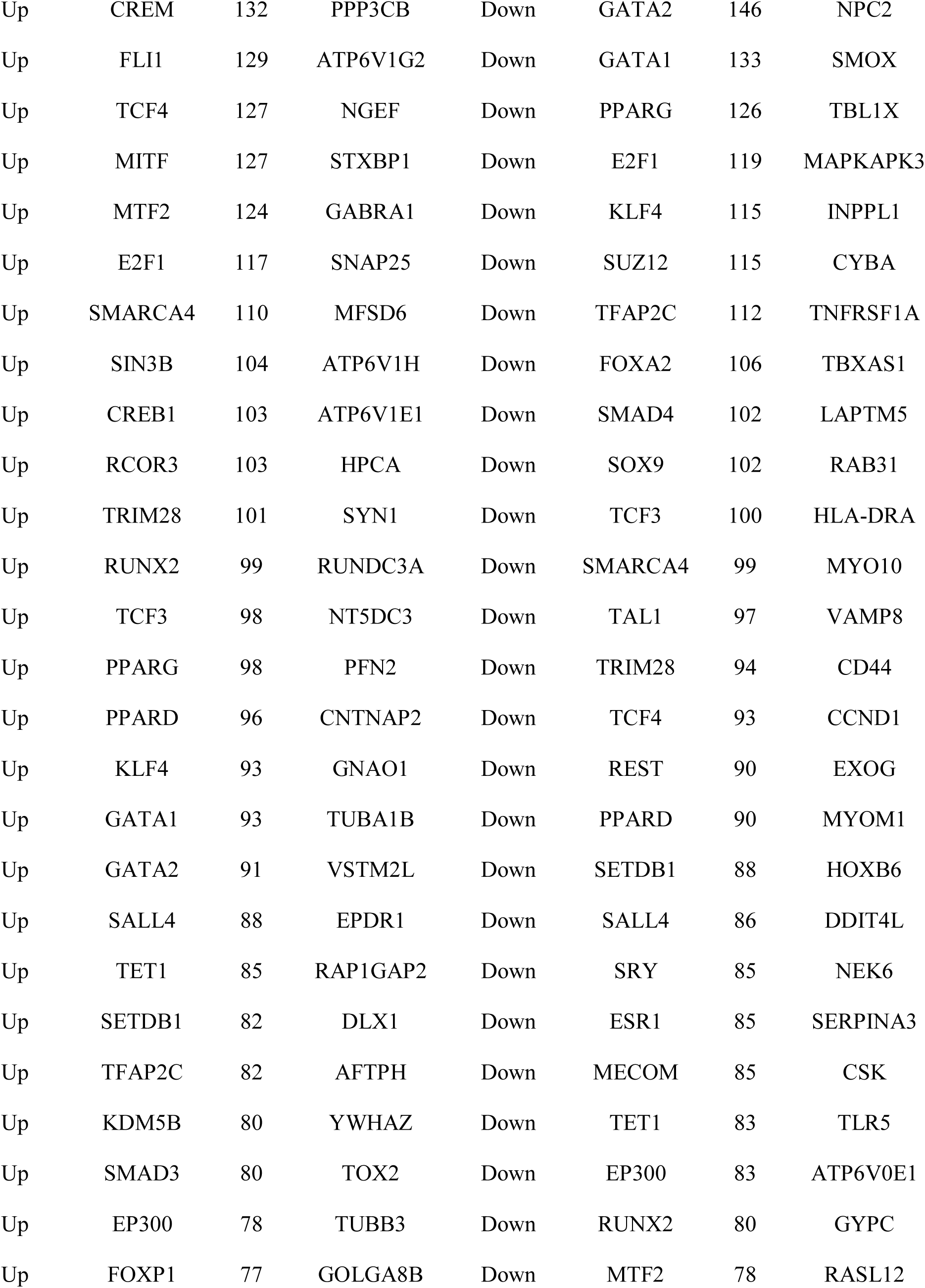
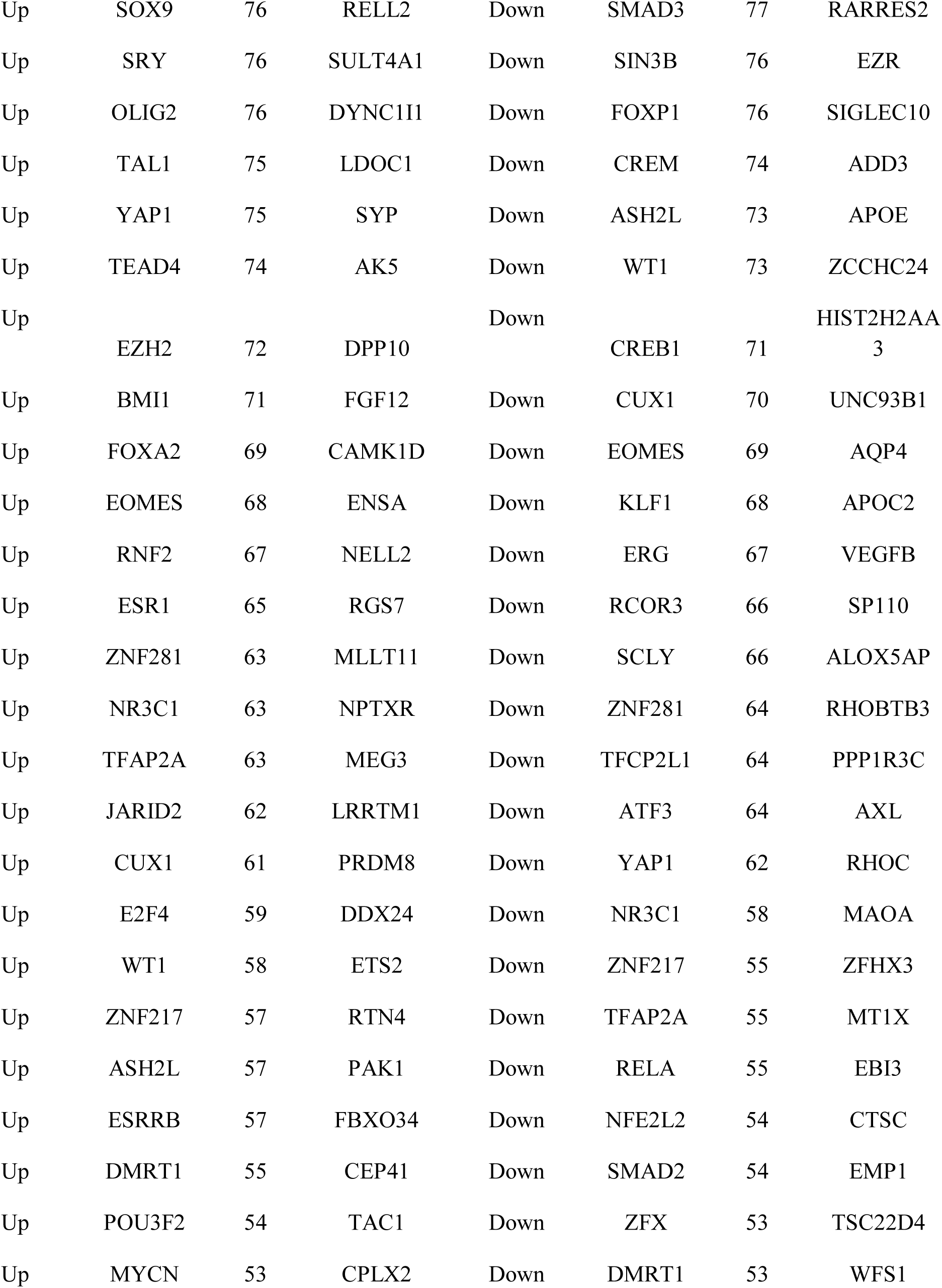
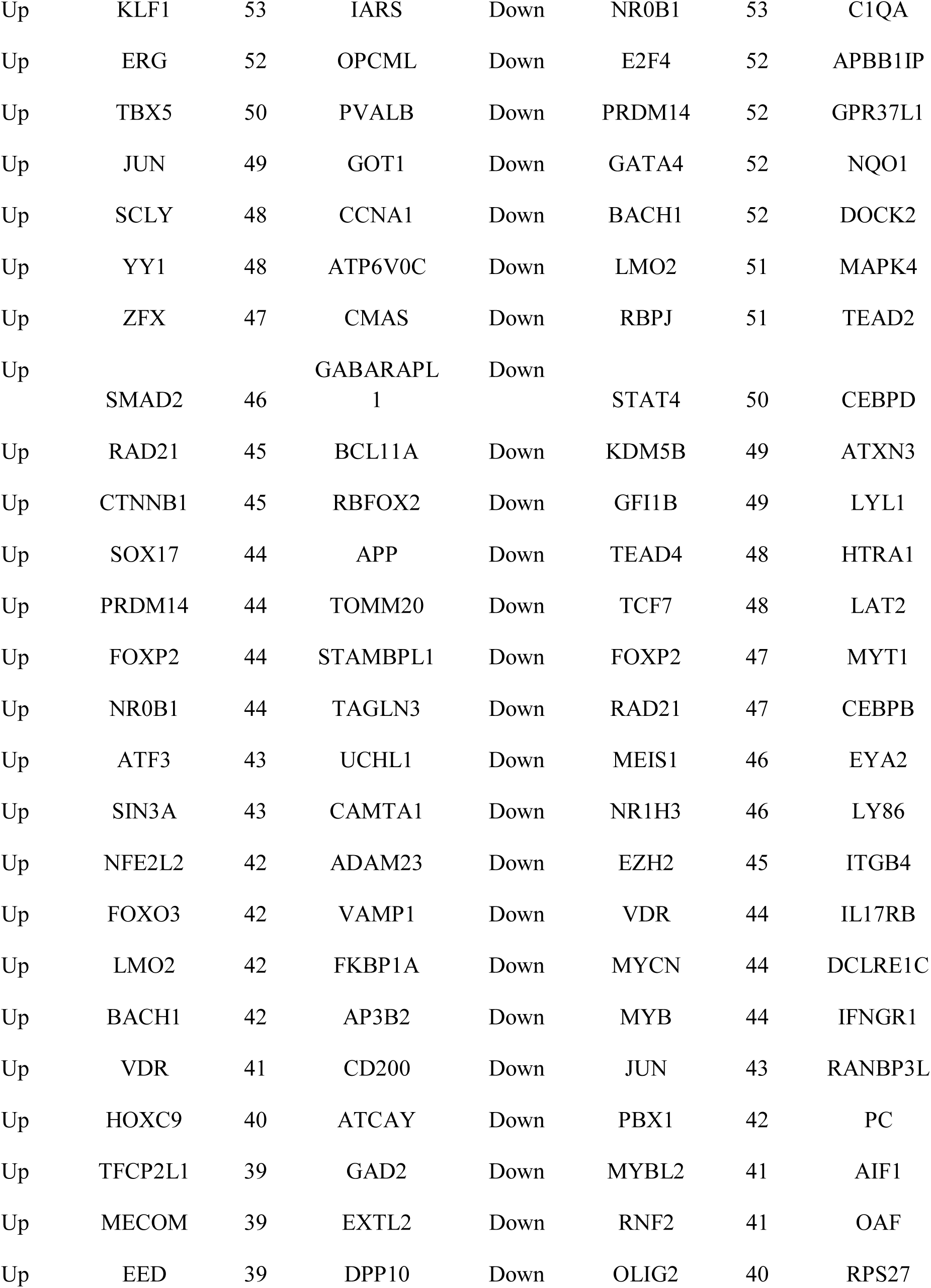
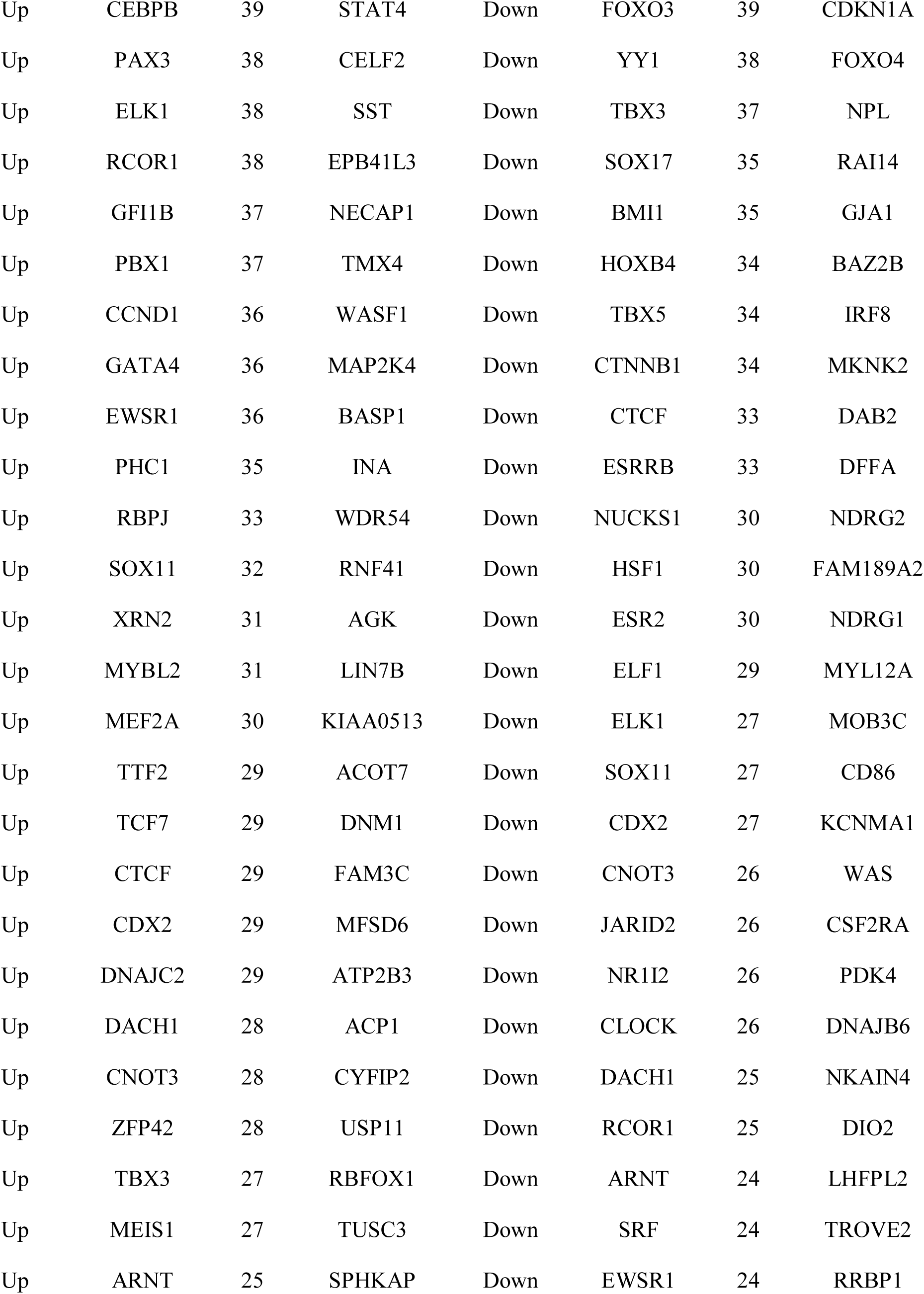
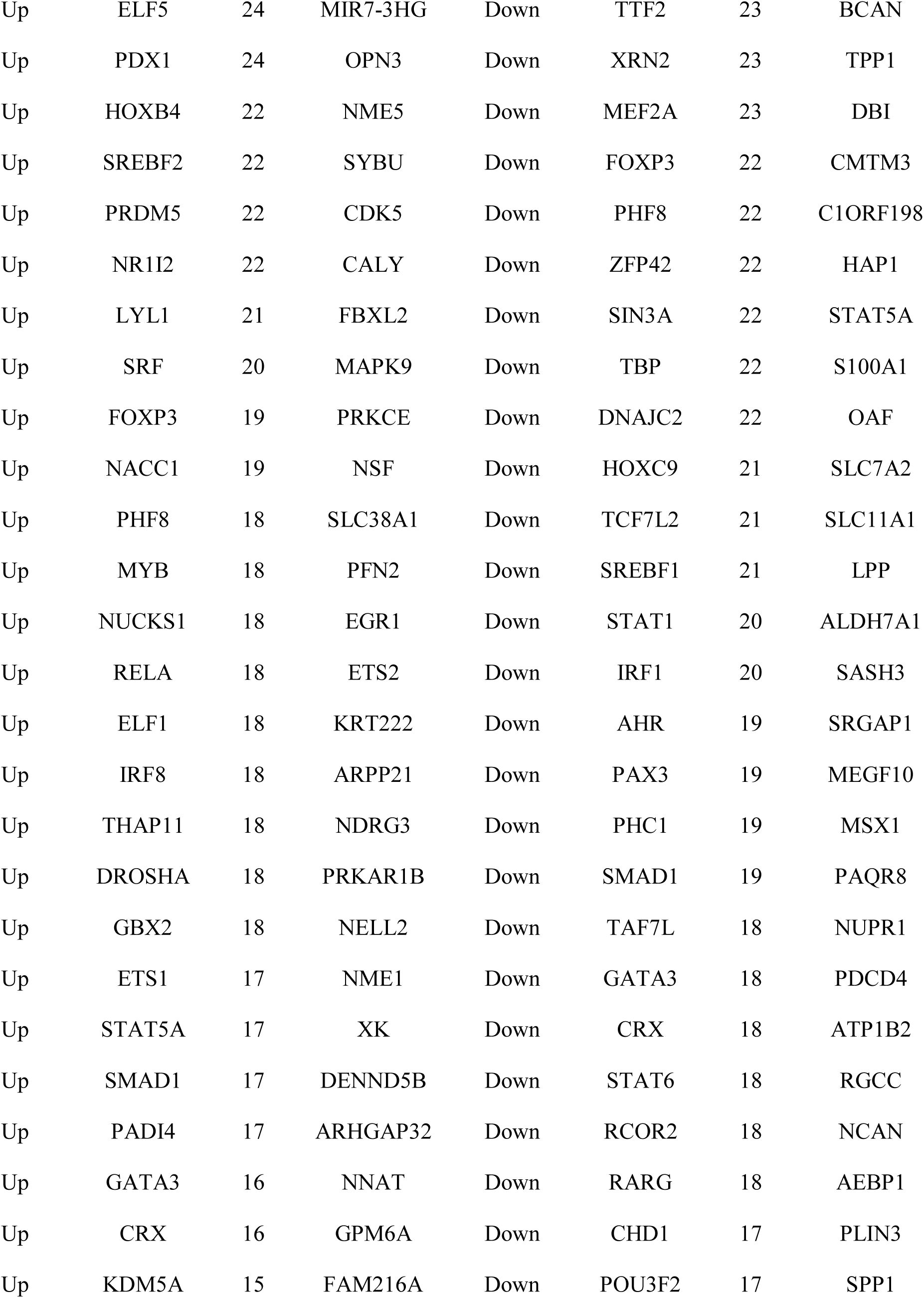
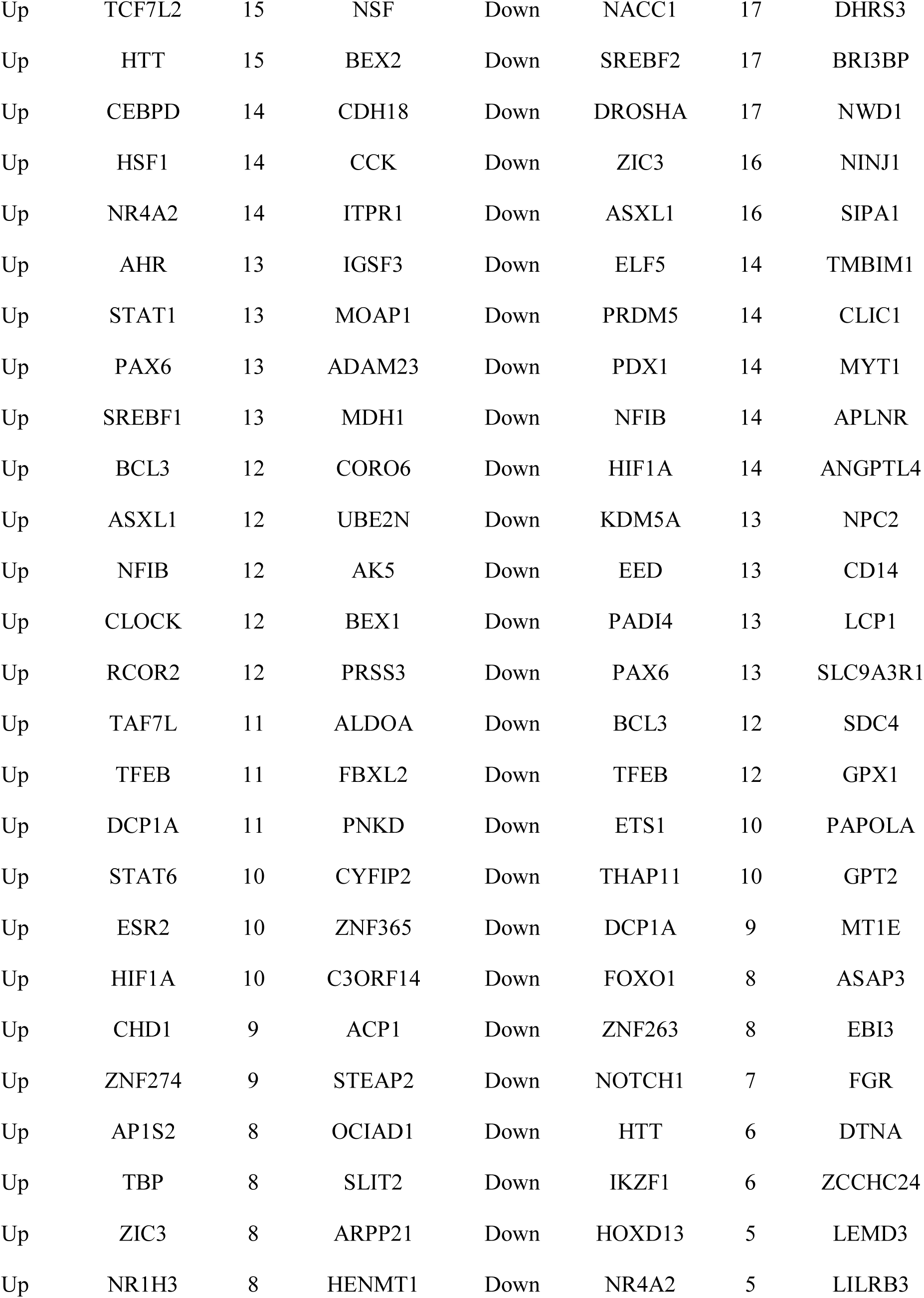
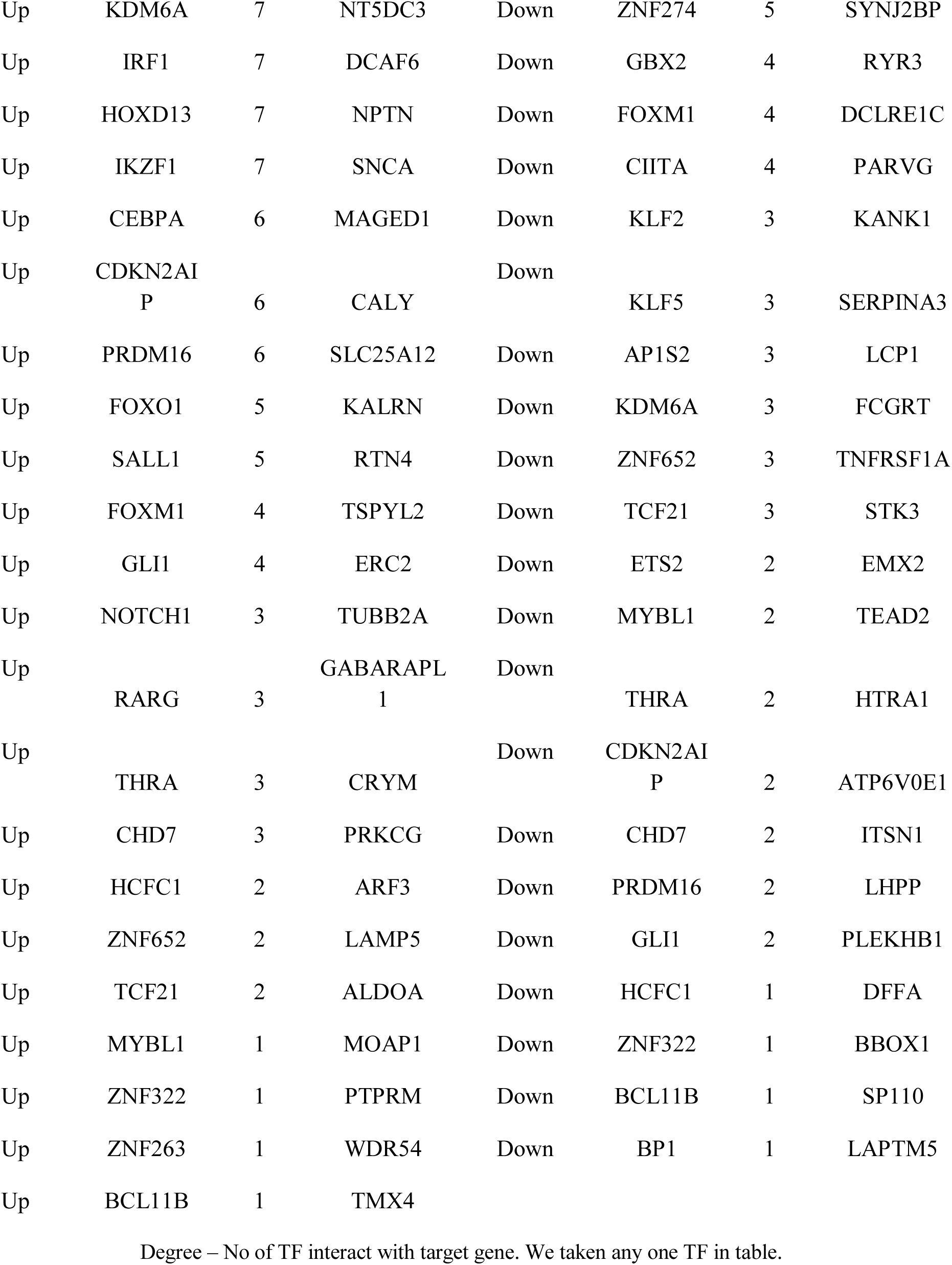
TF - target gene interaction table

### Validation of hub genes

Based on these hub genes between sCJD and normal control, the GLM model was established. AUC values of YWHAZ, GABARAPL1, SSX2IP, YWHAH, UBE2N, HSPB1, HDAC1, CDKN1A, TNFRSF1A and FKBP5 were 0.891, 0.909, 0.873, 0.809, 0.927, 0.973, 0.936, 0.800, 0.918 and 0.945 suggesting that these ten hub genes had high sensitivity and specificity for sCJD prognosis. The ROC results are displayed in Fig. 15. These results indicated that YWHAZ, GABARAPL1, SSX2IP, YWHAH, UBE2N, HSPB1, HDAC1, CDKN1A, TNFRSF1A and FKBP5 may be used as biomarkers for the prognosis of sCJD.

**Fig. 15.**
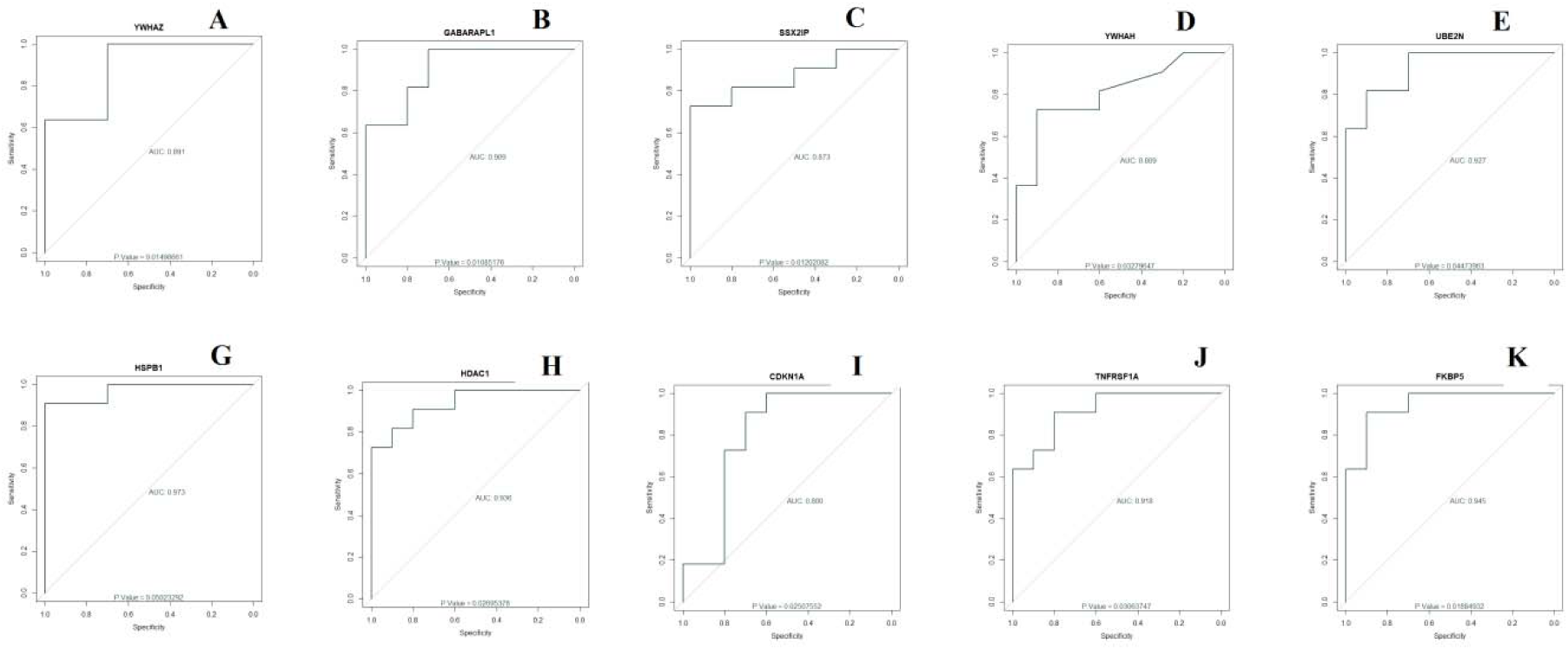
ROC curve validated the sensitivity, specificity of hub genes as a predictive biomarker for sCJD prognosis. A) YWHAZ, B) GABARAPL1, C) SSX2IP, D) YWHAH, E) UBE2N, F) HSPB1, G) HDAC1, H) CDKN1A, I) TNFRSF1A and J) FKBP5

## Discussion

With the development of microarray and high-throughput sequencing technology in current years, integrated bioinformatics methods have been broadly applied in discovering new biomarkers correlated with the diagnosis, prognosis, and treatment of numerous brain diseases [55-56]. sCJD is an prion disease which has brought a heavy burden to European, Australia, and Canadian countries [57]. Understanding the molecular mechanism of sCJD is of essential importance for prognosis, diagnosis and treatment. It has been extensively used to predict therapeutic targets for sCJD since high-throughput sequencing can implement expression levels of genes in human genome simultaneously. The aim of this study was to diagnose several key genes and pathways with similar action highly expressed in sCJD compared to normal controls and discover their potential mechanisms. In the current study, we extract the gene expression profile of GSE124571 downloaded from GEO database and identify 448 up regulated and 443 down regulated genes between RA and normal control using bioinformatics analysis. Less expression of VAMP2 was associated with development of dementia [58], but decrease expression of this gene may be responsible for progression of sCJD. CIRBP (cold inducible RNA binding protein) was linked with neuroinflammation in cerebral ischemia [59], but this gene may be liable for neuroinflammation in sCJD. CRYM (crystallin mu) was involved in progression of Huntington’s disease [60], but this gene may be liable for advancement of sCJD. DOCK3 was involved in growth of hyperactivity disorder [61], but this gene may be associated with progression of sCJD. Genes such as CYFIP2 [62] PC (pyruvate carboxylase) [63] and PLOD1 [64] were responsible for advancement of Alzheimer’s disease, but these genes may be involved in pathogenesis of sCJD. PLOD3 was liable for advancement of brain cancer [65], but this gene may be associated with growth of sCJD. High expression of CRYAB (crystallin alpha B) [66] and DNAJB6 [67] was important for development of Parkinson’s disease, but this gene may be associated with development of sCJD.

In pathway enrichment analysis, up regulated genes were enriched in various pathways. Enriched genes such as GLS2 [68], SLC12A5 [69], GAD1 [70], GAD2 [71], STXBP1 [72], NPTN (neuroplastin) [73] and PPP3CB [74] were associated with development of schizophrenia, but these genes may be important for pathogenesis of sCJD. GLS (glutaminase) was linked with neuroinflammation in brain diseases [75], but this gene may be involved with neuroinflammation in sCJD. Alteration in GNB5 was responsible for advancement of hyperactivity disorder [76], but mutation in this gene may be identified with development of sCJD. Enriched genes such as PRKCB (protein kinase C beta) [77], SNAP25 [78], CALM3 [79], ATP6V0C [80] and SST (somatostatin) [81] were important for pathogenesis of Alzheimer’s disease, but this gene may be linked with progression of sCJD. Enriched genes such as PRKCG (protein kinase C gamma), [82] ITPR1 [83] and VAMP1 [84] were involved in advancement of spinocerebellar ataxia, but these genes may be important for advancement of sCJD. Modification in GABRA1 [85], GABRB3 [86] and GABRG2 [87] were identified with progression of epilepsy, but alteration in these genes may be linked with pathogenesis of sCJD. Mutation in GNAO1 was responsible for progression of movement disorder [88], but Alteration in this gene may associated with development of sCJD. Enriched genes such as STX1A [89], LIN7B [90] and CACNB2 [91] were involved in development of autism, but these genes may be liable for advancement of sCJD. Enriched genes such as NEFL (neurofilament light) [92] and YWHAH (tyrosine 3-monooxygenase/tryptophan 5-monooxygenase activation protein eta) [93] were responsible for development of sCJD. ATP6V1B2 was associated with progression of depression, but this gene may be identified with advancement of sCJD [94]. CHRM1 was associated with pathogenesis of Huntington’s disease [95], but this gene may be liable for advancement of sCJD. Polymorphism in CCKBR (cholecystokinin B receptor) was liable for advancement of Parkinson’s disease [96], this polymorphic gene may be associated with progression of sCJD. Our study found that GNG2, ADCY1, GABARAPL1, GABBR2, SLC32A1, GABRD (gamma-aminobutyric acid type A receptor delta subunit), SLC38A1, GNG3, NSF (N-ethylmaleimide sensitive factor, vesicle fusing ATPase), SYT1, NCALD (neurocalcin delta), CACNG3, CPLX1, KCNJ4, KCNJ12, SYN1, SYN2, CAMK2B, CAMK2G, EPB41L1, ATP6V1A, ATP6V1E1, ATP6V1G2, CAMK1G, PRKCE (protein kinase C epsilon) and GOT1 are up regulated in sCJD and has potential as a novel diagnostic and prognostic biomarker, and therapeutic target. Down regulated genes were enriched in various pathways. Enriched genes such as MAOA (monoamine oxidase A) [97], TGFB3 [98], CEBPD (CCAAT enhancer binding protein delta) [99], NDRG2 [100], SLC11A1 [101], HDAC1 [102], TYROBP (TYRO protein tyrosine kinase binding protein) [103], ALOX5 [104], DOCK2 [105], OLR1 [106], CD14 [107], CD44 [108], CD68 [109], ITPKB (inositol-trisphosphate 3-kinase B) [110], WAS (Wiskott-Aldrich syndrome) [111], CD86 [112] and CD74 [113] were linked with development of Alzheimer’s disease, but these genes may be involved in progression of sCJD. HLA-DPA1 was involved in progression of acute disseminated encephalomyelitis [114], but this gene may be important for development of sCJD. Enriched genes such as HLA-DQA1 [115] and IRF8 [116] were associated with advancement of multiple sclerosis, but these genes may be responsible for pathogenesis of sCJD. Enriched genes such as IFNGR1 [117], PRDX6 [118] and LAMP2 [119] were identified with progression of Parkinson’s disease, but these genes may be linked with advancement of sCJD. NDRG1 was liable for progression of Charcot-Marie-Tooth disease [120], but this gene may be identified with development of sCJD. ITGB4 was liable for progression of bipolar disorder [121], but this gene may be important for advancement of sCJD. SERPINA3 was involved in advancement of sCJD [122]. CLEC5A was important for development of brain cancer [123], but this gene be linked with pathogenesis of sCJD. HSPA1A was responsible for progression of schizophrenia [124], but this gene identified with pathogenesis of sCJD. NPC2 was associated with growth of Niemann-Pick C2 disease [125], but this gene may be important for pathogenesis of sCJD. RNASET2 was linked with progression of cystic leukoencephalopathy [126], but this gene may be liable for advancement of sCJD. Our study found that HLA-DMA (major histocompatibility complex, class II, DM alpha), HLA-DMB (major histocompatibility complex, class II, DM beta), HLA-DOA (major histocompatibility complex, class II, DO alpha), HLA-DRA (major histocompatibility complex, class II, DR alpha), FCGR1A, CYBA (cytochrome b-245 alpha chain), ITGB1, ITGB2, PTPN6, CDKN1A, CEBPA (CCAAT enhancer binding protein alpha), CCND1, ATP8B4, CD300A, CTSC (cathepsin C), TMC6, SERPINB6, FCER1G, FGR (FGR proto-oncogene, Src family tyrosine kinase), SLC2A5, RHOG (ras homolog family member G), PRCP (prolylcarboxypeptidase), PYCARD (PYD and CARD domain containing), VAMP8, CTSH (cathepsin H), ITGAX (integrin subunit alpha X), ADA2, TMBIM1, BST2, LILRB3, RAB31, STOM (stomatin), PLCD1, INPPL1, CSK (C-terminal Src kinase), APBB1IP, PARVG (parvin gamma) and ITGB5 are down regulated in sCJD and has potential as a novel diagnostic and prognostic biomarker, and therapeutic target.

In GO enrichment analysis, up regulated genes were enriched in all GO categories. Enriched genes such as RTN3 [127], VGF (VGF nerve growth factor inducible) [128], SNCA (synuclein alpha) [129], SYP (synaptophysin) [130], NPTX1 [131], NRGN (neurogranin) [132], EGR1 [133], CDK5 [134], MAP1B [135], UCHL1 [136], VPS35 [137], CNTNAP2 [138], KLC1 [139], FZD3 [140], CCK (cholecystokinin) [141], EPHA4 [142], TPM1 [143] and WASF1 [144] were responsible for progression of Alzheimer’s disease, but these genes may be associated in progression of sCJD. PFN2 was liable for development of Charcot-Marie-Tooth disease [145], but this gene may be linked with pathogenesis of sCJD. Enriched genes such as ADCYAP1 [146], NRXN1 [147] LRRTM1 [148], BASP1 [72] and DCLK1 [149] were involved in progression of schizophrenia, but these genes may be responsible for advancement of sCJD. Enriched genes such as SCN1B [150], FGF12 [151], NAPB (NSF attachment protein beta) [152], DNM1 [153] and AP3B2 [154] were important in advancement of epilepsy, but these genes may be identified with progression of sCJD. Enriched genes such as SNCB (synuclein beta) [155], RIT2 [156], TUBB3 [157], PVALB (parvalbumin) [158] and ELAVL4 [159] were involved in progression of Parkinson’s disease, but these genes may be linked with development of sCJD. Enriched genes such as CADPS2 [160], ATP2B2 [161] and ENO2 [162] were important for progression of autism, but these genes may be identified with pathogenesis of sCJD. TAC1 was responsible for advancement of multiple sclerosis [163], but this gene may be associated with progression of sCJD. Enriched genes such NPY (neuropeptide Y) [164], MAP2 [165], APP (amyloid beta precursor protein) [166], STMN2 [167] and PRNP (prion protein) [168] were important for advancement of sCJD. PACSIN1 was liable for advancement of Huntington’s disease [169], but this gene may be involved in progression of sCJD. Our study found that LRFN5, SCN2B, SYT13, AMPH (amphiphysin), ICA1, ADGRL1, PNKD (PNKD, MBL domain containing), SLC8A2, ATP2A2, CLSTN3, CPLX2, PTPRN2, SV2B, GLRB (glycine receptor beta), KALRN (kalirinRhoGEF kinase), SYNGR1, PAK1, PCP4, PDE1A, MAP4, CYGB (cytoglobin), ACOT7, HPCA (hippocalcin), AP1S1, SCAMP5, NGEF (neuronal guanine nucleotide exchange factor), OLFM1, FLRT3, FKBP1A, SVOP (SV2 related protein), SLC9A6, CAP2, MAP2K4, ATCAY (ATCAY, caytaxin), CNTNAP1, ENC1, MYRIP (myosin VIIA and Rab interacting protein), MAGEE1, PPP1R2, ERC2, KCNIP4, ATP1A1, TBR1, NRSN1, MAPK9, MAP2K1, ATP2B1, ARHGAP32, NMNAT2, FRMPD4, SLC6A17, KCTD8, RAP1GAP2, LAMP5, EPB41L3, SLC30A3, NEFM (neurofilament medium), NEFH (neurofilament heavy), NELL2, PTPRN (protein tyrosine phosphatase, receptor type N), ARHGAP44, KIF3C, SLC17A6, GPM6A, DPP6, DMXL2, WDR7, RTN4, STMN1, RGS7, OPA1, SYNGR3, ALDOA (aldolase, fructose-bisphosphate A), BAIAP2L2, GAS7, SYBU (syntabulin), RAB6B, MICAL2, ABLIM2, REEP1, NME1, CDK5R2, DYNC1I1, PTPRT (protein tyrosine phosphatase, receptor type T) and CORO6 are up regulated in sCJD and has potential as a novel diagnostic and prognostic biomarker, and therapeutic target. Down regulated genes were enriched in all GO categories. Enriched genes such as HMOX1 [170], RGCC (regulator of cell cycle) [171], CSF1R [172], HTRA1 [173], BCL6 [174], SYK (spleen associated tyrosine kinase) [175], C1QB [176], ADAM17 [177], PLA2G7 [178], TLR5 [179], PDE9A [180], ITSN1 [181], GJA1 [182], FERMT2 [183], NTRK2 [184], TNFRSF1A [185], S100A1 [186], ALOX5AP [104], APOC2 [187], DIO2 [188] and GSTM3 [189] were responsible for progression of Alzheimer’s disease, but these genes may be liable in progression of sCJD. Enriched genes such as AIF1 [190], ABCA1 [191], AQP4 [192] and GFAP (glial fibrillary acidic protein) [193] were important for pathogenesis of sCJD. Enriched genes such as FGFR3 [194], VEGFB (vascular endothelial growth factor B) [195], GPER1 [196], WASF2 [197], ADRB2 [198], ATXN3 [199], SGK1 [200] and TIMP1 [201] were identified with growth of Parkinson’s disease, but these genes may be linked with progression of sCJD. SLC1A3 was associated with development of hyperactivity disorder [202], but this gene may be liable for progression of sCJD. MEGF10 was involved in progression of schizophrenia [203], but this gene may be liable for advancement of sCJD. Our study found that CMTM3, CEBPB (CCAAT enhancer binding protein beta), PDK4, VSIG4, RFTN2, LY96, VAMP3, SASH3, FCGRT (Fc fragment of IgG receptor and transporter), MERTK (MER proto-oncogene, tyrosine kinase), CHST3, EZR (ezrin), LAT2, MSN (moesin), SLC7A2, CSF3R SCIN (scinderin), GPRC5B, MYOM1, TNFSF14, RASSF2, MYO10, AXL (AXL receptor tyrosine kinase), LILRB1, VSIR (V-set immunoregulatory receptor), HCST (hematopoietic cell signal transducer), STAT5A, MAPKAPK3, C1QA, C1QC, UNC93B1, TAP1, RAC2, RARRES2, GPX1, LGALS9, HAVCR2, EBI3, CD37, LYN (LYN proto-oncogene, Src family tyrosine kinase), TRIB1, HCK (HCK proto-oncogene, Src family tyrosine kinase), HCLS1, HERC5, CYBRD1, DHRS3, SLCO2B1, ADGRV1, FZD9, SLC9A3R1, RAPGEF3, AHNAK (AHNAK nucleoprotein), ANTXR1, PLEK (pleckstrin), CNGB1, SDC4, HSPB1, PDPN (podoplanin), NECAP2, ATP1B2, PIEZO1, SPTBN1, DAB2, MFRP (membrane frizzled-related protein), PTH1R, HEPH (hephaestin), KANK1, KCNMA1, GNA12, DDR1, GRIN2C, LCP1, SLC7A7, RHOQ (ras homolog family member Q), SYTL4, LRP4, EPHX1, CARHSP1, ADD3, SRGAP1, FOXO4, WFS1, RHOBTB3, PPP1R3C, ARHGDIB (Rho GDP dissociation inhibitor beta), RANBP3L, ZFHX3, MAPK4, NEK6, RAB3IL1, NACC2 and FARP1 are down regulated in sCJD and has potential as a novel diagnostic and prognostic biomarker, and therapeutic target.

Up regulated hub genes were identified from PPI network for up regulated genes. YWHAZ was responsible for progression of schizophrenia [204], but this gene may liable for advancement of sCJD. Our study found that SSX2IP, UBE2N, NME5, CAPRIN2 and BEX5 are up regulated in sCJD and has potential as a novel diagnostic and prognostic biomarker, and therapeutic target. Down regulated hub genes were identified from PPI network for down regulated genes. Our study found that FKBP5, MYL12A, SFMBT2, NUPR1, TRIM47 and MS4A7 are down regulated in sCJD and has potential as a novel diagnostic and prognostic biomarker, and therapeutic target.

In module analysis, up regulated hub genes showing the highest node degree in all four significant modules. NCKAP1 was linked with development of Alzheimer’s disease [205], but this gene may be involved in progression of sCJD. RBFOX1 was important for progression autism [206], but this gene may be identified with pathogenesis of sCJD. Our study found that MAGED1, RBFOX2, TUBB2A, USP11, HPRT1 and NCOA7 are up regulated in sCJD and has potential as a novel diagnostic and prognostic biomarker, and therapeutic target. Similarly, down regulated hub genes showing the highest node degree in all four significant modules. Genes such as HSPB8 [207] and BAG3 [208] were responsible for advancement of Alzheimer’s disease, but these genes may be important for progression of sCJD.

In target gene - miRNA network, up regulated target genes showing the highest number of integration with miRNAs. Our study found that MAP3K9 and PGM2L1 are up regulated in sCJD and has potential as a novel diagnostic and prognostic biomarker, and therapeutic target. Similarly, down regulated target genes showing the highest number of integration with miRNAs. Our study found that MKNK2, SYNJ2BP and BRI3BP are down regulated in sCJD and has potential as a novel diagnostic and prognostic biomarker, and therapeutic target

In target gene - TF network, up regulated target genes showing the highest number of integration with TFs. Our study found that CDK14 is up regulated in sCJD and has potential as a novel diagnostic and prognostic biomarker, and therapeutic target. Similarly, down regulated target genes showing the highest number of integration with TFs. Our study found that TST is down regulated in sCJD and has potential as a novel diagnostic and prognostic biomarker, and therapeutic target.

In conclusion, although the current study had certain conditions, including the limited number of cases and the absence of validation in clinical samples, the current analysis diagnosed distinct important genes and pathways closely linked with sCJD, which may contribute to the current knowledge of the complex molecular mechanisms of sCDJ. Of note, the current results credential acceptance by further examination.

## Data Availability

The datasets supporting the conclusions of this article are available in the GEO (Gene Expression Omnibus) (https://www.ncbi.nlm.nih.gov/geo/) repository. [(GSE124571) (https://www.ncbi.nlm.nih.gov/geo/query/acc.cgi?acc=GSE124571)]

## Acknowledgement

We thank Piero Parchi, University of Bologna, DIMES, Neuropathology, Bologna, Italy, very much, the authors who deposited their microarray dataset, GSE124571, into the public GEO database.

## Consent for publication

Not applicable.

## Ethical approval

This article does not contain any studies with human participants or animals performed by any of the authors.

## Informed consent

No informed consent because this study does not contain human or animals participants.

## Author Contributions

B. V. - Writing original draft, and review and editing

C. V. - Software and investigation

I. K. - Supervision and resources

## Authors

Basavaraj Vastrad ORCID ID: 0000-0003-2202-7637

Chanabasayya Vastrad ORCID ID: 0000-0003-3615-4450

Iranna Kotturshetti ORCID ID: 0000-0003-1988-7345

